# Changes in Children’s Fitness and Health Status Over the Course of the COVID-19 Pandemic: A 34-month Longitudinal Study of 331 Primary School Children (September 2019 to June 2022)

**DOI:** 10.1101/2022.12.01.22282992

**Authors:** Gerald Jarnig, Reinhold Kerbl, Mireille N.M. van Poppel

## Abstract

**Background:** Previous studies have shown that coronavirus disease 2019 (COVID-19) mitigation measures lead to reductions in health status and health-related fitness in children, whereas no data are currently available on the development of children after the removal of these mitigation measures. Therefore, we aimed to investigate the influence of different stringent measures, during and after COVID-19, on the longitudinal development of fitness and health status of primary school children.

**METHODS:** A longitudinal cohort study of 331 primary school children in Austria aged 7.7 ± 0.4 years at the start of the observation period was conducted. Before (Sept 2019), and during (June 2020, Sept 2020) and after (March 2021, June 2021, Sept 2021, and June 2022) the COVID-19 mitigation measures, fitness tests were carried out and anthropometric data of the children were collected. Age- and sex-specific standard deviation scores (SDSs) were calculated for seven individual fitness tests of the AUT FIT [1] test battery, body mass index, and waist-to-height ratio. In addition, children were classified into different fitness categories based on the AUT FIT summary score.

**Results:** After the implementation of COVID-19 mitigation measures, cardiorespiratory fitness and action speed decreased dramatically, and the number of children with overweight or obesity increased. At the last measurement after the COVID-19 mitigation measures (June 2022), significant improvements in cardiorespiratory fitness and weight status were observed. However, pre-COVID fitness levels were not achieved and BMI was still higher compared to Sept 2019.

**Interpretation:** Our study showed a trend reversal, in that the negative indirect effects of the COVID-19 measures on fitness and health parameters of children were followed by significant improvements in these parameters after the removal of those mitigation measures. At the end of the observation period, a positive trend in the development of fitness and health status was observed, although pre-COVID levels were not reached. Collaborative efforts are needed to reinforce the observed positive trend, and to continue to improve the level of fitness and health status of primary school children, and to mitigate the long-term negative health consequences of the COVID-19 pandemic.

**Funding:** Austrian Federal Ministry of Arts, Culture, Public Service and Sports (GZ2021-0.361.671).

## INTRODUCTION

In late 2019, when the COVID-19 pandemic began spreading globally from Wuhan, China, heavy restrictions were enforced on public places around the world to best mitigate the spread of the severe acute respiratory syndrome coronavirus 2 (SARS-CoV-2) virus [2].

Children and adolescents were particularly affected by the restrictions due to COVID-19-related school closures and severely limited sports and leisure opportunities [3, 4].

This, in combination with greatly reduced of physical activity [3,5–11], increased screen [5,6,8–10] and sedentary time during leisure time [4, 7], changes in eating habits [8–10,12], and shifted sleep times [8–10,13] led to lifestyle changes in children and adolescents worldwide.

These lifestyle changes were directly related to a decrease in cardiorespiratory fitness [14–17] and a significant increase in the number of children suffering overweight or obesity [18–22].

During the course of the COVID-19 pandemic, dramatic declines in cardiorespiratory fitness and a dramatic increase in body mass index in children and adolescents have been reported [18,23,24].

Cardiorespiratory fitness in childhood is an important health marker [25], and higher CRF levels are associated with lower BMI, waist circumference, and body fat measurements, as well as a lower prevalence of metabolic syndrome later in life [26]. Additionally, childhood obesity is associated with increased cardiovascular risk factors and coronary heart diseases [27, 28].

In Austria, a research project started in September 2019, before the COVID-19-related restrictions in school (13 March 2020), in which data on the fitness and health status of primary school children were collected at seven testing time points (September 2019 to June 2022).

To the best of our knowledge, there are currently no studies reporting changes in cardiorespiratory fitness, BMI, abdominal girth-to-height ratio, and other fitness parameters over the first 34 months of the COVID-19 pandemic using objectively measured data.

The aim of our study was to investigate how children’s fitness and health status changed over the course of the COVID-19 pandemic, specifically children’s development after relaxation of the stringent COVID-19 mitigation measures beginning in June 2021.

## METHODS

This study was originally designed as a randomized controlled trial to investigate the effects of a physical education intervention on the fitness and health status of primary school children in Klagenfurt, Austria. Due to the COVID-19 regulations, the intervention had to be discontinued in March 2020. However, the planned follow-up measurements in June 2020, September 2020, March 2021, June 2021, September 2021, and June 2022 were carried out under the COVID-19-related hygiene regulations valid at the time of testing. Because of the discontinued intervention study, we analyzed the data as that of a longitudinal study and assessed the effects of COVID-19 mitigation measures on the fitness and health status in children aged 7–10 years. The study was registered in the German Clinical Trials Registry (ID DRKS00023824) and approved by the Research Ethics Committee of the University of Graz, Styria, Austria (GZ. 39/23/63 ex 2018/19).

### Selection of Schools and Participants

A list of all 39 primary schools in the greater Klagenfurt area, Austria, was used to select the schools. Using a random number generator, 12 schools were selected and randomly divided into intervention and control groups. All schools agreed to participate in the study. The inclusion criteria were as follows: aged between 7 and 10 years at study baseline and physical ability to perform all of the motor skills tests in the test battery. In spring 2019, all children who were attending first or second grade in any of the 12 schools at that moment were invited to participate in the study. Because those children in the second grade left school in 2021 and thus were not available for follow-up measurements in September 2021 and June 2022, we limited the analysis in this paper to the 516 children who attended first grade at baseline. A total of 433 (83.9%) legal guardians of these children gave their written consent to participate (Figure 1) and reported in writing whether or not their child was a member of a sports club. Meanwhile, 412 children attending first grade in their primary school in school year (SY) 2018/2019 participated in baseline testing (T1) prior to the COVID-19 pandemic (September 2019). Over the course of the different testing phases, children dropped out of the longitudinal study for various reasons (change of school or class, illness, injury, COVID-19, etc.) during the follow-up assessments (September 2019 (T1), June 2020 (T2), September 2020 (T3), March 2021 (T4), June 2021 (T5), September 2021 (T6), and June 2022 (T7)). In total, anthropometric data were collected from 331 children at all test time points and the same number of children completed the six-minute run (6MR) at all possible test time points. For the remaining fitness tests, only those children who completed all of the tests in the remaining test battery at all possible time points were analyzed. Since the tests were carried out, in part, on different test days, the number of study participants varied between the anthropometric data, cardiorespiratory fitness, and the remaining fitness tests, for which a total of 276 children were finally available for the overall assessment. For the medicine ball throw, there were also 276 children present at all testing time points, but due to a lack of reference values for children over the age of 10, only 255 children could be included in the overall analysis (Figure 1).

**Figure 1.**
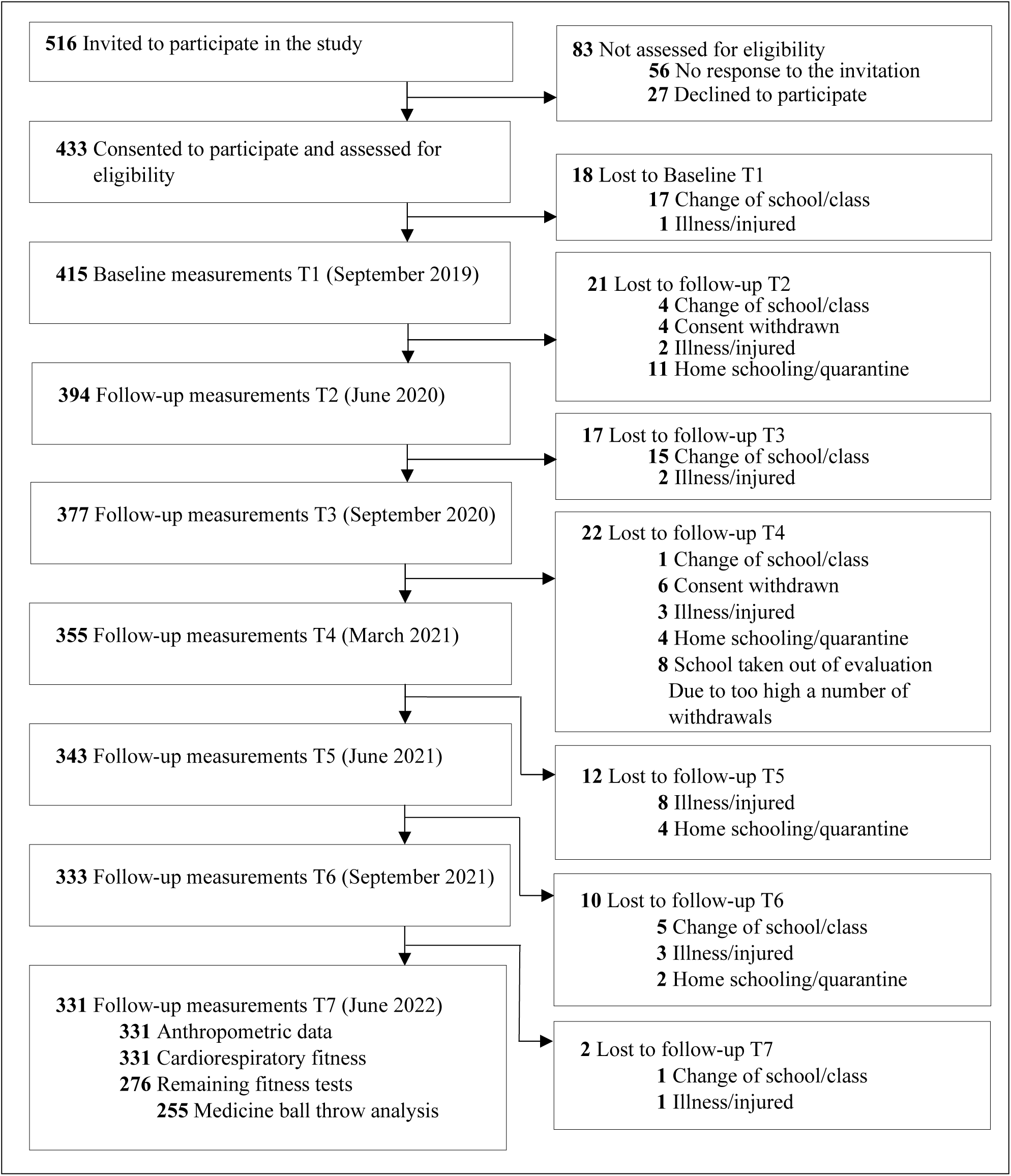
Flow diagram.

### Measurements

All measurements were standardized and piloted with trained personnel (sports scientists) in June 2019 on non-participating children (*n* = 98) of the same age group. Baseline measurements (T1) were carried out in September 2019. When the lockdown was slowly relaxed in spring 2020 (Tables M1 and M2 and Figures M1 and M2), the legal department of the Carinthian Directorate of Education allowed a second test phase (T2) in June 2020, which was performed under strict hygiene measures and the nationwide COVID-19 regulations in place at the time. The cardio running test (6MR) was not allowed at that time because the minimum distance between participants could not be guaranteed. The third test phase (T3) took place in September 2020, during which the complete test battery was carried out. The fourth test phase was conducted in March 2021 (T4), directly after the second major lockdown phase. Due to a very short time window, the complete test battery was not performed at T3. Only anthropometric and health-related fitness data (cardiorespiratory fitness, action speed, and strength) were collected. The fifth, sixth, and seventh test phases were carried out in June 2021 (T5), September 2021 (T6), and June 2022 (T7), respectively, and all fitness tests could be carried out as planned at these time points.

**Table M1.** Levels of COVID-19-related restrictions for children in Austria in schools and sports classes from 31 January 2020 to 31 May 2022 regarding the OxCGRT Stringency Index.

| Period | Index<sup>a</sup> | School Structure | Physical Education in School | Sports and Leisure Facilities | Sports at the Club | Stay at Home Requirements | Sum of Restrictions in the School |
| --- | --- | --- | --- | --- | --- | --- | --- |
| 31.01.2020–23.02.2020 | 0.00 | 0 | 0 | 0 | 0 | 0 | 0 |
| 24.02.2020–08.03.2020 | 11.11 | 0 | 0 | 0 | 0 | 0 | 0 |
| 09.03.2020–10.03.2020 | 19.44 | 0 | 0 | 0 | 0 | 0 | 0 |
| 12.03.2020 | 34.26 | 0 | 0 | 0 | 0 | 0 | 0 |
| 13.03.2020–15.03.2020 | 48.15 | 0 | 0 | 0 | 0 | 0 | 0 |
| 16.03.2020–13.04.2020 | 81.48 | 3 | 3 | 3 | 3 | 3 | 15 |
| 14.04.2020–22.04.2020 | 77.78 | 3 | 3 | 3 | 3 | 3 | 15 |
| 23.04.2020–30.04.2020 | 75.00 | 3 | 3 | 3 | 3 | 3 | 15 |
| 01.05.2020 | 67.59 | 3 | 3 | 2 | 3 | 2 | 13 |
| 02.05.2020–10.05.2020 | 64.81 | 3 | 3 | 2 | 3 | 2 | 13 |
| 11.05.2020–17.05.2020 | 59.26 | 3 | 3 | 2 | 3 | 2 | 13 |
| 18.05.2020–28.05.2020 | 59.26 | 2 | 3 | 2 | 3 | 2 | 12 |
| 29.05.2020–02.06.2020 | 53.70 | 2 | 3 | 2 | 2 | 2 | 11 |
| 03.06.2020 | 50.00 | 2 | 3 | 2 | 2 | 2 | 11 |
| 04.06.2020–1.07.2020 | 47.22 | 2 | 3 | 2 | 2 | 2 | 11 |
| 02.07.2020–09.07.2020 | 47.22 | 2 | 3 | 1 | 1 | 0 | 7 |
| 10.07.2020–05.09.2020 | 35.19 | 0 | 0 | 1 | 1 | 0 | 2 |
| 06.09.2020–13.09.2020 | 36.11 | 0 | 0 | 1 | 1 | 0 | 2 |
| 14.09.2020–16.09.2020 | 36.11 | 1 | 0 | 1 | 1 | 1 | 4 |
| 17.09.2020–28.09.2020 | 37.04 | 1 | 0 | 1 | 1 | 1 | 4 |
| 29.09.2020–12.10.2020 | 40.74 | 1 | 0 | 1 | 1 | 1 | 4 |
| 13.10.2020–16.10.2020 | 44.91 | 1 | 0 | 1 | 1 | 2 | 5 |
| 17.10.2020–22.10.2020 | 58.80 | 1 | 0 | 1 | 1 | 2 | 5 |
| 23.10.2021 | 60.19 | 1 | 0 | 1 | 1 | 2 | 5 |
| 24.10.2020–26.10.2020 | 60.19 | 0 | 0 | 1 | 3 | 2 | 6 |
| 27.10.2020–01.11.2020 | 64.81 | 0 | 0 | 1 | 3 | 2 | 6 |
| 02.11.2020 | 75.00 | 0 | 0 | 1 | 3 | 2 | 6 |
| 03.11.2020–16.11.2020 | 75.00 | 0 | 2 | 1 | 3 | 2 | 8 |
| 17.11.2020–06.12.2020 | 82.41 | 3 | 3 | 1 | 3 | 2 | 12 |
| 07.12.2020–23.12.2020 | 71.30 | 1 | 2 | 1 | 3 | 1 | 8 |
| 24.12.2020–25.12.2020 | 82.41 | 3 | 3 | 1 | 3 | 2 | 12 |
| 26.12.2020–06.01.2021 | 82.41 | 3 | 3 | 1 | 3 | 2 | 12 |
| 07.01.2021–18.01.2021 | 82.41 | 3 | 3 | 1 | 3 | 2 | 12 |
| 19.01.2021–05.02.2021 | 82.41 | 2 | 2 | 1 | 3 | 1 | 9 |
| 06.02.2021–07.02.2021 | 82.41 | 3 | 3 | 1 | 3 | 1 | 11 |
| 08.02.2021–11.02.2021 | 74.07 | 3 | 3 | 1 | 3 | 1 | 11 |
| 12.02.2021–14.02.2021 | 79.63 | 3 | 3 | 1 | 3 | 1 | 11 |
| 15.02.2021–14.03.2021 | 75.93 | 1 | 2 | 1 | 3 | 1 | 8 |
| 15.03.2021–31.03.2021 | 73.15 | 1 | 2 | 1 | 2 | 1 | 7 |
| 01.04.2021–05.04.2021 | 75.00 | 1 | 2 | 1 | 2 | 1 | 7 |
| 06.04.2021–25.04.2021 | 78.70 | 1 | 1 | 1 | 2 | 1 | 6 |
| 26.04.2021–02.05.2021 | 76.85 | 1 | 1 | 1 | 2 | 1 | 6 |
| 03.05.2021–10.05.2021 | 75.00 | 1 | 1 | 1 | 2 | 1 | 6 |
| 11.05.2021–15.05.2021 | 73.15 | 1 | 1 | 1 | 2 | 1 | 6 |
| 16.05.2021–18.05.2021 | 65.74 | 1 | 1 | 1 | 2 | 1 | 6 |
| 19.05.2021–09.06.2021 | 68.52 | 1 | 0 | 0 | 1 | 0 | 2 |
| 10.06.2021–30.06.2021 | 60.19 | 1 | 0 | 0 | 1 | 0 | 2 |
| 01.07.2021–10.08.2021 | 49.07 | 0 | 0 | 0 | 0 | 0 | 0 |
| 11.08.2021–24.08.2021 | 56.02 | 0 | 0 | 0 | 0 | 0 | 0 |
| 25.08.2021–08.09.2021 | 58.80 | 0 | 0 | 0 | 0 | 0 | 0 |
| 09.09.2021–12.09.2021 | 49.07 | 0 | 0 | 0 | 0 | 0 | 0 |
| 13.09.2021–14.09.2021 | 49.07 | 0 | 1 | 0 | 0 | 0 | 1 |
| 15.09.2021–17.09.2021 | 51.85 | 0 | 1 | 0 | 1 | 0 | 2 |
| 18.09.2021–28.09.2021 | 60.19 | 0 | 1 | 0 | 1 | 0 | 2 |
| 29.09.2021–30.09.2021 | 51.85 | 0 | 1 | 0 | 1 | 0 | 2 |
| 01.10.2021–21.11.2021 | 49.31<sub>b</sub> | 0 | 1 | 0 | 1 | 0 | 2 |
| 22.11.2021–28.02.2022 | 50.77<sub>b</sub> | 1 | 1 | 0 | 1 | 0 | 3 |
| 01.03.2022–19.04.2022 | 39.28<sub>b</sub> | 1 | 0 | 0 | 0 | 0 | 1 |
| 20.04.2022–31.05.2022 | 37.96<sub>b</sub> | 0 | 0 | 0 | 0 | 0 | 0 |

<sup>a</sup> COVID-19: Government Stringency Index; <sup>b</sup> average value. 0 = no restrictions, 1 = low restrictions, 2 = medium restrictions, 3 = high restrictions.

**Table M2.**
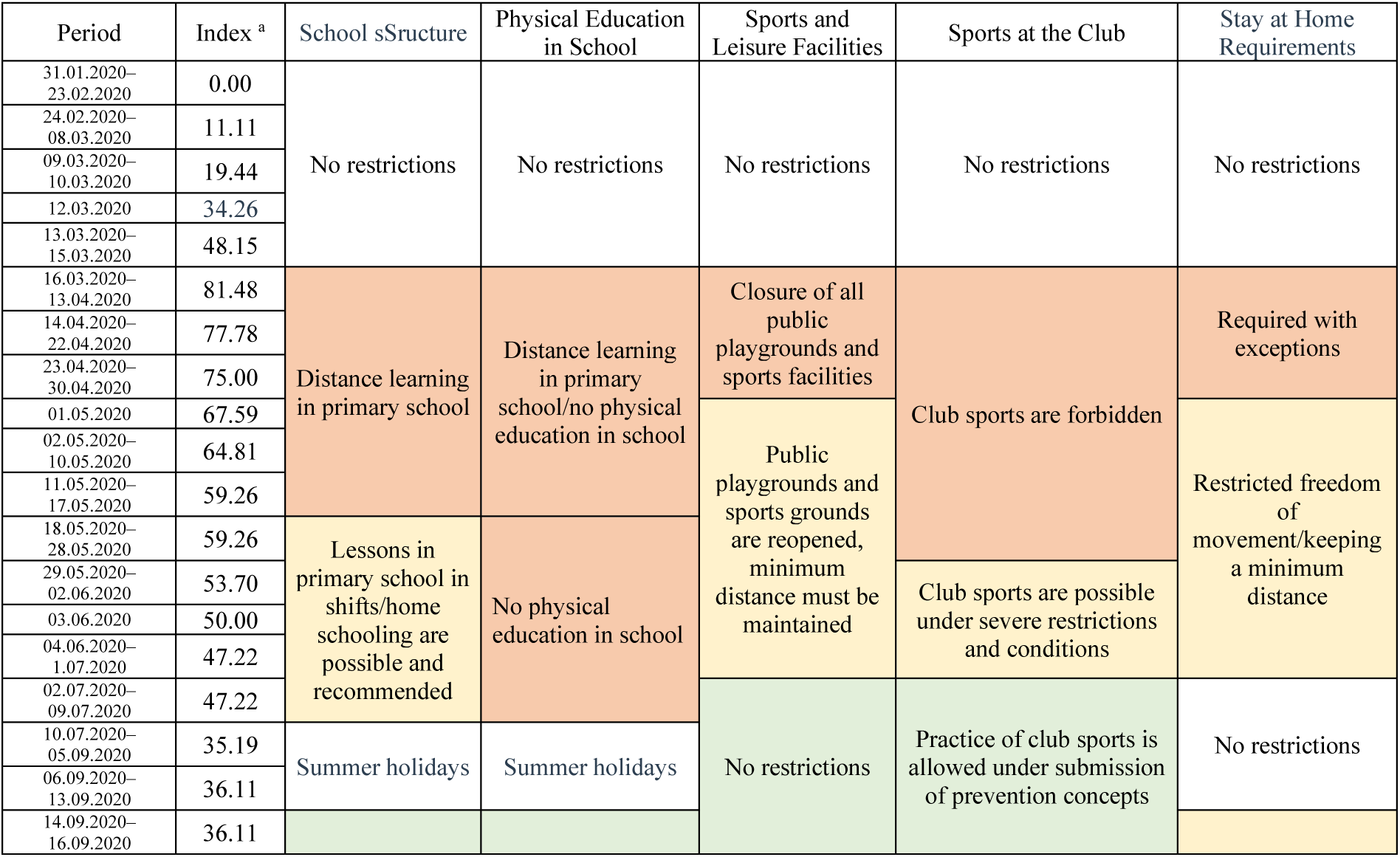
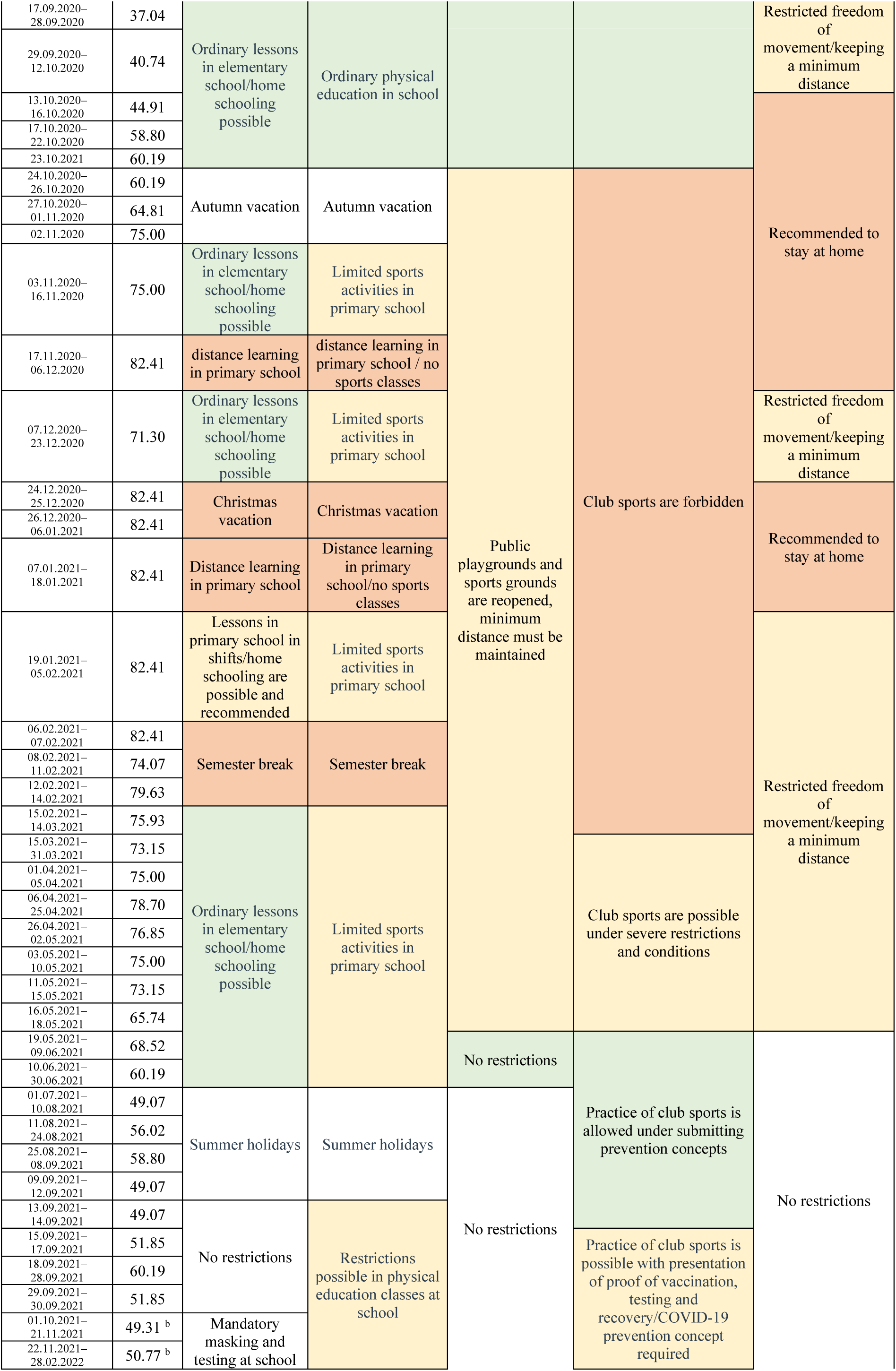
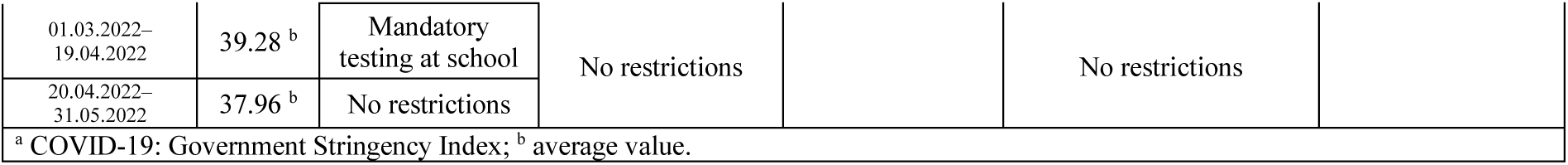
Detailed description of COVID-19 restriction levels for children in Austria in schools and sports classes from 31 January 2020 to 31 May 2022 in relation to the OxCGRT Stringency Index.

**Figure M1.**
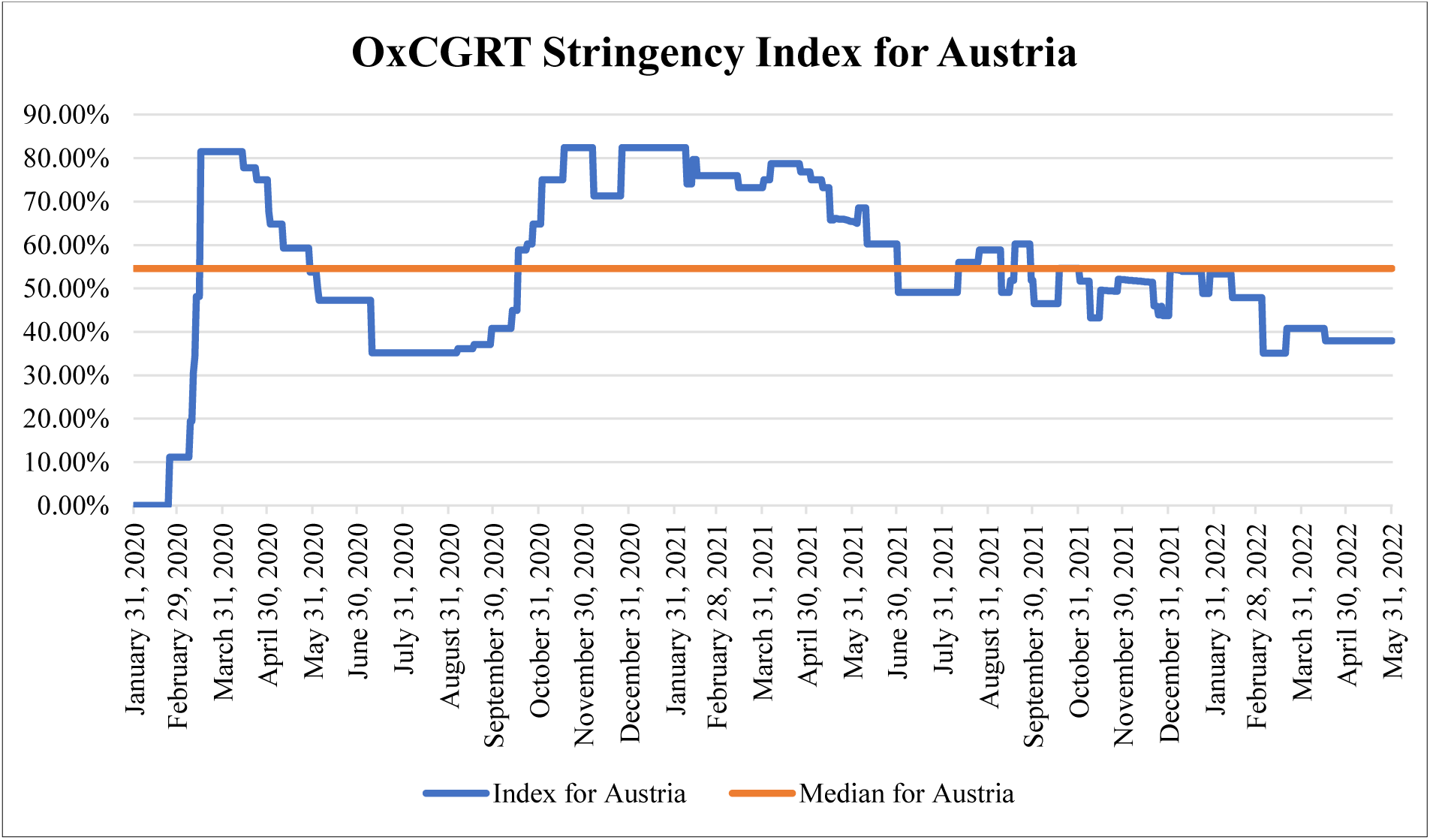
OxCGRT Stringency Index for Austria from 31 January 2020 to 31 May 2022.

**Figure M2.**
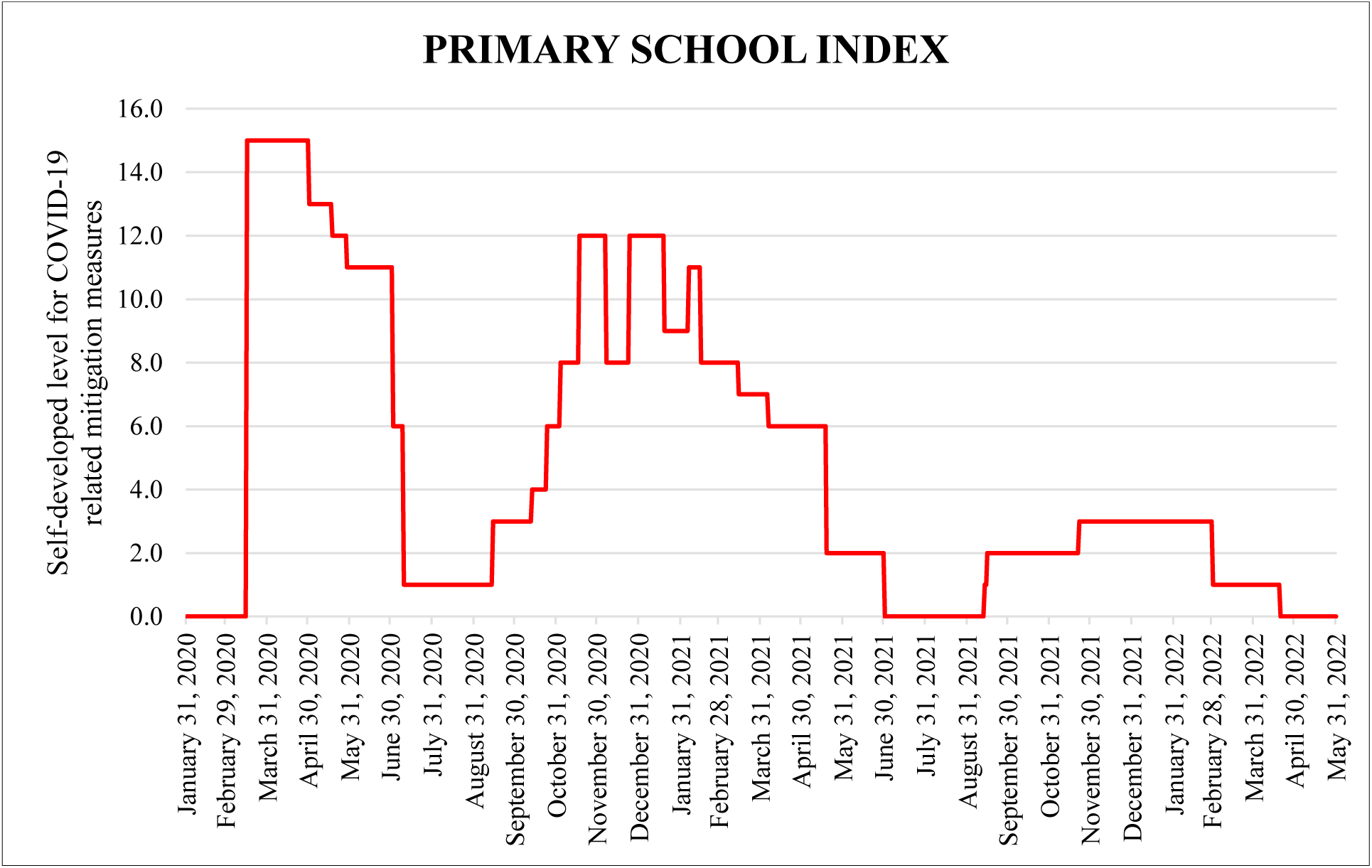
Self-developed levels for COVID-19-related restrictions for children in Austrian schools and sports classes from 31 January 2020 to 31 May 2022.

The intervention began after the baseline measurement in September 2019. In the intervention group, all physical education lessons were planned and implemented by external coaches and sports scientists according to the Austrian school curriculum. The control group received their usual physical education classes at school. The intervention and regular school classes were canceled due to the COVID-19 pandemic on 16 March 2020.

To visualize government-ordered restrictions over the time period considered, the Oxford COVID-19 Government Response Tracker (OxCGRT) [29] provides an internationally comparable level (Figure M1). Based on the OxCGRT classification, a more detailed, self-developed restriction assessment was developed to visualize the impact of the pandemic mitigation measures on children over time (Figure M2). The detailed description of this classification method is based on the Austrian legal decrees published in the Austrian Federal Law Gazette [30].

During the observation period, a sports intervention was designed and implemented from September 2021 to June 2022. For this purpose, the children were again randomly divided into intervention and control groups. The physical activity classes of the children in the intervention groups were planned by external experts according to the Austrian school curriculum and instructed on site by external coaches in the schools. In addition, on days when there were no physical classes, the children were instructed to learn cognitive content with specific movement activities twice a week, while movement activity was given as homework once a week. The planning of these additional specific movement-supporting work materials was also carried out by external experts, provided to all class teachers and implemented by them on site in the school.

### Procedure

For this study, the primary outcomes were the effects of the COVID-19 mitigation measures on children’s fitness and health status, specifically children’s follow-up development after relaxation of the stringent COVID-19 mitigation measures.

#### Anthropometrics

Anthropometric data included height (cm), weight (kg), and abdominal circumference (cm). The children’s height was measured to the nearest 0.1 cm using a portable stadiometer (SECA 213, Hamburg, Germany). Weight was measured to the nearest 0.1 kg using an electronic scale (BOSCH PPW4202/01, Nuremberg, Germany). Abdominal circumference was measured to the nearest 1 cm using a commercial tape measure. BMI was calculated by dividing weight in kilograms by height in meters squared. To calculate the waist-to-height ratio (WtHR), abdominal circumference (cm) was divided by height (cm).

#### Fitness

Different fitness tests were used to assess the fitness of the children (a detailed description can also be found in the work of Jarnig et al. [1]).

#### Endurance

The six-minute run (6MR) was used to analyze cardiorespiratory fitness. The children were instructed to run as far as possible within 6 min. The test was performed in the schoolyards, athletic halls, and sports fields of the schools. A square (6 × 18 m) was marked out with sports poles, then the four corner poles were moved 0.5 m inward. The children had to run around the marked square. A group of six to seven children performed the test simultaneously, and the completed running distance was measured to the nearest meter.

#### Muscular Endurance and Whole-body Coordination

To assess muscular endurance and whole-body coordination, jumping sideways (JS) ability was tested. The instructor marked an area (100 × 50 cm) on the floor, which was divided into two squares (50 × 50 cm) with marking tape. The children stood with both legs in the middle of one square and jumped between the squares with both legs for 15 s after the start command. The aim was to complete as many jumps as possible without touching the marker. If the child touched the marker, that jump was not counted. The instructor counted the number of valid jumps, and each jump across the center line was counted as one jump attempt. Each child had two scoring attempts, and the average of the number of valid jumps from both attempts was used.

#### Lower Body Strength

The standing long jump (SLJ), used to assess lower body muscular strength, is considered as an index for general assessment of muscular fitness in children. The children were instructed to jump as far as they could with both legs from a starting line, and the shortest distance between the starting line and the contact of the child’s heel with the ground was measured with a tape measure to the nearest centimeter. Three scoring attempts were carried out, and the widest of the three jumps was scored.

#### Upper Body Strength

The 1 kg medicine ball throw (MB1kg) was used to measure upper body muscle strength. Each child stood on a starting line, holding a 1 kg medicine ball with both hands while the ball was touching their chest, and threw the ball forward as far as possible with both hands.

The shortest distance between the starting line and the ball’s contact with the ground was measured with a tape measure to the nearest centimeter. Each child had two attempts to throw the ball, and the widest throw was scored.

#### Flexibility

Flexibility was measured with the sit and stretch test. An expensive test box is required to perform the classic sit-and-reach test; therefore, the V-Sit and Reach (VSR) test was chosen as an alternative method, which can be performed with a tape measure and a marking tape. The tape measure was attached to the floor and a heel line was marked with tape. The children sat on the floor with their feet 30 cm apart and their heels on the heel line, placed one hand on top of the other, and slowly stretched forward as far as they could. The distance between the heel line and the maximum position reached that could be held with the fingertips for 2 s was noted. Each child had two scoring attempts, with the longest reach included in the overall score. The standard values of the classic sit and reach test were used as reference values.

#### Action Speed

To assess the children’s action speed, a shuttle run test (4 × 10 SHR) was performed. Two lines (starting and turning line) were marked on the floor at 10 m intervals. Two objects (O1 and O3) were placed behind the turning line and one easy-to-grasp object (O2) was placed in front of the starting line. The children had to run from the starting line across the turning line, pick up O1, run back across the starting line, and set O1 down. Then, they had to pick up O2 and run across the turn line again, put down O2, pick up O3, and run across the starting line with it. The children were instructed to complete this test as fast as possible. Two scoring attempts were made, and the time was measured with a stopwatch to the nearest 0.01 s. Each child had two attempts, and the fastest run was scored.

#### Reaction Speed

To test the reaction speed, a ruler drop test (RD) was performed. A ruler drop stick was constructed (see Jarnig et al. [1]) and held by the instructor. Each child formed a 45° angle with their thumb and outstretched fingers, and the instructor held the ruler drop stick centered in this area. The zero point was held at the level of the bottom of the thumb, and the instructor dropped the rod within 3 s after the command “Ready.” The distance in centimeters that the stick fell was recorded. Each child had one test trial and five scoring attempts. The best and worst attempts were removed from the scoring, and the average score was calculated from the remaining three attempts and included in the overall score.

#### Balance

Existing balance tests are very time-intensive, so the standard single-leg stand (SLS) test was used, in which each leg was assessed for 1 min. The children were instructed to stand with one leg on a thin wooden board, place hands on hips, and hold this position for as long as possible. The test was performed with each leg for a maximum of 45 s (left, SLS-L, and right, SLS-R), and the best result (in seconds) from two attempts for each leg was used. If the child achieved the maximum score (45 s) on the first attempt, a second attempt with the same leg was not performed. Detailed information on the construction of the test apparatus was described by Jarnig et al. [1].

The validity and reliability of these four fitness tests have been described in detail before and have been reported to be suitable for use in field tests [1].

Although Jarnig et al. [1] described, in their reported paper, that the RD is not suitable for widespread use and that the JS test manual needs to be adapted, both tests continued to be performed as described because of the extraordinary attention given to the data due to the COVID-19 pandemic and the fact that there are few direct metric measures that allow direct conclusions to be made about fitness levels before and during the course of the COVID-19 pandemic.

## STATISTICAL ANALYSIS

Descriptive statistics were calculated for all time points (T1, T2, T3, T4, T5, T6, and T7). Continuous variables are presented as the mean (M) and standard deviation (SD) and categorical variables as absolute values (No.) and percentages (%). The analyzed data contain only complete data for all measurement time points of each fitness test, and no imputation of the data was performed.

### Standardization of Anthropometrics

Weight standardization and classification and health risk estimation were performed according to the methodology described in the pilot study for AUT FIT [1].

In short, national reference values were used for BMI standardization and weight classification. Absolute BMI values were converted to EQUI-BMI values (referred to as EQUI BMI_AUT_ in this paper) using the procedure described by Mayer et al. [31] (based on Cole et al. [32]). EQUI-BMI curves can be used to project actual BMI to cutoff values at an age of 18 years relevant to classifying children’s weight into five categories (underweight EQUI BMI_AUT_ <18.5 kg/m^2^, normal weight EQUI BMI_AUT_ 18.5–25.0 kg/m^2^, overweight EQUI BMI_AUT_ ≥25.0 kg/m^2^, obese EQUI BMI_AUT_ ≥30.0 kg/m^2^, morbidly obese EQUI BMI_AUT_ ≥35.0 kg/m^2^) [31].

The cutoff values of 0.5 and 0.6 were used to classify the WtHR and to assess health risk more comprehensively, and the WtHR was divided into no (<0.5), increased (0.5–0.6), and high (≥0.6) health risk [33, 34].

### Standardization of Fitness

To compare the results of the fitness tests (raw scores) with different existing reference values, *z*-scores were formed based on the most current age- and gender-specific reference values. Since no reference values are available for Austrian children of this age, international reference values were used. Current international percentile tables were used for the application of the LMS method to calculate SD scores (6MR (Ref.= German standard values—Düsseldorfer Modell; DüMo) [35], SLJ (Ref. = German standard values— Düsseldorfer Modell; DüMo) [35], 4×10SHR (Ref. = Portuguese standard values; MCA) [36], and VSR (Ref. = Macedonian standard values; MAKFIT) [37]). International reference values were also used for traditional *z*-score standardization (RD (Ref. = German norm values; Fetz) [38], JS (Ref. German norm values—German Motor Test; GMT) [39], and MB1kg (Ref. German norm values—Karlsruhe Testing System; KATS-S) [40]).

Balance was assessed using the point score calculation described by Jarnig et al. [1].

Based on the methodology described in AUT FIT (Jarnig et al. [1]), the SDS or *z*-scores of each fitness test were converted into a nine-point score (STA9). For the classification of each test, the worst performance was represented by 1 point and the best performance by 9 points.

To simplify the presentation of the data for the viewer, the nine categories of assessment were combined into three collective categories. The original categories of “poor,” “very poor,” and “poor” athletic performance were described as “poor” athletic performance. The original categories of “below average,” “average,” and “above average” athletic performance were described as “average” athletic performance. The original categories of “very good,” “excellent,” and “exceptional” athletic performance were described as “good” athletic performance.

### Changes Over Time

Changes in EQUI BMI_AUT_, WtHR, 6MR SDS (DüMo), SLJ SDS (DüMo), MB1kg *z*-score (KATS-K), 4×10SHR SDS (MCA), VSR SDS (MAKFIT), JS *z*-score (GMT), RD *z*-score (Fetz), and SLS (SLS-L and SLS-R) were analyzed over the observation period using mixed analyses of variance (ANOVAs) for sports club membership, group membership in the last SY (intervention: yes or no), and test time points (T1, T2, T3, T4, T5, T6, and T7) were analyzed. In the case of nonsphericity, Greenhouse–Geisser correction was performed. Homogeneity was tested with the Hartley Fmax test. For ANOVAs, partial eta squared (ⴄ_p_^2^) was used to determine the effect size (≥0.01 = small, ≥0.06 = medium, ≥0.14 = large) [41], and only an effect that was at least small was considered relevant.

Changes in the distribution of BMI, WtHR, and fitness classification categories over time were tested with the Friedman test. Post hoc tests were performed using the Wilcoxon signed-rank test.

For the different performance trends in the intervention and control groups in the last SY (September 2021 (T6) to June 2022 (T7)), Δ SDS and Δ *z*-values were calculated and analyzed using an unpaired *t*-test.

All tests were two-sided, and a *p*-value of <0.05 was considered statistically significant. Bonferroni correction was used for the post hoc tests.

All statistical calculations were performed with SPSS version 28 (IBM Corp. Released 2021. IBM SPSS Statistics for Windows, Armonk, NY: IBM Corp).

## RESULTS

The included study population that participated at all measurement time points *(n =* 331) and the group (*n* = 86) that could not be followed up were compared on the variables of age, sex, sports club affiliation, BMI, and abdominal circumference. Children who could not be followed up had poorer results in the 6-minute run and the 4 x 10 m shuttle run, but no differences were found in the anthropometric data, muscle strength, flexibility, coordination, reaction speed, or balance (Table R2).

**Table R1.** Number of study participants in the individual tests.

| Variable | All | Intervention Group<br>SY21–22 | Control Group<br>SY21–22 | Sports Club | No Sports Club |
| --- | --- | --- | --- | --- | --- |
| BMI | 331 | 179 | 152 | 146 | 185 |
| WtHR | 331 | 179 | 152 | 146 | 185 |
| 6MR | 331 | 179 | 152 | 146 | 185 |
| 4×10SHR | 276 | 140 | 136 | 124 | 152 |
| SLJ | 276 | 140 | 136 | 124 | 152 |
| MB1kg | 255 | 128 | 127 | 117 | 138 |
| JS | 276 | 140 | 136 | 124 | 152 |
| VSR | 276 | 140 | 136 | 124 | 152 |
| SLS-L | 276 | 140 | 136 | 124 | 152 |
| SLS-R | 276 | 140 | 136 | 124 | 152 |
| RD | 276 | 140 | 136 | 124 | 152 |

SY21–22 = school year 2021–2022; BMI = body mass index; WtHR = waist-to-height ratio; 6MR = six-minute run; 4×10SHR = 4 x 10 m shuttle run; SLJ = standing long jump; MB1kg = medicine ball throw (1 kg); JS = jumping sideways; VSR = V sit-and-reach test; SLS-L = single leg stands left; SLS-R = single leg stands right.

**Table R2.**
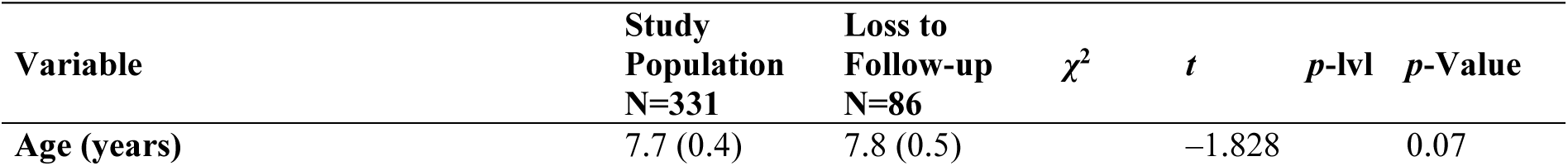
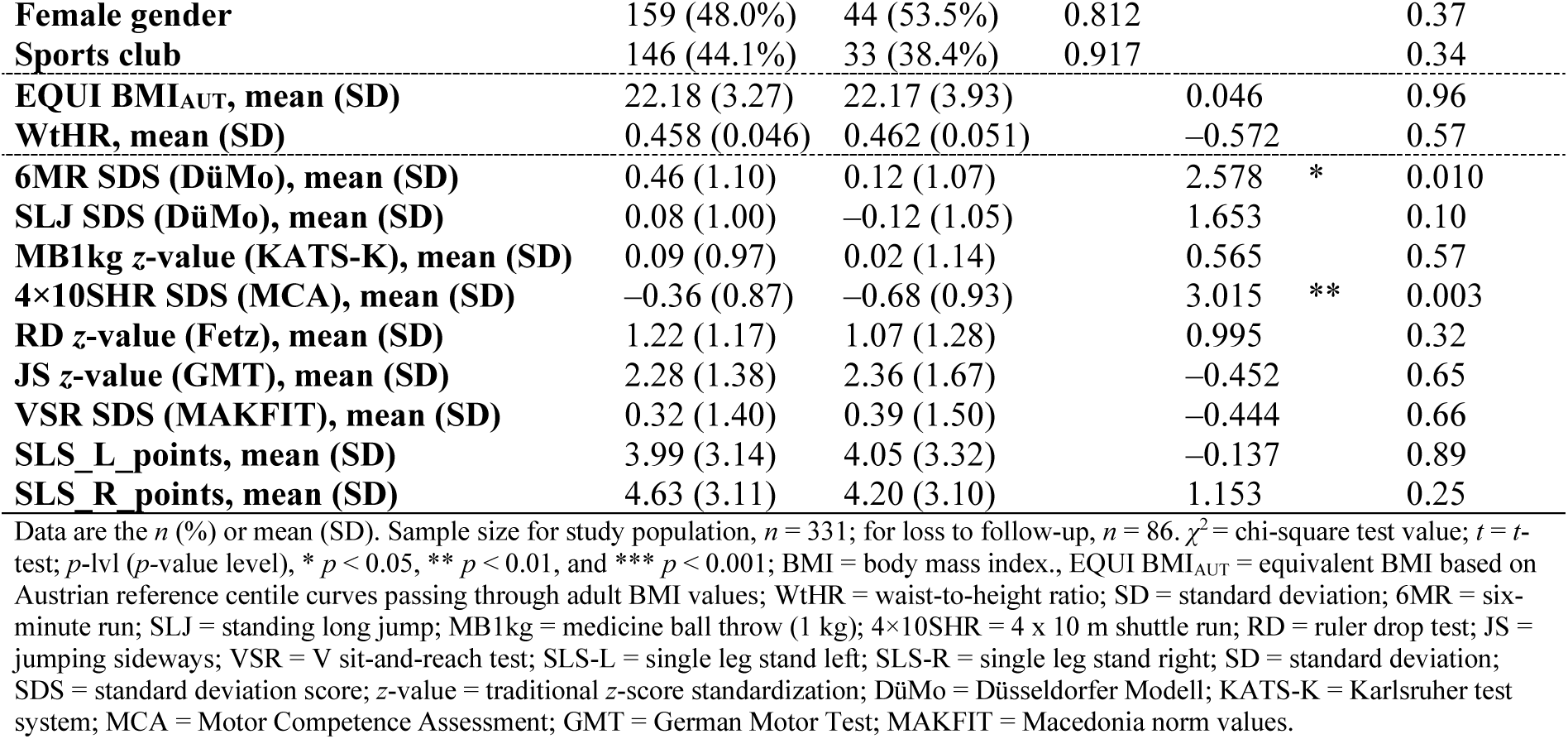
Characteristics of the total sample study population vs. loss to follow-up.

In the sample included in these analyses, the mean age at baseline was 7.7 ± 0.4 years (range = 7–9 years), 179 (54.1%) participated in a physical education (PE) intervention at school between test phases T6 and T7, and 146 (44.1%) children had a sports club membership (Table R3).

**Table R3.**
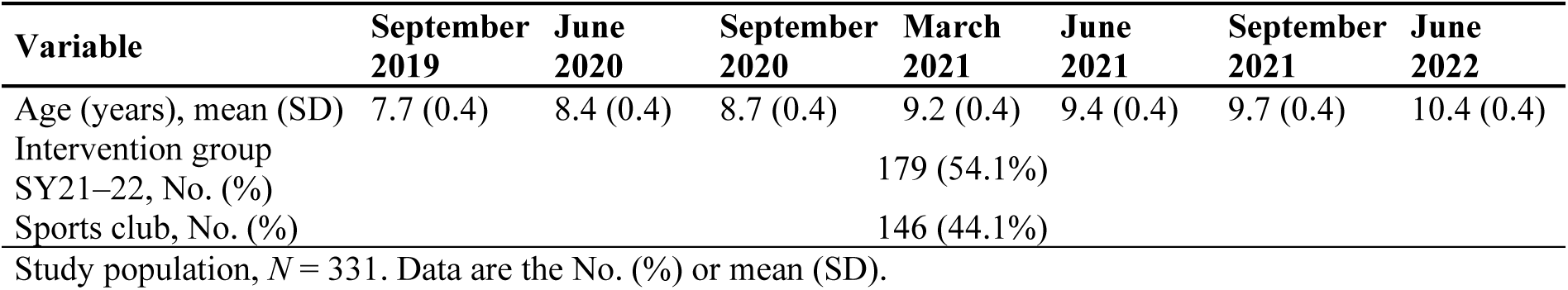
Characteristics of the study participants (anthropometric data).

The PE intervention in the last school year showed significant results in the studied population. When comparing the data from the last two test times (T6 and T7), highly significant differences (*p* < 0.001) were observed between the intervention and control groups in the assessment of health risk, with a better performance in the intervention group (Table X1). Significant differences (*p* = 0.038) were additionally found in reaction speed, with the control group showing better development of eye–hand reaction (Table X1).

Among the children not attending sports clubs, those in the intervention group showed a better and healthier trend in all health-related parameters (weight classification, health risk assessment, cardiorespiratory endurance, strength, power endurance, and action speed) compared to the children in the control group. However, this trend was significant only for health risk assessment (Table X2).

Similarly, for children with sports club membership, significant differences were found only in health risk assessment, but no overall trend was observed in the other health-related parameters, representing better or healthier outcomes in either group (intervention or control group) (Table X2).

Detailed descriptive statistics on the raw scores collected, standard deviation scores (SDS), traditional *z*-scores, and point value calculations are presented in Tables R4, R5, and R6.

**Table R4.**
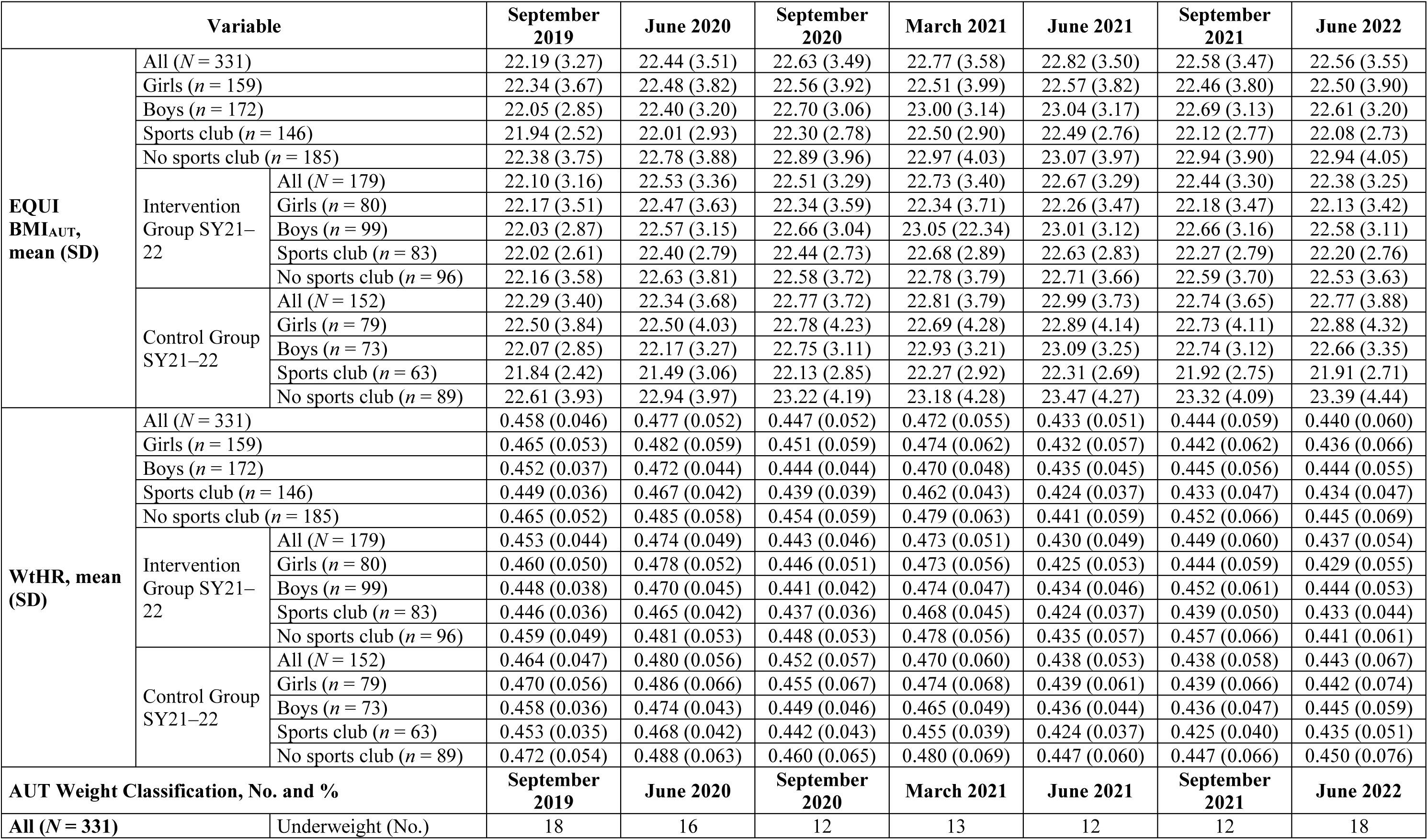
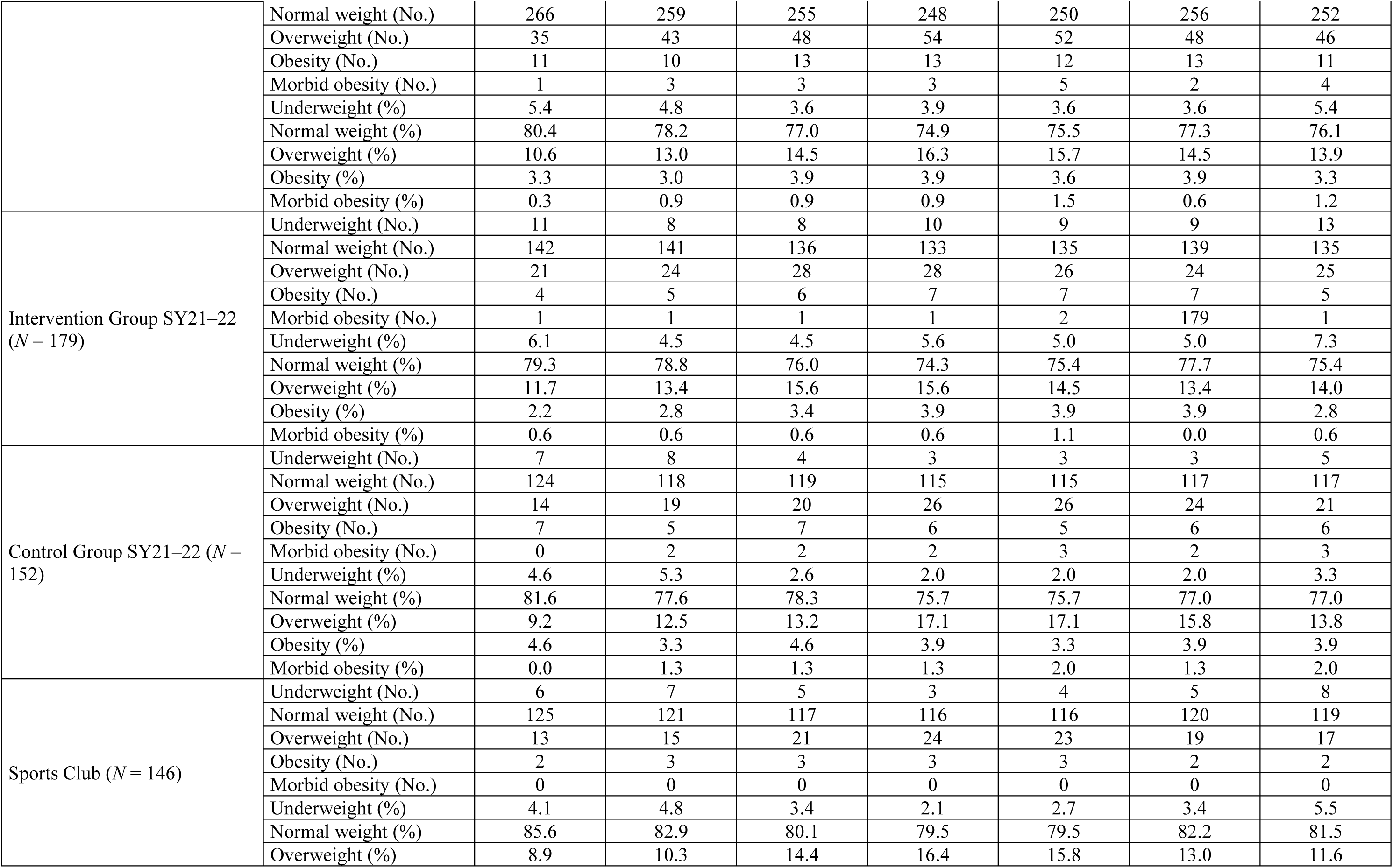
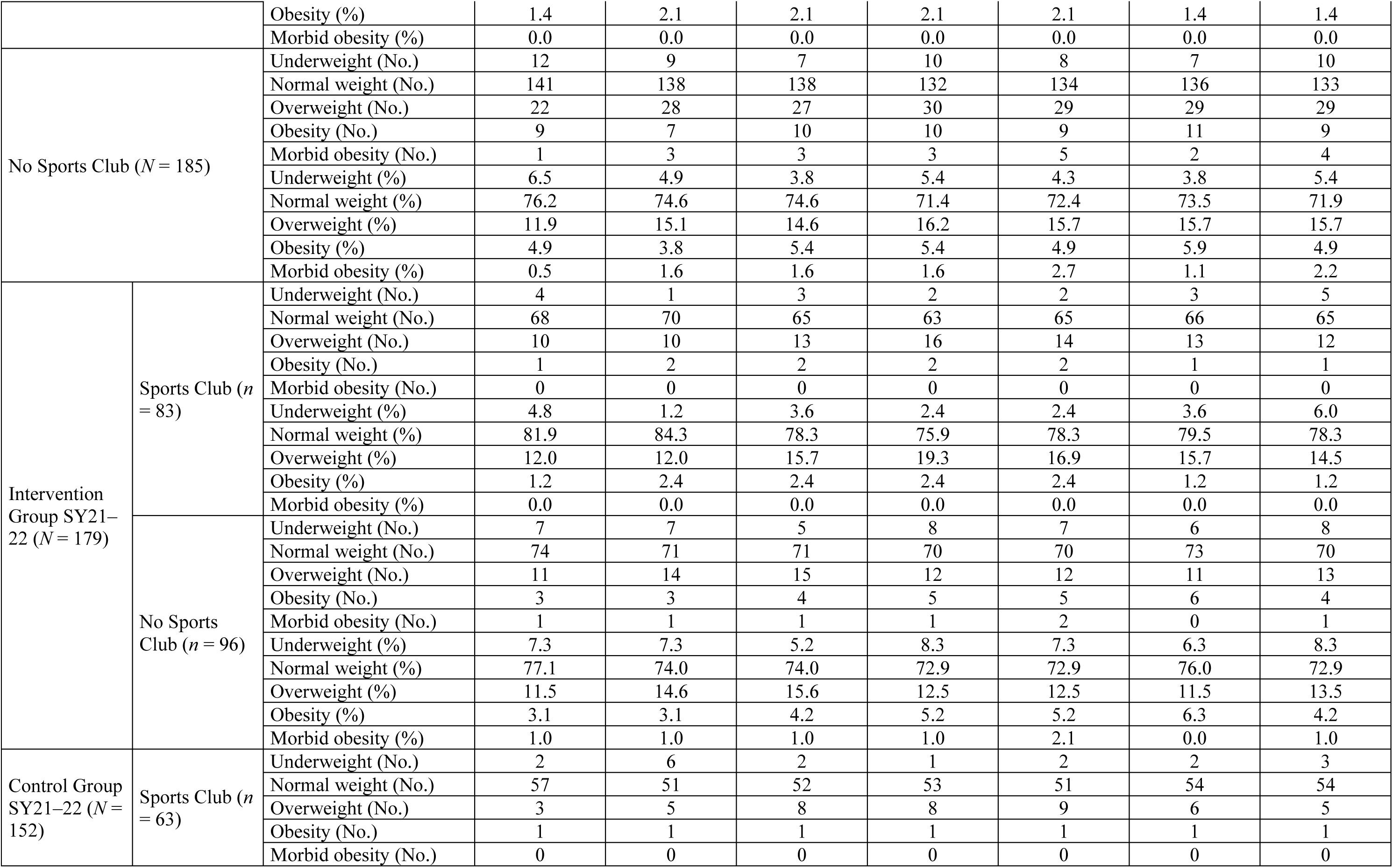
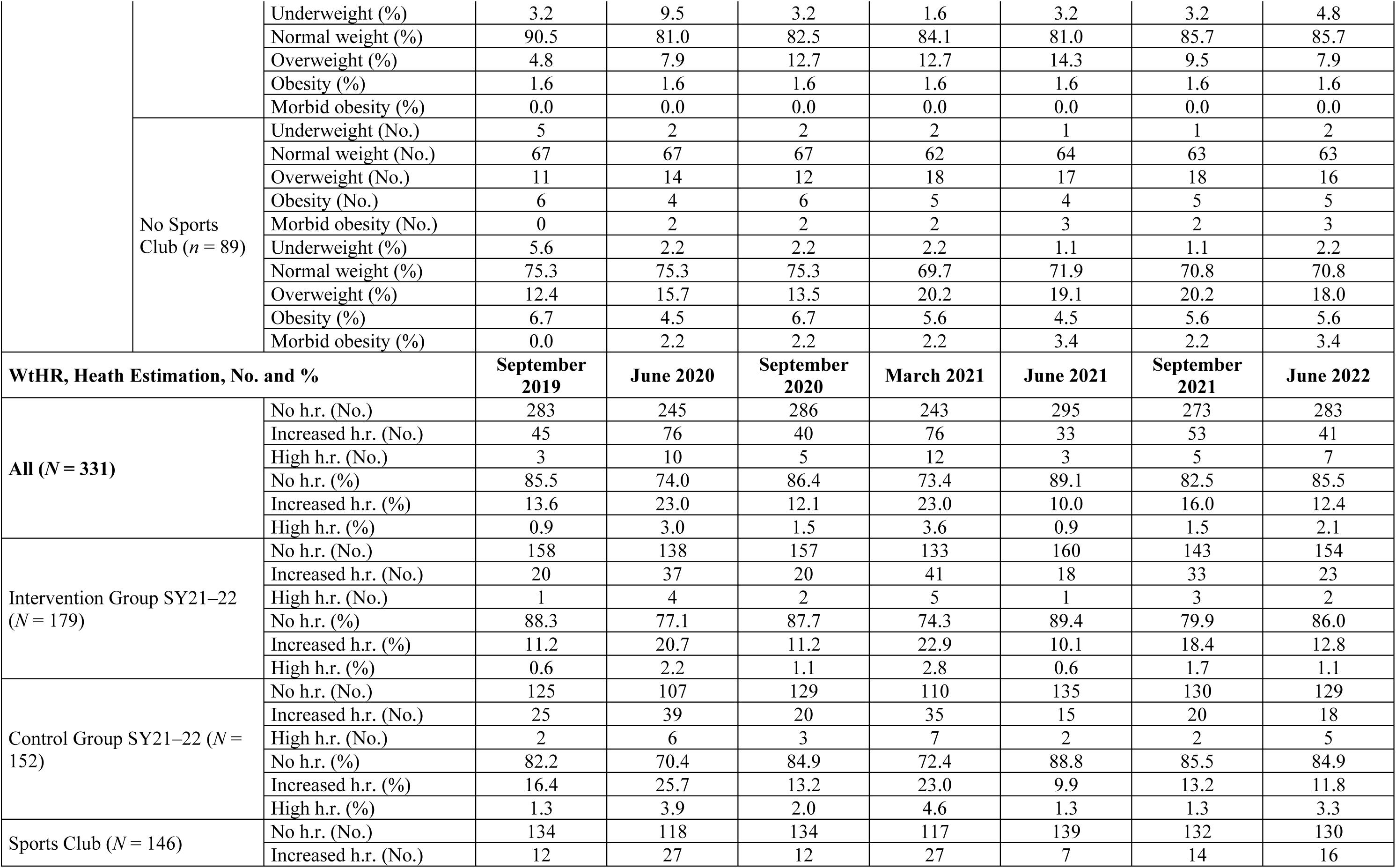
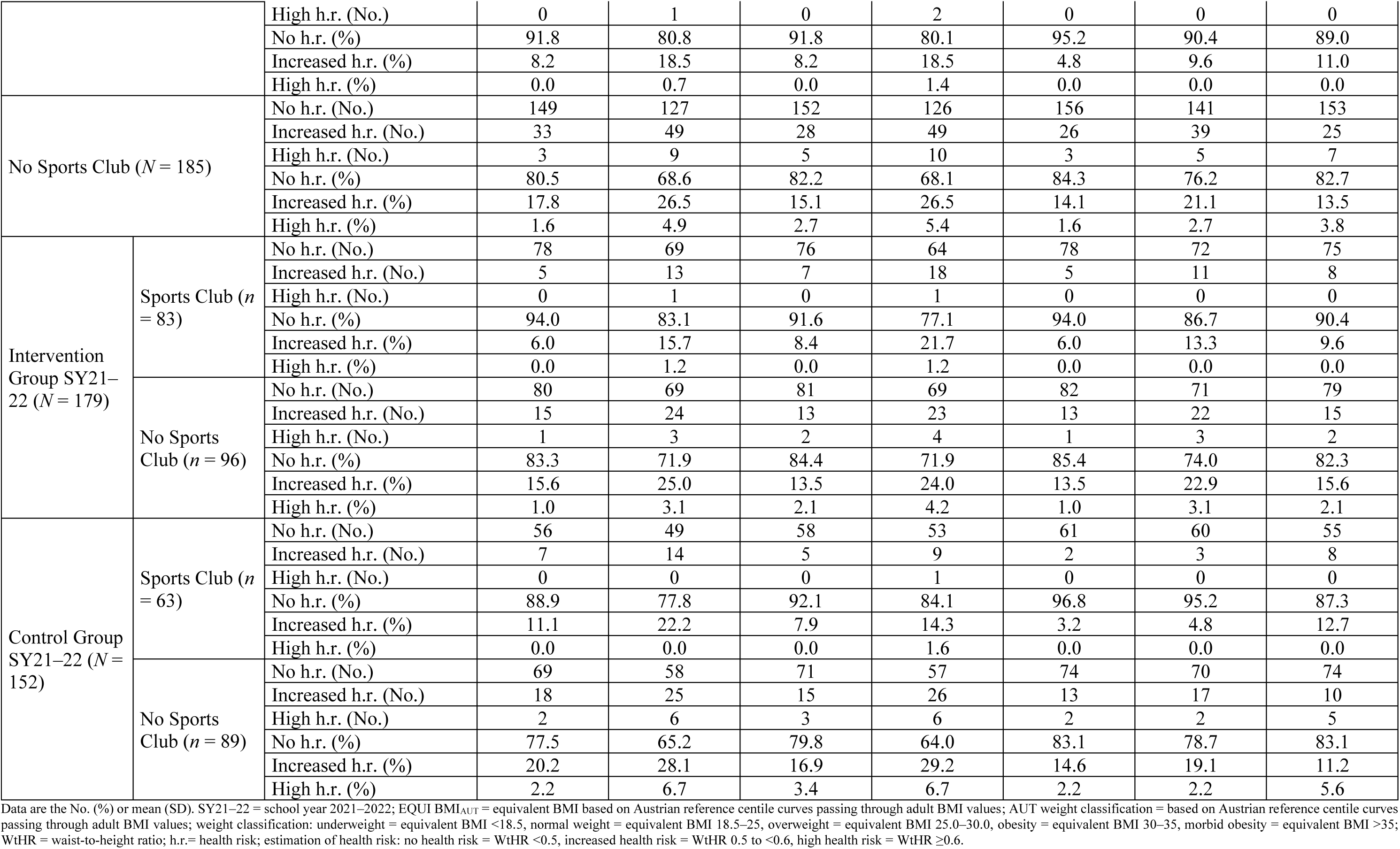
Descriptive statistics of the anthropometrics at all seven measurement time points.

**Table R5.**
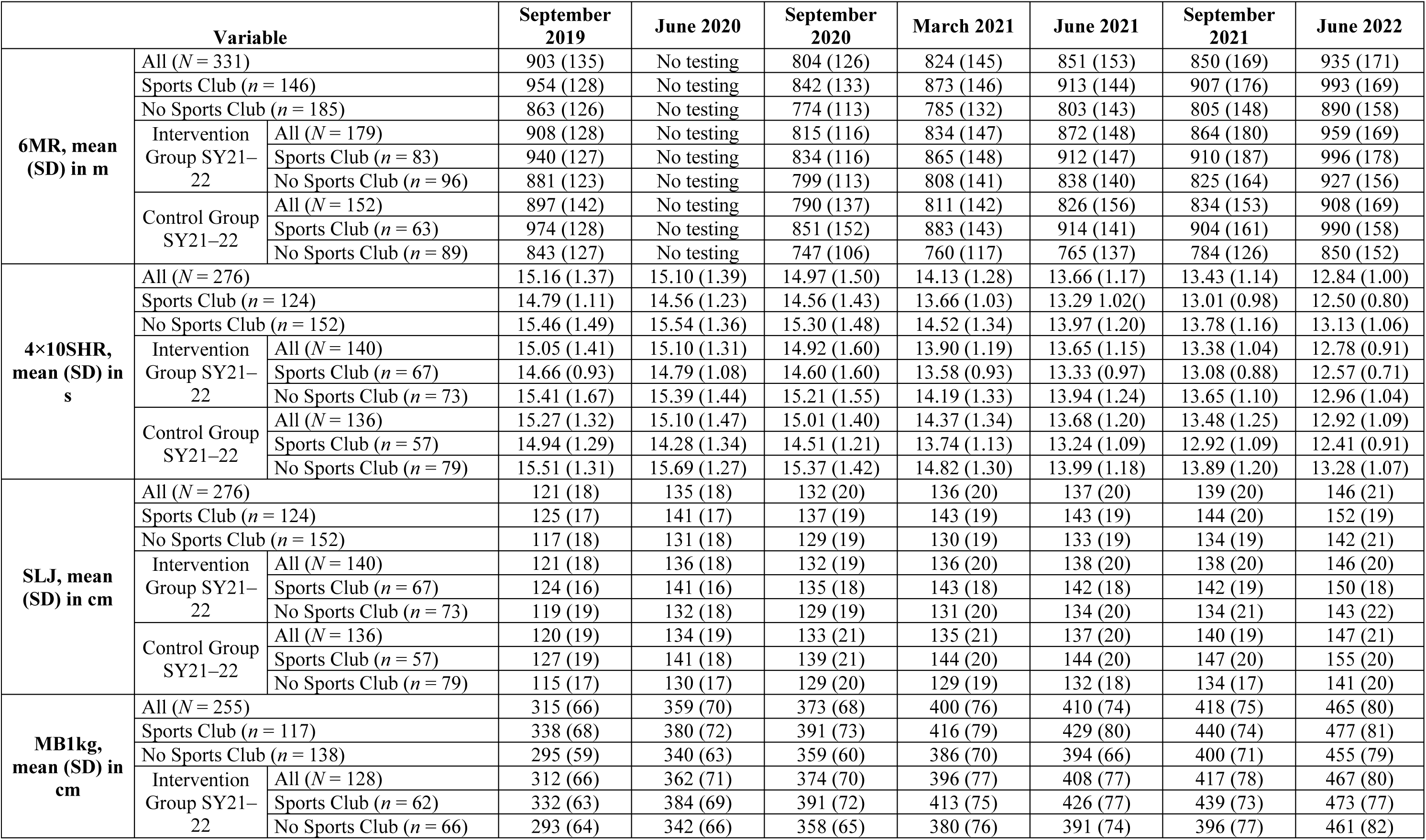
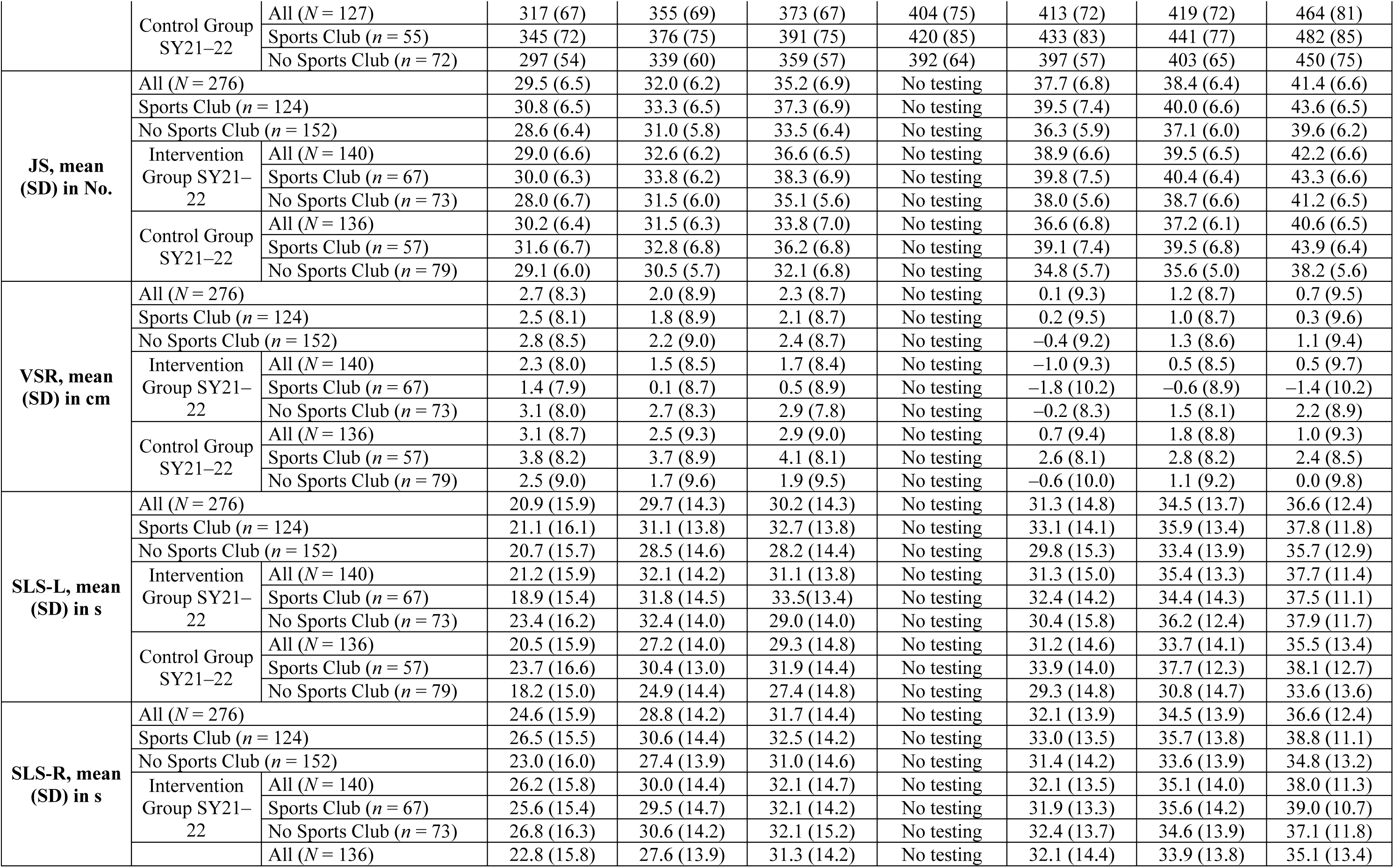
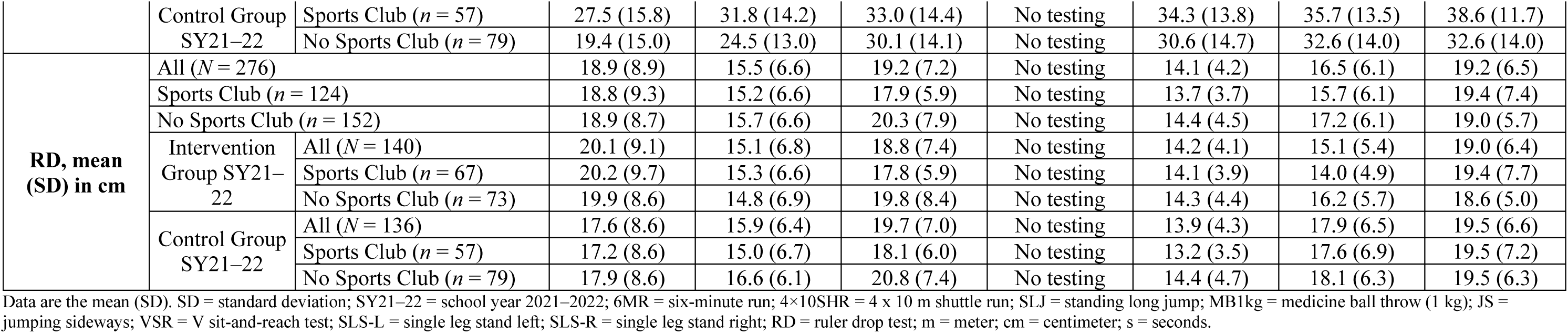
Descriptive statistics of the raw fitness data at all seven measurement time points.

**Table R6.**
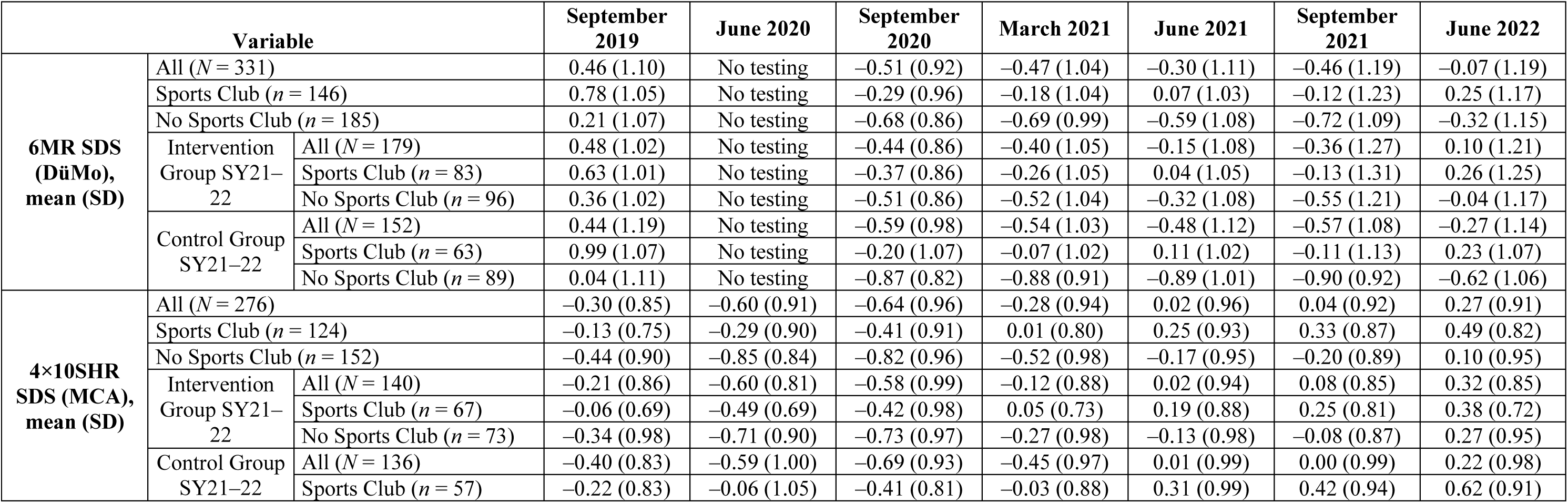
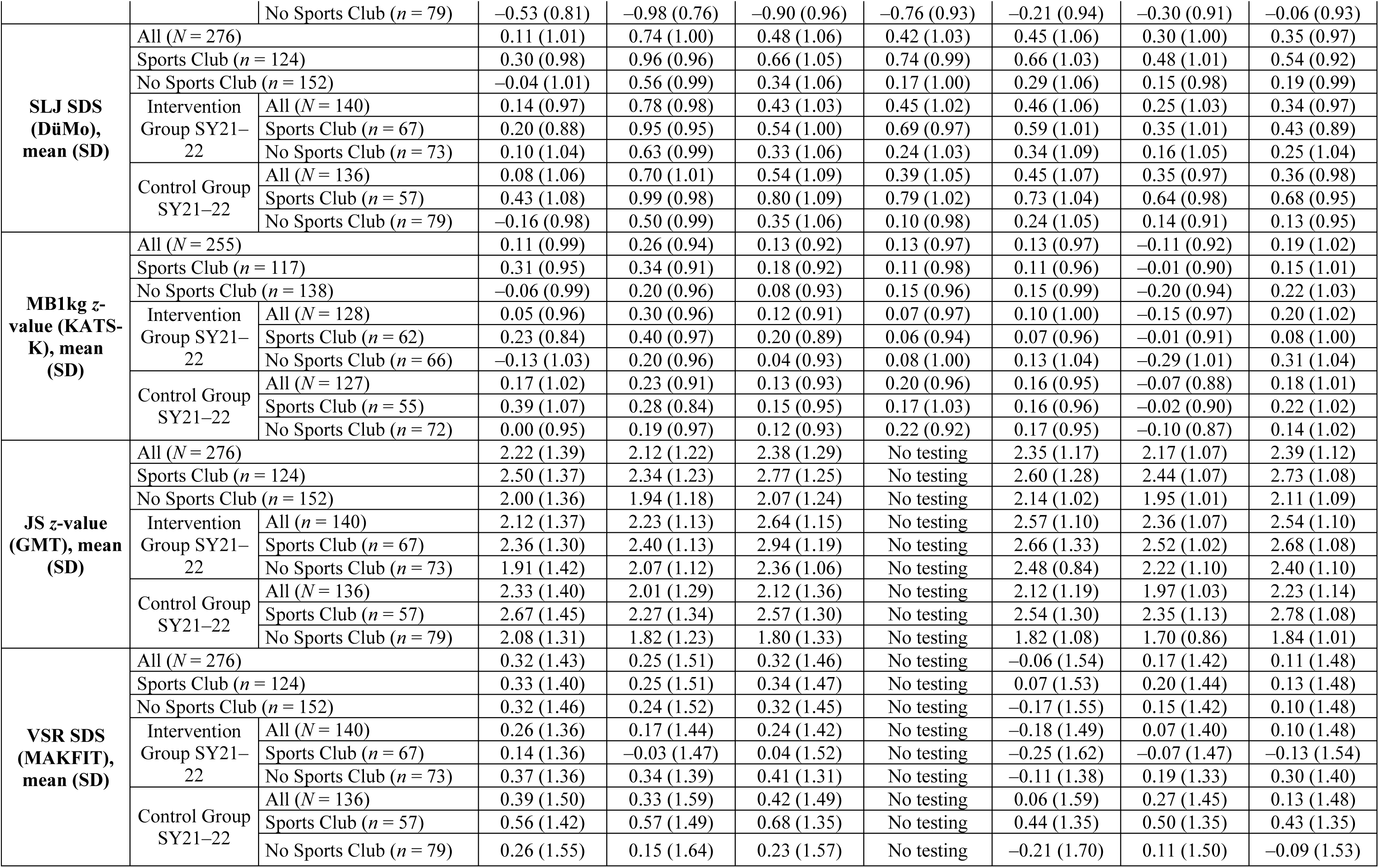
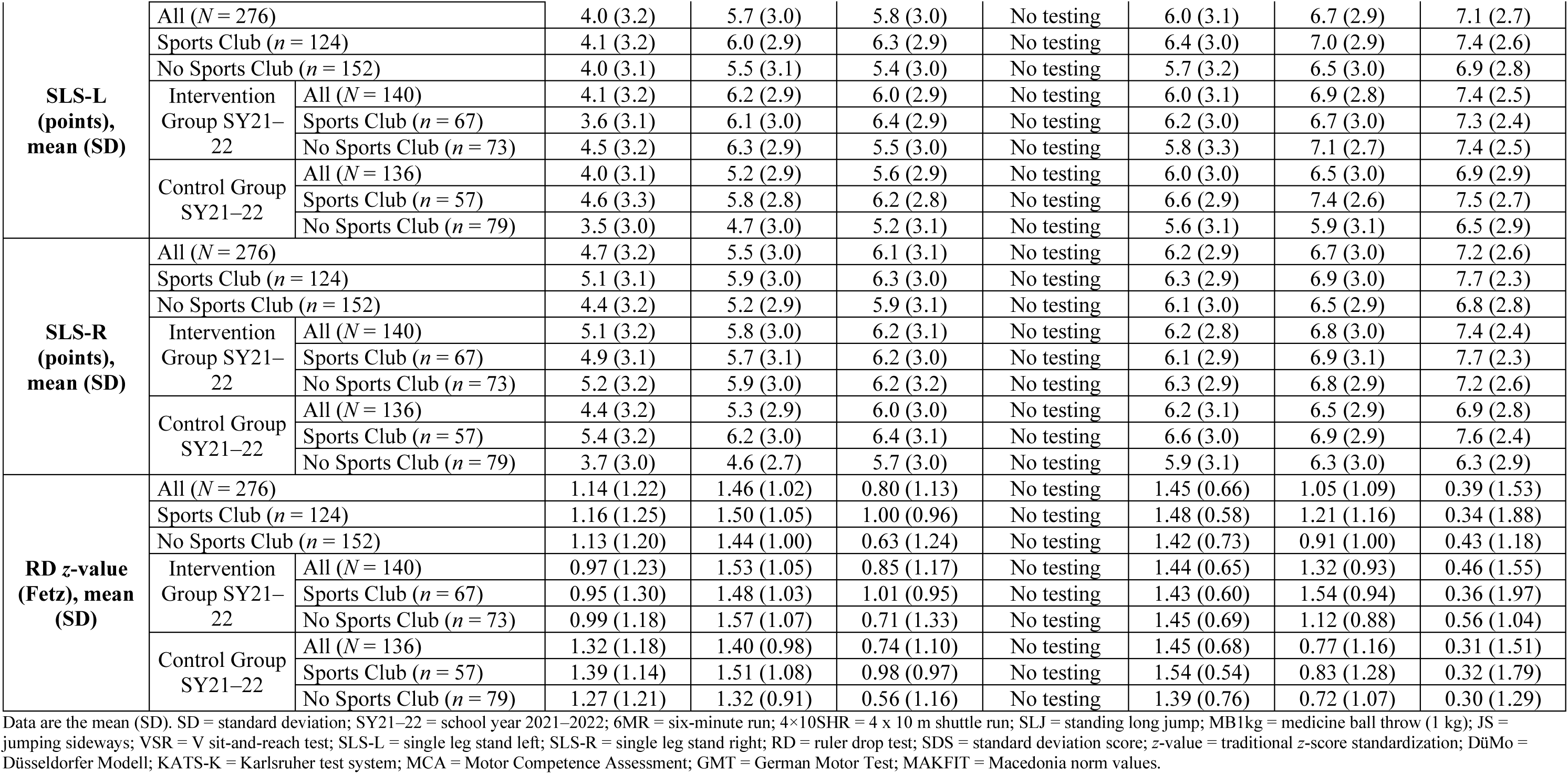
Descriptive statistics of the standard deviation scores (SDSs) and traditional *z*-value calculations for the fitness data at all seven measurement time points.

### Change of Anthropometrics

#### Changes in BMI

Between September 2019 and June 2022, the EQUI BMI_AUT_ score increased from 22.19 to 22.56 (main effect of time: ⴄ_p_^2^ = 0.057; *p* < 0.001). Those children who attended a sports club had a smaller increase in BMI than children without sports club memberships (+0.14 (95% CI, 0.56 to –0.31) versus +0.56 (95% CI, 0.19–0.96)) with small interaction effects over time (time*sports club: ⴄ_p_^2^ = 0.011; *p* = 0.008) (Tables R7 and R8 and Figure R1).

**Table R7.**
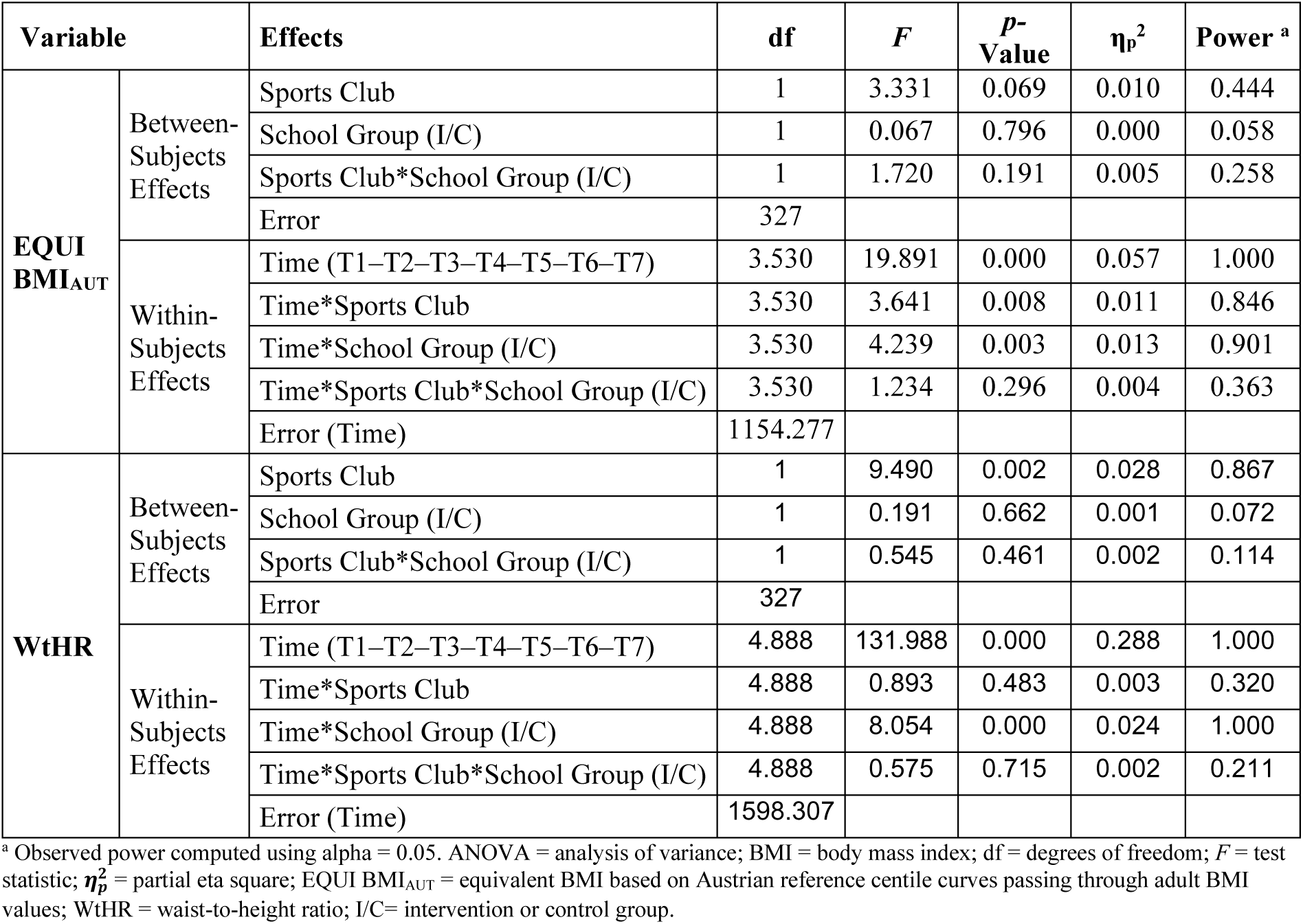
Mixed ANOVAS for the anthropometric data over all measurement time points.

**Table R8.**
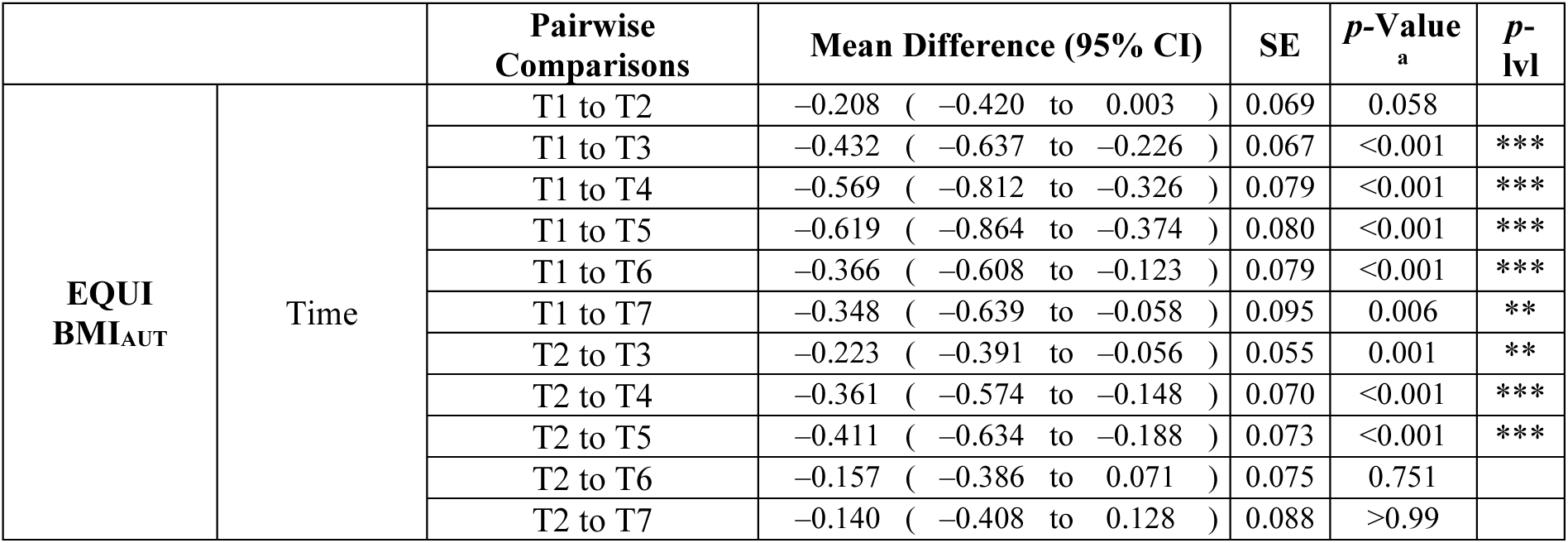
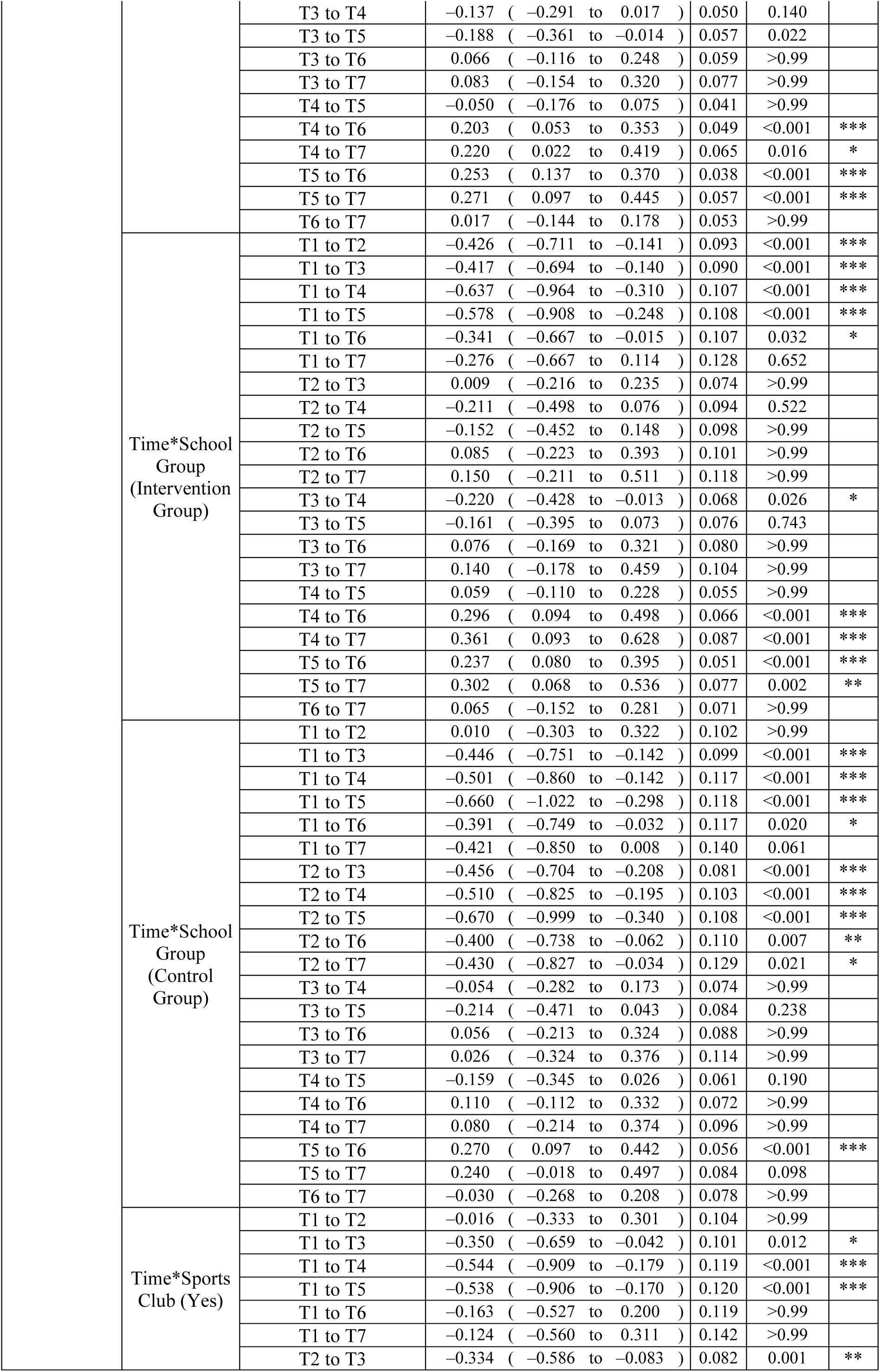
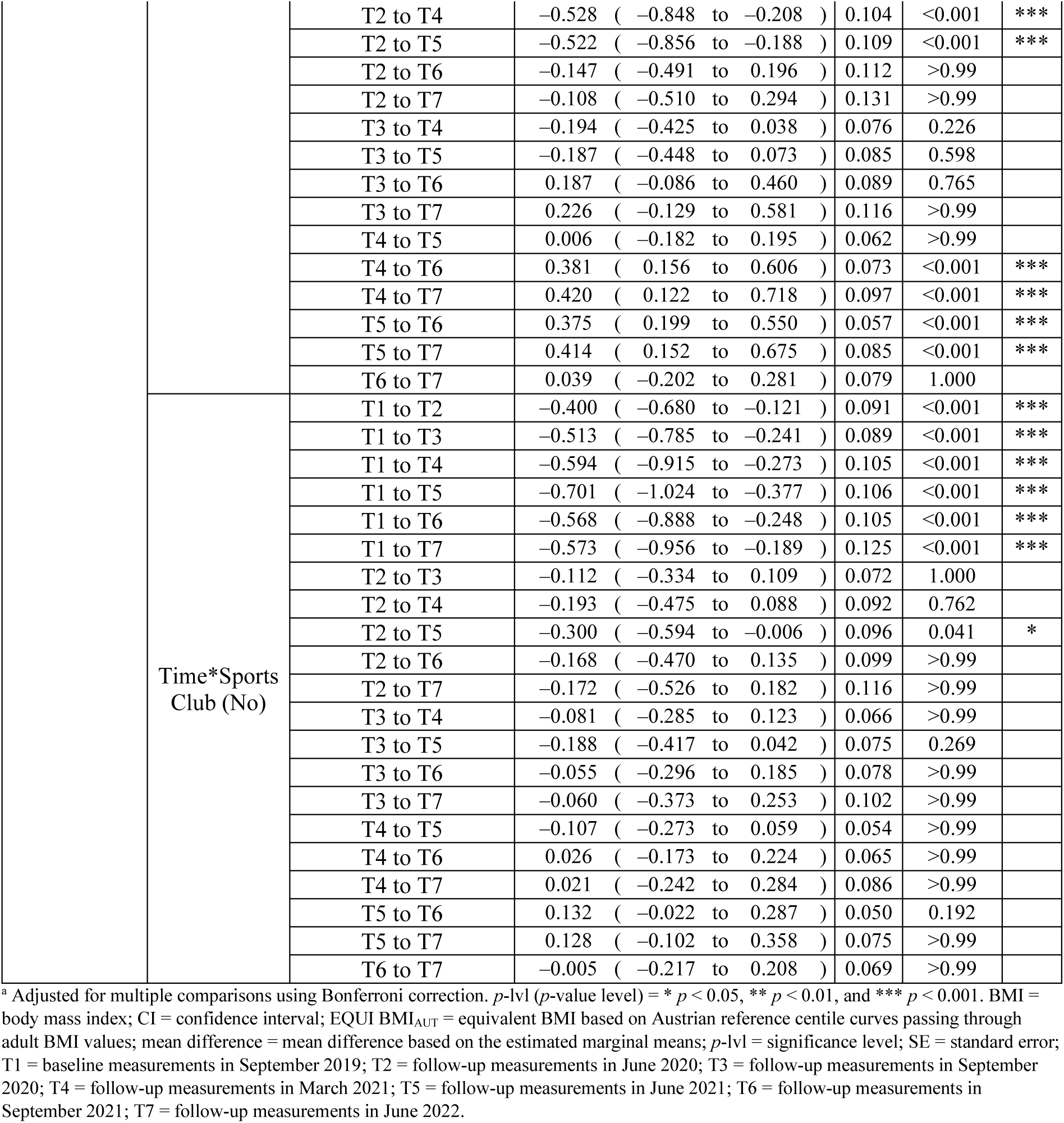
Post hoc tests of EQUI BMI_AUT_ for the main effect of time and interactions for time*school group and time*sports club based on estimated marginal means.

|  |  | Pairwise Comparisons | Mean Difference (95% CI) | SE | p-Value<sup>a</sup> | p-lvl |
| --- | --- | --- | --- | --- | --- | --- |
| EQUI BMI<sub>AUT</sub> | Time | T1 to T2 | -0.208 ( -0.420 to 0.003 ) | 0.069 | 0.058 |  |
|  |  | T1 to T3 | -0.432 ( -0.637 to -0.226 ) | 0.067 | <0.001 | *** |
|  |  | T1 to T4 | -0.569 ( -0.812 to -0.326 ) | 0.079 | <0.001 | *** |
|  |  | T1 to T5 | -0.619 ( -0.864 to -0.374 ) | 0.080 | <0.001 | *** |
|  |  | T1 to T6 | -0.366 ( -0.608 to -0.123 ) | 0.079 | <0.001 | *** |
|  |  | T1 to T7 | -0.348 ( -0.639 to -0.058 ) | 0.095 | 0.006 | ** |
|  |  | T2 to T3 | -0.223 ( -0.391 to -0.056 ) | 0.055 | 0.001 | ** |
|  |  | T2 to T4 | -0.361 ( -0.574 to -0.148 ) | 0.070 | <0.001 | *** |
|  |  | T2 to T5 | -0.411 ( -0.634 to -0.188 ) | 0.073 | <0.001 | *** |
|  |  | T2 to T6 | -0.157 ( -0.386 to 0.071 ) | 0.075 | 0.751 |  |
|  |  | T2 to T7 | -0.140 ( -0.408 to 0.128 ) | 0.088 | >0.99 |  |
|  |  | T3 to T4 | -0.137 ( -0.291 to 0.017 ) | 0.050 | 0.140 |  |
|  |  | T3 to T5 | -0.188 ( -0.361 to -0.014 ) | 0.057 | 0.022 |  |
|  |  | T3 to T6 | 0.066 ( -0.116 to 0.248 ) | 0.059 | >0.99 |  |
|  |  | T3 to T7 | 0.083 ( -0.154 to 0.320 ) | 0.077 | >0.99 |  |
|  |  | T4 to T5 | -0.050 ( -0.176 to 0.075 ) | 0.041 | >0.99 |  |
|  |  | T4 to T6 | 0.203 ( 0.053 to 0.353 ) | 0.049 | <0.001 | *** |
|  |  | T4 to T7 | 0.220 ( 0.022 to 0.419 ) | 0.065 | 0.016 | * |
|  |  | T5 to T6 | 0.253 ( 0.137 to 0.370 ) | 0.038 | <0.001 | *** |
|  |  | T5 to T7 | 0.271 ( 0.097 to 0.445 ) | 0.057 | <0.001 | *** |
|  |  | T6 to T7 | 0.017 ( -0.144 to 0.178 ) | 0.053 | >0.99 |  |
|  | Time*School<br>Group<br>(Intervention<br>Group) | T1 to T2 | -0.426 ( -0.711 to -0.141 ) | 0.093 | <0.001 | *** |
|  |  | T1 to T3 | -0.417 ( -0.694 to -0.140 ) | 0.090 | <0.001 | *** |
|  |  | T1 to T4 | -0.637 ( -0.964 to -0.310 ) | 0.107 | <0.001 | *** |
|  |  | T1 to T5 | -0.578 ( -0.908 to -0.248 ) | 0.108 | <0.001 | *** |
|  |  | T1 to T6 | -0.341 ( -0.667 to -0.015 ) | 0.107 | 0.032 | * |
|  |  | T1 to T7 | -0.276 ( -0.667 to 0.114 ) | 0.128 | 0.652 |  |
|  |  | T2 to T3 | 0.009 ( -0.216 to 0.235 ) | 0.074 | >0.99 |  |
|  |  | T2 to T4 | -0.211 ( -0.498 to 0.076 ) | 0.094 | 0.522 |  |
|  |  | T2 to T5 | -0.152 ( -0.452 to 0.148 ) | 0.098 | >0.99 |  |
|  |  | T2 to T6 | 0.085 ( -0.223 to 0.393 ) | 0.101 | >0.99 |  |
|  |  | T2 to T7 | 0.150 ( -0.211 to 0.511 ) | 0.118 | >0.99 |  |
|  |  | T3 to T4 | -0.220 ( -0.428 to -0.013 ) | 0.068 | 0.026 | * |
|  |  | T3 to T5 | -0.161 ( -0.395 to 0.073 ) | 0.076 | 0.743 |  |
|  |  | T3 to T6 | 0.076 ( -0.169 to 0.321 ) | 0.080 | >0.99 |  |
|  |  | T3 to T7 | 0.140 ( -0.178 to 0.459 ) | 0.104 | >0.99 |  |
|  |  | T4 to T5 | 0.059 ( -0.110 to 0.228 ) | 0.055 | >0.99 |  |
|  |  | T4 to T6 | 0.296 ( 0.094 to 0.498 ) | 0.066 | <0.001 | *** |
|  |  | T4 to T7 | 0.361 ( 0.093 to 0.628 ) | 0.087 | <0.001 | *** |
|  |  | T5 to T6 | 0.237 ( 0.080 to 0.395 ) | 0.051 | <0.001 | *** |
|  |  | T5 to T7 | 0.302 ( 0.068 to 0.536 ) | 0.077 | 0.002 | ** |
|  |  | T6 to T7 | 0.065 ( -0.152 to 0.281 ) | 0.071 | >0.99 |  |
|  | Time*School<br>Group<br>(Control<br>Group) | T1 to T2 | 0.010 ( -0.303 to 0.322 ) | 0.102 | >0.99 |  |
|  |  | T1 to T3 | -0.446 ( -0.751 to -0.142 ) | 0.099 | <0.001 | *** |
|  |  | T1 to T4 | -0.501 ( -0.860 to -0.142 ) | 0.117 | <0.001 | *** |
|  |  | T1 to T5 | -0.660 ( -1.022 to -0.298 ) | 0.118 | <0.001 | *** |
|  |  | T1 to T6 | -0.391 ( -0.749 to -0.032 ) | 0.117 | 0.020 | * |
|  |  | T1 to T7 | -0.421 ( -0.850 to 0.008 ) | 0.140 | 0.061 |  |
|  |  | T2 to T3 | -0.456 ( -0.704 to -0.208 ) | 0.081 | <0.001 | *** |
|  |  | T2 to T4 | -0.510 ( -0.825 to -0.195 ) | 0.103 | <0.001 | *** |
|  |  | T2 to T5 | -0.670 ( -0.999 to -0.340 ) | 0.108 | <0.001 | *** |
|  |  | T2 to T6 | -0.400 ( -0.738 to -0.062 ) | 0.110 | 0.007 | ** |
|  |  | T2 to T7 | -0.430 ( -0.827 to -0.034 ) | 0.129 | 0.021 | * |
|  |  | T3 to T4 | -0.054 ( -0.282 to 0.173 ) | 0.074 | >0.99 |  |
|  |  | T3 to T5 | -0.214 ( -0.471 to 0.043 ) | 0.084 | 0.238 |  |
|  |  | T3 to T6 | 0.056 ( -0.213 to 0.324 ) | 0.088 | >0.99 |  |
|  |  | T3 to T7 | 0.026 ( -0.324 to 0.376 ) | 0.114 | >0.99 |  |
|  |  | T4 to T5 | -0.159 ( -0.345 to 0.026 ) | 0.061 | 0.190 |  |
|  |  | T4 to T6 | 0.110 ( -0.112 to 0.332 ) | 0.072 | >0.99 |  |
|  |  | T4 to T7 | 0.080 ( -0.214 to 0.374 ) | 0.096 | >0.99 |  |
|  |  | T5 to T6 | 0.270 ( 0.097 to 0.442 ) | 0.056 | <0.001 | *** |
|  |  | T5 to T7 | 0.240 ( -0.018 to 0.497 ) | 0.084 | 0.098 |  |
|  |  | T6 to T7 | -0.030 ( -0.268 to 0.208 ) | 0.078 | >0.99 |  |
|  | Time*Sports<br>Club (Yes) | T1 to T2 | -0.016 ( -0.333 to 0.301 ) | 0.104 | >0.99 |  |
|  |  | T1 to T3 | -0.350 ( -0.659 to -0.042 ) | 0.101 | 0.012 | * |
|  |  | T1 to T4 | -0.544 ( -0.909 to -0.179 ) | 0.119 | <0.001 | *** |
|  |  | T1 to T5 | -0.538 ( -0.906 to -0.170 ) | 0.120 | <0.001 | *** |
|  |  | T1 to T6 | -0.163 ( -0.527 to 0.200 ) | 0.119 | >0.99 |  |
|  |  | T1 to T7 | -0.124 ( -0.560 to 0.311 ) | 0.142 | >0.99 |  |
|  |  | T2 to T3 | -0.334 ( -0.586 to -0.083 ) | 0.082 | 0.001 | ** |
|  |  | T2 to T4 | -0.528 ( -0.848 to -0.208 ) | 0.104 | <0.001 | *** |
|  |  | T2 to T5 | -0.522 ( -0.856 to -0.188 ) | 0.109 | <0.001 | *** |
|  |  | T2 to T6 | -0.147 ( -0.491 to 0.196 ) | 0.112 | >0.99 |  |
|  |  | T2 to T7 | -0.108 ( -0.510 to 0.294 ) | 0.131 | >0.99 |  |
|  |  | T3 to T4 | -0.194 ( -0.425 to 0.038 ) | 0.076 | 0.226 |  |
|  |  | T3 to T5 | -0.187 ( -0.448 to 0.073 ) | 0.085 | 0.598 |  |
|  |  | T3 to T6 | 0.187 ( -0.086 to 0.460 ) | 0.089 | 0.765 |  |
|  |  | T3 to T7 | 0.226 ( -0.129 to 0.581 ) | 0.116 | >0.99 |  |
|  |  | T4 to T5 | 0.006 ( -0.182 to 0.195 ) | 0.062 | >0.99 |  |
|  |  | T4 to T6 | 0.381 ( 0.156 to 0.606 ) | 0.073 | <0.001 | *** |
|  |  | T4 to T7 | 0.420 ( 0.122 to 0.718 ) | 0.097 | <0.001 | *** |
|  |  | T5 to T6 | 0.375 ( 0.199 to 0.550 ) | 0.057 | <0.001 | *** |
|  |  | T5 to T7 | 0.414 ( 0.152 to 0.675 ) | 0.085 | <0.001 | *** |
|  |  | T6 to T7 | 0.039 ( -0.202 to 0.281 ) | 0.079 | 1.000 |  |
|  | Time*Sports Club (No) | T1 to T2 | -0.400 ( -0.680 to -0.121 ) | 0.091 | <0.001 | *** |
|  |  | T1 to T3 | -0.513 ( -0.785 to -0.241 ) | 0.089 | <0.001 | *** |
|  |  | T1 to T4 | -0.594 ( -0.915 to -0.273 ) | 0.105 | <0.001 | *** |
|  |  | T1 to T5 | -0.701 ( -1.024 to -0.377 ) | 0.106 | <0.001 | *** |
|  |  | T1 to T6 | -0.568 ( -0.888 to -0.248 ) | 0.105 | <0.001 | *** |
|  |  | T1 to T7 | -0.573 ( -0.956 to -0.189 ) | 0.125 | <0.001 | *** |
|  |  | T2 to T3 | -0.112 ( -0.334 to 0.109 ) | 0.072 | 1.000 |  |
|  |  | T2 to T4 | -0.193 ( -0.475 to 0.088 ) | 0.092 | 0.762 |  |
|  |  | T2 to T5 | -0.300 ( -0.594 to -0.006 ) | 0.096 | 0.041 | * |
|  |  | T2 to T6 | -0.168 ( -0.470 to 0.135 ) | 0.099 | >0.99 |  |
|  |  | T2 to T7 | -0.172 ( -0.526 to 0.182 ) | 0.116 | >0.99 |  |
|  |  | T3 to T4 | -0.081 ( -0.285 to 0.123 ) | 0.066 | >0.99 |  |
|  |  | T3 to T5 | -0.188 ( -0.417 to 0.042 ) | 0.075 | 0.269 |  |
|  |  | T3 to T6 | -0.055 ( -0.296 to 0.185 ) | 0.078 | >0.99 |  |
|  |  | T3 to T7 | -0.060 ( -0.373 to 0.253 ) | 0.102 | >0.99 |  |
|  |  | T4 to T5 | -0.107 ( -0.273 to 0.059 ) | 0.054 | >0.99 |  |
|  |  | T4 to T6 | 0.026 ( -0.173 to 0.224 ) | 0.065 | >0.99 |  |
|  |  | T4 to T7 | 0.021 ( -0.242 to 0.284 ) | 0.086 | >0.99 |  |
|  |  | T5 to T6 | 0.132 ( -0.022 to 0.287 ) | 0.050 | 0.192 |  |
|  |  | T5 to T7 | 0.128 ( -0.102 to 0.358 ) | 0.075 | >0.99 |  |
|  |  | T6 to T7 | -0.005 ( -0.217 to 0.208 ) | 0.069 | >0.99 |  |

<sup>a</sup> Adjusted for multiple comparisons using Bonferroni correction. $p$ -lvl ( $p$ -value level) = \* $p < 0.05$ , \*\* $p < 0.01$ , and \*\*\* $p < 0.001$ . BMI = body mass index; CI = confidence interval; EQUI BMI<sub>AUT</sub> = equivalent BMI based on Austrian reference centile curves passing through adult BMI values; mean difference = mean difference based on the estimated marginal means; $p$ -lvl = significance level; SE = standard error; T1 = baseline measurements in September 2019; T2 = follow-up measurements in June 2020; T3 = follow-up measurements in September 2020; T4 = follow-up measurements in March 2021; T5 = follow-up measurements in June 2021; T6 = follow-up measurements in September 2021; T7 = follow-up measurements in June 2022.

**Figure R1.**
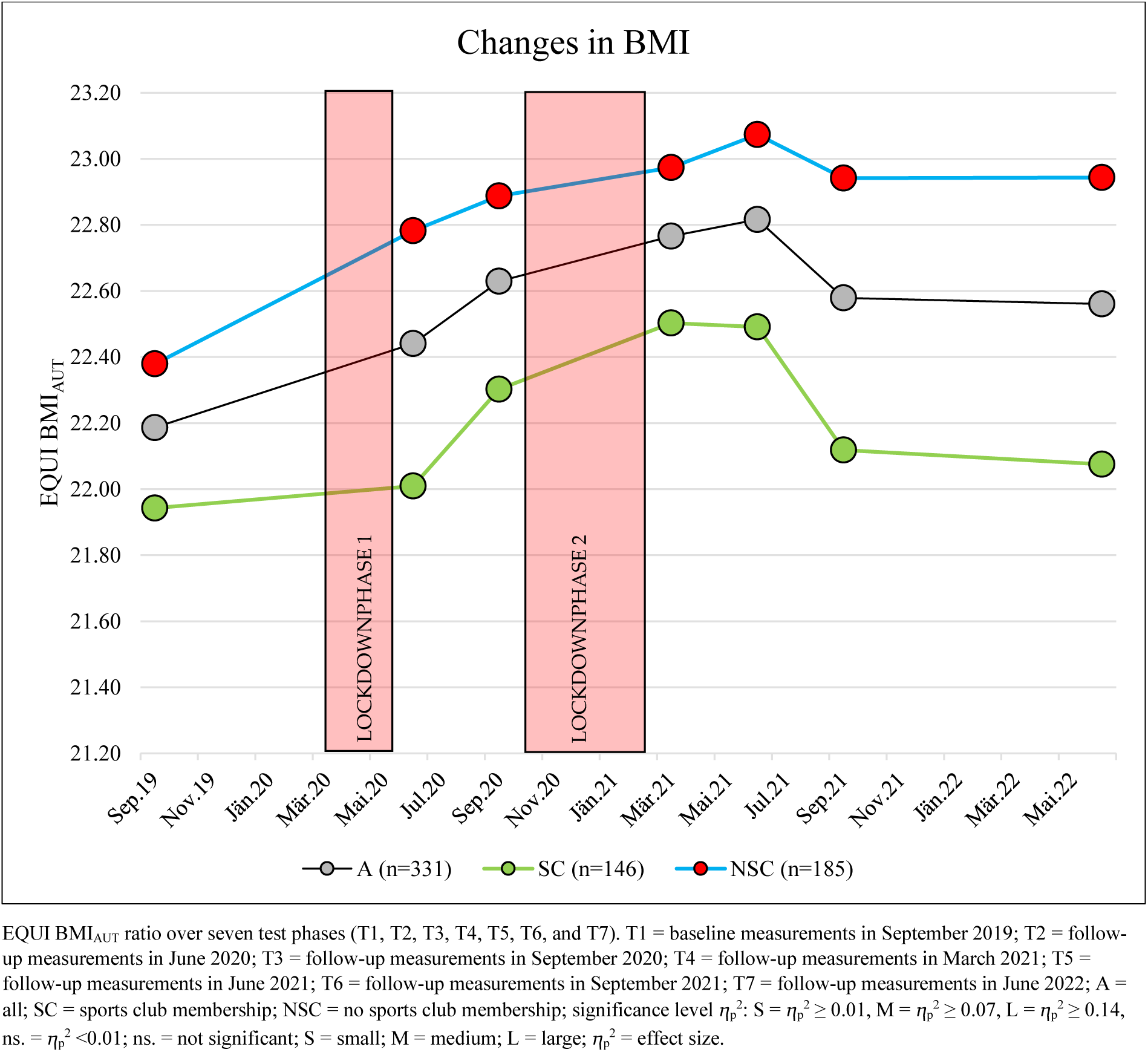
Changes in body mass index from September 2019 to June 2022.

The highest value compared to the baseline level in September 2019 was observed in the EQUI BMI_AUT_ in June 2021 with 22.82; at this point, an increase of +0.63 (95% CI, 0.37– 0.86) was found. Between June 2021 and June 2022, the EQUI BMI_AUT_ showed a significant improvement of –0.26 (95% CI, –0.10 to –0.45), and this trend was observed in all subgroups (Table R8 and Figure R1).

Changes in BMI in the school groups: (subgroup: intervention and control group SY21–22) Small interaction effects over time (time*school group: ⴄ_p_^2^ = 0.013; *p* = 0.003) were found, which were mainly related to the time before the sports intervention (September 2019 to September 2021). No interaction effects over time (time*sports club*school group: ⴄ_p_^2^ = 0.004; *p* = 0.30) and no significant changes were found in the intervention or control groups during the observation period from September 2021 to June 2022 (I-A = –0.06 (95% CI, – 0.28 to 0.15); *p* > 0.99; C-A = 0.07 (95% CI, –0.21 to 0.27); *p* > 0.99) (Table R7 and R8 and Figure R2).

**Figure R2.**
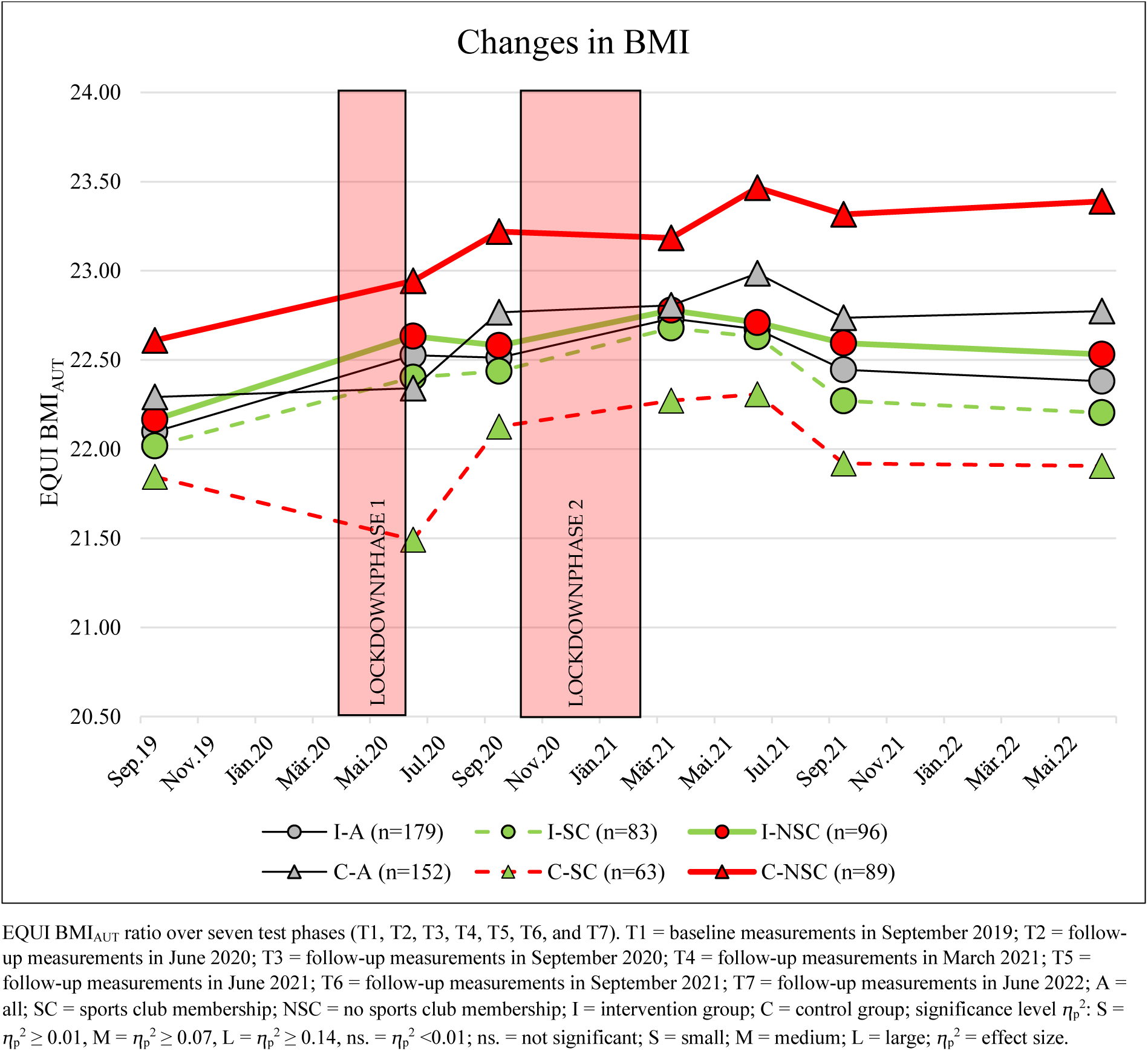
Changes in BMI from September 2019 to June 2022 in the subgroups: Intervention and control groups divided by sports club membership.

#### Changes in Waist-to-Height Ratio (WtHR)

Between September 2019 and June 2022, the WtHR scores changed significantly with large effects between test phases (main effect of time: ⴄ_p_^2^ = 0.288; *p* < 0.001). No differences were found between those children with a sports club membership and those without (Tables R7 and Table R9 and Figure R3).

**Table R9.**
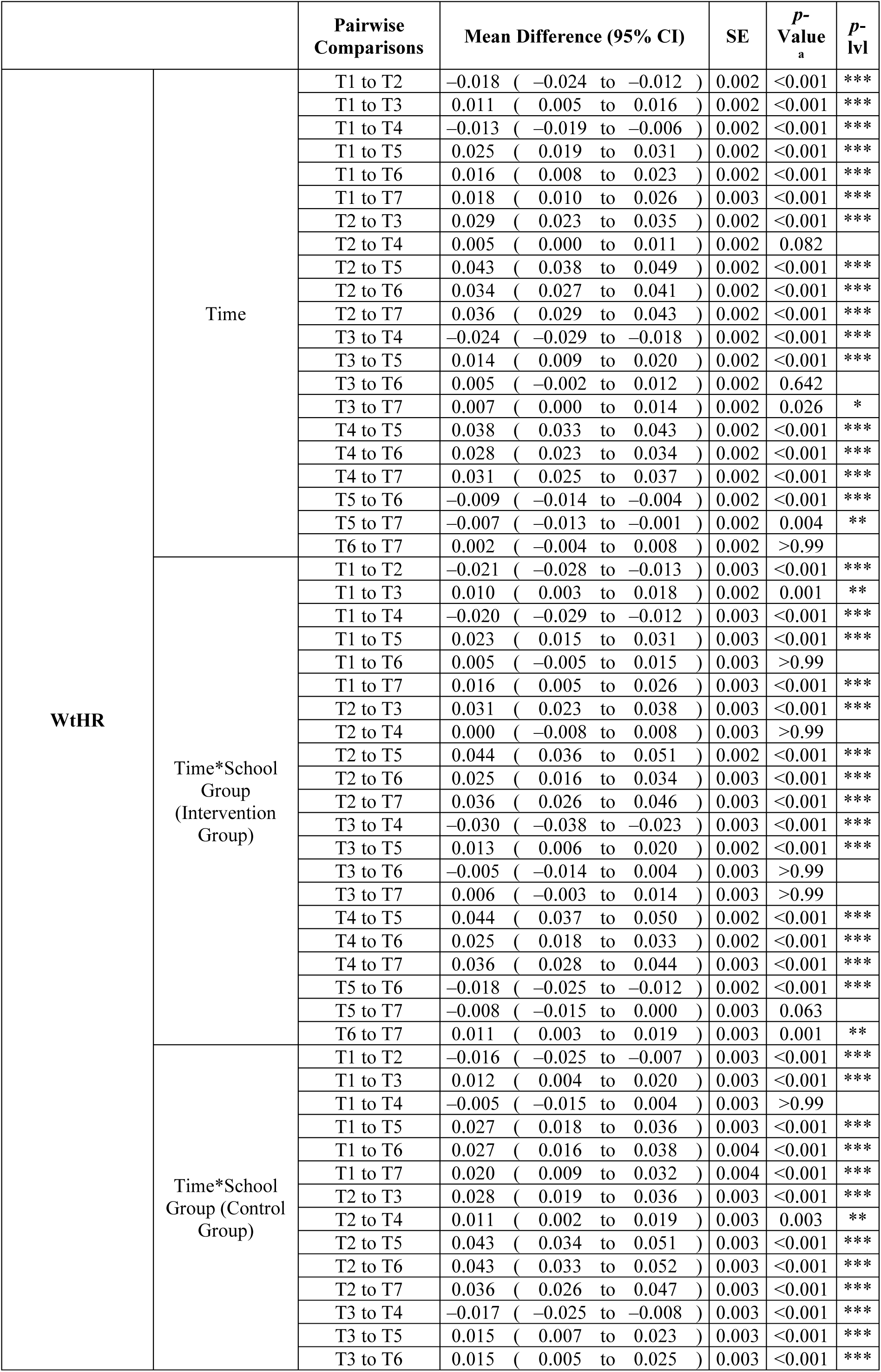
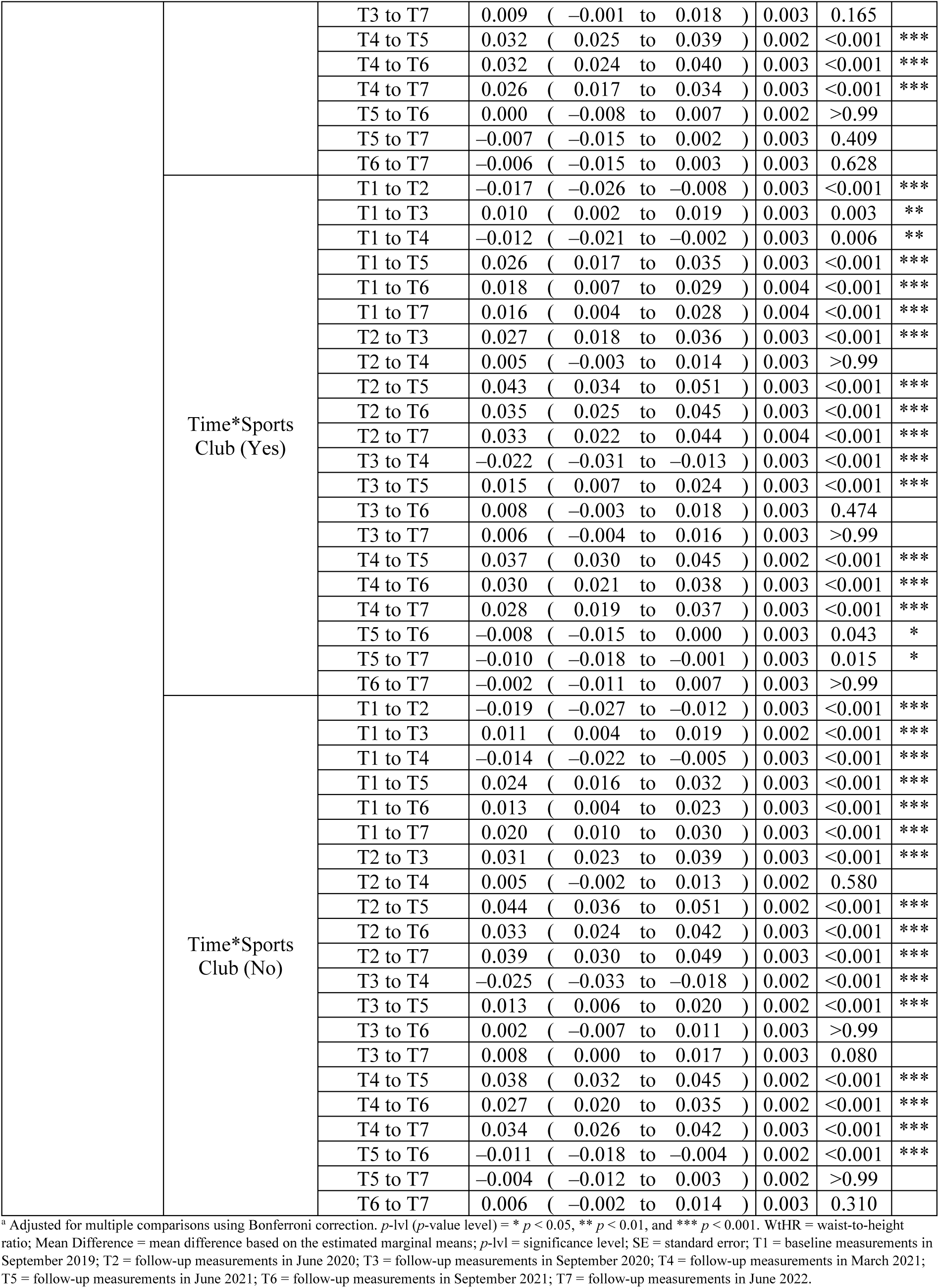
Post hoc tests of the waist-to-height ratio (WtHR) for the main effect of time and interactions for time*school group and time*sports club membership based on estimated marginal means.

**Figure R3.**
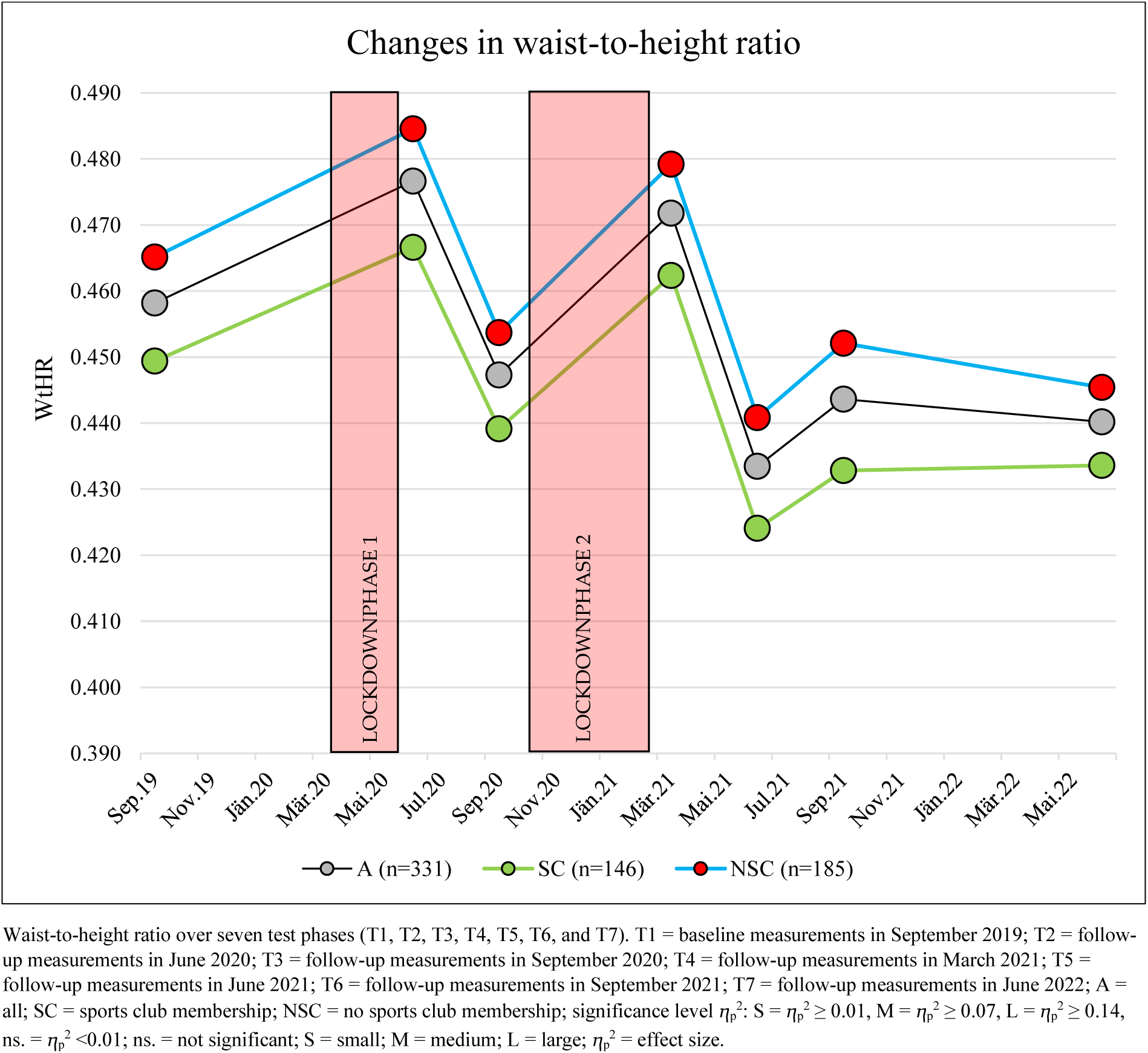
Changes in the waist-to-height ratio from September 2019 to June 2022.

The WtHR increased significantly in each of the lockdown phases (between T1 and T2 or T3 and T4) and then decreased significantly in the phases following the stringent COVID-19 mitigation measures (between T2 and T3 or after T4) (Table R9 and Figure R3).

During the last observation period (September 2021 to June 2022), there was no significant change in the WtHR (*p* > 0.99) (Table R9 and Figure R3).

Changes in the WtHR in the school groups: (subgroup: intervention and control group SY21– 22)

Small interaction effects were found over time (time*school group: ⴄ_p_^2^ = 0.024; *p* < 0.001), which were mainly related to the time before the sports intervention (September 2019 to September 2021). No interaction effects were found over time (time*sports club*school group: ⴄ_p_^2^ = 0.002; *p* = 0.72). During the observation period from September 2021 to June 2022, significant changes were found in the intervention group (I-A = –0.012 (95% CI, –0.019 to –0.003); *p* = 0.003), while the control group showed no significant changes in this time period (C-A = 0.005 (95% CI, –0.003 to 0.015); *p* = 0.63) (Table R9 and Figure R4).

**Figure R4.**
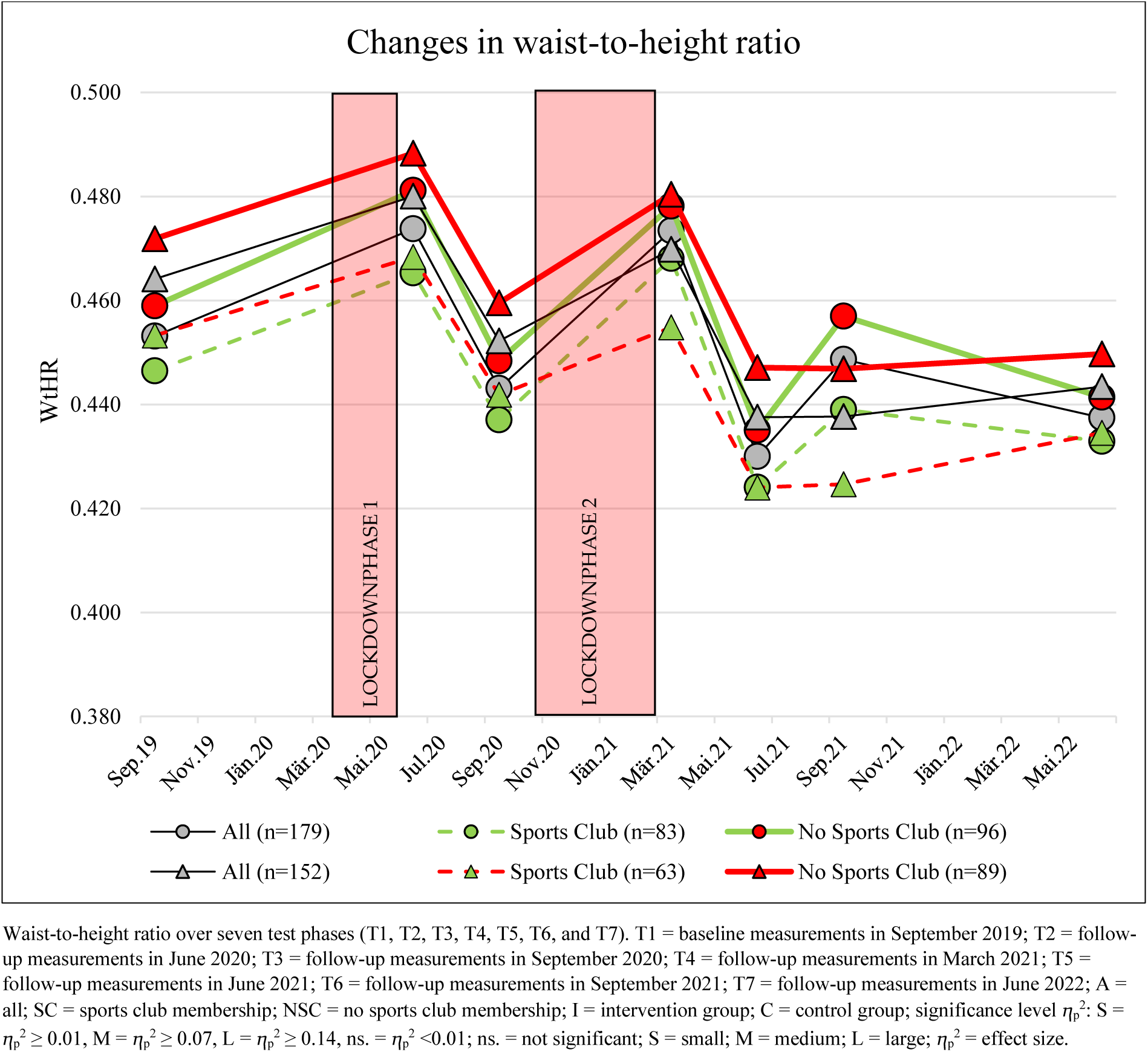
Changes in the waist-to-height ratio from September 2019 to June 2022 in the subgroups: Intervention and control groups divided by sports club membership.

These changes were also found in all subgroups of the intervention and control groups (Table R9 and Figure R4).

### Changes in Fitness

Over the overall observation period, significant changes, with different levels of effects (related to the main effect—time) were found in the area of health- and performance-related parameters (Table R10 and Figure R5).

**Table R10.**
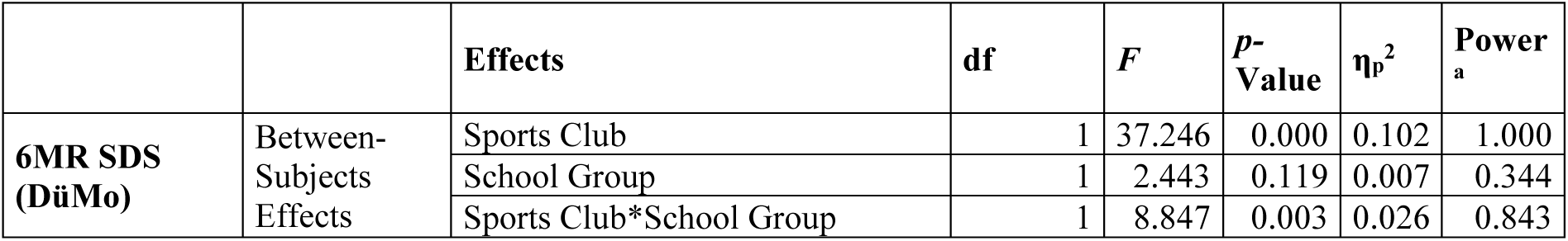
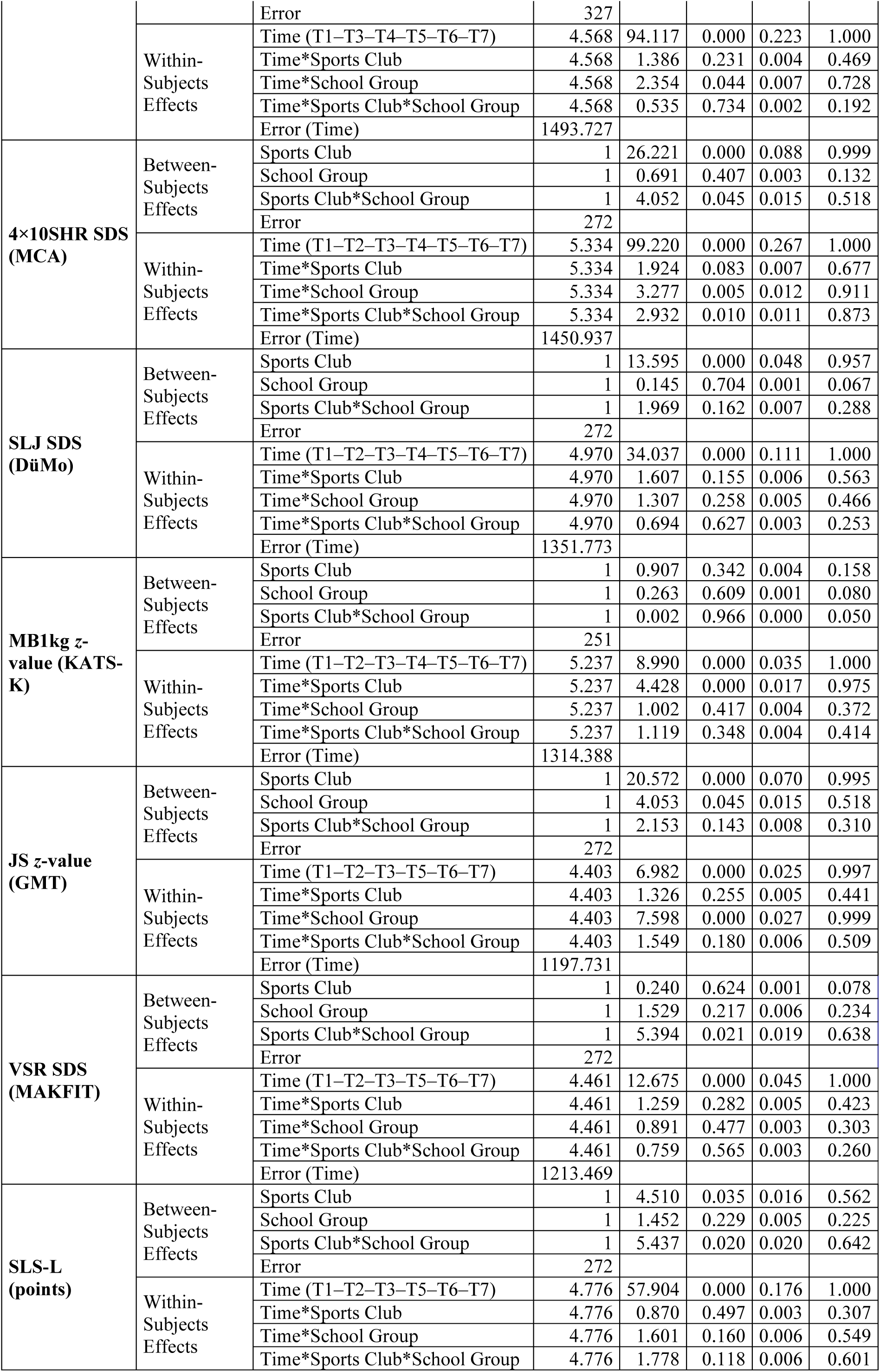
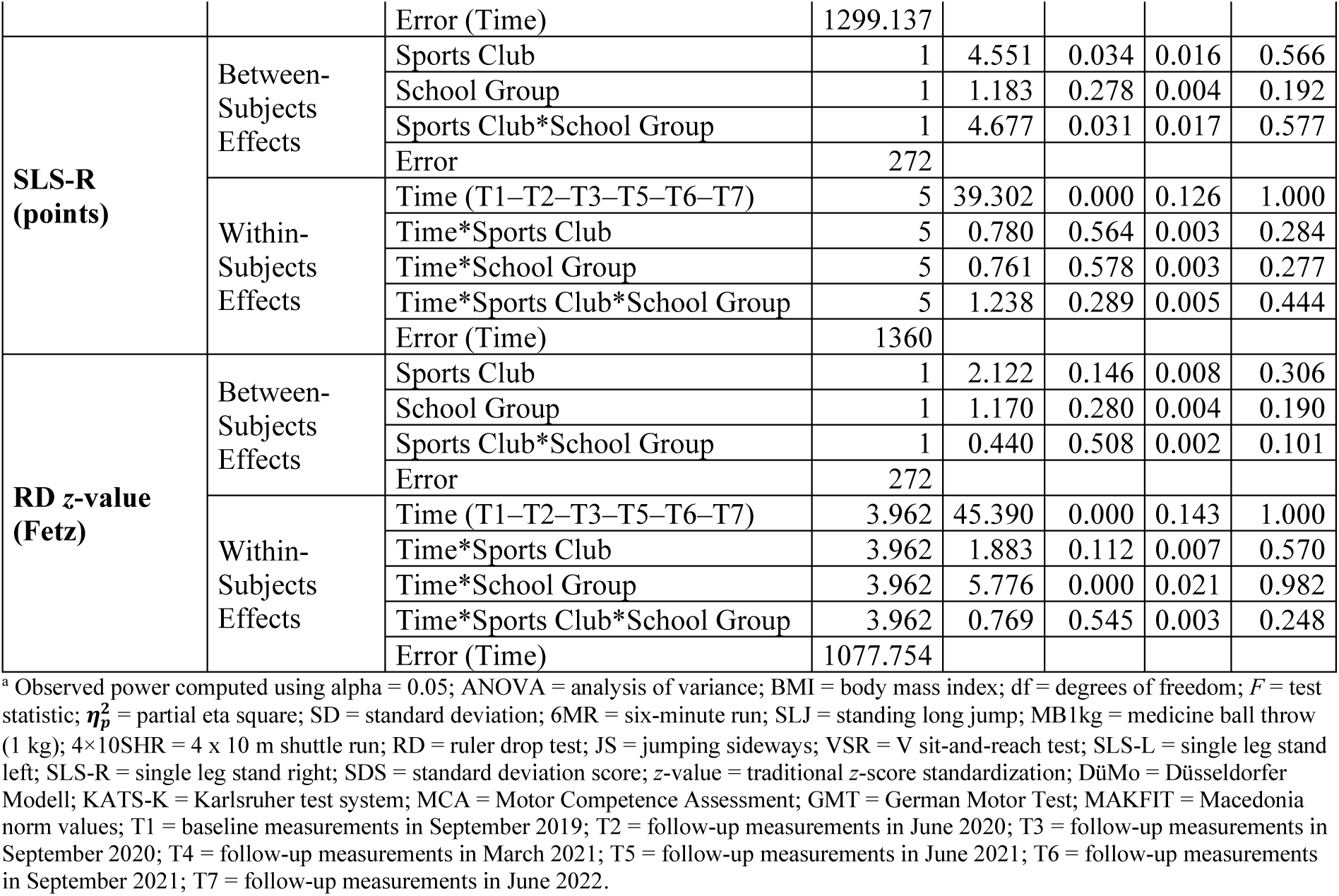
Mixed ANOVAS for the individual fitness tests over all measurement time points.

**Figure R5.**
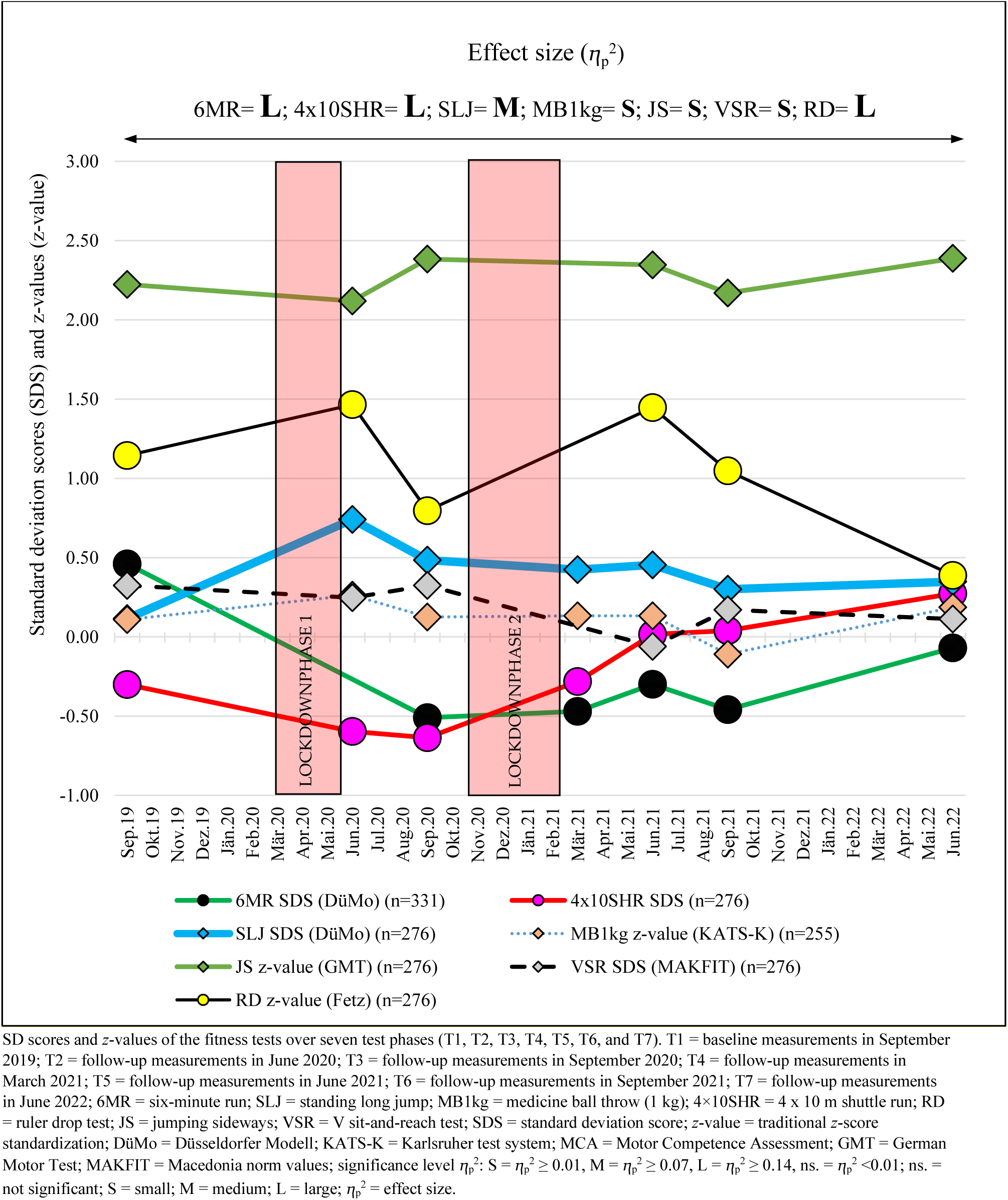
Changes in standard deviation scores (SDS) and *z*-values (*z*-value) of fitness tests 6MR, 4×10SHR, SLJ, MB1kg, JS, VSR, and RD for the main effect of time.

Large effects were observed in cardiorespiratory fitness (6MR), action speed (4×10SHR), and eye–hand reaction speed (RD) (Table R10 and Figure R5).

Medium effects were observed in the standing long jump, the parameter correlated with total body strength, while small effects were found in the remaining fitness tests (MB1kg, VSR, and SJ) (Table R10 and Figure R5).

#### Six-Minute Run (6MR)

Cardiorespiratory fitness (6MR) showed significant changes, with a large effect over the observation period (T1 to T7: main effect of time: ⴄ_p_^2^ = 0.223; *p* < 0.001). An extreme decrease was observed between September 2019 and September 2020 (T1 = 0.46 to T3 = – 0.51 (–0.97 (95% CI, –1.15 to –0.84); *p* < 0.001). Children who reported sports club membership subsequently showed a trend of a slight increase by September 2021 (+0.17 (95% CI, –0.11 to 0.43); *p* > 0.99), but without scientific significance (Tables R10 and R11 and Figures R5 and R6). Children without sports club membership showed a continuous worsening trend between September 2020 and September 2021 (–0.04 (95% CI, –0.27 to 0.20); *p* > 0.99), but without scientific significance (Table R11 and Figure R6). A significant increase in SDS scores was demonstrated between March 2021 and June 2021 (+0.17 (95% CI, 0.04–0.30); *p* = 0.002); this sharp increase was followed by a significant worsening (–0.16 (95% CI, –0.01 to –0.31); *p* = 0.036) between June 2021 and September 2021 (Table R11 and Figure R6).

**Table R11.** Post hoc tests of 6MR for the main effect of time and interactions for time*school group and time*sports club membership based on estimated marginal means.

|  |  | Pairwise Comparisons | Mean Difference (95% CI) | SE | *p*-Value<sub>a</sub> | *p*-lvl |
| --- | --- | --- | --- | --- | --- | --- |
| 6MR SDS (DüMo) | Time | T1 to T3 | 0.990 ( 0.836 to 1.145 ) | 0.052 | <0.001 | *** |
|  |  | T1 to T4 | 0.937 ( 0.780 to 1.094 ) | 0.053 | <0.001 | *** |
|  |  | T1 to T5 | 0.771 ( 0.598 to 0.944 ) | 0.059 | <0.001 | *** |
|  |  | T1 to T6 | 0.929 ( 0.745 to 1.113 ) | 0.062 | <0.001 | *** |
|  |  | T1 to T7 | 0.548 ( 0.372 to 0.723 ) | 0.059 | <0.001 | *** |
|  |  | T3 to T4 | -0.053 ( -0.192 to 0.085 ) | 0.047 | >0.99 |  |
|  |  | T3 to T5 | -0.220 ( -0.364 to -0.075 ) | 0.049 | <0.001 | *** |
|  |  | T3 to T6 | -0.062 ( -0.240 to 0.116 ) | 0.060 | >0.99 |  |
|  |  | T3 to T7 | -0.443 ( -0.610 to -0.276 ) | 0.056 | <0.001 | *** |
|  |  | T4 to T5 | -0.166 ( -0.295 to -0.037 ) | 0.044 | 0.002 | ** |
|  |  | T4 to T6 | -0.009 ( -0.175 to 0.158 ) | 0.056 | >0.99 |  |
|  |  | T4 to T7 | -0.390 ( -0.554 to -0.225 ) | 0.056 | <0.001 | *** |
|  |  | T5 to T6 | 0.158 ( 0.005 to 0.310 ) | 0.052 | 0.036 | * |
|  |  | T5 to T7 | -0.223 ( -0.391 to -0.056 ) | 0.057 | 0.001 | ** |
|  |  | T6 to T7 | -0.381 ( -0.557 to -0.205 ) | 0.060 | <0.001 | *** |
|  | Time*School Group (Intervention Group) | T1 to T3 | 0.930 ( 0.722 to 1.138 ) | 0.070 | <0.001 | *** |
|  |  | T1 to T4 | 0.885 ( 0.674 to 1.097 ) | 0.072 | <0.001 | *** |
|  |  | T1 to T5 | 0.633 ( 0.399 to 0.866 ) | 0.079 | <0.001 | *** |
|  |  | T1 to T6 | 0.835 ( 0.587 to 1.083 ) | 0.084 | <0.001 | *** |
|  |  | T1 to T7 | 0.385 ( 0.149 to 0.622 ) | 0.080 | <0.001 | *** |
|  |  | T3 to T4 | -0.045 ( -0.231 to 0.142 ) | 0.063 | >0.99 |  |
|  |  | T3 to T5 | -0.297 ( -0.491 to -0.103 ) | 0.066 | <0.001 | *** |
|  |  | T3 to T6 | -0.095 ( -0.334 to 0.144 ) | 0.081 | >0.99 |  |
|  |  | T3 to T7 | -0.545 ( -0.770 to -0.320 ) | 0.076 | <0.001 | *** |
|  |  | T4 to T5 | -0.252 ( -0.426 to -0.079 ) | 0.059 | <0.001 | *** |
|  |  | T4 to T6 | -0.050 ( -0.274 to 0.174 ) | 0.076 | >0.99 |  |
|  |  | T4 to T7 | -0.500 ( -0.722 to -0.278 ) | 0.075 | <0.001 | *** |
|  |  | T5 to T6 | 0.202 ( -0.003 to 0.407 ) | 0.069 | 0.057 |  |
|  |  | T5 to T7 | -0.248 ( -0.473 to -0.022 ) | 0.076 | 0.019 | * |
|  |  | T6 to T7 | -0.450 ( -0.687 to -0.213 ) | 0.080 | <0.001 | *** |
|  | Time*School Group (Control Group) | T1 to T3 | 1.051 ( 0.822 to 1.279 ) | 0.077 | <0.001 | *** |
|  |  | T1 to T4 | 0.989 ( 0.757 to 1.221 ) | 0.079 | <0.001 | *** |
|  |  | T1 to T5 | 0.909 ( 0.653 to 1.165 ) | 0.087 | <0.001 | *** |
|  |  | T1 to T6 | 1.022 ( 0.750 to 1.294 ) | 0.092 | <0.001 | *** |
|  |  | T1 to T7 | 0.710 ( 0.450 to 0.970 ) | 0.088 | <0.001 | *** |
|  |  | T3 to T4 | -0.062 ( -0.267 to 0.143 ) | 0.069 | >0.99 |  |
|  |  | T3 to T5 | -0.142 ( -0.355 to 0.071 ) | 0.072 | 0.746 |  |
|  |  | T3 to T6 | -0.029 ( -0.292 to 0.234 ) | 0.089 | >0.99 |  |
|  |  | T3 to T7 | -0.341 ( -0.588 to -0.094 ) | 0.084 | <0.001 | *** |
|  |  | T4 to T5 | -0.080 ( -0.271 to 0.110 ) | 0.064 | >0.99 |  |
|  |  | T4 to T6 | 0.033 ( -0.213 to 0.279 ) | 0.083 | >0.99 |  |
|  |  | T4 to T7 | -0.279 ( -0.523 to -0.036 ) | 0.082 | 0.012 | * |
|  |  | T5 to T6 | 0.113 ( -0.112 to 0.339 ) | 0.076 | >0.99 |  |
|  |  | T5 to T7 | -0.199 ( -0.446 to 0.049 ) | 0.084 | 0.271 |  |
|  |  | T6 to T7 | -0.312 ( -0.573 to -0.052 ) | 0.088 | 0.007 | ** |
|  | Time*Sports Club (Yes) | T1 to T3 | 1.092 ( 0.860 to 1.323 ) | 0.078 | <0.001 | *** |
|  |  | T1 to T4 | 0.972 ( 0.737 to 1.208 ) | 0.080 | <0.001 | *** |
|  |  | T1 to T5 | 0.736 ( 0.476 to 0.997 ) | 0.088 | <0.001 | *** |
|  |  | T1 to T6 | 0.932 ( 0.656 to 1.208 ) | 0.093 | <0.001 | *** |
|  |  | T1 to T7 | 0.565 ( 0.302 to 0.829 ) | 0.089 | <0.001 | *** |
|  |  | T3 to T4 | -0.119 ( -0.327 to 0.089 ) | 0.070 | >0.99 |  |
|  |  | T3 to T5 | -0.355 ( -0.572 to -0.139 ) | 0.073 | <0.001 | *** |
|  |  | T3 to T6 | -0.160 ( -0.427 to 0.107 ) | 0.090 | >0.99 |  |
|  |  | T3 to T7 | -0.526 ( -0.777 to -0.276 ) | 0.085 | <0.001 | *** |
|  |  | T4 to T5 | -0.236 ( -0.430 to -0.043 ) | 0.065 | 0.005 | ** |
|  |  | T4 to T6 | -0.041 ( -0.290 to 0.209 ) | 0.084 | >0.99 |  |
|  |  | T4 to T7 | -0.407 ( -0.654 to -0.160 ) | 0.084 | <0.001 | *** |
|  |  | T5 to T6 | 0.195 ( -0.033 to 0.424 ) | 0.077 | 0.180 |  |
|  |  | T5 to T7 | -0.171 ( -0.422 to 0.080 ) | 0.085 | 0.673 |  |
|  |  | T6 to T7 | -0.367 ( -0.631 to -0.102 ) | 0.089 | 0.001 | ** |
|  | Time*Sports Club (No) | T1 to T3 | 0.889 ( 0.685 to 1.093 ) | 0.069 | <0.001 | *** |
|  |  | T1 to T4 | 0.902 ( 0.694 to 1.109 ) | 0.070 | <0.001 | *** |
|  |  | T1 to T5 | 0.805 ( 0.576 to 1.034 ) | 0.077 | <0.001 | *** |
|  |  | T1 to T6 | 0.925 ( 0.682 to 1.168 ) | 0.082 | <0.001 | *** |
|  |  | T1 to T7 | 0.530 ( 0.298 to 0.762 ) | 0.078 | <0.001 | *** |
|  |  | T3 to T4 | 0.013 ( -0.171 to 0.196 ) | 0.062 | >0.99 |  |
|  |  | T3 to T5 | -0.084 ( -0.275 to 0.107 ) | 0.064 | >0.99 |  |
|  |  | T3 to T6 | 0.036 ( -0.199 to 0.271 ) | 0.079 | >0.99 |  |
|  |  | T3 to T7 | -0.359 ( -0.580 to -0.139 ) | 0.075 | <0.001 | *** |
|  |  | T4 to T5 | -0.097 ( -0.267 to 0.074 ) | 0.058 | >0.99 |  |
|  |  | T4 to T6 | 0.024 ( -0.196 to 0.243 ) | 0.074 | >0.99 |  |
|  |  | T4 to T7 | -0.372 ( -0.590 to -0.154 ) | 0.074 | <0.001 | *** |
|  |  | T5 to T6 | 0.120 ( -0.081 to 0.322 ) | 0.068 | >0.99 |  |
|  |  | T5 to T7 | -0.275 ( -0.497 to -0.054 ) | 0.075 | 0.004 | ** |
|  |  | T6 to T7 | -0.396 ( -0.628 to -0.163 ) | 0.079 | <0.001 | *** |

<sup>a</sup> Adjusted for multiple comparisons using Bonferroni correction. *p*-lvl (*p*-value level) = \* *p* < 0.05, \*\* *p* < 0.01, and \*\*\* *p* < 0.001; CI = confidence interval; 6MR = six-minute run; DüMo = Düsseldorfer Modell; Mean Difference = mean difference based on the estimated marginal means; *p*-lvl = significance level; SE = standard error; T1 = baseline measurements in September 2019; T3 = follow-up measurements in September 2020; T4 = follow-up measurements in March 2021; T5 = follow-up measurements in June 2021; T6 = follow-up measurements in September 2021; T7 = follow-up measurements in June 2022.

**Figure R6.**
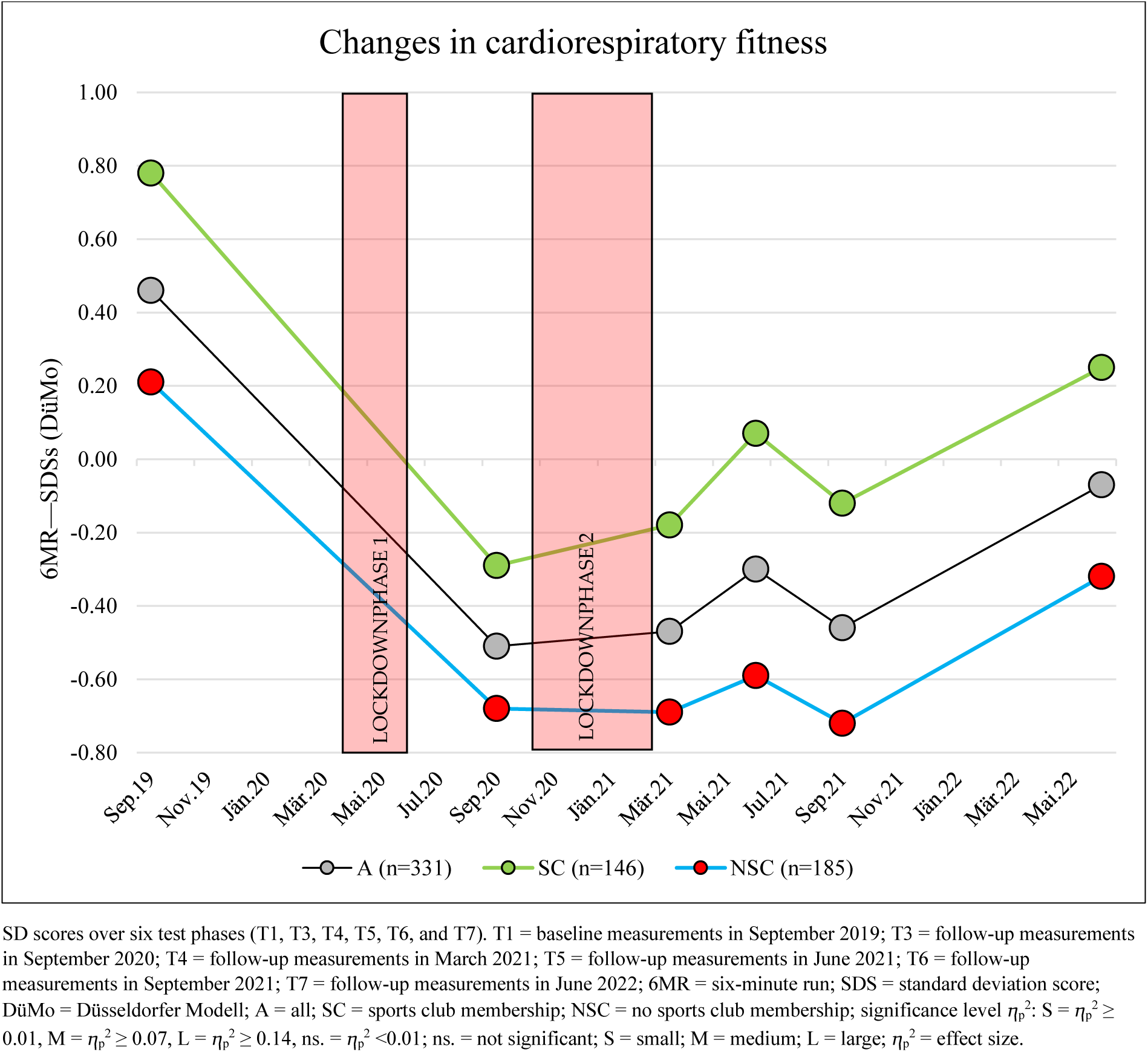
Changes in cardiorespiratory fitness from September 2019 to June 2022.

No interaction effects were found over time (time*sports club: ⴄ_p_^2^ = 0.004; *p* = 0.23) (Table R10 and Figures R5 and R6).

Changes in 6MR in the school groups: (subgroup: intervention and control group SY21–22)

No interaction effects were found over time (time*school group: ⴄ_p_^2^ = 0.007; *p* = 0.044) (Table R10 and Figure R7).

**Figure R7.**
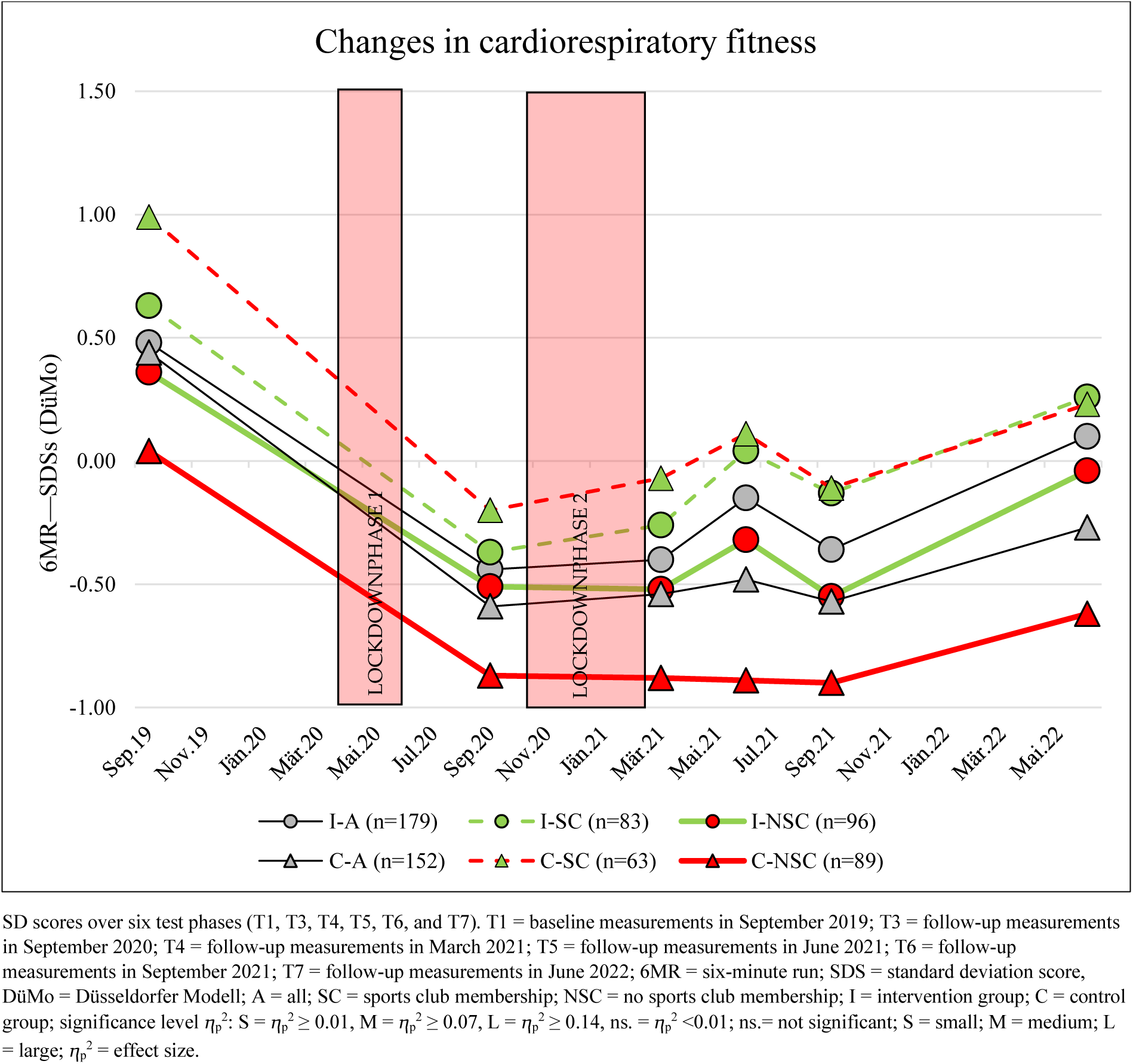
Changes in cardiorespiratory fitness from September 2019 to June 2022 in the subgroups: Intervention and control groups divided by sports club membership.

During the observation period from September 2021 to June 2022, significant improvements were found in the sports intervention and control groups (I-A = +0.46 (95% CI, 0.21–0.69), *p* < 0.001; C-A = +0.30 (95% CI, 0.05–0.57), *p* = 0.007) (Table R11 and Figure R7).

This trend toward improvement was observed in all subgroups (sports club membership vs. no sports club membership) of the intervention and control groups (Table R11 and Figure R7).

#### Action Speed (4×10SHR)

Action speed (4×10SHR) showed significant changes with a large effect over the observation period (T1 to T7: main effect of time: ⴄ_p_^2^ = 0.267; *p* < 0.001) (Table R10 and Figure R8).

**Figure R8.**
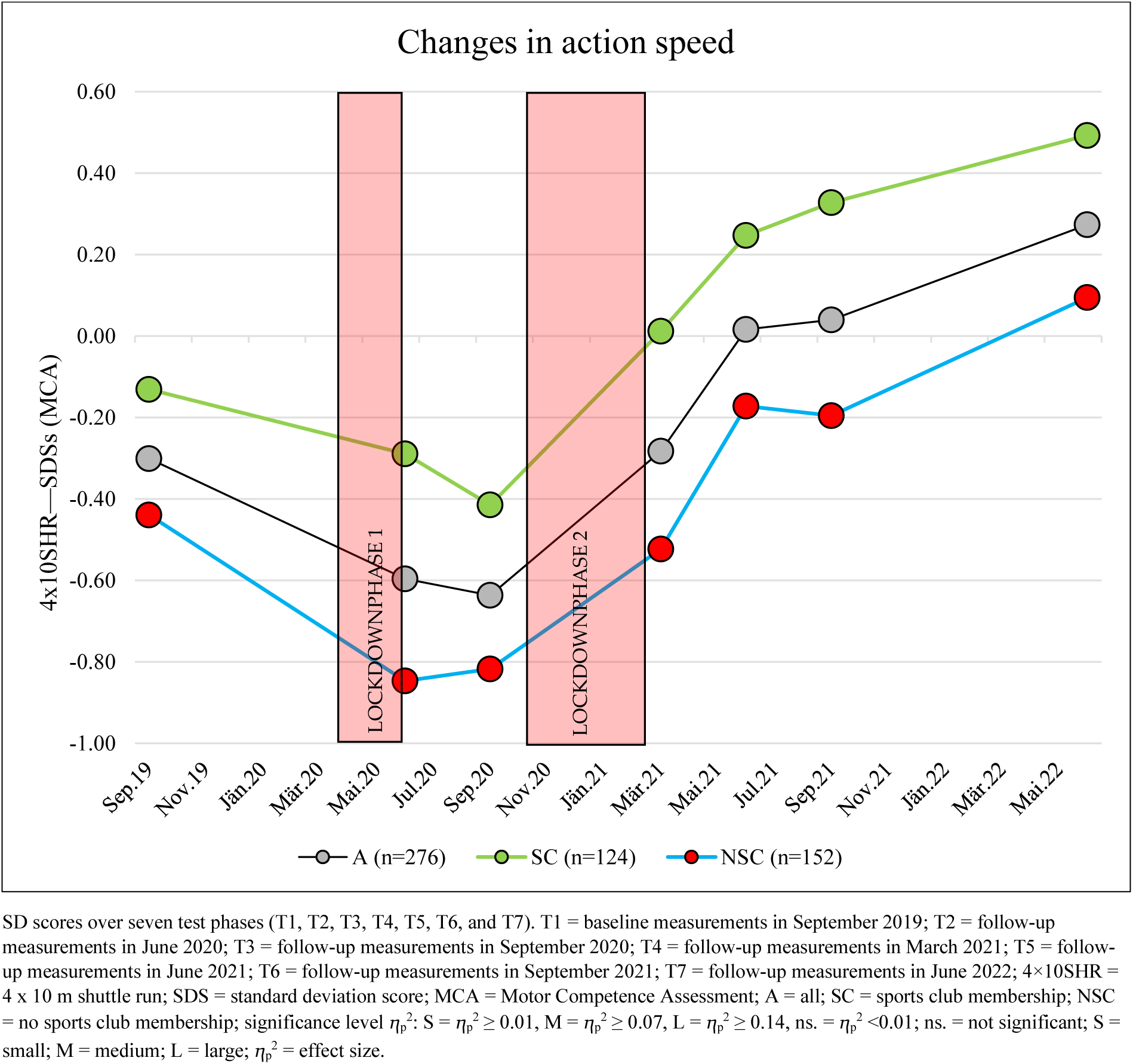
Changes in action speed from September 2019 to June 2022.

A dramatic decline was observed between September 2019 and September 2020 (–0.34 (95% CI, –0.49 to –0.17); p < 0.001), followed by a strong improvement in action speed by June 2021 (+0.66 (95% CI, 0.49–0.82); *p* < 0.001). Between June 2021 and September 2021, there was a slight improvement without scientific significance (+0.02 (95% CI, –0.08 to 0.15); *p* > 0.99). Finally, between September 2021 and June 2022, a strong improvement was demonstrated (+0.23 (95% CI, 0.11–0.35); *p* < 0.001) (Table R12 and Figure R8).

**Table R12.** Post hoc tests of 4×10SHR for the main effect of time and interactions for time*school group and time*sports club membership based on estimated marginal means.

|  |  | Pairwise Comparisons | Mean Difference (95% CI) | SE | *p*-Value<sub>a</sub> | *p*-lvl |
| --- | --- | --- | --- | --- | --- | --- |
| 4×10SHR SDS (MCA) | Time | T1 to T2 | 0.270 ( 0.100 to 0.441 ) | 0.056 | <0.001 | *** |
|  |  | T1 to T3 | 0.328 ( 0.165 to 0.490 ) | 0.053 | <0.001 | *** |
|  |  | T1 to T4 | -0.035 ( -0.201 to 0.131 ) | 0.054 | >0.99 |  |
|  |  | T1 to T5 | -0.328 ( -0.488 to -0.168 ) | 0.052 | <0.001 | *** |
|  |  | T1 to T6 | -0.358 ( -0.508 to -0.208 ) | 0.049 | <0.001 | *** |
|  |  | T1 to T7 | -0.588 ( -0.747 to -0.429 ) | 0.052 | <0.001 | *** |
|  |  | T2 to T3 | 0.057 ( -0.093 to 0.208 ) | 0.049 | >0.99 |  |
|  |  | T2 to T4 | -0.305 ( -0.451 to -0.159 ) | 0.048 | <0.001 | *** |
|  |  | T2 to T5 | -0.598 ( -0.748 to -0.447 ) | 0.049 | <0.001 | *** |
|  |  | T2 to T6 | -0.628 ( -0.775 to -0.481 ) | 0.048 | <0.001 | *** |
|  |  | T2 to T7 | -0.859 ( -1.004 to -0.713 ) | 0.047 | <0.001 | *** |
|  |  | T3 to T4 | -0.362 ( -0.503 to -0.222 ) | 0.046 | <0.001 | *** |
|  |  | T3 to T5 | -0.655 ( -0.818 to -0.493 ) | 0.053 | <0.001 | *** |
|  |  | T3 to T6 | -0.686 ( -0.843 to -0.529 ) | 0.051 | <0.001 | *** |
|  |  | T3 to T7 | -0.916 ( -1.070 to -0.762 ) | 0.050 | <0.001 | *** |
|  |  | T4 to T5 | -0.293 ( -0.436 to -0.150 ) | 0.047 | <0.001 | *** |
|  |  | T4 to T6 | -0.323 ( -0.458 to -0.189 ) | 0.044 | <0.001 | *** |
|  |  | T4 to T7 | -0.554 ( -0.683 to -0.424 ) | 0.042 | <0.001 | *** |
|  |  | T5 to T6 | -0.031 ( -0.145 to 0.084 ) | 0.037 | >0.99 |  |
|  |  | T5 to T7 | -0.261 ( -0.383 to -0.139 ) | 0.040 | <0.001 | *** |
|  |  | T6 to T7 | -0.230 ( -0.354 to -0.106 ) | 0.040 | <0.001 | *** |
|  | Time*School Group (Intervention Group) | T1 to T2 | 0.399 ( 0.162 to 0.637 ) | 0.077 | <0.001 | *** |
|  |  | T1 to T3 | 0.377 ( 0.150 to 0.603 ) | 0.074 | <0.001 | *** |
|  |  | T1 to T4 | -0.089 ( -0.321 to 0.143 ) | 0.076 | >0.99 |  |
|  |  | T1 to T5 | -0.230 ( -0.453 to -0.007 ) | 0.073 | 0.037 | * |
|  |  | T1 to T6 | -0.282 ( -0.491 to -0.073 ) | 0.068 | 0.001 | ** |
|  |  | T1 to T7 | -0.523 ( -0.745 to -0.301 ) | 0.072 | <0.001 | *** |
|  |  | T2 to T3 | -0.022 ( -0.232 to 0.187 ) | 0.068 | >0.99 |  |
|  |  | T2 to T4 | -0.488 ( -0.692 to -0.284 ) | 0.066 | <0.001 | *** |
|  |  | T2 to T5 | -0.629 ( -0.839 to -0.419 ) | 0.068 | <0.001 | *** |
|  |  | T2 to T6 | -0.681 ( -0.886 to -0.476 ) | 0.067 | <0.001 | *** |
|  |  | T2 to T7 | -0.922 ( -1.125 to -0.719 ) | 0.066 | <0.001 | *** |
|  |  | T3 to T4 | -0.466 ( -0.662 to -0.270 ) | 0.064 | <0.001 | *** |
|  |  | T3 to T5 | -0.607 ( -0.833 to -0.380 ) | 0.074 | <0.001 | *** |
|  |  | T3 to T6 | -0.659 ( -0.878 to -0.439 ) | 0.071 | <0.001 | *** |
|  |  | T3 to T7 | -0.899 ( -1.114 to -0.685 ) | 0.070 | <0.001 | *** |
|  |  | T4 to T5 | -0.141 ( -0.340 to 0.058 ) | 0.065 | 0.652 |  |
|  |  | T4 to T6 | -0.193 ( -0.380 to -0.005 ) | 0.061 | 0.038 | * |
|  |  | T4 to T7 | -0.434 ( -0.614 to -0.253 ) | 0.059 | <0.001 | *** |
|  |  | T5 to T6 | -0.052 ( -0.212 to 0.108 ) | 0.052 | >0.99 |  |
|  |  | T5 to T7 | -0.293 ( -0.463 to -0.123 ) | 0.055 | <0.001 | *** |
|  |  | T6 to T7 | -0.241 ( -0.414 to -0.068 ) | 0.056 | <0.001 | *** |
|  | Time*School Group (Control Group) | T1 to T2 | 0.141 ( -0.103 to 0.385 ) | 0.080 | >0.99 |  |
|  |  | T1 to T3 | 0.279 ( 0.046 to 0.511 ) | 0.076 | 0.006 | ** |
|  |  | T1 to T4 | 0.020 ( -0.218 to 0.258 ) | 0.078 | >0.99 |  |
|  |  | T1 to T5 | -0.425 ( -0.655 to -0.196 ) | 0.075 | <0.001 | *** |
|  |  | T1 to T6 | -0.434 ( -0.649 to -0.219 ) | 0.070 | <0.001 | *** |
|  |  | T1 to T7 | -0.654 ( -0.882 to -0.426 ) | 0.074 | <0.001 | *** |
|  |  | T2 to T3 | 0.137 ( -0.078 to 0.352 ) | 0.070 | >0.99 |  |
|  |  | T2 to T4 | -0.121 ( -0.331 to 0.088 ) | 0.068 | >0.99 |  |
|  |  | T2 to T5 | -0.566 ( -0.782 to -0.351 ) | 0.070 | <0.001 | *** |
|  |  | T2 to T6 | -0.576 ( -0.786 to -0.365 ) | 0.069 | <0.001 | *** |
|  |  | T2 to T7 | -0.795 ( -1.004 to -0.587 ) | 0.068 | <0.001 | *** |
|  |  | T3 to T4 | -0.259 ( -0.461 to -0.057 ) | 0.066 | 0.002 | ** |
|  |  | T3 to T5 | -0.704 ( -0.937 to -0.471 ) | 0.076 | <0.001 | *** |
|  |  | T3 to T6 | -0.713 ( -0.938 to -0.488 ) | 0.073 | <0.001 | *** |
|  |  | T3 to T7 | -0.933 ( -1.153 to -0.712 ) | 0.072 | <0.001 | *** |
|  |  | T4 to T5 | -0.445 ( -0.650 to -0.240 ) | 0.067 | <0.001 | *** |
|  |  | T4 to T6 | -0.454 ( -0.647 to -0.261 ) | 0.063 | <0.001 | *** |
|  |  | T4 to T7 | -0.674 ( -0.859 to -0.488 ) | 0.060 | <0.001 | *** |
|  |  | T5 to T6 | -0.009 ( -0.173 to 0.155 ) | 0.053 | >0.99 |  |
|  |  | T5 to T7 | -0.229 ( -0.403 to -0.054 ) | 0.057 | 0.002 | ** |
|  |  | T6 to T7 | -0.220 ( -0.397 to -0.042 ) | 0.058 | 0.004 | ** |
|  | Time*Sports Club<br>(Yes) | T1 to T2 | 0.135 ( -0.118 to 0.388 ) | 0.083 | >0.99 |  |
|  |  | T1 to T3 | 0.277 ( 0.036 to 0.519 ) | 0.079 | 0.011 | * |
|  |  | T1 to T4 | -0.146 ( -0.393 to 0.100 ) | 0.080 | >0.99 |  |
|  |  | T1 to T5 | -0.389 ( -0.627 to -0.152 ) | 0.078 | <0.001 | *** |
|  |  | T1 to T6 | -0.471 ( -0.694 to -0.249 ) | 0.073 | <0.001 | *** |
|  |  | T1 to T7 | -0.640 ( -0.876 to -0.403 ) | 0.077 | <0.001 | *** |
|  |  | T2 to T3 | 0.142 ( -0.081 to 0.365 ) | 0.073 | >0.99 |  |
|  |  | T2 to T4 | -0.281 ( -0.498 to -0.064 ) | 0.071 | 0.002 | ** |
|  |  | T2 to T5 | -0.524 ( -0.748 to -0.301 ) | 0.073 | <0.001 | *** |
|  |  | T2 to T6 | -0.606 ( -0.825 to -0.388 ) | 0.071 | <0.001 | *** |
|  |  | T2 to T7 | -0.775 ( -0.991 to -0.558 ) | 0.070 | <0.001 | *** |
|  |  | T3 to T4 | -0.423 ( -0.633 to -0.214 ) | 0.068 | <0.001 | *** |
|  |  | T3 to T5 | -0.667 ( -0.908 to -0.425 ) | 0.079 | <0.001 | *** |
|  |  | T3 to T6 | -0.749 ( -0.982 to -0.515 ) | 0.076 | <0.001 | *** |
|  |  | T3 to T7 | -0.917 ( -1.145 to -0.688 ) | 0.074 | <0.001 | *** |
|  |  | T4 to T5 | -0.243 ( -0.455 to -0.031 ) | 0.069 | 0.011 | * |
|  |  | T4 to T6 | -0.325 ( -0.525 to -0.125 ) | 0.065 | <0.001 | *** |
|  |  | T4 to T7 | -0.493 ( -0.686 to -0.301 ) | 0.063 | <0.001 | *** |
|  |  | T5 to T6 | -0.082 ( -0.252 to 0.088 ) | 0.055 | >0.99 |  |
|  |  | T5 to T7 | -0.250 ( -0.431 to -0.069 ) | 0.059 | <0.001 | *** |
|  |  | T6 to T7 | -0.168 ( -0.352 to 0.016 ) | 0.060 | 0.114 |  |
|  | Time*Sports Club<br>(No) | T1 to T2 | 0.406 ( 0.178 to 0.633 ) | 0.074 | <0.001 | *** |
|  |  | T1 to T3 | 0.378 ( 0.161 to 0.596 ) | 0.071 | <0.001 | *** |
|  |  | T1 to T4 | 0.077 ( -0.145 to 0.299 ) | 0.072 | >0.99 |  |
|  |  | T1 to T5 | -0.266 ( -0.480 to -0.051 ) | 0.070 | 0.004 | ** |
|  |  | T1 to T6 | -0.245 ( -0.446 to -0.044 ) | 0.065 | 0.005 | ** |
|  |  | T1 to T7 | -0.537 ( -0.750 to -0.324 ) | 0.069 | <0.001 | *** |
|  |  | T2 to T3 | -0.027 ( -0.228 to 0.174 ) | 0.065 | >0.99 |  |
|  |  | T2 to T4 | -0.329 ( -0.524 to -0.133 ) | 0.064 | <0.001 | *** |
|  |  | T2 to T5 | -0.671 ( -0.872 to -0.470 ) | 0.066 | <0.001 | *** |
|  |  | T2 to T6 | -0.650 ( -0.847 to -0.454 ) | 0.064 | <0.001 | *** |
|  |  | T2 to T7 | -0.943 ( -1.137 to -0.748 ) | 0.063 | <0.001 | *** |
|  |  | T3 to T4 | -0.301 ( -0.490 to -0.113 ) | 0.061 | <0.001 | *** |
|  |  | T3 to T5 | -0.644 ( -0.861 to -0.426 ) | 0.071 | <0.001 | *** |
|  |  | T3 to T6 | -0.623 ( -0.833 to -0.413 ) | 0.069 | <0.001 | *** |
|  |  | T3 to T7 | -0.915 ( -1.121 to -0.709 ) | 0.067 | <0.001 | *** |
|  |  | T4 to T5 | -0.343 ( -0.534 to -0.151 ) | 0.062 | <0.001 | *** |
|  |  | T4 to T6 | -0.322 ( -0.502 to -0.142 ) | 0.059 | <0.001 | *** |
|  |  | T4 to T7 | -0.614 ( -0.787 to -0.441 ) | 0.056 | <0.001 | *** |
|  |  | T5 to T6 | 0.021 ( -0.132 to 0.174 ) | 0.050 | >0.99 |  |
|  |  | T5 to T7 | -0.271 ( -0.434 to -0.108 ) | 0.053 | <0.001 | *** |
|  |  | T6 to T7 | -0.292 ( -0.458 to -0.126 ) | 0.054 | <0.001 | *** |

<sup>a</sup> Adjusted for multiple comparisons using Bonferroni correction. $p$ -lvl ( $p$ -value level) = \* $p < 0.05$ , \*\* $p < 0.01$ , and \*\*\* $p < 0.001$ ; CI = confidence interval; 4x10SHR = 4 x 10 m shuttle run; MCA = Motor Competence Assessment; Mean Difference = mean difference based on the estimated marginal means; $p$ -lvl = significance level; SE = standard error; T1 = baseline measurements in September 2019; T2 = follow-up measurements in June 2020; T3 = follow-up measurements in September 2020; T4 = follow-up measurements in March 2021; T5 = follow-up measurements in June 2021; T6 = follow-up measurements in September 2021; T7 = follow-up measurements in June 2022.

No interaction effects were found over time (time*sports club: ⴄ_p_^2^ = 0.007; *p* = 0.08) (Table R10 and Figure R8).

Changes in 4x10SHR in the school groups: (subgroup: intervention and control group SY21– 22)

Small interaction effects were found over time (time*school group: ⴄ_p_^2^ = 0.012, *p* = 0.005; time*sports club*school group: ⴄ_p_^2^ = 0.011, *p* = 0.010), which were mainly related to the time before the sports intervention (September 2019 to September 2021). Significant changes were found during the observation period from September 2021 to June 2022 in the intervention and control groups (I-A = +0.24 (95% CI, 0.07–0.41), *p* < 0.001; C-A = +0.22 (95% CI, 0.04–0.40), *p* < 0.001). This trend of improvement was observed in all subgroups of the intervention and control groups (Tables R10 and R12 and Figure R9).

**Figure R9.**
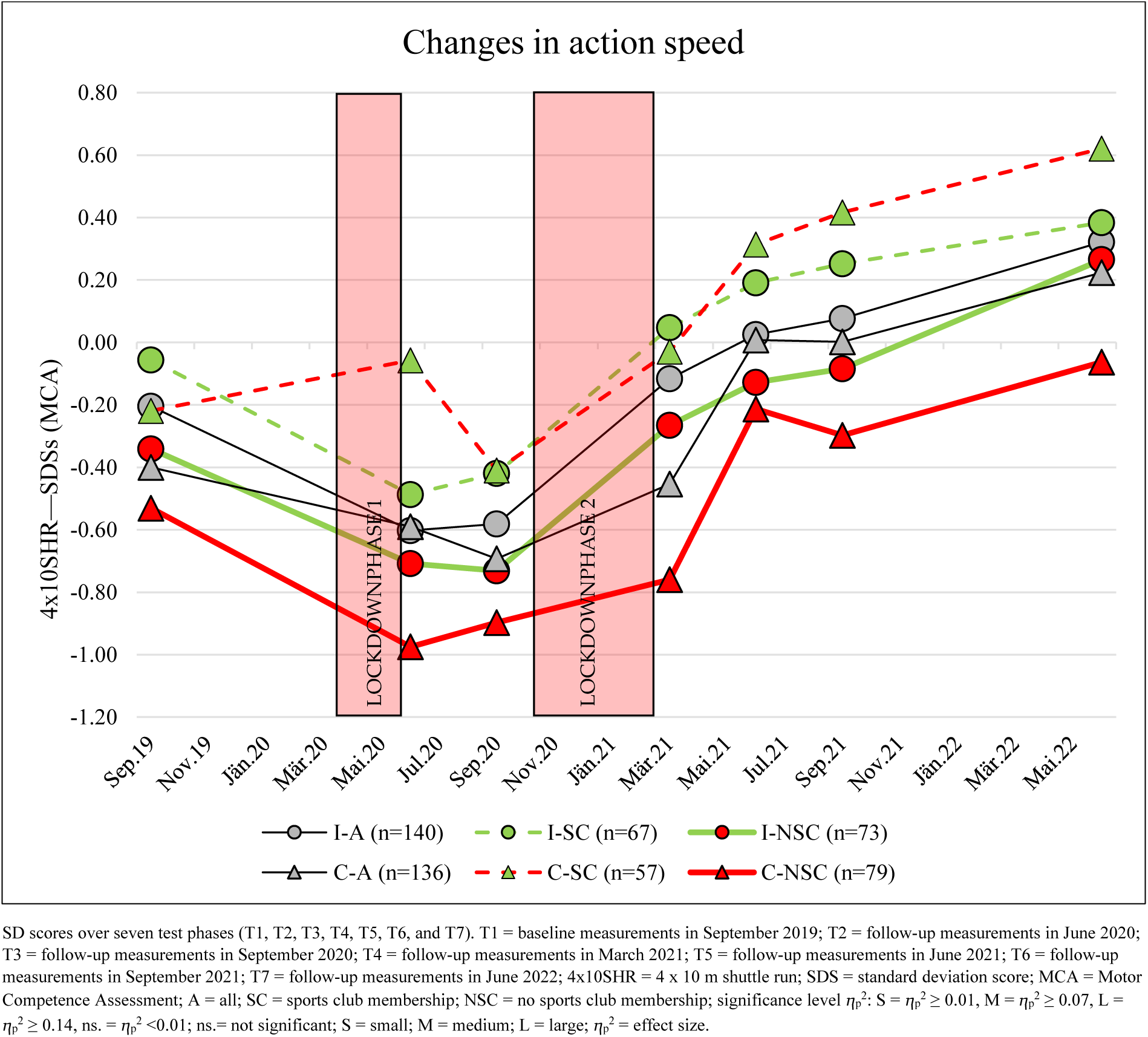
Changes in action speed from September 2019 to June 2022 in the subgroups: Intervention and control groups divided by sports club membership.

#### Standing Long Jump (SLJ)

The standing long jump (SLJ), the leg strength parameter, showed significant changes, with a medium effect over the observation period (T1 to T7: main effect of time: ⴄ_p_^2^ = 0.111; *p* < 0.001) (Table R10 and Figure R10).

**Figure R10.**
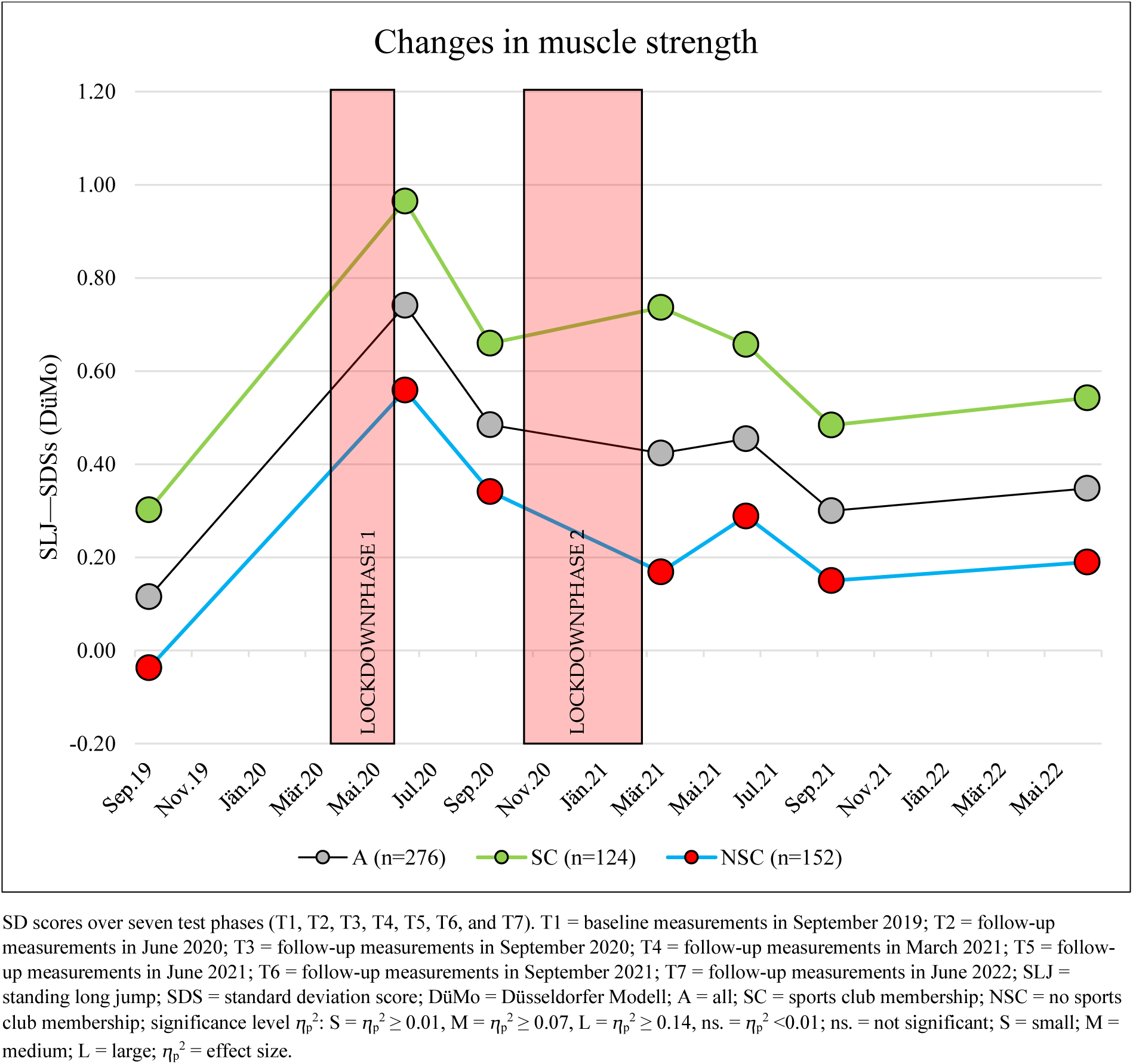
Changes in muscle strength from September 2019 to June 2022.

A strong improvement in SDS was observed between September 2019 and June 2020 (+0.63 (95% CI, 0.47–0.78); *p* < 0.001). Between June 2020 and September 2021, the SDS showed a decline of –0.44 (95% CI, –0.30 to –0.58); *p* < 0.001) (Table R13 and Figure R10). During the observation period from September 2021 to June 2022, standing long jump performance stabilized and showed a slight trend of improvement, which, however, did not represent scientific significance (+0.05 (95% CI, –0.05 to 0.15); *p* > 0.99) (Table R13 and Figure R10).

**Table R13.** Post hoc tests of SLJ for the main effect of time and interactions for time*school group and time*sports club membership based on estimated marginal means.

|  |  | Pairwise Comparisons | Mean Difference (95% CI) | SE | *p*-Value<sub>a</sub> | *p*-lvl |
| --- | --- | --- | --- | --- | --- | --- |
| SLJ SDS (DüMo) | Time | T1 to T2 | −0.624 ( −0.779 to −0.470 ) | 0.050 | <0.001 | *** |
|  |  | T1 to T3 | −0.366 ( −0.533 to −0.199 ) | 0.055 | <0.001 | *** |
|  |  | T1 to T4 | −0.316 ( −0.474 to −0.159 ) | 0.051 | <0.001 | *** |
|  |  | T1 to T5 | −0.337 ( −0.505 to −0.169 ) | 0.055 | <0.001 | *** |
|  |  | T1 to T6 | −0.183 ( −0.344 to −0.023 ) | 0.052 | 0.011 | * |
|  |  | T1 to T7 | −0.232 ( −0.392 to −0.073 ) | 0.052 | <0.001 | *** |
|  |  | T2 to T3 | 0.258 ( 0.111 to 0.405 ) | 0.048 | <0.001 | *** |
|  |  | T2 to T4 | 0.308 ( 0.167 to 0.448 ) | 0.046 | <0.001 | *** |
|  |  | T2 to T5 | 0.287 ( 0.142 to 0.432 ) | 0.047 | <0.001 | *** |
|  |  | T2 to T6 | 0.441 ( 0.299 to 0.583 ) | 0.046 | <0.001 | *** |
|  |  | T2 to T7 | 0.392 ( 0.244 to 0.539 ) | 0.048 | <0.001 | *** |
|  |  | T3 to T4 | 0.050 ( −0.098 to 0.197 ) | 0.048 | >0.99 |  |
|  |  | T3 to T5 | 0.029 ( −0.124 to 0.182 ) | 0.050 | >0.99 |  |
|  |  | T3 to T6 | 0.183 ( 0.043 to 0.322 ) | 0.045 | 0.002 | ** |
|  |  | T3 to T7 | 0.134 ( −0.017 to 0.284 ) | 0.049 | 0.147 |  |
|  |  | T4 to T5 | −0.021 ( −0.133 to 0.092 ) | 0.037 | >0.99 |  |
|  |  | T4 to T6 | 0.133 ( 0.015 to 0.251 ) | 0.039 | 0.014 | * |
|  |  | T4 to T7 | 0.084 ( −0.034 to 0.202 ) | 0.039 | 0.631 |  |
|  |  | T5 to T6 | 0.154 ( 0.048 to 0.259 ) | 0.034 | <0.001 | *** |
|  |  | T5 to T7 | 0.105 ( −0.014 to 0.223 ) | 0.039 | 0.153 |  |
|  |  | T6 to T7 | −0.049 ( −0.149 to 0.051 ) | 0.033 | >0.99 |  |
|  | Time*School Group (Intervention Group) | T1 to T2 | −0.639 ( −0.855 to −0.424 ) | 0.070 | <0.001 | *** |
|  |  | T1 to T3 | −0.287 ( −0.521 to −0.054 ) | 0.076 | 0.004 | ** |
|  |  | T1 to T4 | −0.317 ( −0.537 to −0.098 ) | 0.071 | <0.001 | *** |
|  |  | T1 to T5 | −0.321 ( −0.555 to −0.087 ) | 0.076 | 0.001 | ** |
|  |  | T1 to T6 | −0.109 ( −0.333 to 0.114 ) | 0.073 | >0.99 |  |
|  |  | T1 to T7 | −0.195 ( −0.417 to 0.028 ) | 0.072 | 0.163 |  |
|  |  | T2 to T3 | 0.352 ( 0.147 to 0.557 ) | 0.067 | <0.001 | *** |
|  |  | T2 to T4 | 0.322 ( 0.126 to 0.518 ) | 0.064 | <0.001 | *** |
|  |  | T2 to T5 | 0.318 ( 0.116 to 0.521 ) | 0.066 | <0.001 | *** |
|  |  | T2 to T6 | 0.530 ( 0.332 to 0.728 ) | 0.065 | <0.001 | *** |
|  |  | T2 to T7 | 0.445 ( 0.239 to 0.651 ) | 0.067 | <0.001 | *** |
|  |  | T3 to T4 | −0.030 ( −0.236 to 0.176 ) | 0.067 | >0.99 |  |
|  |  | T3 to T5 | −0.034 ( −0.248 to 0.180 ) | 0.070 | >0.99 |  |
|  |  | T3 to T6 | 0.178 ( −0.016 to 0.372 ) | 0.063 | 0.111 |  |
|  |  | T3 to T7 | 0.093 ( −0.117 to 0.303 ) | 0.069 | >0.99 |  |
|  |  | T4 to T5 | −0.004 ( −0.161 to 0.154 ) | 0.051 | >0.99 |  |
|  |  | T4 to T6 | 0.208 ( 0.043 to 0.373 ) | 0.054 | 0.003 | ** |
|  |  | T4 to T7 | 0.123 ( −0.042 to 0.288 ) | 0.054 | 0.481 |  |
|  |  | T5 to T6 | 0.212 ( 0.065 to 0.359 ) | 0.048 | <0.001 | *** |
|  |  | T5 to T7 | 0.127 ( −0.039 to 0.292 ) | 0.054 | 0.414 |  |
|  |  | T6 to T7 | −0.085 ( −0.225 to 0.055 ) | 0.046 | >0.99 |  |
|  | Time*School Group (Control Group) | T1 to T2 | −0.609 ( −0.830 to −0.387 ) | 0.072 | <0.001 | *** |
|  |  | T1 to T3 | −0.445 ( −0.684 to −0.205 ) | 0.078 | <0.001 | *** |
|  |  | T1 to T4 | −0.315 ( −0.541 to −0.090 ) | 0.073 | <0.001 | *** |
|  |  | T1 to T5 | −0.353 ( −0.594 to −0.112 ) | 0.078 | <0.001 | *** |
|  |  | T1 to T6 | −0.257 ( −0.487 to −0.027 ) | 0.075 | 0.015 | * |
|  |  | T1 to T7 | −0.270 ( −0.499 to −0.042 ) | 0.074 | 0.007 | ** |
|  |  | T2 to T3 | 0.164 ( −0.047 to 0.375 ) | 0.069 | 0.368 |  |
|  |  | T2 to T4 | 0.293 ( 0.092 to 0.495 ) | 0.066 | <0.001 | *** |
|  |  | T2 to T5 | 0.256 ( 0.048 to 0.464 ) | 0.068 | 0.004 | ** |
|  |  | T2 to T6 | 0.351 ( 0.148 to 0.555 ) | 0.066 | <0.001 | *** |
|  |  | T2 to T7 | 0.338 ( 0.127 to 0.550 ) | 0.069 | <0.001 | *** |
|  |  | T3 to T4 | 0.129 ( −0.082 to 0.341 ) | 0.069 | >0.99 |  |
|  |  | T3 to T5 | 0.092 ( −0.128 to 0.311 ) | 0.072 | >0.99 |  |
|  |  | T3 to T6 | 0.187 ( −0.012 to 0.387 ) | 0.065 | 0.091 |  |
|  |  | T3 to T7 | 0.174 ( -0.042 to 0.390 ) | 0.070 | 0.294 |  |
|  |  | T4 to T5 | -0.038 ( -0.199 to 0.124 ) | 0.053 | >0.99 |  |
|  |  | T4 to T6 | 0.058 ( -0.112 to 0.228 ) | 0.055 | >0.99 |  |
|  |  | T4 to T7 | 0.045 ( -0.124 to 0.214 ) | 0.055 | >0.99 |  |
|  |  | T5 to T6 | 0.096 ( -0.055 to 0.246 ) | 0.049 | >0.99 |  |
|  |  | T5 to T7 | 0.083 ( -0.087 to 0.253 ) | 0.055 | >0.99 |  |
|  |  | T6 to T7 | -0.013 ( -0.156 to 0.130 ) | 0.047 | >0.99 |  |
|  | Time*Sports Club<br>(Yes) | T1 to T2 | -0.655 ( -0.884 to -0.425 ) | 0.075 | <0.001 | *** |
|  |  | T1 to T3 | -0.359 ( -0.608 to -0.111 ) | 0.081 | <0.001 | *** |
|  |  | T1 to T4 | -0.429 ( -0.663 to -0.196 ) | 0.076 | <0.001 | *** |
|  |  | T1 to T5 | -0.351 ( -0.601 to -0.102 ) | 0.081 | <0.001 | *** |
|  |  | T1 to T6 | -0.184 ( -0.423 to 0.054 ) | 0.078 | 0.390 |  |
|  |  | T1 to T7 | -0.241 ( -0.477 to -0.004 ) | 0.077 | 0.043 | * |
|  |  | T2 to T3 | 0.296 ( 0.077 to 0.514 ) | 0.071 | <0.001 | *** |
|  |  | T2 to T4 | 0.225 ( 0.016 to 0.435 ) | 0.068 | 0.022 | * |
|  |  | T2 to T5 | 0.303 ( 0.088 to 0.519 ) | 0.070 | <0.001 | *** |
|  |  | T2 to T6 | 0.471 ( 0.260 to 0.681 ) | 0.069 | <0.001 | *** |
|  |  | T2 to T7 | 0.414 ( 0.195 to 0.634 ) | 0.072 | <0.001 | *** |
|  |  | T3 to T4 | -0.070 ( -0.290 to 0.149 ) | 0.072 | >0.99 |  |
|  |  | T3 to T5 | 0.008 ( -0.220 to 0.236 ) | 0.074 | >0.99 |  |
|  |  | T3 to T6 | 0.175 ( -0.032 to 0.382 ) | 0.067 | 0.210 |  |
|  |  | T3 to T7 | 0.118 ( -0.105 to 0.342 ) | 0.073 | >0.99 |  |
|  |  | T4 to T5 | 0.078 ( -0.090 to 0.245 ) | 0.055 | >0.99 |  |
|  |  | T4 to T6 | 0.245 ( 0.069 to 0.421 ) | 0.057 | <0.001 | *** |
|  |  | T4 to T7 | 0.189 ( 0.013 to 0.364 ) | 0.057 | 0.023 | * |
|  |  | T5 to T6 | 0.167 ( 0.011 to 0.324 ) | 0.051 | 0.025 | * |
|  |  | T5 to T7 | 0.111 ( -0.066 to 0.287 ) | 0.058 | >0.99 |  |
|  |  | T6 to T7 | -0.056 ( -0.205 to 0.092 ) | 0.048 | >0.99 |  |
|  | Time*Sports Club<br>(No) | T1 to T2 | -0.593 ( -0.800 to -0.387 ) | 0.067 | <0.001 | *** |
|  |  | T1 to T3 | -0.373 ( -0.597 to -0.149 ) | 0.073 | <0.001 | *** |
|  |  | T1 to T4 | -0.204 ( -0.414 to 0.007 ) | 0.069 | 0.069 |  |
|  |  | T1 to T5 | -0.323 ( -0.547 to -0.098 ) | 0.073 | <0.001 | *** |
|  |  | T1 to T6 | -0.183 ( -0.398 to 0.032 ) | 0.070 | 0.202 |  |
|  |  | T1 to T7 | -0.224 ( -0.438 to -0.011 ) | 0.070 | 0.030 | * |
|  |  | T2 to T3 | 0.220 ( 0.024 to 0.417 ) | 0.064 | 0.014 | * |
|  |  | T2 to T4 | 0.390 ( 0.201 to 0.578 ) | 0.061 | <0.001 | *** |
|  |  | T2 to T5 | 0.271 ( 0.076 to 0.465 ) | 0.063 | <0.001 | *** |
|  |  | T2 to T6 | 0.411 ( 0.221 to 0.601 ) | 0.062 | <0.001 | *** |
|  |  | T2 to T7 | 0.369 ( 0.171 to 0.567 ) | 0.064 | <0.001 | *** |
|  |  | T3 to T4 | 0.169 ( -0.028 to 0.367 ) | 0.064 | 0.191 |  |
|  |  | T3 to T5 | 0.050 ( -0.155 to 0.256 ) | 0.067 | >0.99 |  |
|  |  | T3 to T6 | 0.190 ( 0.004 to 0.377 ) | 0.061 | 0.040 | * |
|  |  | T3 to T7 | 0.149 ( -0.053 to 0.350 ) | 0.066 | 0.517 |  |
|  |  | T4 to T5 | -0.119 ( -0.270 to 0.032 ) | 0.049 | 0.339 |  |
|  |  | T4 to T6 | 0.021 ( -0.138 to 0.179 ) | 0.052 | >0.99 |  |
|  |  | T4 to T7 | -0.021 ( -0.179 to 0.138 ) | 0.052 | >0.99 |  |
|  |  | T5 to T6 | 0.140 ( -0.001 to 0.281 ) | 0.046 | 0.053 |  |
|  |  | T5 to T7 | 0.098 ( -0.060 to 0.257 ) | 0.052 | >0.99 |  |
|  |  | T6 to T7 | -0.042 ( -0.176 to 0.092 ) | 0.044 | >0.99 |  |

<sup>a</sup> Adjusted for multiple comparisons using Bonferroni correction. $p$ -lvl ( $p$ -value level) = \* $p < 0.05$ , \*\* $p < 0.01$ , and \*\*\* = $p < 0.001$ ; CI = confidence interval; SLJ = standing long jump; DüMo = Düsseldorfer Modell; Mean Difference = mean difference based on the estimated marginal means; $p$ -lvl = significance level; SE = standard error; T1 = baseline measurements in September 2019; T2 = follow-up measurements in June 2020; T3 = follow-up measurements in September 2020; T4 = follow-up measurements in March 2021; T5 = follow-up measurements in June 2021; T6 = follow-up measurements in September 2021; T7 = follow-up measurements in June 2022.

No interaction effects were found over time (time*sports club: ⴄ_p_^2^= 0.006, *p* = 0.16) (Table R10 and Figure R10).

Changes in SLJ in the school groups: (subgroup: intervention and control groups SY21–22) No interaction effects were found over time (time*school group: ⴄ_p_^2^ = 0.005, *p* = 0.026; time*sports club*school group: ⴄ_p_^2^= 0.003, *p* = 0.63) (Tables R10 and R13 and Figure R11). No significant changes were found in the intervention or control groups during the observation period from September 2021 to June 2022 (I-A = +0.09 (95% CI, –0.06 to 0.23), *p* > 0.99; C-A = +0.01 (95% CI, –0.13 to 0.16), *p* > 0.99). Only in the control group without sports club membership was a “minimal” tendential decline detected between September 2021 and June 2022 (–0.01 SDS) (Table R13 and Figure R11).

**Figure R11.**
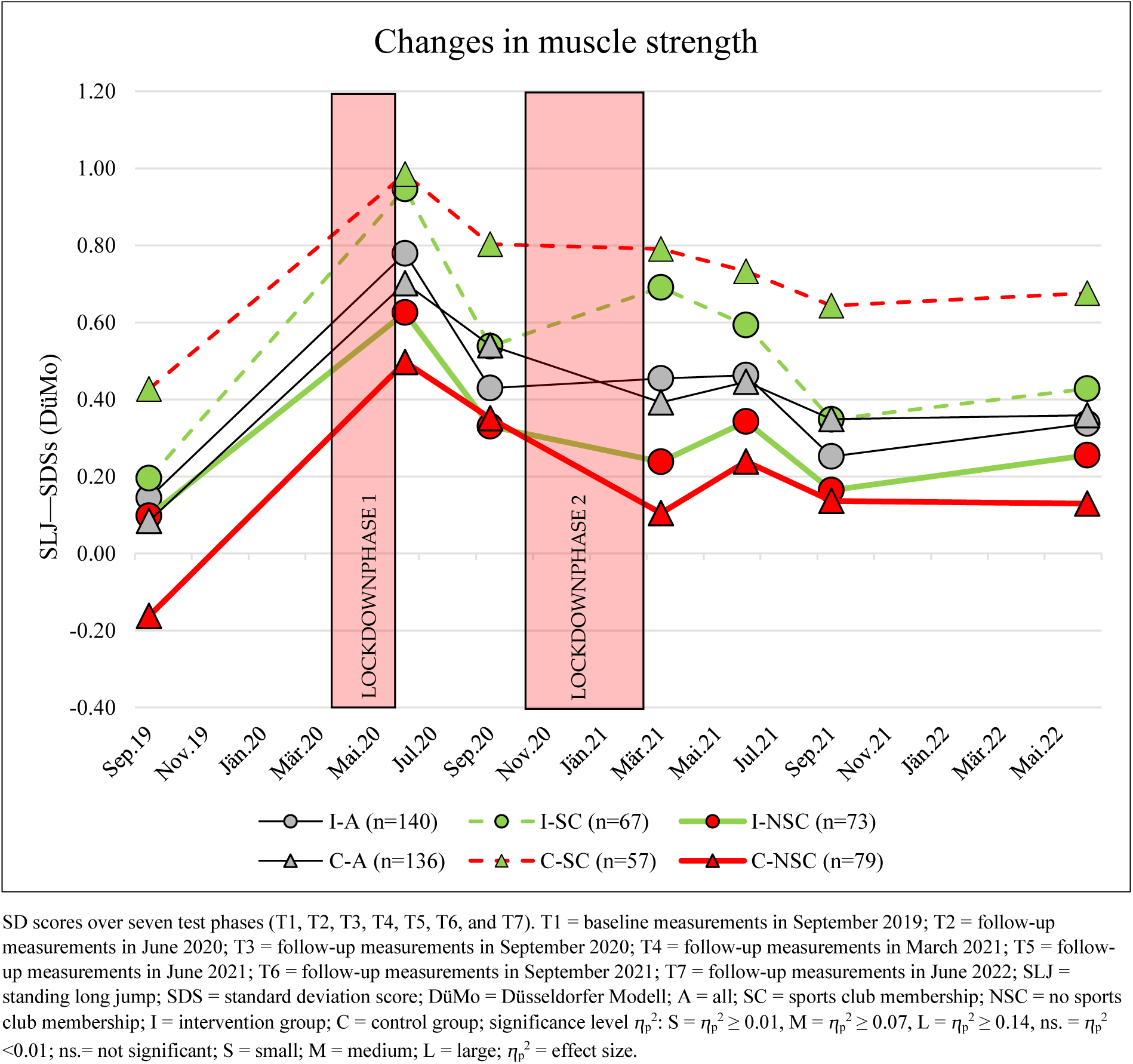
Changes in muscle strength from September 2019 to June 2022 in the subgroups: Intervention and control groups divided by sports club membership.

#### Upper Body Muscle Strength (MB1kg)

The medicine ball throw (MB1kg), the upper body strength parameter, showed significant changes with a small effect over the observation period (T1 to T7: main effect of time: ⴄ_p_^2^ = 0.035; *p* < 0.001) (Table R10 and Figures R5 and R12).

**Figure R12.**
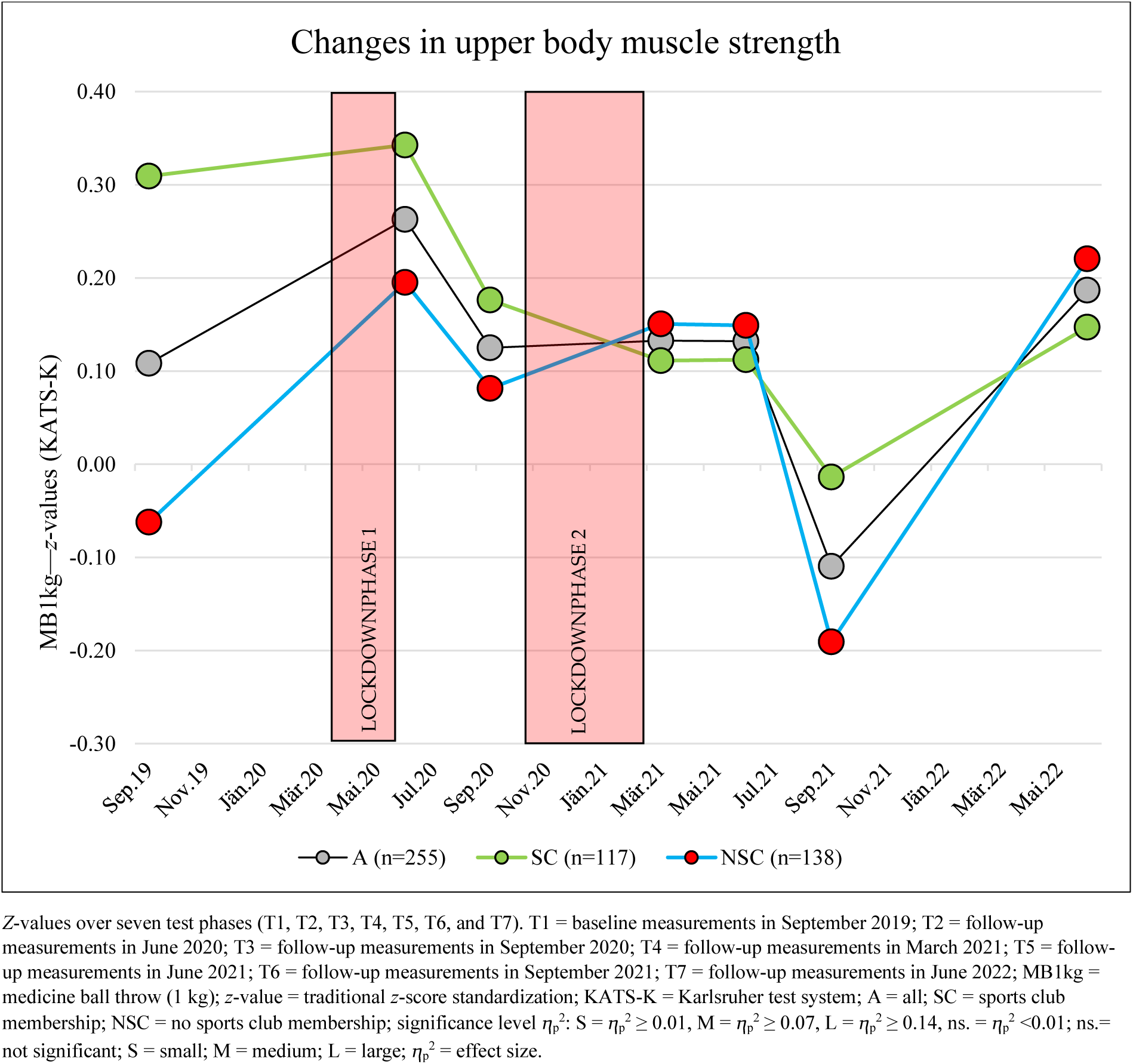
Changes in upper body muscle strength from September 2019 to June 2022.

An improvement in performance without scientific significance was observed in the medicine ball throw between September 2019 and June 2020 (+0.15 (95% CI, –0.05 to 0.33); *p* = 0.47). Declines in performance were observed during the school vacations of summer 2020 (–0.13 (95% CI, –0.31 to 0.03); *p* = 0.22) and summer 2021 (–0.24 (95% CI, –0.38 to –0.09); *p* < 0.001) (Tables R10 and R14 and Figure R12).

**Table R14.** Post hoc tests of MB1kg for the main effect of time and interactions for time*school group and time*sports club membership based on estimated marginal means.

| | | Pairwise Comparisons | Mean Difference (95% CI) | SE | $p$-Value<sub>a</sub> | $p$-lvl |
| --- | --- | --- | --- | --- | --- | --- |
| MB1kg z-value (KATS-K) | Time | T1 to T2 | −0.143 ( −0.334 to 0.048 ) | 0.062 | 0.470 |  |
|  |  | T1 to T3 | −0.003 ( −0.170 to 0.165 ) | 0.055 | >0.99 |  |
|  |  | T1 to T4 | −0.007 ( −0.199 to 0.186 ) | 0.063 | >0.99 |  |
|  |  | T1 to T5 | −0.007 ( −0.194 to 0.180 ) | 0.061 | >0.99 |  |
|  |  | T1 to T6 | 0.229 ( 0.066 to 0.393 ) | 0.053 | <0.001 | *** |
|  |  | T1 to T7 | −0.063 ( −0.259 to 0.133 ) | 0.064 | >0.99 |  |
|  |  | T2 to T3 | 0.140 ( −0.027 to 0.307 ) | 0.054 | 0.223 |  |
|  |  | T2 to T4 | 0.136 ( −0.024 to 0.297 ) | 0.052 | 0.203 |  |
|  |  | T2 to T5 | 0.136 ( −0.016 to 0.288 ) | 0.049 | 0.135 |  |
|  |  | T2 to T6 | 0.372 ( 0.206 to 0.538 ) | 0.054 | <0.001 | *** |
|  |  | T2 to T7 | 0.080 ( −0.077 to 0.237 ) | 0.051 | >0.99 |  |
|  |  | T3 to T4 | −0.004 ( −0.166 to 0.159 ) | 0.053 | >0.99 |  |
|  |  | T3 to T5 | −0.004 ( −0.168 to 0.160 ) | 0.053 | >0.99 |  |
|  |  | T3 to T6 | 0.232 ( 0.098 to 0.366 ) | 0.044 | <0.001 | *** |
|  |  | T3 to T7 | −0.060 ( −0.222 to 0.102 ) | 0.053 | >0.99 |  |
|  |  | T4 to T5 | 0.000 ( −0.144 to 0.143 ) | 0.047 | >0.99 |  |
|  |  | T4 to T6 | 0.236 ( 0.070 to 0.402 ) | 0.054 | <0.001 | *** |
|  |  | T4 to T7 | −0.056 ( −0.214 to 0.101 ) | 0.051 | >0.99 |  |
|  |  | T5 to T6 | 0.236 ( 0.089 to 0.383 ) | 0.048 | <0.001 | *** |
|  |  | T5 to T7 | −0.056 ( −0.206 to 0.094 ) | 0.049 | >0.99 |  |
|  |  | T6 to T7 | −0.292 ( −0.431 to −0.153 ) | 0.045 | <0.001 | *** |
|  | Time*School Group (Intervention Group) | T1 to T2 | −0.248 ( −0.517 to 0.020 ) | 0.087 | 0.102 |  |
|  |  | T1 to T3 | −0.068 ( −0.304 to 0.167 ) | 0.077 | >0.99 |  |
|  |  | T1 to T4 | −0.019 ( −0.289 to 0.252 ) | 0.088 | >0.99 |  |
|  |  | T1 to T5 | −0.049 ( −0.312 to 0.214 ) | 0.086 | >0.99 |  |
|  |  | T1 to T6 | 0.199 ( −0.031 to 0.428 ) | 0.075 | 0.178 |  |
|  |  | T1 to T7 | −0.143 ( −0.419 to 0.132 ) | 0.090 | >0.99 |  |
|  |  | T2 to T3 | 0.180 ( −0.055 to 0.415 ) | 0.076 | 0.404 |  |
|  |  | T2 to T4 | 0.230 ( 0.004 to 0.455 ) | 0.073 | 0.042 | * |
|  |  | T2 to T5 | 0.199 ( −0.014 to 0.413 ) | 0.070 | 0.095 |  |
|  |  | T2 to T6 | 0.447 ( 0.214 to 0.680 ) | 0.076 | <0.001 | *** |
|  |  | T2 to T7 | 0.105 ( −0.115 to 0.325 ) | 0.072 | >0.99 |  |
|  |  | T3 to T4 | 0.049 ( −0.179 to 0.278 ) | 0.074 | >0.99 |  |
|  |  | T3 to T5 | 0.019 ( −0.211 to 0.249 ) | 0.075 | >0.99 |  |
|  |  | T3 to T6 | 0.267 ( 0.078 to 0.456 ) | 0.061 | <0.001 | *** |
|  |  | T3 to T7 | −0.075 ( −0.302 to 0.152 ) | 0.074 | >0.99 |  |
|  |  | T4 to T5 | −0.030 ( −0.232 to 0.172 ) | 0.066 | >0.99 |  |
|  |  | T4 to T6 | 0.217 ( −0.016 to 0.451 ) | 0.076 | 0.096 |  |
|  |  | T4 to T7 | −0.124 ( −0.346 to 0.097 ) | 0.072 | >0.99 |  |
|  |  | T5 to T6 | 0.248 ( 0.042 to 0.454 ) | 0.067 | 0.006 | ** |
|  |  | T5 to T7 | −0.094 ( −0.305 to 0.117 ) | 0.069 | >0.99 |  |
|  |  | T6 to T7 | -0.342 ( -0.538 to -0.146 ) | 0.064 | <0.001 | *** |
|  | Time*School<br>Group (Control<br>Group) | T1 to T2 | -0.037 ( -0.309 to 0.234 ) | 0.088 | >0.99 |  |
|  |  | T1 to T3 | 0.063 ( -0.176 to 0.301 ) | 0.078 | >0.99 |  |
|  |  | T1 to T4 | 0.006 ( -0.268 to 0.280 ) | 0.089 | >0.99 |  |
|  |  | T1 to T5 | 0.035 ( -0.231 to 0.301 ) | 0.087 | >0.99 |  |
|  |  | T1 to T6 | 0.259 ( 0.027 to 0.492 ) | 0.076 | 0.015 | * |
|  |  | T1 to T7 | 0.017 ( -0.262 to 0.296 ) | 0.091 | >0.99 |  |
|  |  | T2 to T3 | 0.100 ( -0.138 to 0.337 ) | 0.077 | >0.99 |  |
|  |  | T2 to T4 | 0.043 ( -0.185 to 0.272 ) | 0.074 | >0.99 |  |
|  |  | T2 to T5 | 0.073 ( -0.143 to 0.289 ) | 0.070 | >0.99 |  |
|  |  | T2 to T6 | 0.297 ( 0.061 to 0.533 ) | 0.077 | 0.003 | ** |
|  |  | T2 to T7 | 0.055 ( -0.168 to 0.278 ) | 0.073 | >0.99 |  |
|  |  | T3 to T4 | -0.057 ( -0.288 to 0.175 ) | 0.075 | >0.99 |  |
|  |  | T3 to T5 | -0.027 ( -0.260 to 0.206 ) | 0.076 | >0.99 |  |
|  |  | T3 to T6 | 0.197 ( 0.006 to 0.388 ) | 0.062 | 0.037 | * |
|  |  | T3 to T7 | -0.045 ( -0.275 to 0.185 ) | 0.075 | >0.99 |  |
|  |  | T4 to T5 | 0.030 ( -0.175 to 0.234 ) | 0.067 | >0.99 |  |
|  |  | T4 to T6 | 0.254 ( 0.017 to 0.490 ) | 0.077 | 0.024 | * |
|  |  | T4 to T7 | 0.012 ( -0.213 to 0.236 ) | 0.073 | >0.99 |  |
|  |  | T5 to T6 | 0.224 ( 0.015 to 0.433 ) | 0.068 | 0.024 | * |
|  |  | T5 to T7 | -0.018 ( -0.231 to 0.195 ) | 0.070 | >0.99 |  |
|  |  | T6 to T7 | -0.242 ( -0.441 to -0.044 ) | 0.065 | 0.005 | ** |
|  | Time*Sports Club<br>(Yes) | T1 to T2 | -0.025 ( -0.306 to 0.256 ) | 0.092 | >0.99 |  |
|  |  | T1 to T3 | 0.139 ( -0.107 to 0.385 ) | 0.080 | >0.99 |  |
|  |  | T1 to T4 | 0.200 ( -0.084 to 0.483 ) | 0.092 | 0.662 |  |
|  |  | T1 to T5 | 0.199 ( -0.076 to 0.475 ) | 0.090 | 0.568 |  |
|  |  | T1 to T6 | 0.328 ( 0.088 to 0.569 ) | 0.078 | <0.001 | *** |
|  |  | T1 to T7 | 0.163 ( -0.126 to 0.452 ) | 0.094 | >0.99 |  |
|  |  | T2 to T3 | 0.164 ( -0.081 to 0.410 ) | 0.080 | 0.863 |  |
|  |  | T2 to T4 | 0.225 ( -0.011 to 0.461 ) | 0.077 | 0.080 |  |
|  |  | T2 to T5 | 0.225 ( 0.001 to 0.448 ) | 0.073 | 0.047 | * |
|  |  | T2 to T6 | 0.353 ( 0.109 to 0.597 ) | 0.080 | <0.001 | *** |
|  |  | T2 to T7 | 0.188 ( -0.042 to 0.419 ) | 0.075 | 0.271 |  |
|  |  | T3 to T4 | 0.060 ( -0.179 to 0.300 ) | 0.078 | >0.99 |  |
|  |  | T3 to T5 | 0.060 ( -0.181 to 0.301 ) | 0.079 | >0.99 |  |
|  |  | T3 to T6 | 0.189 ( -0.008 to 0.387 ) | 0.064 | 0.076 |  |
|  |  | T3 to T7 | 0.024 ( -0.214 to 0.262 ) | 0.078 | >0.99 |  |
|  |  | T4 to T5 | 0.000 ( -0.212 to 0.211 ) | 0.069 | >0.99 |  |
|  |  | T4 to T6 | 0.129 ( -0.116 to 0.373 ) | 0.080 | >0.99 |  |
|  |  | T4 to T7 | -0.037 ( -0.269 to 0.195 ) | 0.076 | >0.99 |  |
|  |  | T5 to T6 | 0.129 ( -0.087 to 0.345 ) | 0.070 | >0.99 |  |
|  |  | T5 to T7 | -0.036 ( -0.257 to 0.184 ) | 0.072 | >0.99 |  |
|  |  | T6 to T7 | -0.165 ( -0.370 to 0.040 ) | 0.067 | 0.297 |  |
|  | Time*Sports Club<br>(No) | T1 to T2 | -0.261 ( -0.519 to -0.002 ) | 0.084 | 0.046 | * |
|  |  | T1 to T3 | -0.145 ( -0.372 to 0.082 ) | 0.074 | >0.99 |  |
|  |  | T1 to T4 | -0.213 ( -0.473 to 0.048 ) | 0.085 | 0.270 |  |
|  |  | T1 to T5 | -0.213 ( -0.466 to 0.040 ) | 0.082 | 0.216 |  |
|  |  | T1 to T6 | 0.130 ( -0.092 to 0.351 ) | 0.072 | >0.99 |  |
|  |  | T1 to T7 | -0.289 ( -0.554 to -0.023 ) | 0.087 | 0.020 | * |
|  |  | T2 to T3 | 0.116 ( -0.110 to 0.342 ) | 0.074 | >0.99 |  |
|  |  | T2 to T4 | 0.048 ( -0.170 to 0.265 ) | 0.071 | >0.99 |  |
|  |  | T2 to T5 | 0.047 ( -0.158 to 0.253 ) | 0.067 | >0.99 |  |
|  |  | T2 to T6 | 0.390 ( 0.166 to 0.615 ) | 0.073 | <0.001 | *** |
|  |  | T2 to T7 | -0.028 ( -0.241 to 0.184 ) | 0.069 | >0.99 |  |
|  |  | T3 to T4 | -0.068 ( -0.288 to 0.152 ) | 0.072 | >0.99 |  |
|  |  | T3 to T5 | -0.068 ( -0.290 to 0.153 ) | 0.072 | >0.99 |  |
|  |  | T3 to T6 | 0.275 ( 0.093 to 0.456 ) | 0.059 | <0.001 | *** |
|  |  | T3 to T7 | -0.144 ( -0.363 to 0.075 ) | 0.071 | 0.936 |  |
|  |  | T4 to T5 | -0.001 ( -0.195 to 0.194 ) | 0.063 | >0.99 |  |
|  |  | T4 to T6 | 0.343 ( 0.118 to 0.567 ) | 0.073 | <0.001 | *** |
|  |  | T4 to T7 | -0.076 ( -0.290 to 0.137 ) | 0.070 | >0.99 |  |
|  |  | T5 to T6 | 0.343 ( 0.145 to 0.542 ) | 0.065 | <0.001 | *** |
|  |  | T5 to T7 | -0.076 ( -0.279 to 0.127 ) | 0.066 | >0.99 |  |
|  |  | T6 to T7 | -0.419 ( -0.608 to -0.230 ) | 0.062 | <0.001 | *** |

<sup>a</sup> Adjusted for multiple comparisons using Bonferroni correction. *p*-lvl (*p*-value level) = \* *p* < 0.05, \*\* *p* < 0.01, and \*\*\* *p* < 0.001. CI = confidence interval; MB1kg = medicine ball throw (1 kg); KATS-K = Karlsruher test system; Mean Different = mean difference based on the estimated marginal means; *p*-lvl = significance level; SE = standard error; T1 = baseline measurements in September 2019; T2 = follow-up measurements in June 2020; T3 = follow-up measurements in September 2020; T4 = follow-up measurements in March 2021; T5 = follow-up measurements in June 2021; T6 = follow-up measurements in September 2021; T7 = follow-up measurements in June 2022.

The observation period from September 2021 to June 2022 showed a significant improvement in performance (+0.30 (95% CI, 0.15–0.43); *p* < 0.001) (Table R14 and Figure R12).

Small interaction effects over time (time*sports club: ⴄ_p_^2^= 0.017, *p* < 0.001) were found (Tables R10 and R14 and Figures R5 and R12).

Changes in medicine ball throwing in the school groups: (subgroup: intervention and control groups SY21–22)

No interaction effects were found over time (time*school group: ⴄ_p_^2^ = 0.004, *p* = 0.42; time*sports club*school group: ⴄ_p_^2^= 0.004, *p* = 0.35) (Table R10 and Figure R13).

**Figure R13.**
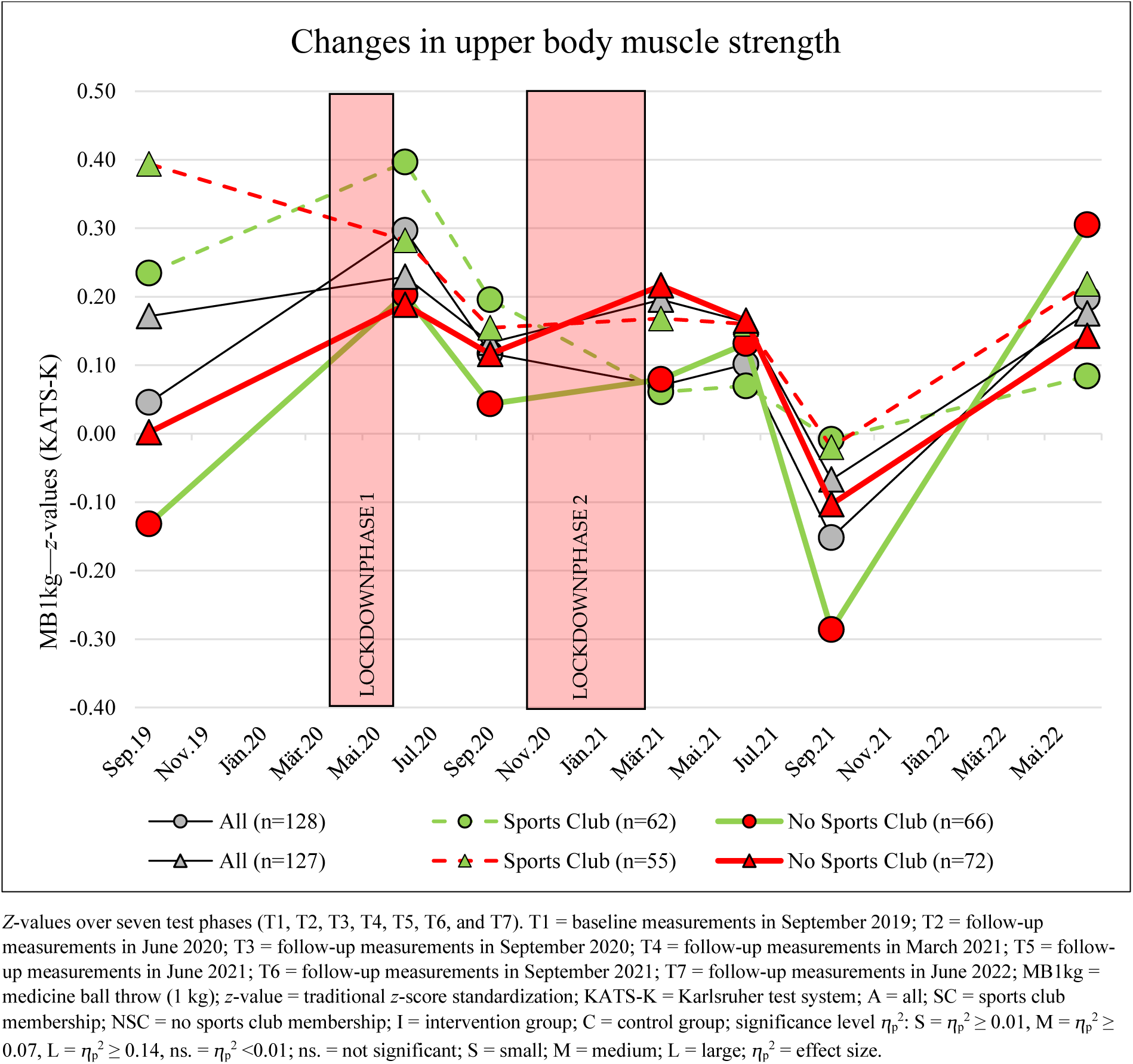
Changes in upper body muscle strength from September 2019 to June 2022 in the subgroups: Intervention and control groups divided by sports club membership.

Significant changes were observed between September 2021 and June 2022 in the intervention and control groups (I-A = +0.35 (95% CI, 0.15–0.54), *p* < 0.001; C-A = +0.25 (95% CI, 0.05–0.44), *p* = 0.005) (Table R14 and Figure R13).

The highest increase was observed between September 2021 and June 2022 in the intervention group without sports club membership (+0.60 SDS) (Table R14 and Figure R13).

#### Jumping Sideways (JS)

Significant changes were seen in jumping sideways (JS), the parameter for whole-body coordination and muscle endurance, with a small effect over the observation period (T1–T7: main effect of time: ⴄ_p_^2^ = 0.002; *p* < 0.001) (Table R10 and Figure R5).

At the end of the observation period (June 2022), no significant change from baseline (September 2019) was shown (+0.17 (95% CI, –0.04 to 0.39); *p* = 0.28) (Table R15 and Figure R14).

**Table R15.** Post hoc tests of JS for the main effect of time and interactions for time*school group and time*sports club membership based on estimated marginal means.

|  |  | Pairwise Comparisons | Mean Difference (95% CI) | SE | *p*-Value<sub>a</sub> | *p*-lvl |
| --- | --- | --- | --- | --- | --- | --- |
| JS *z*-value (GMT) | Time | T1 to T2 | 0.115 ( -0.118 to 0.348 ) | 0.079 | >0.99 |  |
|  |  | T1 to T3 | -0.164 ( -0.383 to 0.056 ) | 0.074 | 0.422 |  |
|  |  | T1 to T5 | -0.121 ( -0.345 to 0.103 ) | 0.076 | >0.99 |  |
|  |  | T1 to T6 | 0.058 ( -0.153 to 0.268 ) | 0.071 | >0.99 |  |
|  |  | T1 to T7 | -0.173 ( -0.389 to 0.043 ) | 0.073 | 0.278 |  |
|  |  | T2 to T3 | -0.278 ( -0.470 to -0.087 ) | 0.065 | <0.001 | *** |
|  |  | T2 to T5 | -0.236 ( -0.425 to -0.046 ) | 0.064 | 0.004 | ** |
|  |  | T2 to T6 | -0.057 ( -0.247 to 0.134 ) | 0.064 | >0.99 |  |
|  |  | T2 to T7 | -0.288 ( -0.471 to -0.104 ) | 0.062 | <0.001 | *** |
|  |  | T3 to T5 | 0.043 ( -0.125 to 0.210 ) | 0.057 | >0.99 |  |
|  |  | T3 to T6 | 0.221 ( 0.047 to 0.396 ) | 0.059 | 0.003 | ** |
|  |  | T3 to T7 | -0.009 ( -0.200 to 0.182 ) | 0.065 | >0.99 |  |
|  |  | T5 to T6 | 0.179 ( 0.020 to 0.338 ) | 0.054 | 0.015 | * |
|  |  | T5 to T7 | -0.052 ( -0.224 to 0.120 ) | 0.058 | >0.99 |  |
|  |  | T6 to T7 | -0.231 ( -0.384 to -0.078 ) | 0.052 | <0.001 | *** |
|  | Time*School Group (Intervention Group) | T1 to T2 | -0.102 ( -0.427 to 0.223 ) | 0.110 | >0.99 |  |
|  |  | T1 to T3 | -0.520 ( -0.826 to -0.214 ) | 0.103 | <0.001 | *** |
|  |  | T1 to T5 | -0.438 ( -0.751 to -0.126 ) | 0.106 | <0.001 | *** |
|  |  | T1 to T6 | -0.238 ( -0.532 to 0.055 ) | 0.099 | 0.255 |  |
|  |  | T1 to T7 | -0.410 ( -0.712 to -0.109 ) | 0.102 | 0.001 | ** |
|  |  | T2 to T3 | -0.418 ( -0.685 to -0.151 ) | 0.090 | <0.001 | *** |
|  |  | T2 to T5 | -0.336 ( -0.600 to -0.071 ) | 0.089 | 0.003 | ** |
|  |  | T2 to T6 | -0.136 ( -0.401 to 0.130 ) | 0.090 | >0.99 |  |
|  |  | T2 to T7 | -0.308 ( -0.564 to -0.052 ) | 0.086 | 0.007 | ** |
|  |  | T3 to T5 | 0.082 ( -0.152 to 0.316 ) | 0.079 | >0.99 |  |
|  |  | T3 to T6 | 0.282 ( 0.039 to 0.525 ) | 0.082 | 0.010 | * |
|  |  | T3 to T7 | 0.110 ( -0.157 to 0.376 ) | 0.090 | >0.99 |  |
|  |  | T5 to T6 | 0.200 ( -0.022 to 0.422 ) | 0.075 | 0.120 |  |
|  |  | T5 to T7 | 0.028 ( -0.212 to 0.268 ) | 0.081 | >0.99 |  |
|  |  | T6 to T7 | -0.172 ( -0.386 to 0.041 ) | 0.072 | 0.262 |  |
|  | Time*School Group (Control Group) | T1 to T2 | 0.332 ( -0.002 to 0.666 ) | 0.113 | 0.053 |  |
|  |  | T1 to T3 | 0.193 ( -0.121 to 0.507 ) | 0.106 | >0.99 |  |
|  |  | T1 to T5 | 0.196 ( -0.125 to 0.518 ) | 0.108 | >0.99 |  |
|  |  | T1 to T6 | 0.354 ( 0.052 to 0.655 ) | 0.102 | 0.009 | ** |
|  |  | T1 to T7 | 0.065 ( -0.245 to 0.374 ) | 0.105 | >0.99 |  |
|  |  | T2 to T3 | -0.139 ( -0.413 to 0.135 ) | 0.093 | >0.99 |  |
|  |  | T2 to T5 | -0.135 ( -0.407 to 0.136 ) | 0.092 | >0.99 |  |
|  |  | T2 to T6 | 0.022 ( -0.251 to 0.295 ) | 0.092 | >0.99 |  |
|  |  | T2 to T7 | -0.267 ( -0.530 to -0.004 ) | 0.089 | 0.043 | * |
|  |  | T3 to T5 | 0.003 ( -0.237 to 0.244 ) | 0.081 | >0.99 |  |
|  |  | T3 to T6 | 0.161 ( -0.089 to 0.410 ) | 0.084 | 0.865 |  |
|  |  | T3 to T7 | -0.128 ( -0.402 to 0.146 ) | 0.092 | >0.99 |  |
|  |  | T5 to T6 | 0.157 ( -0.071 to 0.385 ) | 0.077 | 0.627 |  |
|  |  | T5 to T7 | -0.132 ( -0.378 to 0.115 ) | 0.083 | >0.99 |  |
|  |  | T6 to T7 | -0.289 ( -0.508 to -0.070 ) | 0.074 | 0.002 | ** |
|  | Time*Sports Club (Yes) | T1 to T2 | 0.182 ( -0.164 to 0.528 ) | 0.117 | >0.99 |  |
|  |  | T1 to T3 | -0.239 ( -0.565 to 0.087 ) | 0.110 | 0.464 |  |
|  |  | T1 to T5 | -0.084 ( -0.417 to 0.250 ) | 0.112 | >0.99 |  |
|  |  | T1 to T6 | 0.082 ( -0.230 to 0.395 ) | 0.106 | >0.99 |  |
|  |  | T1 to T7 | -0.217 ( -0.538 to 0.104 ) | 0.108 | 0.693 |  |
|  |  | T2 to T3 | -0.421 ( -0.705 to -0.136 ) | 0.096 | <0.001 | *** |
|  |  | T2 to T5 | -0.265 ( -0.547 to 0.016 ) | 0.095 | 0.085 |  |
|  |  | T2 to T6 | -0.100 ( -0.383 to 0.183 ) | 0.096 | >0.99 |  |
|  |  | T2 to T7 | -0.399 ( -0.671 to -0.126 ) | 0.092 | <0.001 | *** |
|  |  | T3 to T5 | 0.155 ( -0.094 to 0.404 ) | 0.084 | 0.987 |  |
|  |  | T3 to T6 | 0.321 ( 0.062 to 0.580 ) | 0.087 | 0.004 | ** |
|  |  | T3 to T7 | 0.022 ( -0.262 to 0.306 ) | 0.096 | >0.99 |  |
|  |  | T5 to T6 | 0.166 ( -0.071 to 0.402 ) | 0.080 | 0.582 |  |
|  |  | T5 to T7 | -0.134 ( -0.389 to 0.122 ) | 0.086 | >0.99 |  |
|  |  | T6 to T7 | -0.299 ( -0.526 to -0.072 ) | 0.077 | 0.002 | ** |
|  | Time*Sports Club<br>(No) | T1 to T2 | 0.048 ( -0.264 to 0.360 ) | 0.105 | >0.99 |  |
|  |  | T1 to T3 | -0.088 ( -0.382 to 0.206 ) | 0.099 | >0.99 |  |
|  |  | T1 to T5 | -0.158 ( -0.458 to 0.142 ) | 0.101 | >0.99 |  |
|  |  | T1 to T6 | 0.034 ( -0.248 to 0.315 ) | 0.095 | >0.99 |  |
|  |  | T1 to T7 | -0.129 ( -0.418 to 0.161 ) | 0.098 | >0.99 |  |
|  |  | T2 to T3 | -0.136 ( -0.392 to 0.120 ) | 0.087 | >0.99 |  |
|  |  | T2 to T5 | -0.206 ( -0.460 to 0.048 ) | 0.086 | 0.253 |  |
|  |  | T2 to T6 | -0.014 ( -0.269 to 0.241 ) | 0.086 | >0.99 |  |
|  |  | T2 to T7 | -0.176 ( -0.422 to 0.069 ) | 0.083 | 0.517 |  |
|  |  | T3 to T5 | -0.070 ( -0.294 to 0.154 ) | 0.076 | >0.99 |  |
|  |  | T3 to T6 | 0.122 ( -0.112 to 0.355 ) | 0.079 | >0.99 |  |
|  |  | T3 to T7 | -0.040 ( -0.296 to 0.216 ) | 0.086 | >0.99 |  |
|  |  | T5 to T6 | 0.192 ( -0.021 to 0.405 ) | 0.072 | 0.121 |  |
|  |  | T5 to T7 | 0.030 ( -0.201 to 0.260 ) | 0.078 | >0.99 |  |
|  |  | T6 to T7 | -0.162 ( -0.367 to 0.043 ) | 0.069 | 0.296 |  |

<sup>a</sup> Adjusted for multiple comparisons using Bonferroni correction. *p*-lvl (*p*-value level) = \* *p* < 0.05, \*\* *p* < 0.01, and \*\*\* *p* < 0.001; CI = confidence interval; JS = jumping sideways; GMT = German Motor Test; Mean Difference = mean difference based on the estimated marginal means; *p*-lvl = significance level; SE = standard error; T1 = baseline measurements in September 2019; T2 = follow-up measurements in June 2020; T3 = follow-up measurements in September 2020; T5 = follow-up measurements in June 2021; T6 = follow-up measurements in September 2021; T7 = follow-up measurements in June 2022.

**Figure R14.**
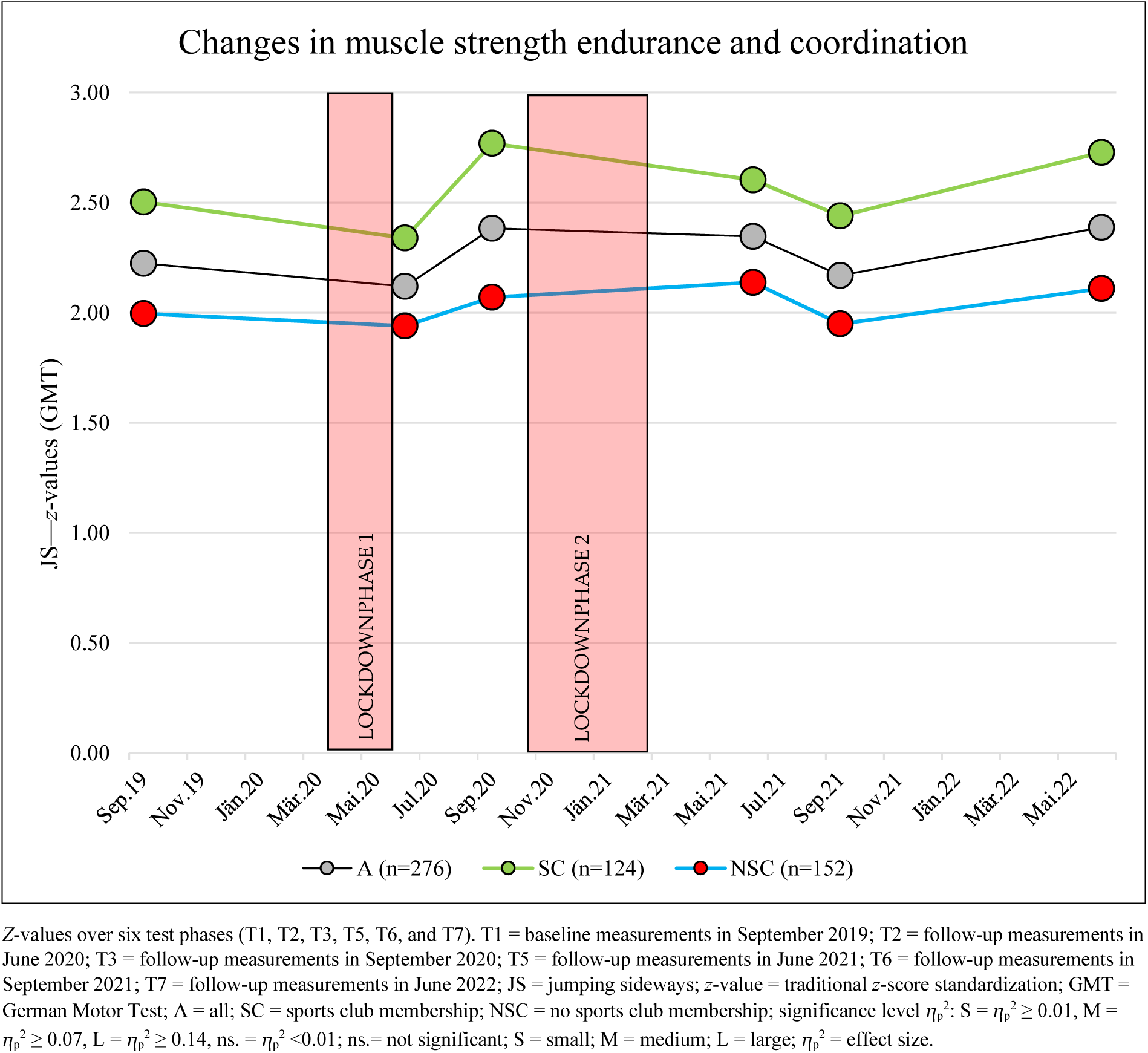
Changes in muscle strength endurance and coordination from September 2019 to June 2022.

No significant change was observed between September 2019 and June 2020 (–0.10 (95% CI, –0.35 to 0.12); *p* > 0.99). In the following observation period (June 2020 to September 2020), a significant improvement in performance was observed (+0.26 (95% CI, 0.09–0.47); *p* < 0.001). No changes in performance were shown in SY 2020/2021 (September 2020 to June 2021) (–0.03 (95% CI, –0.21 to 0.13); *p* > 0.99). From June 2021 to September 2021, a significant reduction was found (–0.03 (95% CI, –0.21 to 0.13); *p* > 0.99) (Table R15 and Figure R14).

Finally, between September 2021 and June 2022, a strong improvement in jumping sideways was detected (+0.22 (95% CI, 0.08–0.38); *p* < 0.001) (Table R15 and Figure R14).

No interaction effects were found over time (time*sports club: ⴄ_p_^2^= 0.005; *p* = 0.26) (Tables R10 and R11 and Figures R5 and R14).

Changes in JS in the school groups: (subgroup: intervention and control groups SY21–22) Small interaction effects over time (time*school group: ⴄ_p_^2^= 0.027, *p* < 0.001) were found, which were mainly related to the time before the sports intervention (September 2019 to September 2021) (Table R10 and Figure R5).

Significant changes were found in the control group during the observation period from September 2021 to June 2022 (C-A = +0.26 (95% CI, 0.07–0.51), *p* = 0.002). These improvements were more than threefold higher in those children with sports club membership (+0.43 SDS) than in those without (+0.14). No changes in performance were demonstrated at this time point in the intervention group (I-A = +0.18 (95% CI, –0.04 to 0.39), *p* = 0.26) (Table R15 and Figure R15).

#### Flexibility (VSR)

Flexibility (VSR) showed significant changes, with a medium effect over the observation period (T1–T7: main effect of time: ⴄ_p_^2^ = 0.045; *p* < 0.001) (Tables R10 and R16 and Figures R5 and R16).

**Table R16.** Post hoc tests of VSR for the main effect of time and interactions for time*school group and time*sports club membership based on estimated marginal means.

|  |  | Pairwise Comparisons | Mean Difference (95% CI) |  | SE | p-Value<sub>a</sub> | p-lvl |
| --- | --- | --- | --- | --- | --- | --- | --- |
| VSR SDS (MAKFIT) | Time | T1 to T2 | 0.074 | ( -0.082 to 0.229 ) | 0.053 | >0.99 |  |
|  |  | T1 to T3 | -0.006 | ( -0.160 to 0.148 ) | 0.052 | >0.99 |  |
|  |  | T1 to T5 | 0.369 | ( 0.186 to 0.552 ) | 0.062 | <0.001 | *** |
|  |  | T1 to T6 | 0.150 | ( -0.009 to 0.308 ) | 0.054 | 0.083 |  |
|  |  | T1 to T7 | 0.197 | ( 0.004 to 0.390 ) | 0.065 | 0.040 | * |
|  |  | T2 to T3 | -0.080 | ( -0.221 to 0.062 ) | 0.048 | >0.99 |  |
|  |  | T2 to T5 | 0.295 | ( 0.111 to 0.479 ) | 0.062 | <0.001 | *** |
|  |  | T2 to T6 | 0.076 | ( -0.087 to 0.239 ) | 0.055 | >0.99 |  |
|  |  | T2 to T7 | 0.124 | ( -0.064 to 0.311 ) | 0.063 | 0.785 |  |
|  |  | T3 to T5 | 0.375 | ( 0.219 to 0.530 ) | 0.053 | <0.001 | *** |
|  |  | T3 to T6 | 0.156 | ( 0.002 to 0.309 ) | 0.052 | 0.044 | * |
|  |  | T3 to T7 | 0.203 | ( 0.031 to 0.375 ) | 0.058 | 0.008 | ** |
|  |  | T5 to T6 | -0.219 | ( -0.364 to -0.075 ) | 0.049 | <0.001 | *** |
|  |  | T5 to T7 | -0.172 | ( -0.355 to 0.012 ) | 0.062 | 0.090 |  |
|  |  | T6 to T7 | 0.048 | ( -0.113 to 0.209 ) | 0.054 | >0.99 |  |
|  | Time*School<br>Group<br>(Intervention<br>Group) | T1 to T2 | 0.097 | ( -0.120 to 0.314 ) | 0.073 | >0.99 |  |
|  |  | T1 to T3 | 0.028 | ( -0.186 to 0.243 ) | 0.073 | >0.99 |  |
|  |  | T1 to T5 | 0.438 | ( 0.183 to 0.693 ) | 0.086 | <0.001 | *** |
|  |  | T1 to T6 | 0.191 | ( -0.030 to 0.413 ) | 0.075 | 0.164 |  |
|  |  | T1 to T7 | 0.158 | ( -0.112 to 0.427 ) | 0.091 | >0.99 |  |
|  |  | T2 to T3 | -0.069 | ( -0.266 to 0.129 ) | 0.067 | >0.99 |  |
|  |  | T2 to T5 | 0.341 | ( 0.084 to 0.598 ) | 0.087 | 0.001 | ** |
|  |  | T2 to T6 | 0.094 | ( -0.133 to 0.321 ) | 0.077 | >0.99 |  |
|  |  | T2 to T7 | 0.061 | ( -0.202 to 0.323 ) | 0.088 | >0.99 |  |
|  |  | T3 to T5 | 0.410 | ( 0.193 to 0.627 ) | 0.073 | <0.001 | *** |
|  |  | T3 to T6 | 0.163 | ( -0.051 to 0.377 ) | 0.072 | 0.374 |  |
|  |  | T3 to T7 | 0.129 | ( -0.111 to 0.369 ) | 0.081 | >0.99 |  |
|  |  | T5 to T6 | -0.247 | ( -0.448 to -0.045 ) | 0.068 | 0.005 | ** |
|  |  | T5 to T7 | -0.280 | ( -0.536 to -0.024 ) | 0.086 | 0.020 | * |
|  |  | T6 to T7 | -0.034 | ( -0.259 to 0.191 ) | 0.076 | >0.99 |  |
|  | Time*School<br>Group (Control<br>Group) | T1 to T2 | 0.050 | ( -0.173 to 0.273 ) | 0.075 | >0.99 |  |
|  |  | T1 to T3 | -0.041 | ( -0.261 to 0.180 ) | 0.075 | >0.99 |  |
|  |  | T1 to T5 | 0.300 | ( 0.037 to 0.562 ) | 0.089 | 0.012 | * |
|  |  | T1 to T6 | 0.108 | ( -0.120 to 0.335 ) | 0.077 | >0.99 |  |
|  |  | T1 to T7 | 0.237 | ( -0.040 to 0.513 ) | 0.093 | 0.176 |  |
|  |  | T2 to T3 | -0.091 | ( -0.293 to 0.112 ) | 0.068 | >0.99 |  |
|  |  | T2 to T5 | 0.249 | ( -0.014 to 0.513 ) | 0.089 | 0.082 |  |
|  |  | T2 to T6 | 0.058 | ( -0.176 to 0.291 ) | 0.079 | >0.99 |  |
|  |  | T2 to T7 | 0.187 | ( -0.082 to 0.456 ) | 0.091 | 0.613 |  |
|  |  | T3 to T5 | 0.340 | ( 0.117 to 0.563 ) | 0.075 | <0.001 | *** |
|  |  | T3 to T6 | 0.148 | ( -0.072 to 0.368 ) | 0.074 | 0.701 |  |
|  |  | T3 to T7 | 0.278 | ( 0.031 to 0.524 ) | 0.083 | 0.015 | * |
|  |  | T5 to T6 | -0.192 | ( -0.399 to 0.015 ) | 0.070 | 0.097 |  |
|  |  | T5 to T7 | -0.063 | ( -0.325 to 0.200 ) | 0.089 | >0.99 |  |
|  |  | T6 to T7 | 0.129 | ( -0.102 to 0.360 ) | 0.078 | >0.99 |  |
|  | Time*Sports Club<br>(Yes) | T1 to T2 | 0.077 | ( -0.154 to 0.308 ) | 0.078 | >0.99 |  |
|  |  | T1 to T3 | -0.012 | ( -0.241 to 0.217 ) | 0.077 | >0.99 |  |
|  |  | T1 to T5 | 0.255 | ( -0.017 to 0.527 ) | 0.092 | 0.089 |  |
|  |  | T1 to T6 | 0.131 | ( -0.104 to 0.367 ) | 0.080 | >0.99 |  |
|  |  | T1 to T7 | 0.187 | ( -0.100 to 0.474 ) | 0.097 | 0.819 |  |
|  |  | T2 to T3 | -0.089 | ( -0.299 to 0.121 ) | 0.071 | >0.99 |  |
|  |  | T2 to T5 | 0.178 | ( -0.096 to 0.451 ) | 0.092 | 0.828 |  |
|  |  | T2 to T6 | 0.054 | ( -0.187 to 0.296 ) | 0.082 | >0.99 |  |
|  |  | T2 to T7 | 0.110 | ( -0.169 to 0.389 ) | 0.094 | >0.99 |  |
|  |  | T3 to T5 | 0.267 | ( 0.035 to 0.498 ) | 0.078 | 0.011 | * |
|  |  | T3 to T6 | 0.143 | ( -0.085 to 0.371 ) | 0.077 | 0.963 |  |
|  |  | T3 to T7 | 0.199 | ( -0.057 to 0.455 ) | 0.086 | 0.331 |  |
|  |  | T5 to T6 | -0.123 | ( -0.338 to 0.091 ) | 0.073 | >0.99 |  |
|  |  | T5 to T7 | -0.068 | ( -0.340 to 0.205 ) | 0.092 | >0.99 |  |
|  |  | T6 to T7 | 0.056 | ( -0.184 to 0.295 ) | 0.081 | >0.99 |  |
|  | Time*Sports Club<br>(No) | T1 to T2 | 0.070 | ( -0.138 to 0.279 ) | 0.070 | >0.99 |  |
|  |  | T1 to T3 | 0.000 | ( -0.206 to 0.206 ) | 0.070 | >0.99 |  |
|  |  | T1 to T5 | 0.483 | ( 0.238 to 0.728 ) | 0.083 | <0.001 | *** |
|  |  | T1 to T6 | 0.168 | ( -0.044 to 0.380 ) | 0.072 | 0.298 |  |
|  |  | T1 to T7 | 0.208 | ( -0.051 to 0.466 ) | 0.087 | 0.271 |  |
|  |  | T2 to T3 | -0.071 | ( -0.260 to 0.119 ) | 0.064 | >0.99 |  |
|  |  | T2 to T5 | 0.412 | ( 0.166 to 0.659 ) | 0.083 | <0.001 | *** |
|  |  | T2 to T6 | 0.097 | ( -0.120 to 0.315 ) | 0.074 | >0.99 |  |
|  |  | T2 to T7 | 0.137 | ( -0.114 to 0.389 ) | 0.085 | >0.99 |  |
|  |  | T3 to T5 | 0.483 | ( 0.275 to 0.691 ) | 0.070 | <0.001 | *** |
|  |  | T3 to T6 | 0.168 | ( -0.037 to 0.374 ) | 0.069 | 0.239 |  |
|  |  | T3 to T7 | 0.208 | ( -0.023 to 0.438 ) | 0.078 | 0.119953 |  |
|  |  | T5 to T6 | -0.315 | ( -0.508 to -0.121 ) | 0.065 | <0.001 | *** |
|  |  | T5 to T7 | -0.275 | ( -0.521 to -0.030 ) | 0.083 | 0.016 | * |
|  |  | T6 to T7 | 0.040 | ( -0.176 to 0.255 ) | 0.073 | >0.99 |  |

<sup>a</sup> Adjusted for multiple comparisons using Bonferroni correction. *p*-lvl (*p*-value level) = \* *p* < 0.05, \*\* *p* < 0.01, and \*\*\* *p* < 0.001; CI = confidence interval; VSR = V sit-and-reach test; MAKFIT = Macedonia norm values; Mean Difference = mean difference based on the estimated marginal means; *p*-lvl = significance level; SE = standard error; T1 = baseline measurements in September 2019; T2 = follow-up measurements in June 2020; T3 = follow-up measurements in September 2020; T5 = follow-up measurements in June 2021; T6 = follow-up measurements in September 2021; T7 = follow-up measurements in June 2022.

**Figure R15.**
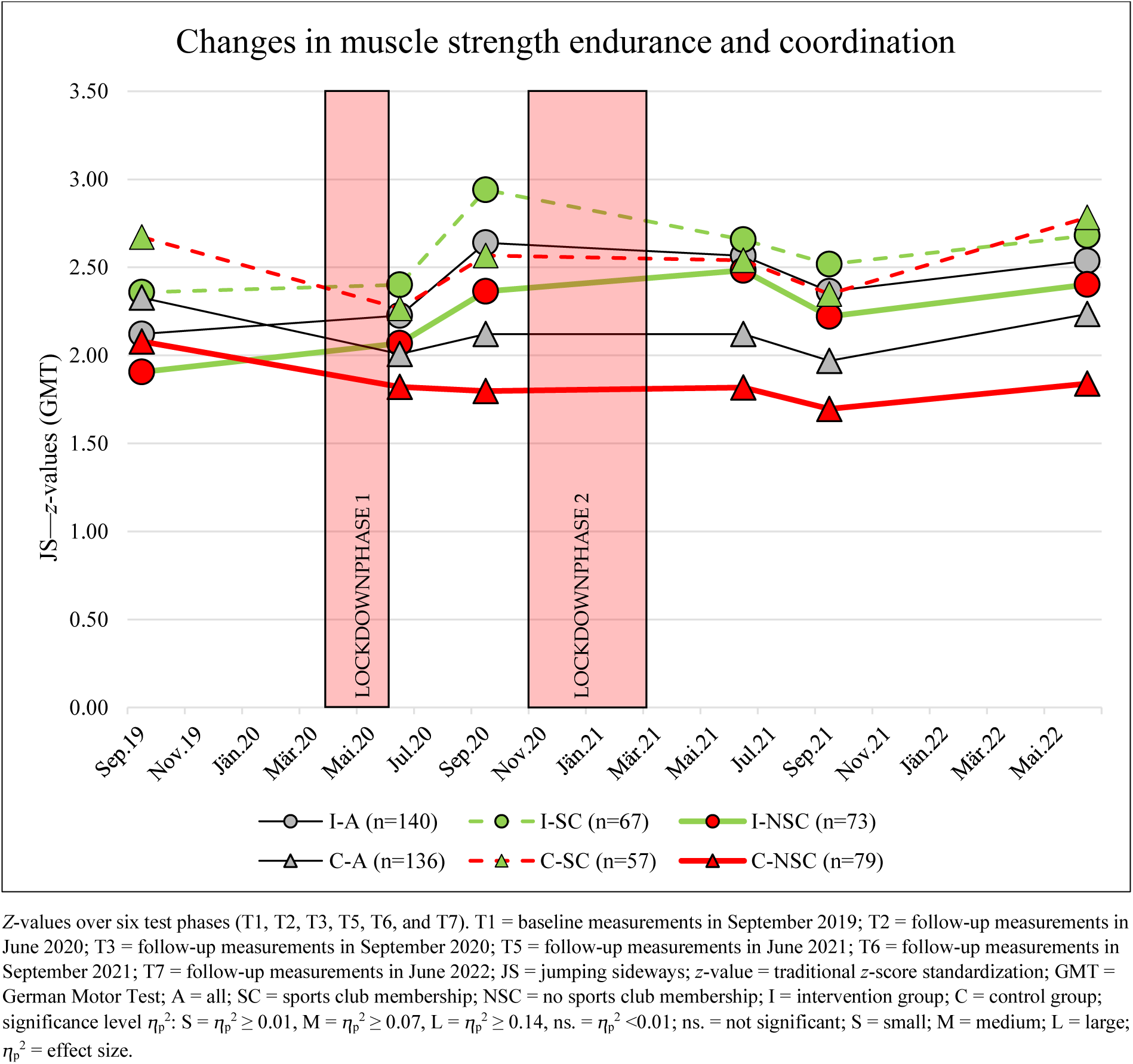
Changes in muscle strength endurance and coordination from September 2019 to June 2022 in the subgroups: Intervention and control groups divided by sports club membership.

**Figure R16.**
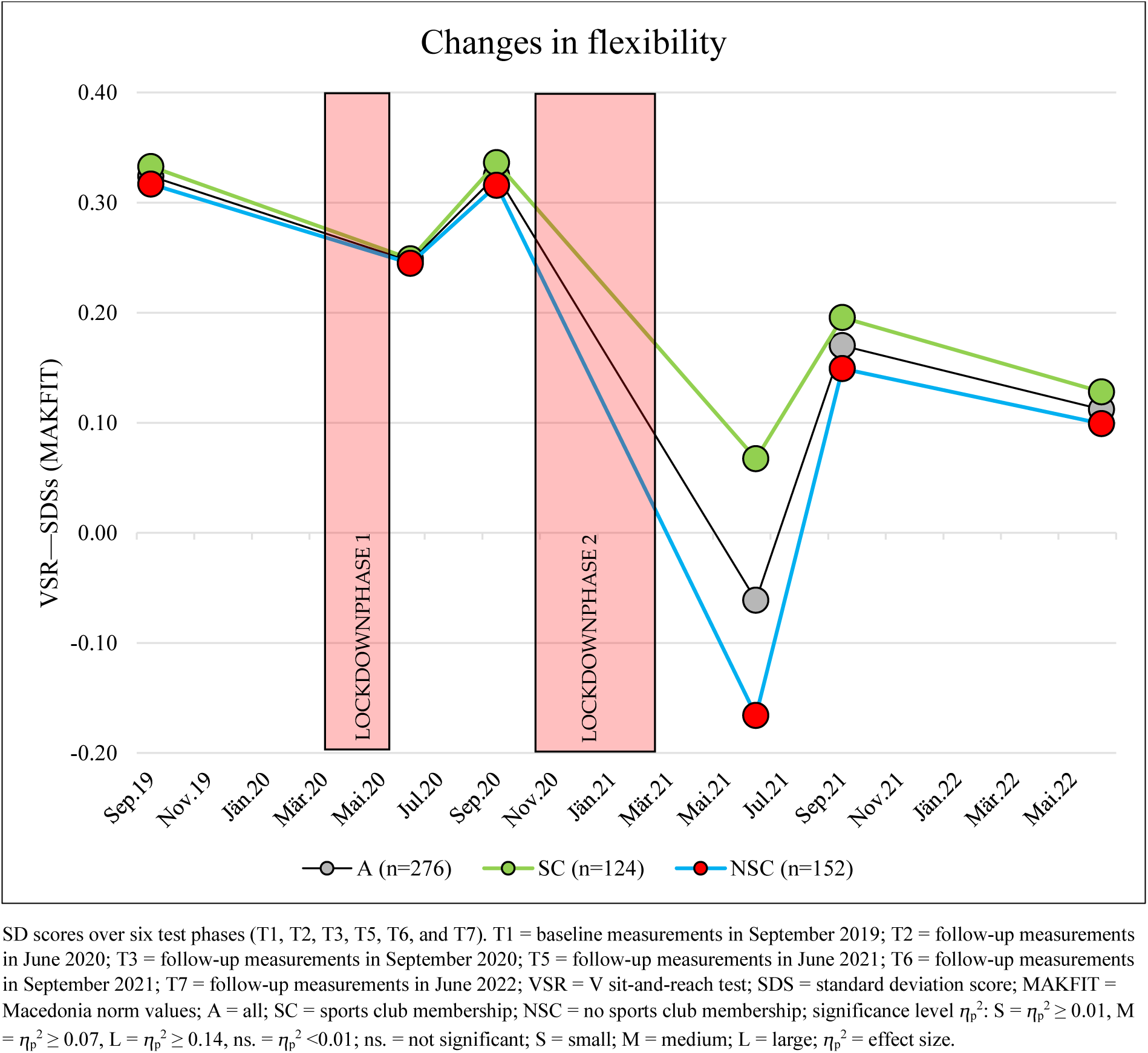
Changes in flexibility from September 2019 to June 2022.

At the end of the observation period (June 2022), a marginally significant worsening from baseline (September 2019) was shown (–0.21 (95% CI, –0.00 to –0.39); *p* = 0.041) (Table R16 and Figure R16).

At T5 (June 2021), a significantly significant change (*p* < 0.001) was observed compared to other test time points (Table R16 and Figure R16).

No significant change in flexibility was found between September 2021 and June 2022 (–0.06 (95% CI, –0.21 to 0.11); *p* > 0.99). (Table R16 and Figure R16).

No interaction effects were detected over time (time*sports club: ⴄ_p_^2^ = 0.005, *p* = 0.28) (Tables R10 and R16 and Figures R5 and R16).

Changes in VSR in the school groups: (subgroup: intervention and control groups SY21–22)

No interaction effects were found over time (time*school group: ⴄ_p_^2^ = 0.003, *p* = 0.48; time*sports club*school group: ⴄ_p_^2^ = 0.003, *p* = 0.57) and no significant changes were monitored during the observation period from September 2021 to June 2022 in the intervention and control groups (I-A = +0.03 (95% CI, –0.19 to 0.26); *p* > 0.99; C-A = –0.14 (95% CI, –0.36 to 0.10); *p* > 0.99) (Tables R10 and R16 and Figures R5 and R17).

**Figure R17.**
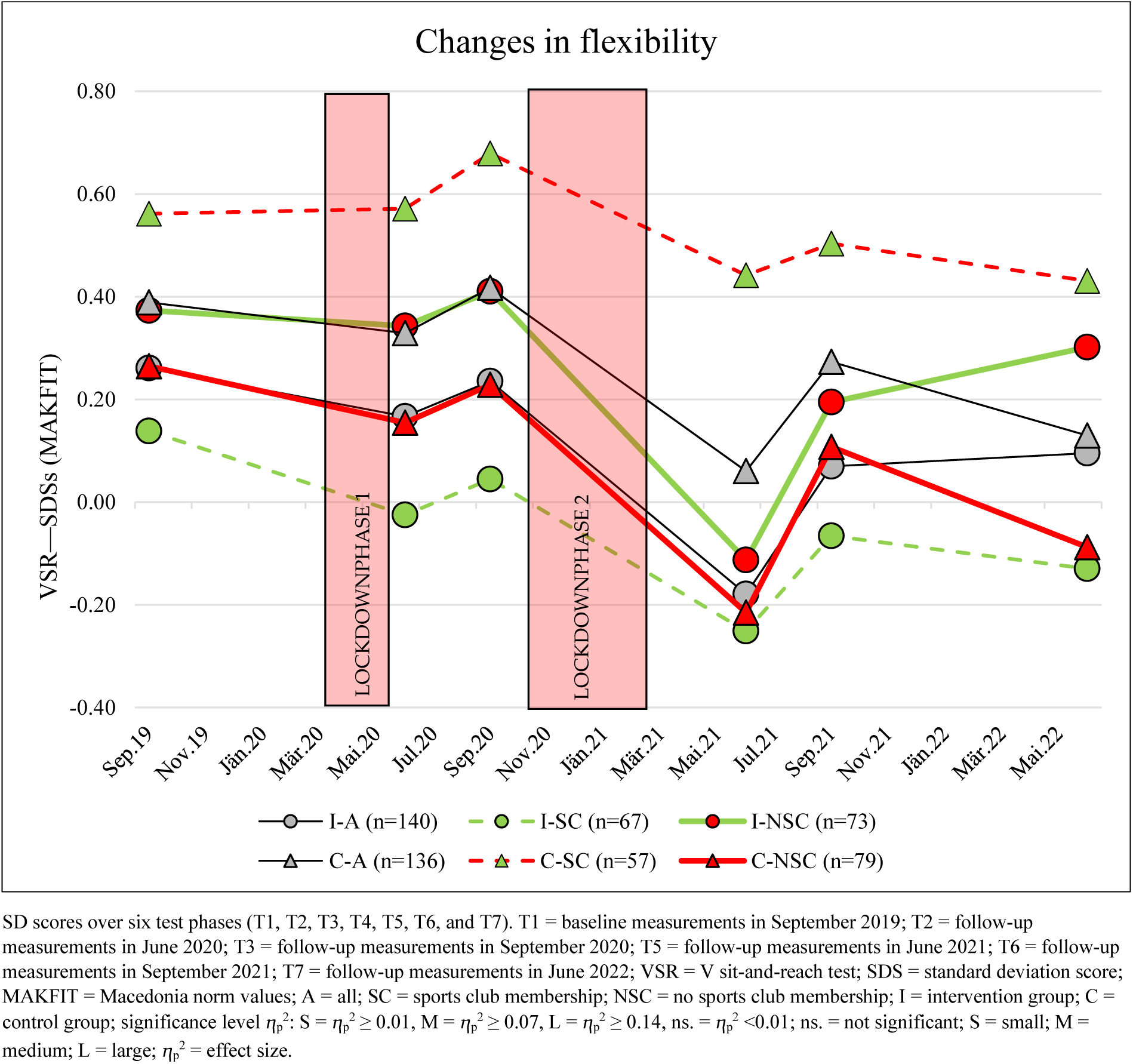
Changes in flexibility from September 2019 to June 2022 in the subgroups: Intervention and control groups divided by sports club membership.

Significant changes were found in the control group during the observation period from September 2021 to June 2022 (C-A = +0.26 (95% CI, 0.07–0.51), *p* = 0.002). These improvements were more than threefold higher in those children with sports club membership (+0.43 SDS) than those without (+0.14). No changes in performance were demonstrated at this time point in the intervention group (I-A = +0.18 (95% CI, –0.04 to 0.39), *p* = 0.26) (Table R16 and Figure R17).

There was a different trend for children who had no sports club membership (intervention group: VSR SDS = +0.11; control group: VSR SDS = –0.20) (Table R16 and Figure R17).

#### Balance (SLS-L and SLS-R)

For the one-leg stand, both legs showed significant changes with large effects during the observation period from September 2019 to June 2022 (main effect of time: SLS-L: ⴄ_p_^2^ = 0.176; *p* < 0.001; SLS-R: ⴄ_p_^2^= 0.126; *p* < 0.001) (Table R10 and Figure R18).

**Figure R18.**
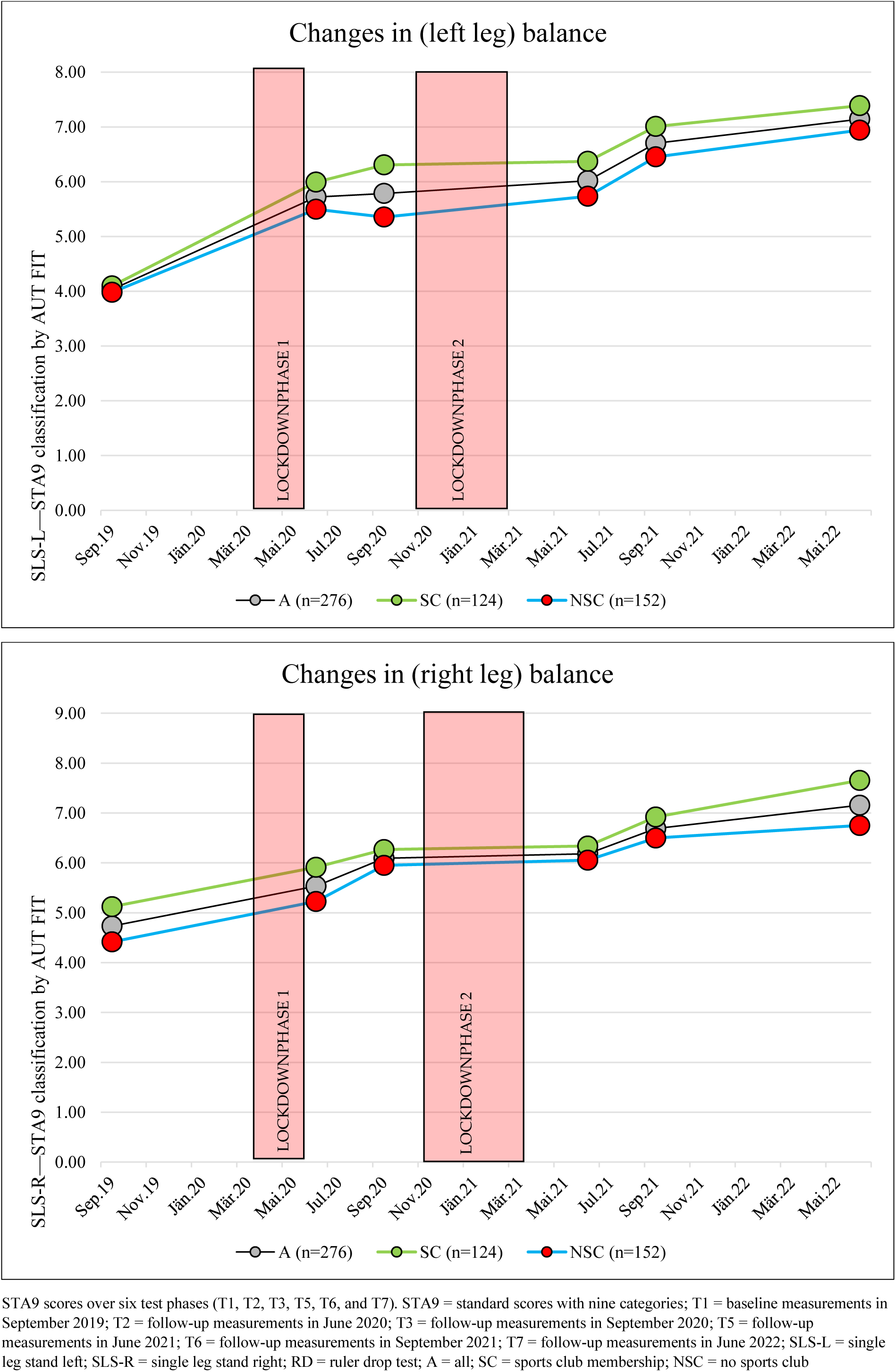
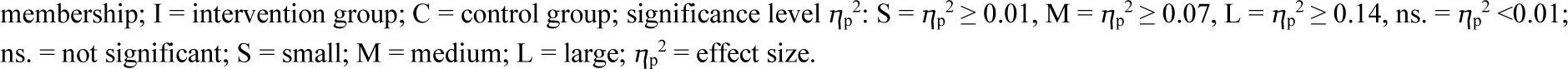
Changes in balance from September 2019 to June 2022.

No interaction effects were found over time (time*school group: SLS-L: ⴄ_p_^2^ = 0.003; *p* = 0.50; SLS-R: ⴄ_p_^2^= 0.003; *p* = 0.56) (Table R10 and Figure R18).

Throughout the whole observation time period (September 2019 to June 2022), a continuous increase in single leg standing performance was observed for both variants (left and right leg) of the single leg stand (Tables R10, R17, and R18 and Figure R18).

**Table R17.** Post hoc tests of SLS-L for the main effect of time and interactions for time*school group and time*sports club membership based on estimated marginal means.

|  |  | Pairwise Comparisons | Mean Difference (95% CI) | SE | p-Value<sup>a</sup> | p-lvl |
| --- | --- | --- | --- | --- | --- | --- |
| SLS-L (points) | Time | T1 to T2 | -1.684 ( -2.270 to -1.097 ) | 0.198 | <0.001 | *** |
|  |  | T1 to T3 | -1.760 ( -2.386 to -1.134 ) | 0.211 | <0.001 | *** |
|  |  | T1 to T5 | -1.991 ( -2.651 to -1.332 ) | 0.223 | <0.001 | *** |
|  |  | T1 to T6 | -2.688 ( -3.319 to -2.057 ) | 0.213 | <0.001 | *** |
|  |  | T1 to T7 | -3.107 ( -3.731 to -2.484 ) | 0.211 | <0.001 | *** |
|  |  | T2 to T3 | -0.076 ( -0.611 to 0.459 ) | 0.181 | >0.99 |  |
|  |  | T2 to T5 | -0.307 ( -0.873 to 0.259 ) | 0.191 | >0.99 |  |
|  |  | T2 to T6 | -1.004 ( -1.556 to -0.453 ) | 0.186 | <0.001 | *** |
|  |  | T2 to T7 | -1.424 ( -2.033 to -0.815 ) | 0.206 | <0.001 | *** |
|  |  | T3 to T5 | -0.231 ( -0.818 to 0.356 ) | 0.198 | >0.99 |  |
|  |  | T3 to T6 | -0.928 ( -1.513 to -0.343 ) | 0.198 | <0.001 | *** |
|  |  | T3 to T7 | -1.348 ( -1.948 to -0.747 ) | 0.203 | <0.001 | *** |
|  |  | T5 to T6 | -0.697 ( -1.262 to -0.132 ) | 0.191 | 0.005 | ** |
|  |  | T5 to T7 | -1.116 ( -1.703 to -0.530 ) | 0.198 | <0.001 | *** |
|  |  | T6 to T7 | -0.419 ( -0.952 to 0.113 ) | 0.180 | 0.306 |  |
|  | Time*School Group (Intervention Group) | T1 to T2 | -2.157 ( -2.975 to -1.339 ) | 0.276 | <0.001 | *** |
|  |  | T1 to T3 | -1.924 ( -2.797 to -1.050 ) | 0.295 | <0.001 | *** |
|  |  | T1 to T5 | -1.969 ( -2.889 to -1.049 ) | 0.311 | <0.001 | *** |
|  |  | T1 to T6 | -2.824 ( -3.704 to -1.944 ) | 0.297 | <0.001 | *** |
|  |  | T1 to T7 | -3.309 ( -4.179 to -2.439 ) | 0.294 | <0.001 | *** |
|  |  | T2 to T3 | 0.234 ( -0.513 to 0.981 ) | 0.252 | >0.99 |  |
|  |  | T2 to T5 | 0.188 ( -0.601 to 0.978 ) | 0.267 | >0.99 |  |
|  |  | T2 to T6 | -0.667 ( -1.436 to 0.102 ) | 0.260 | 0.161 |  |
|  |  | T2 to T7 | -1.152 ( -2.002 to -0.302 ) | 0.287 | 0.001 | ** |
|  |  | T3 to T5 | -0.046 ( -0.864 to 0.773 ) | 0.277 | >0.99 |  |
|  |  | T3 to T6 | -0.901 ( -1.717 to -0.085 ) | 0.276 | 0.018 | * |
|  |  | T3 to T7 | -1.386 ( -2.224 to -0.547 ) | 0.283 | <0.001 | *** |
|  |  | T5 to T6 | -0.855 ( -1.644 to -0.067 ) | 0.266 | 0.022 | * |
|  |  | T5 to T7 | -1.340 ( -2.158 to -0.522 ) | 0.276 | <0.001 | *** |
|  |  | T6 to T7 | -0.485 ( -1.227 to 0.258 ) | 0.251 | 0.815 |  |
|  | Time*School Group (Control Group) | T1 to T2 | -1.210 ( -2.051 to -0.370 ) | 0.284 | <0.001 | *** |
|  |  | T1 to T3 | -1.596 ( -2.493 to -0.699 ) | 0.303 | <0.001 | *** |
|  |  | T1 to T5 | -2.013 ( -2.958 to -1.068 ) | 0.319 | <0.001 | *** |
|  |  | T1 to T6 | -2.552 ( -3.456 to -1.648 ) | 0.305 | <0.001 | *** |
|  |  | T1 to T7 | -2.906 ( -3.800 to -2.012 ) | 0.302 | <0.001 | *** |
|  |  | T2 to T3 | -0.386 ( -1.153 to 0.382 ) | 0.259 | >0.99 |  |
|  |  | T2 to T5 | -0.803 ( -1.614 to 0.008 ) | 0.274 | 0.055 |  |
|  |  | T2 to T6 | -1.342 ( -2.131 to -0.552 ) | 0.267 | <0.001 | *** |
|  |  | T2 to T7 | -1.696 ( -2.568 to -0.823 ) | 0.295 | <0.001 | *** |
|  |  | T3 to T5 | -0.417 ( -1.258 to 0.424 ) | 0.284 | >0.99 |  |
|  |  | T3 to T6 | -0.956 ( -1.794 to -0.118 ) | 0.283 | 0.013 | * |
|  |  | T3 to T7 | -1.310 ( -2.171 to -0.449 ) | 0.291 | <0.001 | *** |
|  |  | T5 to T6 | -0.539 ( -1.349 to 0.271 ) | 0.274 | 0.748 |  |
|  |  | T5 to T7 | -0.893 ( -1.733 to -0.053 ) | 0.284 | 0.027 | * |
|  |  | T6 to T7 | -0.354 ( -1.117 to 0.409 ) | 0.258 | >0.99 |  |
|  | Time*Sports Club (Yes) | T1 to T2 | -1.843 ( -2.714 to -0.972 ) | 0.294 | <0.001 | *** |
|  |  | T1 to T3 | -2.157 ( -3.088 to -1.227 ) | 0.314 | <0.001 | *** |
|  |  | T1 to T5 | -2.247 ( -3.227 to -1.267 ) | 0.331 | <0.001 | *** |
|  |  | T1 to T6 | -2.898 ( -3.836 to -1.961 ) | 0.317 | <0.001 | *** |
|  |  | T1 to T7 | -3.256 ( -4.182 to -2.329 ) | 0.313 | <0.001 | *** |
|  |  | T2 to T3 | -0.315 ( -1.110 to 0.481 ) | 0.269 | >0.99 |  |
|  |  | T2 to T5 | -0.404 ( -1.245 to 0.436 ) | 0.284 | >0.99 |  |
|  |  | T2 to T6 | -1.056 ( -1.875 to -0.236 ) | 0.277 | 0.003 | ** |
|  |  | T2 to T7 | -1.413 ( -2.318 to -0.508 ) | 0.306 | <0.001 | *** |
|  |  | T3 to T5 | -0.090 ( -0.962 to 0.782 ) | 0.295 | >0.99 |  |
|  |  | T3 to T6 | -0.741 ( -1.610 to 0.128 ) | 0.293 | 0.182 |  |
|  |  | T3 to T7 | -1.098 ( -1.991 to -0.205 ) | 0.301 | 0.005 | ** |
|  |  | T5 to T6 | -0.651 ( -1.491 to 0.189 ) | 0.284 | 0.337 |  |
|  |  | T5 to T7 | -1.008 ( -1.879 to -0.137 ) | 0.294 | 0.011 | * |
|  |  | T6 to T7 | -0.357 ( -1.148 to 0.434 ) | 0.267 | >0.99 |  |
|  | Time*Sports Club (No) | T1 to T2 | -1.525 ( -2.310 to -0.740 ) | 0.265 | <0.001 | *** |
|  |  | T1 to T3 | -1.362 ( -2.200 to -0.524 ) | 0.283 | <0.001 | *** |
|  |  | T1 to T5 | -1.735 ( -2.618 to -0.852 ) | 0.298 | <0.001 | *** |
|  |  | T1 to T6 | -2.478 ( -3.323 to -1.634 ) | 0.285 | <0.001 | *** |
|  |  | T1 to T7 | -2.959 ( -3.794 to -2.125 ) | 0.282 | <0.001 | *** |
|  |  | T2 to T3 | 0.163 ( -0.554 to 0.879 ) | 0.242 | >0.99 |  |
|  |  | T2 to T5 | -0.210 ( -0.968 to 0.547 ) | 0.256 | >0.99 |  |
|  |  | T2 to T6 | -0.953 ( -1.691 to -0.215 ) | 0.249 | 0.002 | ** |
|  |  | T2 to T7 | -1.435 ( -2.250 to -0.619 ) | 0.275 | <0.001 | *** |
|  |  | T3 to T5 | -0.373 ( -1.159 to 0.413 ) | 0.265 | >0.99 |  |
|  |  | T3 to T6 | -1.116 ( -1.899 to -0.333 ) | 0.264 | <0.001 | *** |
|  |  | T3 to T7 | -1.597 ( -2.401 to -0.793 ) | 0.272 | <0.001 | *** |
|  |  | T5 to T6 | -0.743 ( -1.500 to 0.014 ) | 0.256 | 0.059 |  |
|  |  | T5 to T7 | -1.224 ( -2.009 to -0.440 ) | 0.265 | <0.001 | *** |
|  |  | T6 to T7 | -0.481 ( -1.194 to 0.231 ) | 0.241 | 0.697 |  |

<sup>a</sup> Adjusted for multiple comparisons using Bonferroni correction. $p$ -lvl ( $p$ -value level) = \* $p < 0.05$ , \*\* $p < 0.01$ , \*\*\* $p < 0.001$ ; CI = confidence interval; SLS-L = single leg stand left; Mean Difference = mean difference based on the estimated marginal means; $p$ -lvl = significance level; SE = standard error; T1 = baseline measurements in September 2019; T2 = follow-up measurements in June 2020; T3 = follow-up measurements in September 2020; T5 = follow-up measurements in June 2021; T6 = follow-up measurements in September 2021; T7 = follow-up measurements in June 2022.

No interaction effects over time were observed (SLS-L: time*school group: ⴄ_p_^2^= 0.006, *p* = 0.16; time*sports club*school group: ⴄ_p_^2^ = 0.006, *p* = 0.12; SLS-R: time*school group: ⴄ_p_^2^ = 0.003, *p* = 0.58; time*sports club*school group: ⴄ_p_^2^= 0.005, *p* = 0.29) and no significant changes were found in the intervention and control groups during the observation period from September 2021 to June 2022 (Tables R10, R17, and R18 and Figures R18 and R19).

**Table R18.**
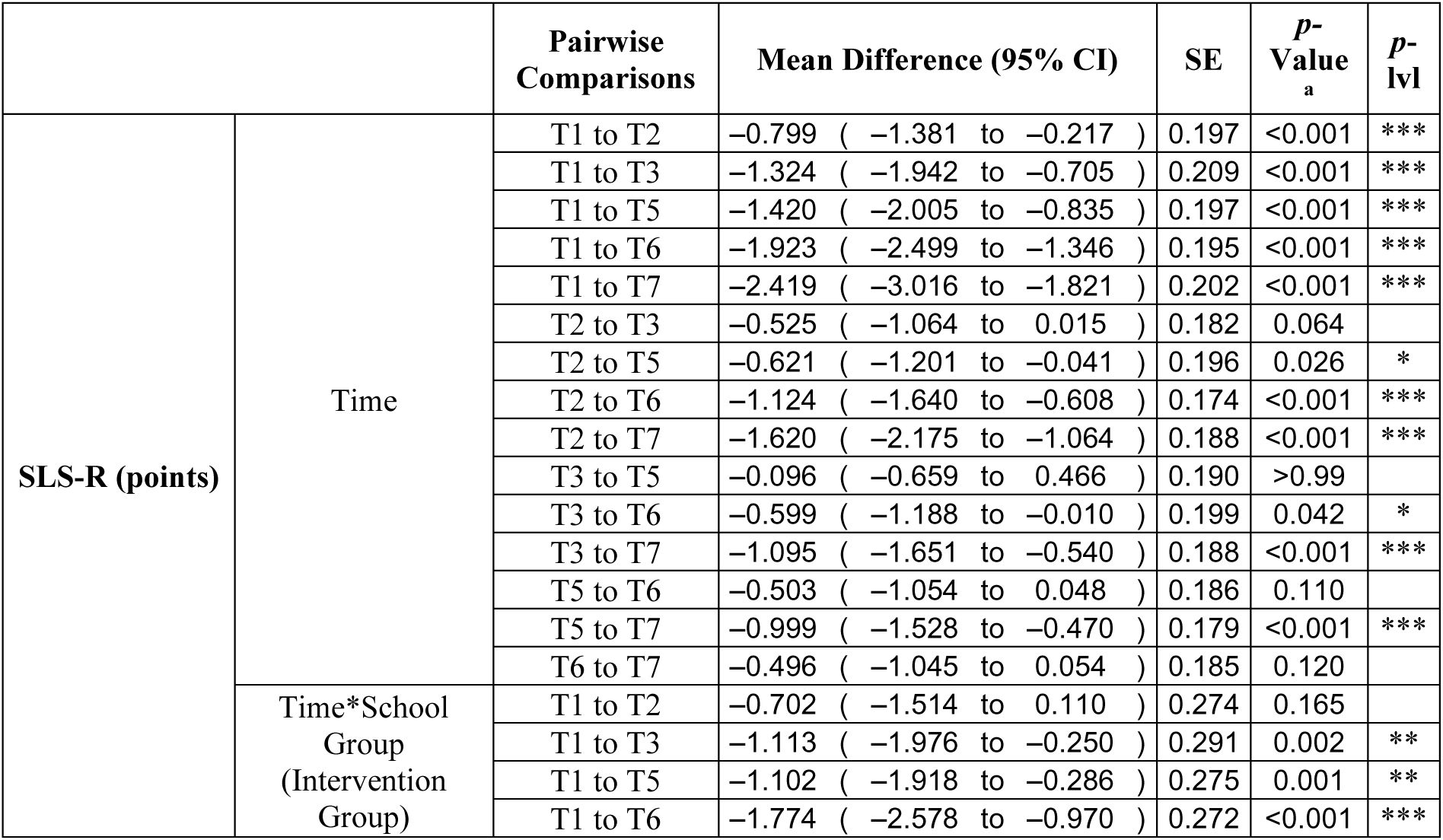
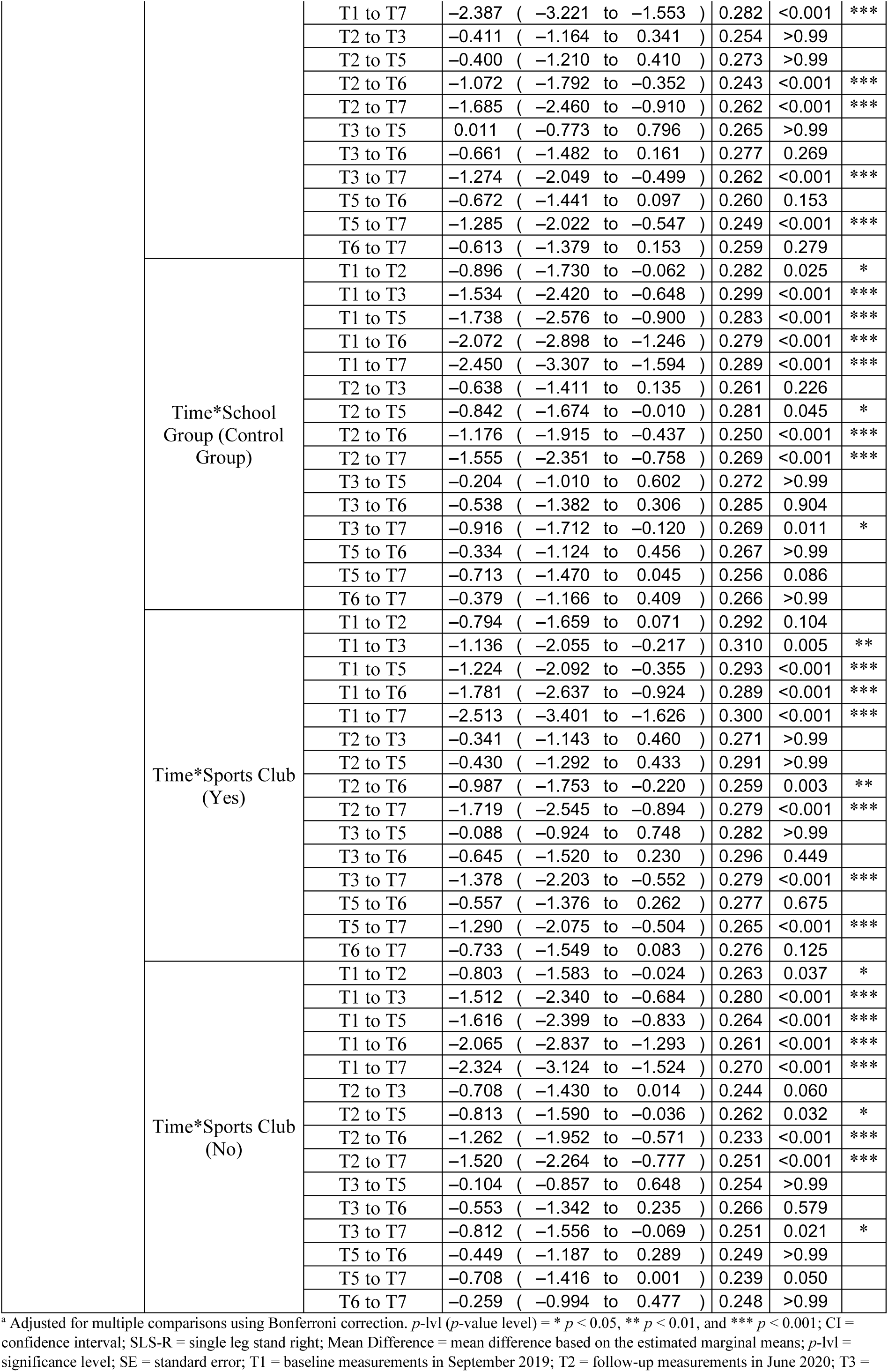
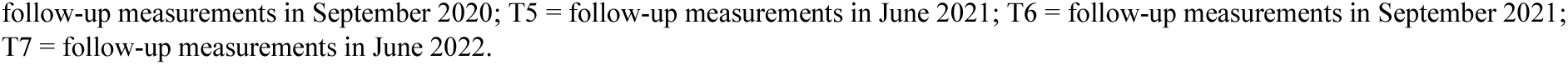
Post hoc tests of SLS-R for the main effect of time and interactions for time*school group and time*sports club membership based on estimated marginal means.

**Figure R19.**
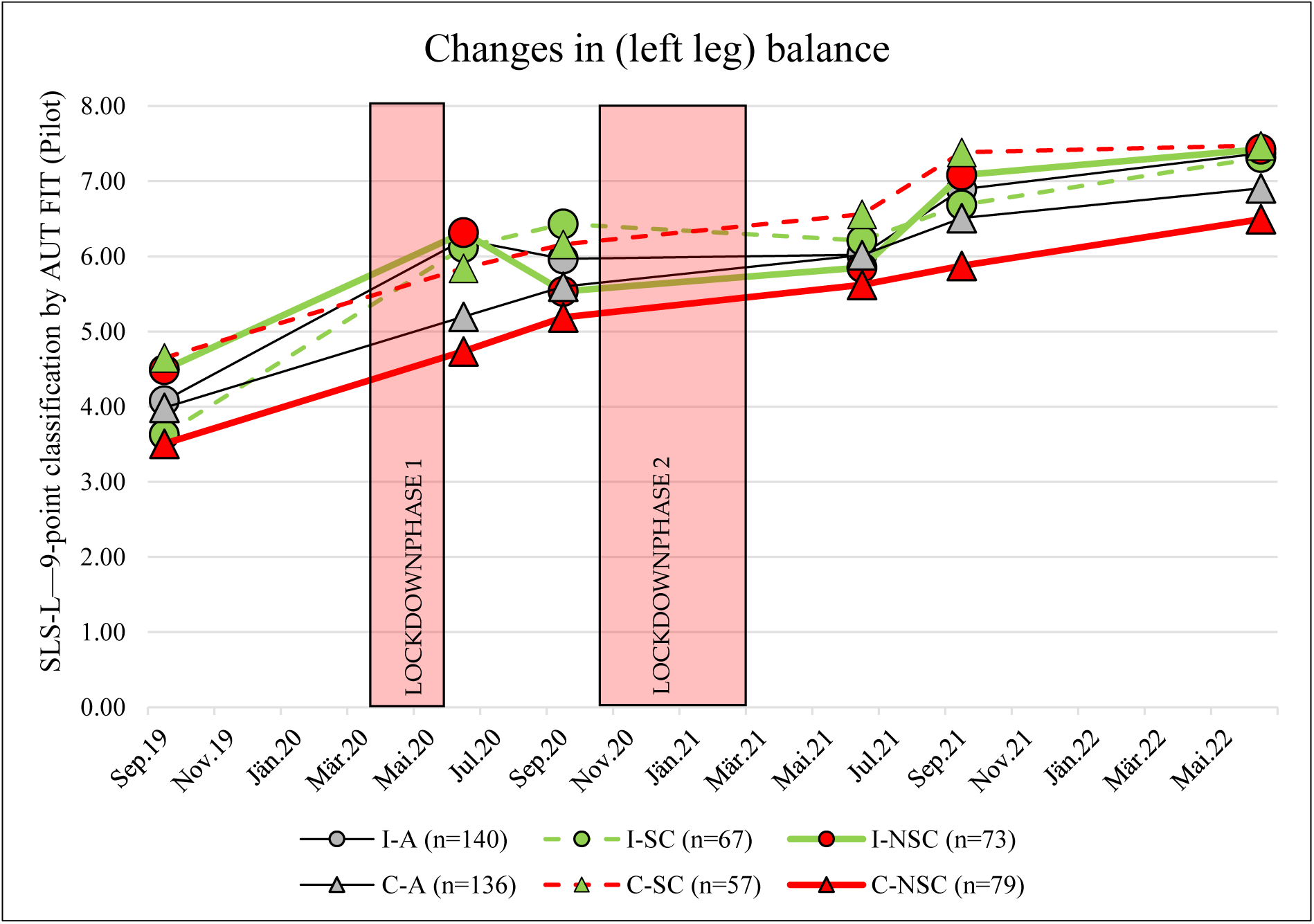
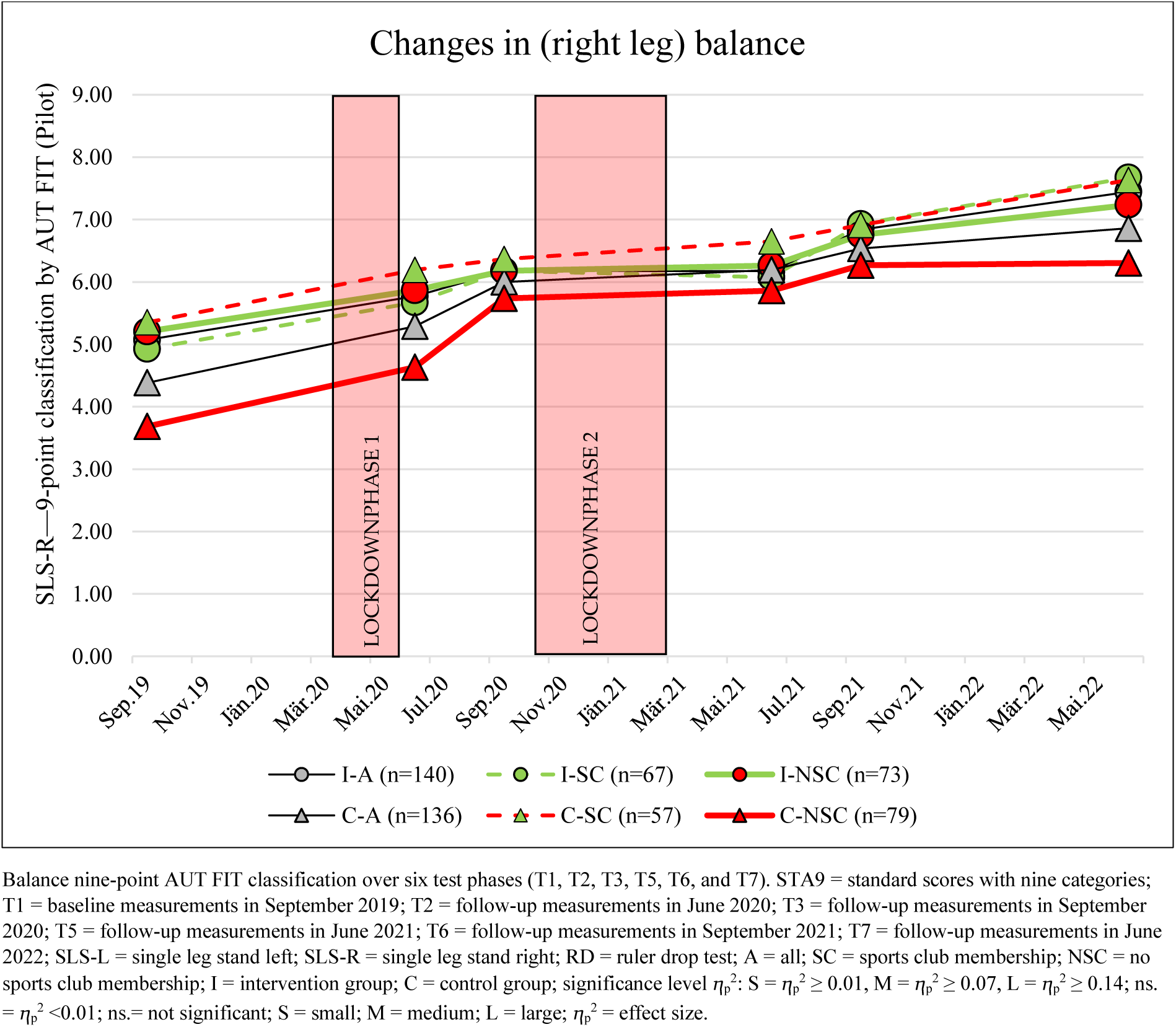
Changes in balance from September 2019 to June 2022 in the subgroups: Intervention and control groups divided by sports club membership.

#### Reaction Speed (RD)

Reaction speed (RD) showed significant changes, with a large effect over the observation period (T1–T7: main effect of time: ⴄ_p_^2^= 0.143; *p* < 0.001) (Table R10 and Figures R5 and R20).

**Figure R20.**
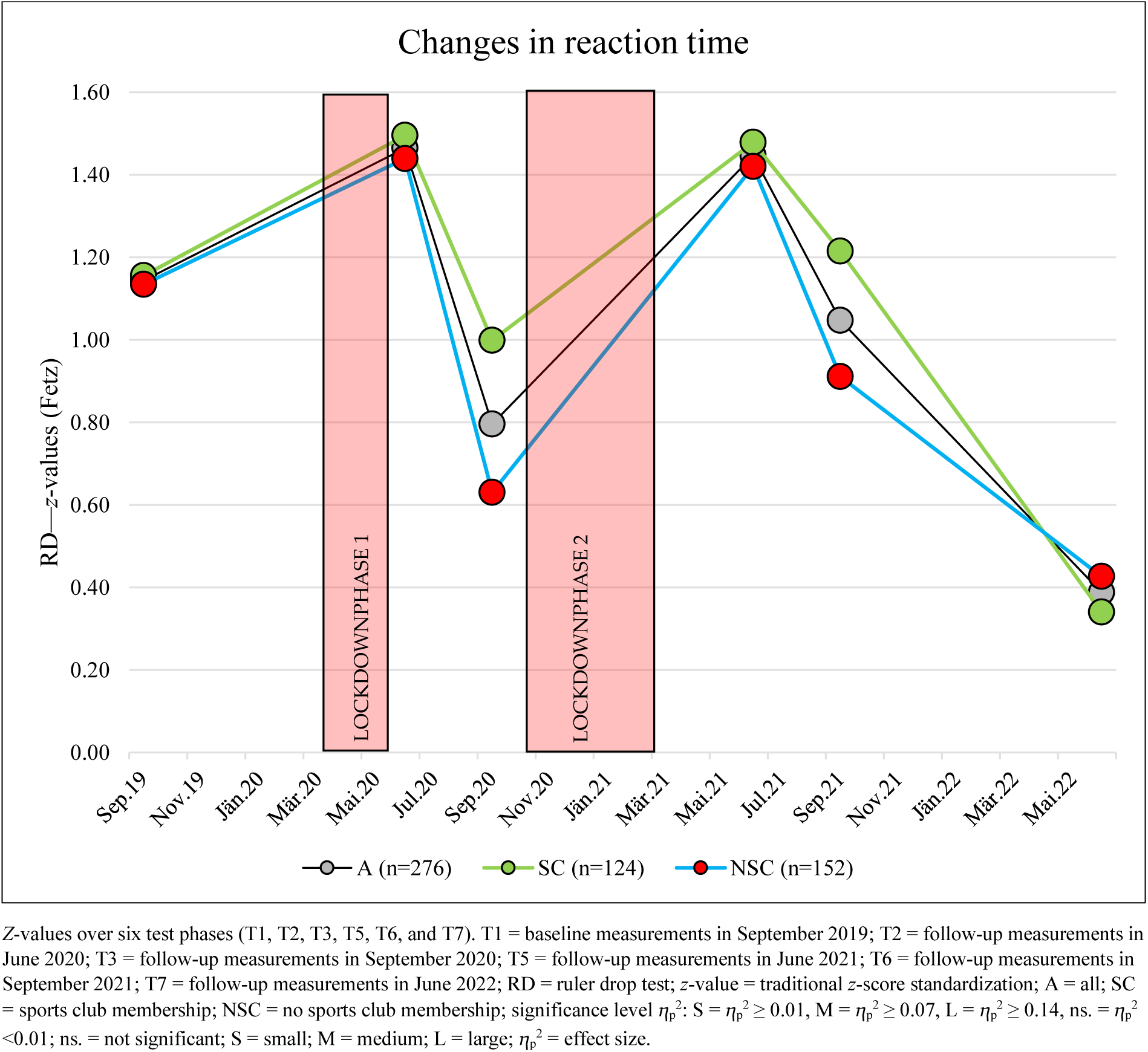
Changes in reaction speed from September 2019 to June 2022.

Over the whole observation period (September 2019 to June 2022), a decrease in performance on the reaction test was demonstrated (–0.75 (95% CI, –1.10 to –0.43), *p* < 0.001) (Tables R10 and R19 and Figures R5 and R20).

**Table R19.** Post hoc tests of RD for the main effect of time and interactions for time*school group and time*sports club membership based on estimated marginal means.

|  |  | Pairwise Comparisons | Mean Difference (95% CI) | SE | *p*-Value<sub>a</sub> | *p*-lvl |
| --- | --- | --- | --- | --- | --- | --- |
| RD *z*-value (Fetz) | Time | T1 to T2 | −0.319 ( −0.562 to −0.076 ) | 0.082 | 0.002 | ** |
|  |  | T1 to T3 | 0.336 ( 0.077 to 0.595 ) | 0.087 | 0.002 | ** |
|  |  | T1 to T5 | −0.301 ( −0.533 to −0.069 ) | 0.078 | 0.002 | ** |
|  |  | T1 to T6 | 0.099 ( −0.151 to 0.349 ) | 0.084 | >0.99 |  |
|  |  | T1 to T7 | 0.766 ( 0.432 to 1.100 ) | 0.113 | <0.001 | *** |
|  |  | T2 to T3 | 0.655 ( 0.437 to 0.873 ) | 0.074 | <0.001 | *** |
|  |  | T2 to T5 | 0.018 ( −0.167 to 0.204 ) | 0.063 | >0.99 |  |
|  |  | T2 to T6 | 0.418 ( 0.185 to 0.651 ) | 0.079 | <0.001 | *** |
|  |  | T2 to T7 | 1.085 ( 0.772 to 1.398 ) | 0.106 | <0.001 | *** |
|  |  | T3 to T5 | −0.637 ( −0.839 to −0.434 ) | 0.069 | <0.001 | *** |
|  |  | T3 to T6 | −0.237 ( −0.473 to −0.001 ) | 0.080 | 0.048 | * |
|  |  | T3 to T7 | 0.430 ( 0.113 to 0.747 ) | 0.107 | 0.001 | ** |
|  |  | T5 to T6 | 0.400 ( 0.203 to 0.596 ) | 0.066 | <0.001 | *** |
|  |  | T5 to T7 | 1.067 ( 0.793 to 1.341 ) | 0.093 | <0.001 | *** |
|  |  | T6 to T7 | 0.667 ( 0.383 to 0.952 ) | 0.096 | <0.001 | *** |
|  | Time*School Group (Intervention Group) | T1 to T2 | −0.553 ( −0.892 to −0.214 ) | 0.114 | <0.001 | *** |
|  |  | T1 to T3 | 0.114 ( −0.247 to 0.475 ) | 0.122 | >0.99 |  |
|  |  | T1 to T5 | −0.466 ( −0.789 to −0.142 ) | 0.109 | <0.001 | *** |
|  |  | T1 to T6 | −0.355 ( −0.704 to −0.007 ) | 0.118 | 0.042 | * |
|  |  | T1 to T7 | 0.515 ( 0.049 to 0.980 ) | 0.157 | 0.018 | * |
|  |  | T2 to T3 | 0.667 ( 0.364 to 0.971 ) | 0.103 | <0.001 | *** |
|  |  | T2 to T5 | 0.087 ( −0.171 to 0.346 ) | 0.087 | >0.99 |  |
|  |  | T2 to T6 | 0.197 ( −0.128 to 0.522 ) | 0.110 | >0.99 |  |
|  |  | T2 to T7 | 1.067 ( 0.631 to 1.504 ) | 0.148 | <0.001 | *** |
|  |  | T3 to T5 | −0.580 ( −0.863 to −0.297 ) | 0.096 | <0.001 | *** |
|  |  | T3 to T6 | −0.470 ( −0.799 to −0.140 ) | 0.111 | <0.001 | *** |
|  |  | T3 to T7 | 0.400 ( −0.042 to 0.843 ) | 0.149 | 0.117 |  |
|  |  | T5 to T6 | 0.110 ( −0.165 to 0.385 ) | 0.093 | >0.99 |  |
|  |  | T5 to T7 | 0.980 ( 0.598 to 1.363 ) | 0.129 | <0.001 | *** |
|  |  | T6 to T7 | 0.870 ( 0.473 to 1.267 ) | 0.134 | <0.001 | *** |
|  | Time*School Group (Control Group) | T1 to T2 | −0.085 ( −0.433 to 0.263 ) | 0.118 | >0.99 |  |
|  |  | T1 to T3 | 0.557 ( 0.187 to 0.928 ) | 0.125 | <0.001 | *** |
|  |  | T1 to T5 | −0.136 ( −0.468 to 0.197 ) | 0.112 | >0.99 |  |
|  |  | T1 to T6 | 0.553 ( 0.195 to 0.912 ) | 0.121 | <0.001 | *** |
|  |  | T1 to T7 | 1.018 ( 0.539 to 1.496 ) | 0.161 | <0.001 | *** |
|  |  | T2 to T3 | 0.643 ( 0.331 to 0.955 ) | 0.105 | <0.001 | *** |
|  |  | T2 to T5 | −0.050 ( −0.316 to 0.215 ) | 0.090 | >0.99 |  |
|  |  | T2 to T6 | 0.639 ( 0.305 to 0.973 ) | 0.113 | <0.001 | *** |
|  |  | T2 to T7 | 1.103 ( 0.654 to 1.552 ) | 0.152 | <0.001 | *** |
|  |  | T3 to T5 | −0.693 ( −0.984 to −0.402 ) | 0.098 | <0.001 | *** |
|  |  | T3 to T6 | −0.004 ( −0.342 to 0.334 ) | 0.114 | >0.99 |  |
|  |  | T3 to T7 | 0.460 ( 0.006 to 0.915 ) | 0.153 | 0.044 | * |
|  |  | T5 to T6 | 0.689 ( 0.407 to 0.971 ) | 0.095 | <0.001 | *** |
|  |  | T5 to T7 | 1.153 ( 0.761 to 1.546 ) | 0.133 | <0.001 | *** |
|  |  | T6 to T7 | 0.464 ( 0.056 to 0.872 ) | 0.138 | 0.013 | * |
|  | Time*Sports Club (Yes) | T1 to T2 | −0.323 ( −0.684 to 0.038 ) | 0.122 | 0.129 |  |
|  |  | T1 to T3 | 0.176 ( −0.209 to 0.560 ) | 0.130 | >0.99 |  |
|  |  | T1 to T5 | −0.309 ( −0.654 to 0.035 ) | 0.116 | 0.125 |  |
|  |  | T1 to T6 | −0.013 ( −0.384 to 0.359 ) | 0.125 | >0.99 |  |
|  |  | T1 to T7 | 0.835 ( 0.339 to 1.331 ) | 0.167 | <0.001 | *** |
|  |  | T2 to T3 | 0.499 ( 0.175 to 0.822 ) | 0.109 | <0.001 | *** |
|  |  | T2 to T5 | 0.013 ( −0.262 to 0.289 ) | 0.093 | >0.99 |  |
|  |  | T2 to T6 | 0.310 ( −0.036 to 0.657 ) | 0.117 | 0.126 |  |
|  |  | T2 to T7 | 1.158 ( 0.692 to 1.623 ) | 0.157 | <0.001 | *** |
|  |  | T3 to T5 | −0.485 ( −0.787 to −0.184 ) | 0.102 | <0.001 | *** |
|  |  | T3 to T6 | −0.188 ( −0.539 to 0.162 ) | 0.118 | >0.99 |  |
|  |  | T3 to T7 | 0.659 ( 0.188 to 1.130 ) | 0.159 | <0.001 | *** |
|  |  | T5 to T6 | 0.297 ( 0.004 to 0.589 ) | 0.099 | 0.044 | * |
|  |  | T5 to T7 | 1.144 ( 0.737 to 1.551 ) | 0.138 | <0.001 | *** |
|  |  | T6 to T7 | 0.847 ( 0.424 to 1.270 ) | 0.143 | <0.001 | *** |
|  | Time*Sports Club<br>(No) | T1 to T2 | -0.315 ( -0.641 to 0.010 ) | 0.110 | 0.066 |  |
|  |  | T1 to T3 | 0.496 ( 0.149 to 0.842 ) | 0.117 | <0.001 | *** |
|  |  | T1 to T5 | -0.292 ( -0.602 to 0.019 ) | 0.105 | 0.086 |  |
|  |  | T1 to T6 | 0.210 ( -0.124 to 0.545 ) | 0.113 | 0.956 |  |
|  |  | T1 to T7 | 0.698 ( 0.251 to 1.144 ) | 0.151 | <0.001 | *** |
|  |  | T2 to T3 | 0.811 ( 0.520 to 1.103 ) | 0.098 | <0.001 | *** |
|  |  | T2 to T5 | 0.023 ( -0.225 to 0.271 ) | 0.084 | >0.99 |  |
|  |  | T2 to T6 | 0.526 ( 0.214 to 0.838 ) | 0.105 | <0.001 | *** |
|  |  | T2 to T7 | 1.013 ( 0.594 to 1.432 ) | 0.142 | <0.001 | *** |
|  |  | T3 to T5 | -0.788 ( -1.059 to -0.516 ) | 0.092 | <0.001 | *** |
|  |  | T3 to T6 | -0.285 ( -0.602 to 0.031 ) | 0.107 | 0.119 |  |
|  |  | T3 to T7 | 0.202 ( -0.223 to 0.626 ) | 0.143 | >0.99 |  |
|  |  | T5 to T6 | 0.502 ( 0.239 to 0.766 ) | 0.089 | <0.001 | *** |
|  |  | T5 to T7 | 0.989 ( 0.623 to 1.356 ) | 0.124 | <0.001 | *** |
|  |  | T6 to T7 | 0.487 ( 0.106 to 0.868 ) | 0.129 | 0.003 | ** |

<sup>a</sup> Adjusted for multiple comparisons using Bonferroni correction. *p*-lvl (*p*-value level) = \* *p* < 0.05, \*\* *p* < 0.01, and \*\*\* *P* < 0.001; CI = confidence interval; RD = ruler drop test; Mean Difference = mean difference based on the estimated marginal means; *p*-lvl = significance level; SE = standard error; T1 = baseline measurements in September 2019; T2 = follow-up measurements in June 2020; T3 = follow-up measurements in September 2020; T5 = follow-up measurements in June 2021; T6 = follow-up measurements in September 2021; T7 = follow-up measurements in June 2022.

Significant increases in performance were found at the test time points of June 2020 and June 2021, respectively, after the school lockdowns in Austria (September 2019 to June 2020 = +0.32 (95% CI, 0.08–0.56), *p* = 0.002; September 2020 to June 2021 = +0.65 (95% CI, 0.43– 0.84), *p* < 0.001) (Table R19 and Figure R20).

In contrast, significant decreases in RD performance were found in all other time periods (June 2020 to September 2020 (–0.66 (95% CI, –0.87 to –0.44), *p* < 0.001), June 2021 to September 2021 (–0.40 (95% CI, –0.60 to –0.20), *p* < 0.001), and September 2021 to June 2022 (–0.66 (95% CI, –0.95 to –0.38), *p* < 0.001) (Table R19 and Figure R20).

No interaction effects were found over time (time*sports club: ⴄ_p_^2^ = 0.007; *p* = 0.11) (Tables R10 and R19 and Figures R5 and R20).

Changes in JS in the school groups: (subgroup: intervention and control groups SY21–22)

Small interaction effects over time (time*school group: ⴄ_p_^2^= 0.021, *p* < 0.001) were found, which were mainly related to the time before the sports intervention (September 2019 to September 2021) (Table R10 and Figure R5).

Significant changes were found during the observation period from September 2021 to June 2022 in the intervention and control groups, with higher significance in the intervention group (I-A = –0.86 (95% CI, –1.27 to –0.47), *p* < 0.001; C-A = –0.46 (95% CI, –0.87 to –0.06), *p* = 0.013) (Table R19 and Figure R21).

**Figure R21.**
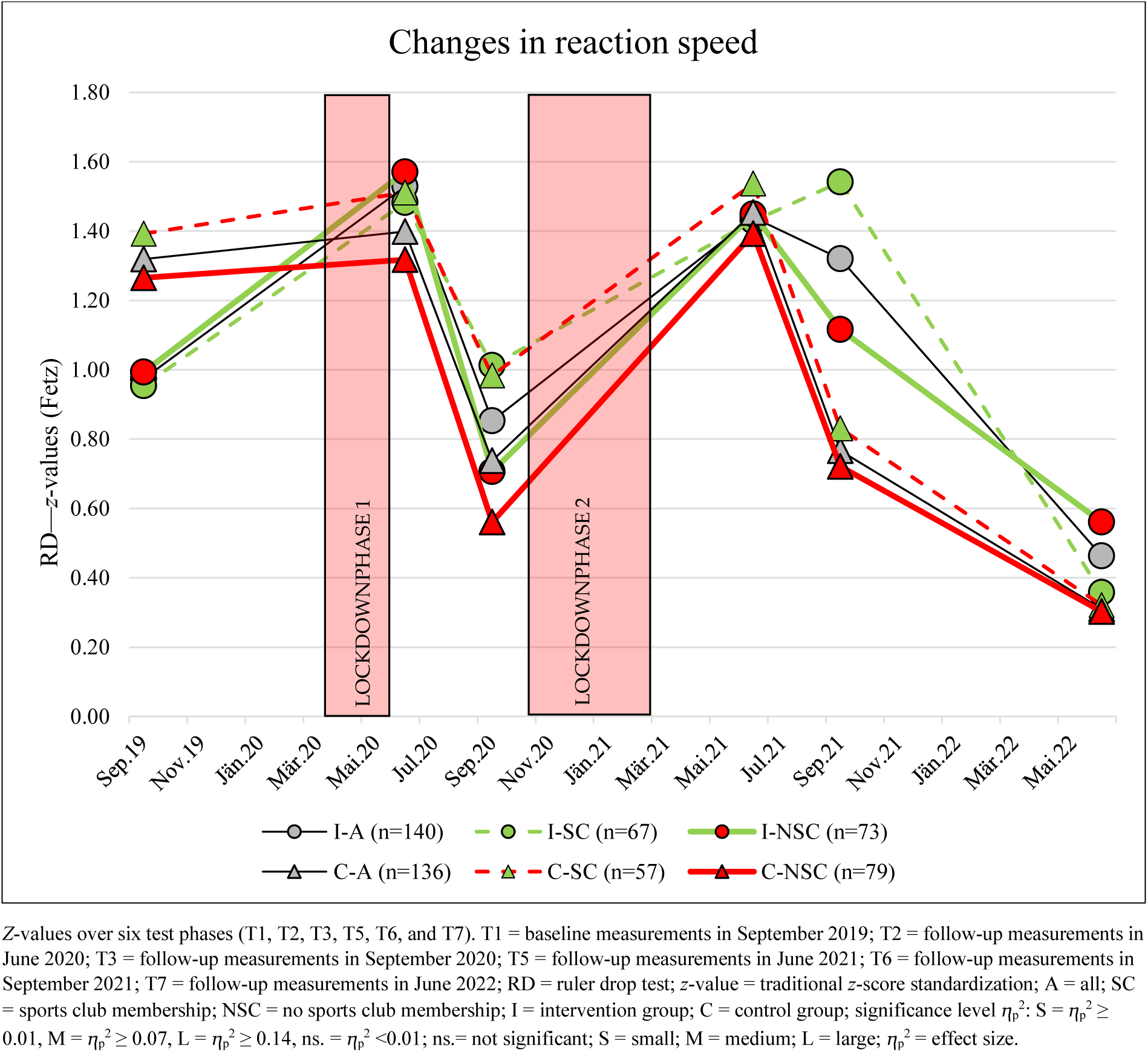
Changes in reaction speed from September 2019 to June 2022 in the subgroups: Intervention and control groups divided by sports club membership.

### Changes in the Classification Groups

Over the overall observation time period (September 2019 to June 2022), highly significant changes (Friedman test: Chi-squared (5–6) = 49.36–387.99, *p* < 0.001) were observed in nine of the 10 classification groups (EQUI BMI_AUT_, WtHR, 6MR, SLJ, 4×10SHR4×10SHR, MB1kg, VSR, RD, and SLS) (Table R20).

**Table R20.** Friedman test for weight classification, health risk estimation, and individual fitness tests using the cutoff values described in AUT FIT for the baseline T1 and follow-up T2, T3, T4, T5, T6, and T7 measurements (overall study population).

| Variable | df | $\chi^2$ | $p$-lvl | $p$-Value |
| --- | --- | --- | --- | --- |
| AUT weight classification, $n = 331$ | 6 | 49.355 | *** | <0.001 |
| WtHR estimation of health risk, $n = 331$ | 6 | 130.369 | *** | <0.001 |
| STA9 Classification, 6MR (DüMo), $n = 331$ | 5 | 318.368 | *** | <0.001 |
| STA9 Classification, SLJ (DüMo), $n = 276$ | 6 | 153.754 | *** | <0.001 |
| STA9 Classification, 4×10SHR SDS (MAC), $n = 276$ | 6 | 387.994 | *** | <0.001 |
| STA9 Classification, MB1kg (KATS-K), $n = 255$ | 6 | 56.057 | *** | <0.001 |
| STA9 Classification, JS (GMT), $n = 276$ | 5 | 19.524 | ** | 0.002 |
| STA9 Classification, VSR (MAKFIT), $n = 276$ | 5 | 70.210 | *** | <0.001 |
| STA9 Classification, RD (Fetz), $n = 276$ | 5 | 179.789 | *** | <0.001 |
| STA9 Classification, SLS (Jarnig), $n = 276$ | 5 | 279.419 | *** | <0.001 |

$n$ = study population; df = degrees of freedom; $\chi^2$ = chi-square test value; $p$ -lvl ( $p$ -value level) = \* $p < 0.05$ , \*\* $p < 0.01$ , and \*\*\* $p < 0.001$ ; $p$ -lvl = significance level; AUT weight classification = based on Austrian reference centile curves passing through adult BMI values; WtHR = waist-to-height ratio; STA9 = standard scores with nine categories; 6MR = six-minute run; SLJ = standing long jump; MB1kg = medicine ball throw (1 kg); 4×10SHR = 4 × 10 m shuttle run; RD = ruler drop test; JS = jumping sideways; VSR = V sit-and-reach test; DüMo = Düsseldorf Modell; KATS-K = Karlsruher test system; MCA = Motor Competence Assessment; GMT = German Motor Test; MAKFIT = Macedonia norm values; T1 = baseline measurements in September 2019; T2 = follow-up measurements in June 2020; T3 = follow-up measurements in September 2020; T4 = follow-up measurements in March 2021; T5 = follow-up measurements in June 2021; T6 = follow-up measurements in September 2021; T7 = follow-up measurements in June 2022.

Only jumping sideways showed “less” significant changes from September 2019 to June 2022 (Friedman test: Chi-square (5) = 19.52, *p* = 0.002) (Table R20).

### Change of Classification in Anthropometrics

#### Changes in Weight Classification

Highly significant changes in the weight classification groups (Friedman test: Chi-square (6) = 49.36, *p* < 0.001) were monitored over the overall observation time period (September 2019 to June 2022) (Table R20 and Figure R22).

**Figure R22.**
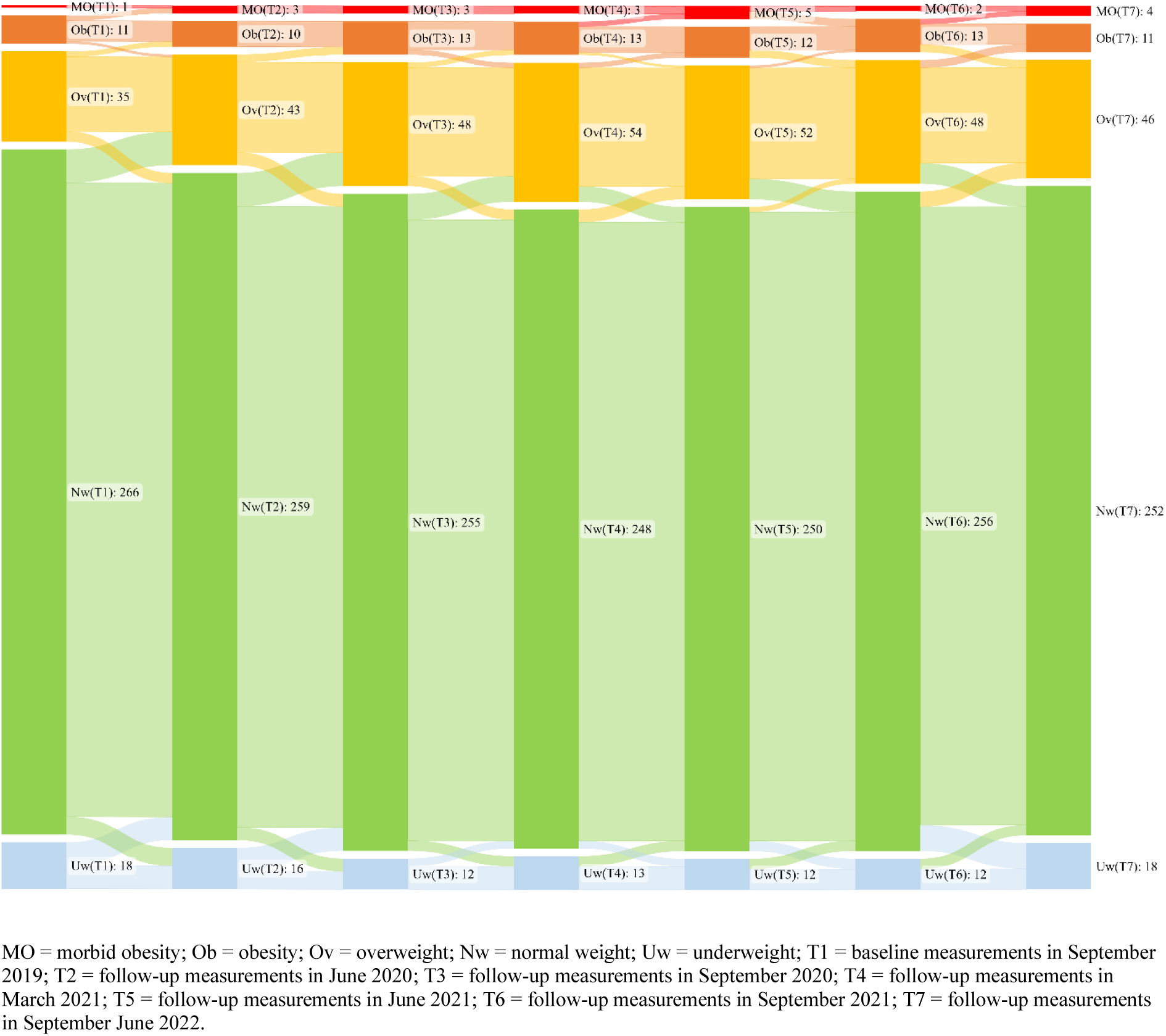
Change in weight classifications between September 2019 and June 2022.

From September 2019 to March 2021, the number of children suffering from overweight, obesity, or extreme obesity increased continuously (September 2019: *n* = 47; March 2021: *n* = 70; *p* < 0.001). Between March 2021 and June 2021, the number became stagnant (March 2021: *n* = 70; June 2021: *n* = 69; *p* > 0.99) and between June 2021 and June 2022 (June 2021: *n* = 69; September 2021: *n* = 61; *p* = 0.17) showed a non-significant trend of health improvement due to decreasing numbers in the overweight, obese, and extreme obesity weight classification groups (Table R21 and Figure R22).

**Table R21.**
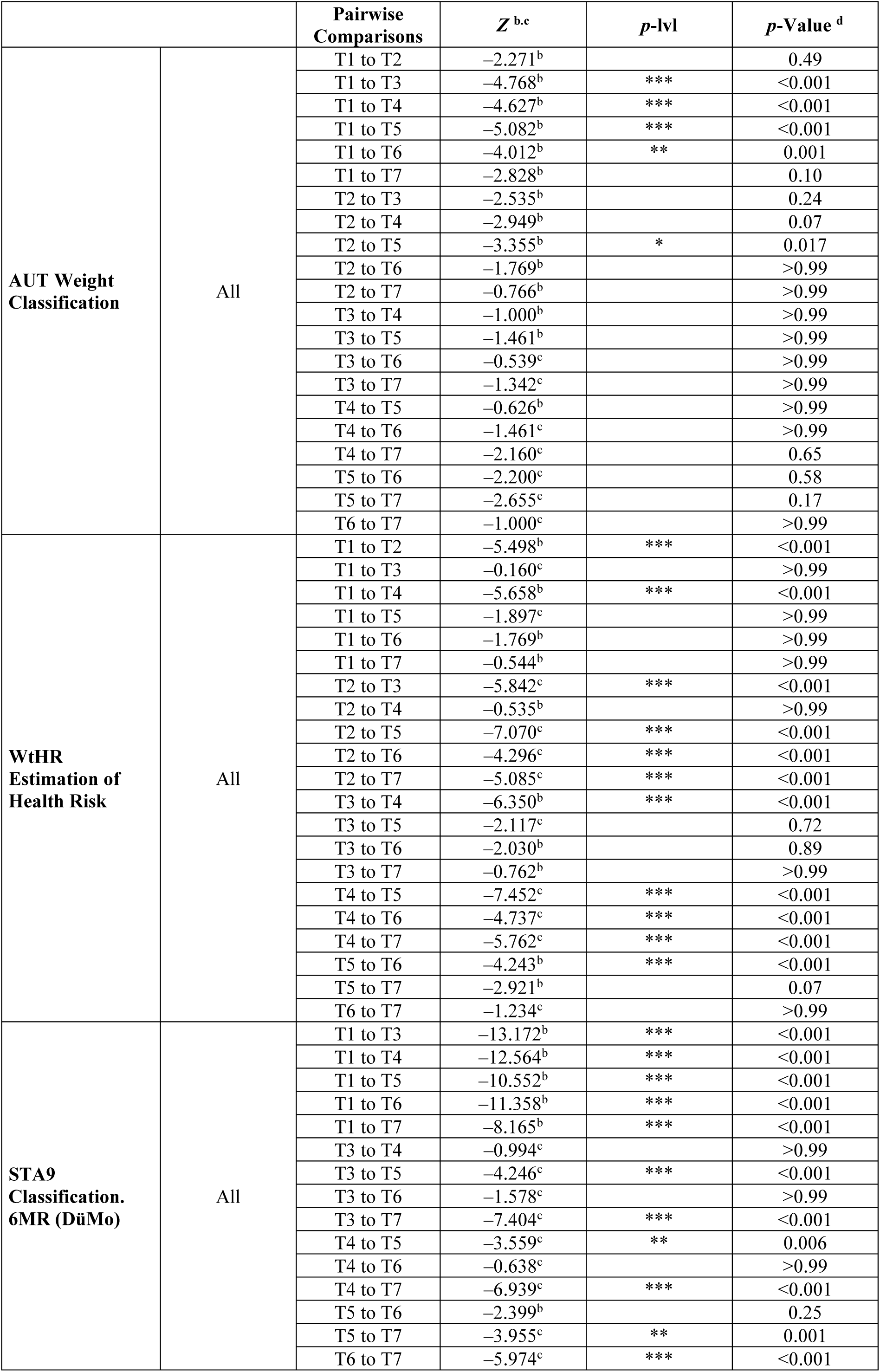
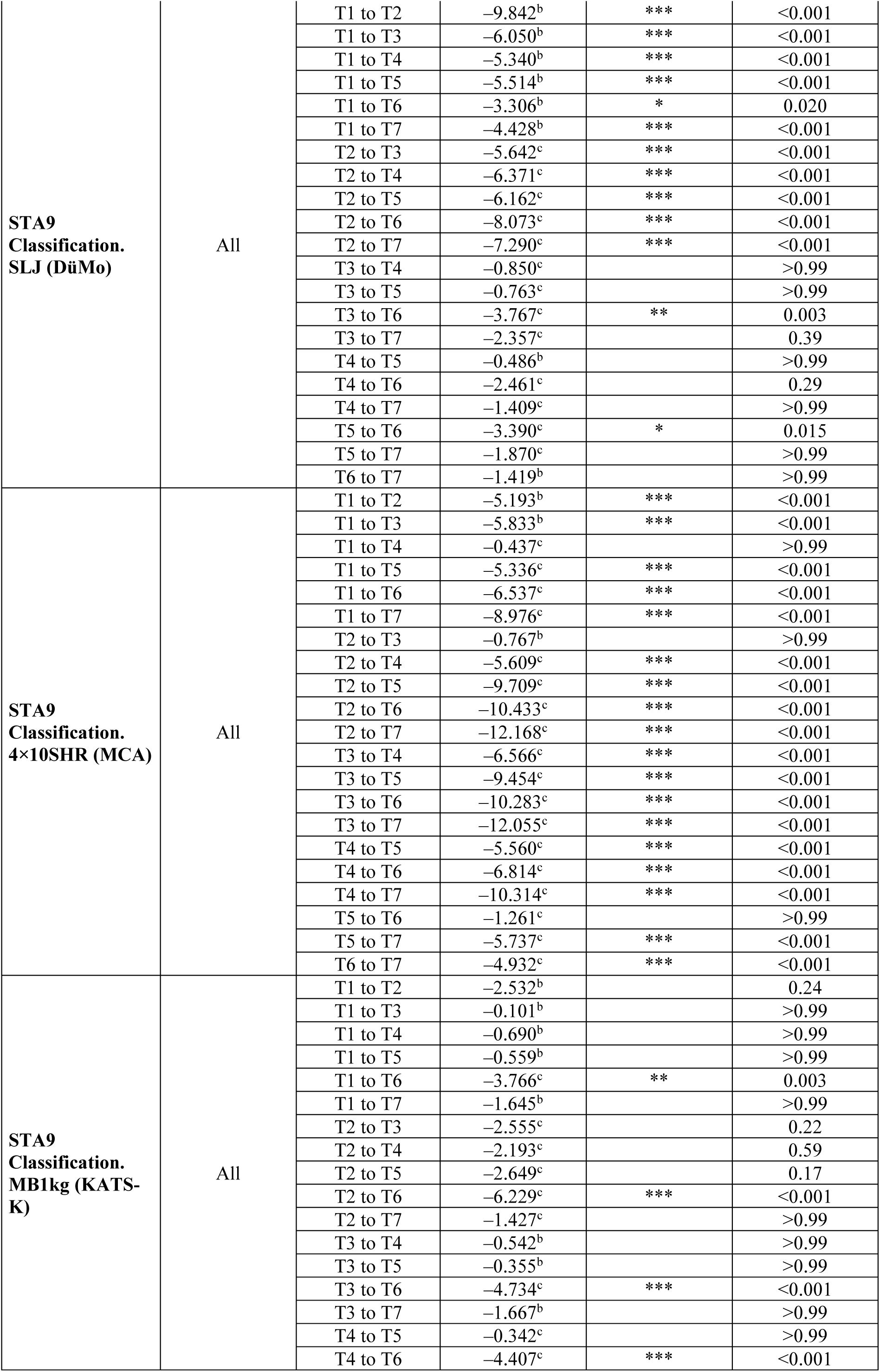
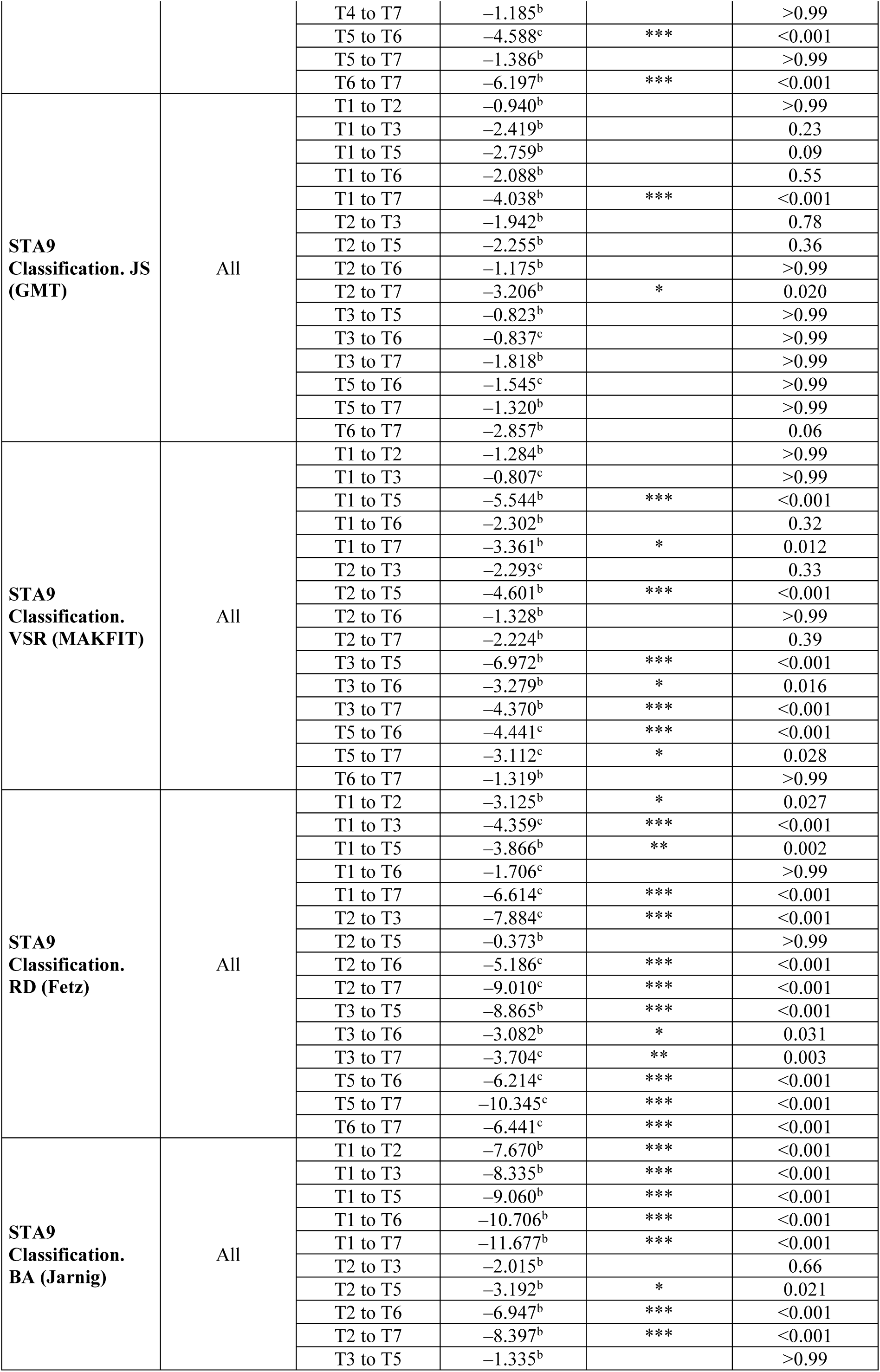
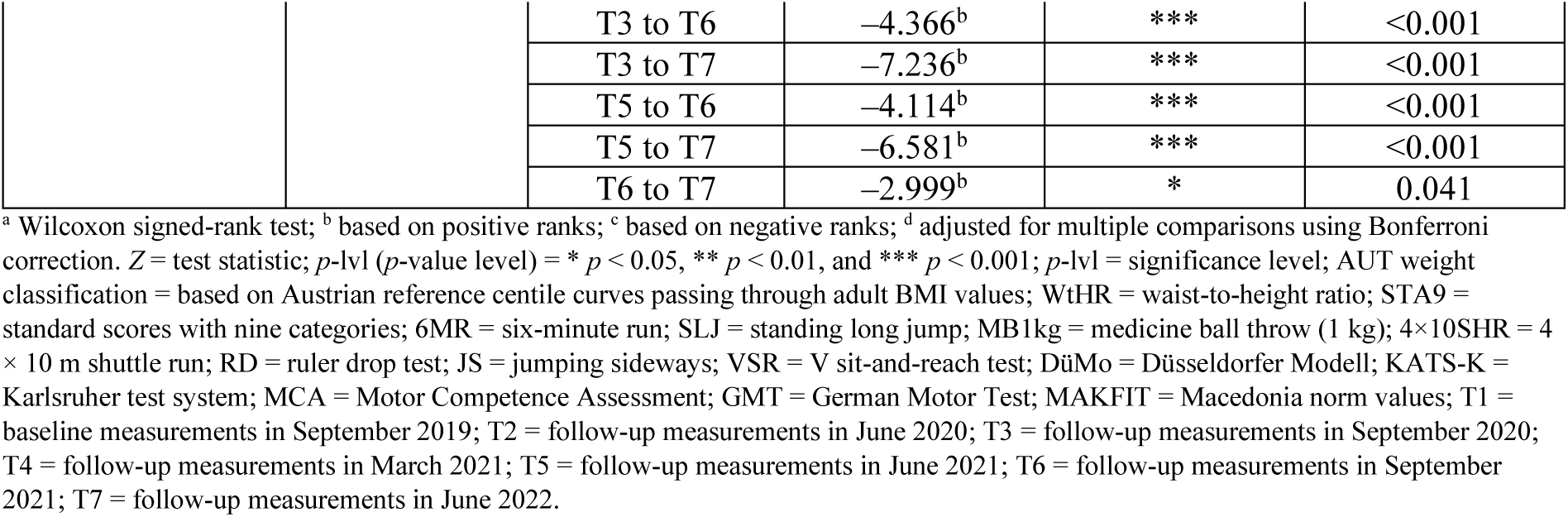
Post hoc analyses using the Wilcoxon test for weight classifications, health risk estimation, and individual fitness tests using the cutoff values described in AUT FIT for the baseline T1 and follow-up T2, T3, T4, T5, T6, and T7 measurements (overall study population).

|  |  | Pairwise Comparisons | Z<sup>b,c</sup> | p-lvl | p-Value<sup>d</sup> |
| --- | --- | --- | --- | --- | --- |
| AUT Weight Classification | All | T1 to T2 | -2.271<sup>b</sup> |  | 0.49 |
|  |  | T1 to T3 | -4.768<sup>b</sup> | *** | <0.001 |
|  |  | T1 to T4 | -4.627<sup>b</sup> | *** | <0.001 |
|  |  | T1 to T5 | -5.082<sup>b</sup> | *** | <0.001 |
|  |  | T1 to T6 | -4.012<sup>b</sup> | ** | 0.001 |
|  |  | T1 to T7 | -2.828<sup>b</sup> |  | 0.10 |
|  |  | T2 to T3 | -2.535<sup>b</sup> |  | 0.24 |
|  |  | T2 to T4 | -2.949<sup>b</sup> |  | 0.07 |
|  |  | T2 to T5 | -3.355<sup>b</sup> | * | 0.017 |
|  |  | T2 to T6 | -1.769<sup>b</sup> |  | >0.99 |
|  |  | T2 to T7 | -0.766<sup>b</sup> |  | >0.99 |
|  |  | T3 to T4 | -1.000<sup>b</sup> |  | >0.99 |
|  |  | T3 to T5 | -1.461<sup>b</sup> |  | >0.99 |
|  |  | T3 to T6 | -0.539<sup>c</sup> |  | >0.99 |
|  |  | T3 to T7 | -1.342<sup>c</sup> |  | >0.99 |
|  |  | T4 to T5 | -0.626<sup>b</sup> |  | >0.99 |
|  |  | T4 to T6 | -1.461<sup>c</sup> |  | >0.99 |
|  |  | T4 to T7 | -2.160<sup>c</sup> |  | 0.65 |
|  |  | T5 to T6 | -2.200<sup>c</sup> |  | 0.58 |
|  |  | T5 to T7 | -2.655<sup>c</sup> |  | 0.17 |
|  |  | T6 to T7 | -1.000<sup>c</sup> |  | >0.99 |
| WtHR Estimation of Health Risk | All | T1 to T2 | -5.498<sup>b</sup> | *** | <0.001 |
|  |  | T1 to T3 | -0.160<sup>c</sup> |  | >0.99 |
|  |  | T1 to T4 | -5.658<sup>b</sup> | *** | <0.001 |
|  |  | T1 to T5 | -1.897<sup>c</sup> |  | >0.99 |
|  |  | T1 to T6 | -1.769<sup>b</sup> |  | >0.99 |
|  |  | T1 to T7 | -0.544<sup>b</sup> |  | >0.99 |
|  |  | T2 to T3 | -5.842<sup>c</sup> | *** | <0.001 |
|  |  | T2 to T4 | -0.535<sup>b</sup> |  | >0.99 |
|  |  | T2 to T5 | -7.070<sup>c</sup> | *** | <0.001 |
|  |  | T2 to T6 | -4.296<sup>c</sup> | *** | <0.001 |
|  |  | T2 to T7 | -5.085<sup>c</sup> | *** | <0.001 |
|  |  | T3 to T4 | -6.350<sup>b</sup> | *** | <0.001 |
|  |  | T3 to T5 | -2.117<sup>c</sup> |  | 0.72 |
|  |  | T3 to T6 | -2.030<sup>b</sup> |  | 0.89 |
|  |  | T3 to T7 | -0.762<sup>b</sup> |  | >0.99 |
|  |  | T4 to T5 | -7.452<sup>c</sup> | *** | <0.001 |
|  |  | T4 to T6 | -4.737<sup>c</sup> | *** | <0.001 |
|  |  | T4 to T7 | -5.762<sup>c</sup> | *** | <0.001 |
|  |  | T5 to T6 | -4.243<sup>b</sup> | *** | <0.001 |
|  |  | T5 to T7 | -2.921<sup>b</sup> |  | 0.07 |
|  |  | T6 to T7 | -1.234<sup>c</sup> |  | >0.99 |
| STA9 Classification. 6MR (DüMo) | All | T1 to T3 | -13.172<sup>b</sup> | *** | <0.001 |
|  |  | T1 to T4 | -12.564<sup>b</sup> | *** | <0.001 |
|  |  | T1 to T5 | -10.552<sup>b</sup> | *** | <0.001 |
|  |  | T1 to T6 | -11.358<sup>b</sup> | *** | <0.001 |
|  |  | T1 to T7 | -8.165<sup>b</sup> | *** | <0.001 |
|  |  | T3 to T4 | -0.994<sup>c</sup> |  | >0.99 |
|  |  | T3 to T5 | -4.246<sup>c</sup> | *** | <0.001 |
|  |  | T3 to T6 | -1.578<sup>c</sup> |  | >0.99 |
|  |  | T3 to T7 | -7.404<sup>c</sup> | *** | <0.001 |
|  |  | T4 to T5 | -3.559<sup>c</sup> | ** | 0.006 |
|  |  | T4 to T6 | -0.638<sup>c</sup> |  | >0.99 |
|  |  | T4 to T7 | -6.939<sup>c</sup> | *** | <0.001 |
|  |  | T5 to T6 | -2.399<sup>b</sup> |  | 0.25 |
|  |  | T5 to T7 | -3.955<sup>c</sup> | ** | 0.001 |
|  |  | T6 to T7 | -5.974<sup>c</sup> | *** | <0.001 |
| **STA9**<br>**Classification.**<br>**SLJ (DüMo)** | All | T1 to T2 | −9.842<sup>b</sup> | *** | <0.001 |
|  |  | T1 to T3 | −6.050<sup>b</sup> | *** | <0.001 |
|  |  | T1 to T4 | −5.340<sup>b</sup> | *** | <0.001 |
|  |  | T1 to T5 | −5.514<sup>b</sup> | *** | <0.001 |
|  |  | T1 to T6 | −3.306<sup>b</sup> | * | 0.020 |
|  |  | T1 to T7 | −4.428<sup>b</sup> | *** | <0.001 |
|  |  | T2 to T3 | −5.642<sup>c</sup> | *** | <0.001 |
|  |  | T2 to T4 | −6.371<sup>c</sup> | *** | <0.001 |
|  |  | T2 to T5 | −6.162<sup>c</sup> | *** | <0.001 |
|  |  | T2 to T6 | −8.073<sup>c</sup> | *** | <0.001 |
|  |  | T2 to T7 | −7.290<sup>c</sup> | *** | <0.001 |
|  |  | T3 to T4 | −0.850<sup>c</sup> |  | >0.99 |
|  |  | T3 to T5 | −0.763<sup>c</sup> |  | >0.99 |
|  |  | T3 to T6 | −3.767<sup>c</sup> | ** | 0.003 |
|  |  | T3 to T7 | −2.357<sup>c</sup> |  | 0.39 |
|  |  | T4 to T5 | −0.486<sup>b</sup> |  | >0.99 |
|  |  | T4 to T6 | −2.461<sup>c</sup> |  | 0.29 |
|  |  | T4 to T7 | −1.409<sup>c</sup> |  | >0.99 |
|  |  | T5 to T6 | −3.390<sup>c</sup> | * | 0.015 |
|  |  | T5 to T7 | −1.870<sup>c</sup> |  | >0.99 |
|  |  | T6 to T7 | −1.419<sup>b</sup> |  | >0.99 |
| **STA9**<br>**Classification.**<br>**4×10SHR (MCA)** | All | T1 to T2 | −5.193<sup>b</sup> | *** | <0.001 |
|  |  | T1 to T3 | −5.833<sup>b</sup> | *** | <0.001 |
|  |  | T1 to T4 | −0.437<sup>c</sup> |  | >0.99 |
|  |  | T1 to T5 | −5.336<sup>c</sup> | *** | <0.001 |
|  |  | T1 to T6 | −6.537<sup>c</sup> | *** | <0.001 |
|  |  | T1 to T7 | −8.976<sup>c</sup> | *** | <0.001 |
|  |  | T2 to T3 | −0.767<sup>b</sup> |  | >0.99 |
|  |  | T2 to T4 | −5.609<sup>c</sup> | *** | <0.001 |
|  |  | T2 to T5 | −9.709<sup>c</sup> | *** | <0.001 |
|  |  | T2 to T6 | −10.433<sup>c</sup> | *** | <0.001 |
|  |  | T2 to T7 | −12.168<sup>c</sup> | *** | <0.001 |
|  |  | T3 to T4 | −6.566<sup>c</sup> | *** | <0.001 |
|  |  | T3 to T5 | −9.454<sup>c</sup> | *** | <0.001 |
|  |  | T3 to T6 | −10.283<sup>c</sup> | *** | <0.001 |
|  |  | T3 to T7 | −12.055<sup>c</sup> | *** | <0.001 |
|  |  | T4 to T5 | −5.560<sup>c</sup> | *** | <0.001 |
|  |  | T4 to T6 | −6.814<sup>c</sup> | *** | <0.001 |
|  |  | T4 to T7 | −10.314<sup>c</sup> | *** | <0.001 |
|  |  | T5 to T6 | −1.261<sup>c</sup> |  | >0.99 |
|  |  | T5 to T7 | −5.737<sup>c</sup> | *** | <0.001 |
|  |  | T6 to T7 | −4.932<sup>c</sup> | *** | <0.001 |
| **STA9**<br>**Classification.**<br>**MB1kg (KATS-K)** | All | T1 to T2 | −2.532<sup>b</sup> |  | 0.24 |
|  |  | T1 to T3 | −0.101<sup>b</sup> |  | >0.99 |
|  |  | T1 to T4 | −0.690<sup>b</sup> |  | >0.99 |
|  |  | T1 to T5 | −0.559<sup>b</sup> |  | >0.99 |
|  |  | T1 to T6 | −3.766<sup>c</sup> | ** | 0.003 |
|  |  | T1 to T7 | −1.645<sup>b</sup> |  | >0.99 |
|  |  | T2 to T3 | −2.555<sup>c</sup> |  | 0.22 |
|  |  | T2 to T4 | −2.193<sup>c</sup> |  | 0.59 |
|  |  | T2 to T5 | −2.649<sup>c</sup> |  | 0.17 |
|  |  | T2 to T6 | −6.229<sup>c</sup> | *** | <0.001 |
|  |  | T2 to T7 | −1.427<sup>c</sup> |  | >0.99 |
|  |  | T3 to T4 | −0.542<sup>b</sup> |  | >0.99 |
|  |  | T3 to T5 | −0.355<sup>b</sup> |  | >0.99 |
|  |  | T3 to T6 | −4.734<sup>c</sup> | *** | <0.001 |
|  |  | T3 to T7 | −1.667<sup>b</sup> |  | >0.99 |
|  |  | T4 to T5 | −0.342<sup>c</sup> |  | >0.99 |
|  |  | T4 to T6 | −4.407<sup>c</sup> | *** | <0.001 |
|  |  | T4 to T7 | −1.185<sup>b</sup> |  | >0.99 |
|  |  | T5 to T6 | −4.588<sup>c</sup> | *** | <0.001 |
|  |  | T5 to T7 | −1.386<sup>b</sup> |  | >0.99 |
|  |  | T6 to T7 | −6.197<sup>b</sup> | *** | <0.001 |
| **STA9**<br>**Classification. JS**<br>**(GMT)** | All | T1 to T2 | −0.940<sup>b</sup> |  | >0.99 |
|  |  | T1 to T3 | −2.419<sup>b</sup> |  | 0.23 |
|  |  | T1 to T5 | −2.759<sup>b</sup> |  | 0.09 |
|  |  | T1 to T6 | −2.088<sup>b</sup> |  | 0.55 |
|  |  | T1 to T7 | −4.038<sup>b</sup> | *** | <0.001 |
|  |  | T2 to T3 | −1.942<sup>b</sup> |  | 0.78 |
|  |  | T2 to T5 | −2.255<sup>b</sup> |  | 0.36 |
|  |  | T2 to T6 | −1.175<sup>b</sup> |  | >0.99 |
|  |  | T2 to T7 | −3.206<sup>b</sup> | * | 0.020 |
|  |  | T3 to T5 | −0.823<sup>b</sup> |  | >0.99 |
|  |  | T3 to T6 | −0.837<sup>c</sup> |  | >0.99 |
|  |  | T3 to T7 | −1.818<sup>b</sup> |  | >0.99 |
|  |  | T5 to T6 | −1.545<sup>c</sup> |  | >0.99 |
|  |  | T5 to T7 | −1.320<sup>b</sup> |  | >0.99 |
|  |  | T6 to T7 | −2.857<sup>b</sup> |  | 0.06 |
| **STA9**<br>**Classification.**<br>**VSR (MAKFIT)** | All | T1 to T2 | −1.284<sup>b</sup> |  | >0.99 |
|  |  | T1 to T3 | −0.807<sup>c</sup> |  | >0.99 |
|  |  | T1 to T5 | −5.544<sup>b</sup> | *** | <0.001 |
|  |  | T1 to T6 | −2.302<sup>b</sup> |  | 0.32 |
|  |  | T1 to T7 | −3.361<sup>b</sup> | * | 0.012 |
|  |  | T2 to T3 | −2.293<sup>c</sup> |  | 0.33 |
|  |  | T2 to T5 | −4.601<sup>b</sup> | *** | <0.001 |
|  |  | T2 to T6 | −1.328<sup>b</sup> |  | >0.99 |
|  |  | T2 to T7 | −2.224<sup>b</sup> |  | 0.39 |
|  |  | T3 to T5 | −6.972<sup>b</sup> | *** | <0.001 |
|  |  | T3 to T6 | −3.279<sup>b</sup> | * | 0.016 |
|  |  | T3 to T7 | −4.370<sup>b</sup> | *** | <0.001 |
|  |  | T5 to T6 | −4.441<sup>c</sup> | *** | <0.001 |
|  |  | T5 to T7 | −3.112<sup>c</sup> | * | 0.028 |
|  |  | T6 to T7 | −1.319<sup>b</sup> |  | >0.99 |
| **STA9**<br>**Classification.**<br>**RD (Fetz)** | All | T1 to T2 | −3.125<sup>b</sup> | * | 0.027 |
|  |  | T1 to T3 | −4.359<sup>c</sup> | *** | <0.001 |
|  |  | T1 to T5 | −3.866<sup>b</sup> | ** | 0.002 |
|  |  | T1 to T6 | −1.706<sup>c</sup> |  | >0.99 |
|  |  | T1 to T7 | −6.614<sup>c</sup> | *** | <0.001 |
|  |  | T2 to T3 | −7.884<sup>c</sup> | *** | <0.001 |
|  |  | T2 to T5 | −0.373<sup>b</sup> |  | >0.99 |
|  |  | T2 to T6 | −5.186<sup>c</sup> | *** | <0.001 |
|  |  | T2 to T7 | −9.010<sup>c</sup> | *** | <0.001 |
|  |  | T3 to T5 | −8.865<sup>b</sup> | *** | <0.001 |
|  |  | T3 to T6 | −3.082<sup>b</sup> | * | 0.031 |
|  |  | T3 to T7 | −3.704<sup>c</sup> | ** | 0.003 |
|  |  | T5 to T6 | −6.214<sup>c</sup> | *** | <0.001 |
|  |  | T5 to T7 | −10.345<sup>c</sup> | *** | <0.001 |
|  |  | T6 to T7 | −6.441<sup>c</sup> | *** | <0.001 |
| **STA9**<br>**Classification.**<br>**BA (Jarnig)** | All | T1 to T2 | −7.670<sup>b</sup> | *** | <0.001 |
|  |  | T1 to T3 | −8.335<sup>b</sup> | *** | <0.001 |
|  |  | T1 to T5 | −9.060<sup>b</sup> | *** | <0.001 |
|  |  | T1 to T6 | −10.706<sup>b</sup> | *** | <0.001 |
|  |  | T1 to T7 | −11.677<sup>b</sup> | *** | <0.001 |
|  |  | T2 to T3 | −2.015<sup>b</sup> |  | 0.66 |
|  |  | T2 to T5 | −3.192<sup>b</sup> | * | 0.021 |
|  |  | T2 to T6 | −6.947<sup>b</sup> | *** | <0.001 |
|  |  | T2 to T7 | −8.397<sup>b</sup> | *** | <0.001 |
|  |  | T3 to T5 | −1.335<sup>b</sup> |  | >0.99 |

|  |  |  |  |  |
| --- | --- | --- | --- | --- |
|  | T3 to T6 | −4.366<sup>b</sup> | *** | <0.001 |
|  | T3 to T7 | −7.236<sup>b</sup> | *** | <0.001 |
|  | T5 to T6 | −4.114<sup>b</sup> | *** | <0.001 |
|  | T5 to T7 | −6.581<sup>b</sup> | *** | <0.001 |
|  | T6 to T7 | −2.999<sup>b</sup> | * | 0.041 |

<sup>a</sup> Wilcoxon signed-rank test; <sup>b</sup> based on positive ranks; <sup>c</sup> based on negative ranks; <sup>d</sup> adjusted for multiple comparisons using Bonferroni correction. Z = test statistic; *p*-lvl (*p*-value level) = \* *p* < 0.05, \*\* *p* < 0.01, and \*\*\* *p* < 0.001; *p*-lvl = significance level; AUT weight classification = based on Austrian reference centile curves passing through adult BMI values; WtHR = waist-to-height ratio; STA9 = standard scores with nine categories; 6MR = six-minute run; SLJ = standing long jump; MB1kg = medicine ball throw (1 kg); 4×10SHR = 4 × 10 m shuttle run; RD = ruler drop test; JS = jumping sideways; VSR = V sit-and-reach test; DüMo = Düsseldorf Modell; KATS-K = Karlsruher test system; MCA = Motor Competence Assessment; GMT = German Motor Test; MAKFIT = Macedonia norm values; T1 = baseline measurements in September 2019; T2 = follow-up measurements in June 2020; T3 = follow-up measurements in September 2020; T4 = follow-up measurements in March 2021; T5 = follow-up measurements in June 2021; T6 = follow-up measurements in September 2021; T7 = follow-up measurements in June 2022.

Comparing the number of children in the weight classification groups from the baseline level (September 2019) before COVID-19 and the final test time point (June 2022), a nonsignificant increase (based on positive ranks) in the children in the higher weight groups (overweight, obese, and extremely obese) was observed (*z* = –2.83, *p* = 0.10) (Table R21 and Figure R22).

#### Changes in Health Risk Estimation

Highly significant changes in the estimation of health risk (Friedman test: Chi-square (6) = 130.37, *p* < 0.001) were observed over the overall observation time period (September 2019 to June 2022) (Table R20 and Figure R23).

**Figure R23.**
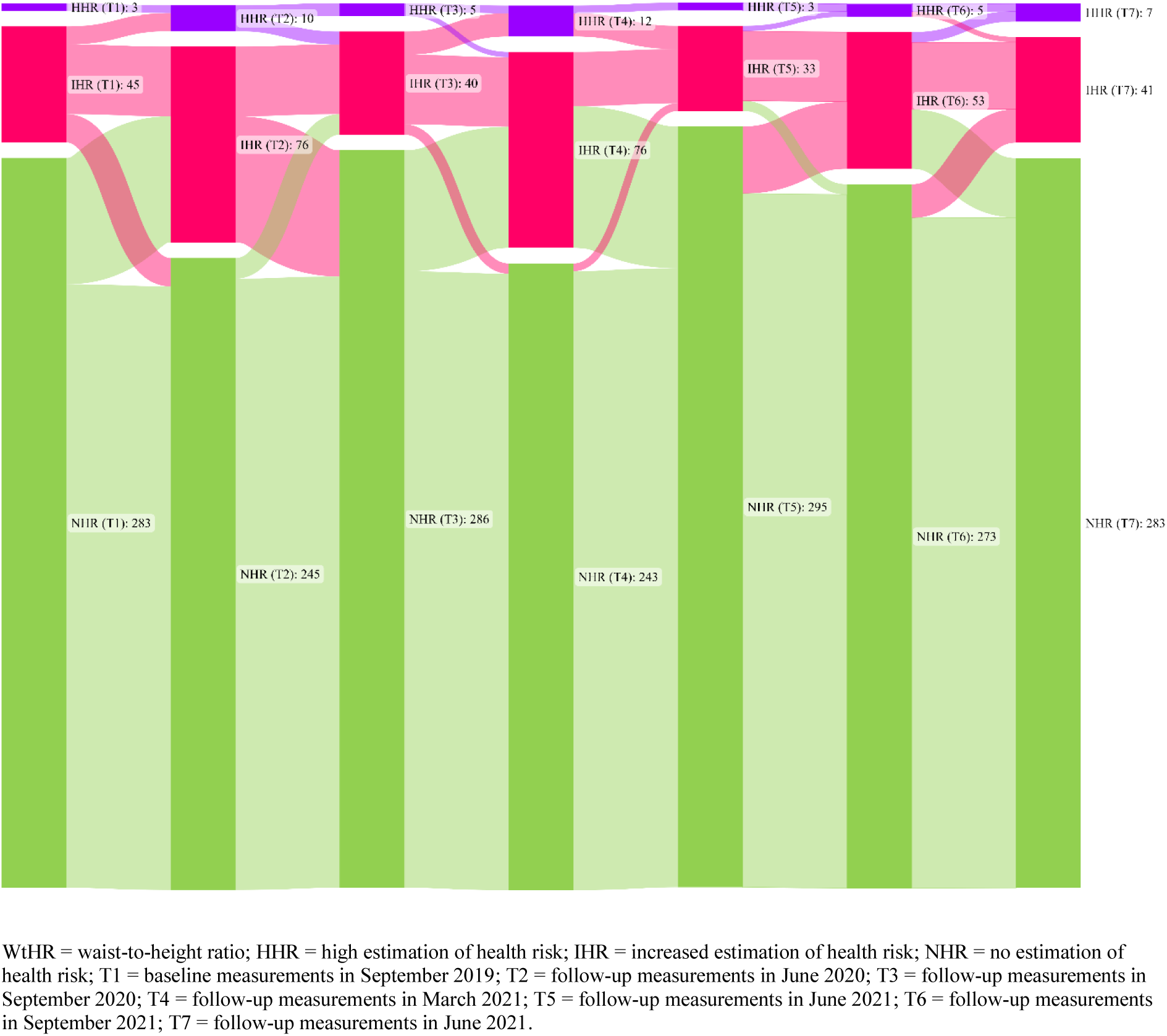
Changes in health risk estimation between September 2019 and June 2022.

The number of children with increased or high health risk increased dramatically after the first (September 2019 (*n* = 48), June 2020 (*n* = 86), *p* < 0.001) and second (September 2020 (*n* = 45), March 2021 (*n* = 88), *p* < 0.001) lockdown periods. In contrast, values during the first open phase (June 2020 (*n* = 86) to September 2020 (*n* = 45), *p* < 0.001) and the second open phase (March 2021 (*n* = 88) to June 2021 (*n* = 36), *p* < 0.001) showed a significant improvement, i.e., a decrease in the children’s mean abdominal circumference (Table R21 and Figure R23).

Comparing the number of children in all three classification groups (no, increased, and high health risk) from baseline (September 2019) before COVID-19 and the last test time point (June 2022), there were no significant changes over the overall observation time period (*z* = – 0.54, *p* > 0.99) (Table R21 and Figure R23).

### Changes in the Classification of Performance in Fitness

#### Six-Minute Run (6MR)

Highly significant changes (Friedman test: Chi-square (5) = 318.37, *p* < 0.001) were observed in cardiorespiratory fitness over the overall observation time period (September 2019 to June 2022) (Table R20 and Figure R24).

**Figure R24.**
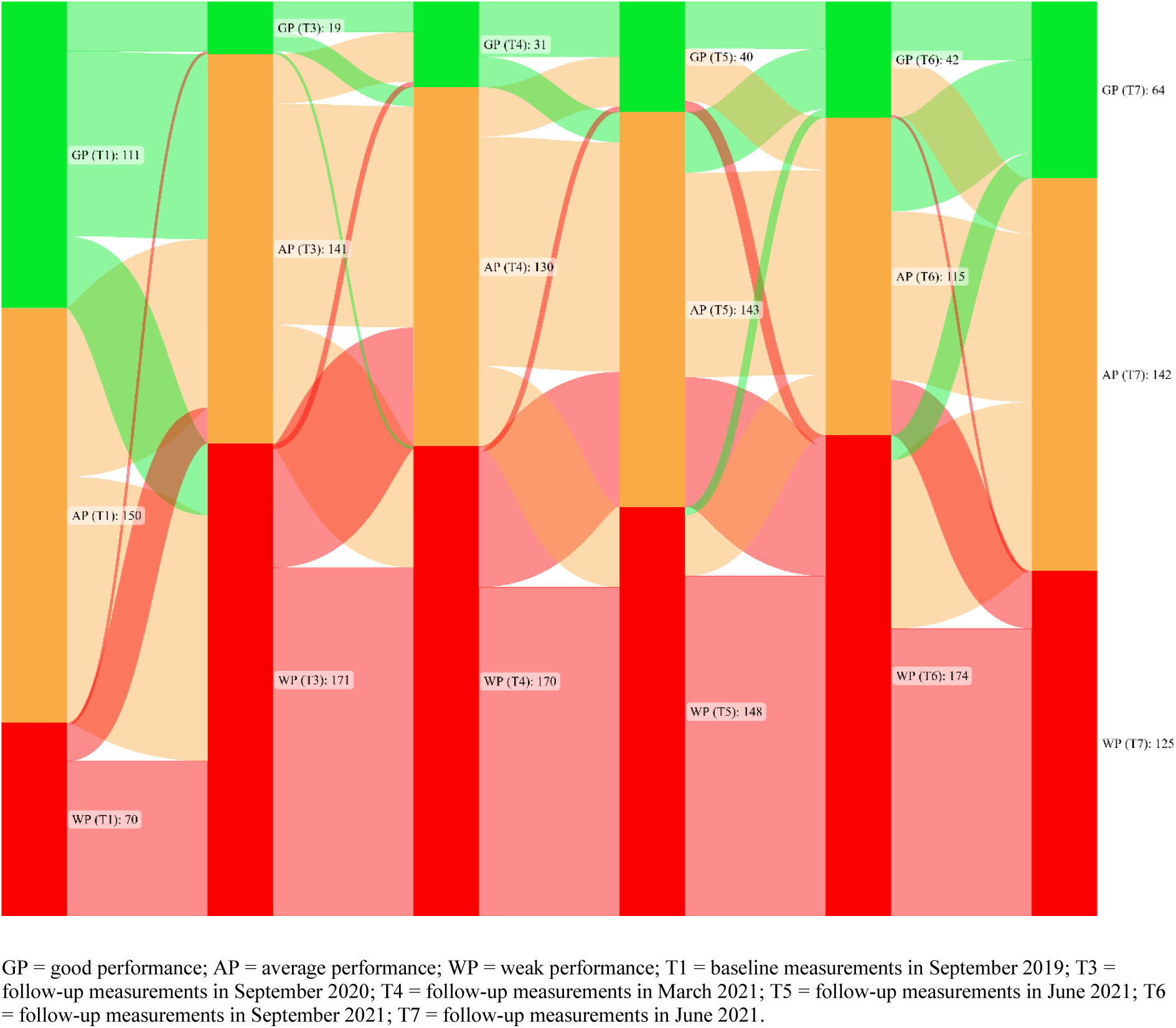
Changes in cardiorespiratory fitness performance groups between September 2019 and June 2022.

Between the baseline measurement in September 2019 and the first measurement time point of cardiorespiratory fitness (September 2020) after the COVID-19 pandemic outbreak, there was a dramatic increase in children in the low performance groups (September 2019 (*n* = 70), September 2020 (*n* = 171), *p* < 0.001). This worrisome decline in cardiorespiratory endurance stagnated at this low level (September 2020 (*n* = 171), September 2021 (*n* = 174), *p* > 0.99), with little variation between testing phases. In the last observation period between September 2021 and June 2022, the number of children in the weak classification group decreased significantly (September 2021 (*n* = 174), June 2022 (*n* = 125), *p* < 0.001), but stayed significantly above the pre-COVID-19 baseline level in September 2019 (*n* = 70, *p* < 0.001) (Table R21 and Figure R24).

Comparing the number of children in all three classification groups (weak, average, and good performance) from baseline (September 2019) before COVID-19 against the last test time point (June 2022), highly significant changes were seen over the overall observation time period (*z* = –8.17, *p* < 0.001) (Table R21 and Figure R24).

#### Standing Long Jump (SLJ)

Highly significant changes (Friedman test: Chi-square (6) = 153.75, *p* < 0.001) were monitored for the standing long jump over the overall observation time period (September 2019 to June 2022) (Table R20 and Figure R25).

**Figure R25.**
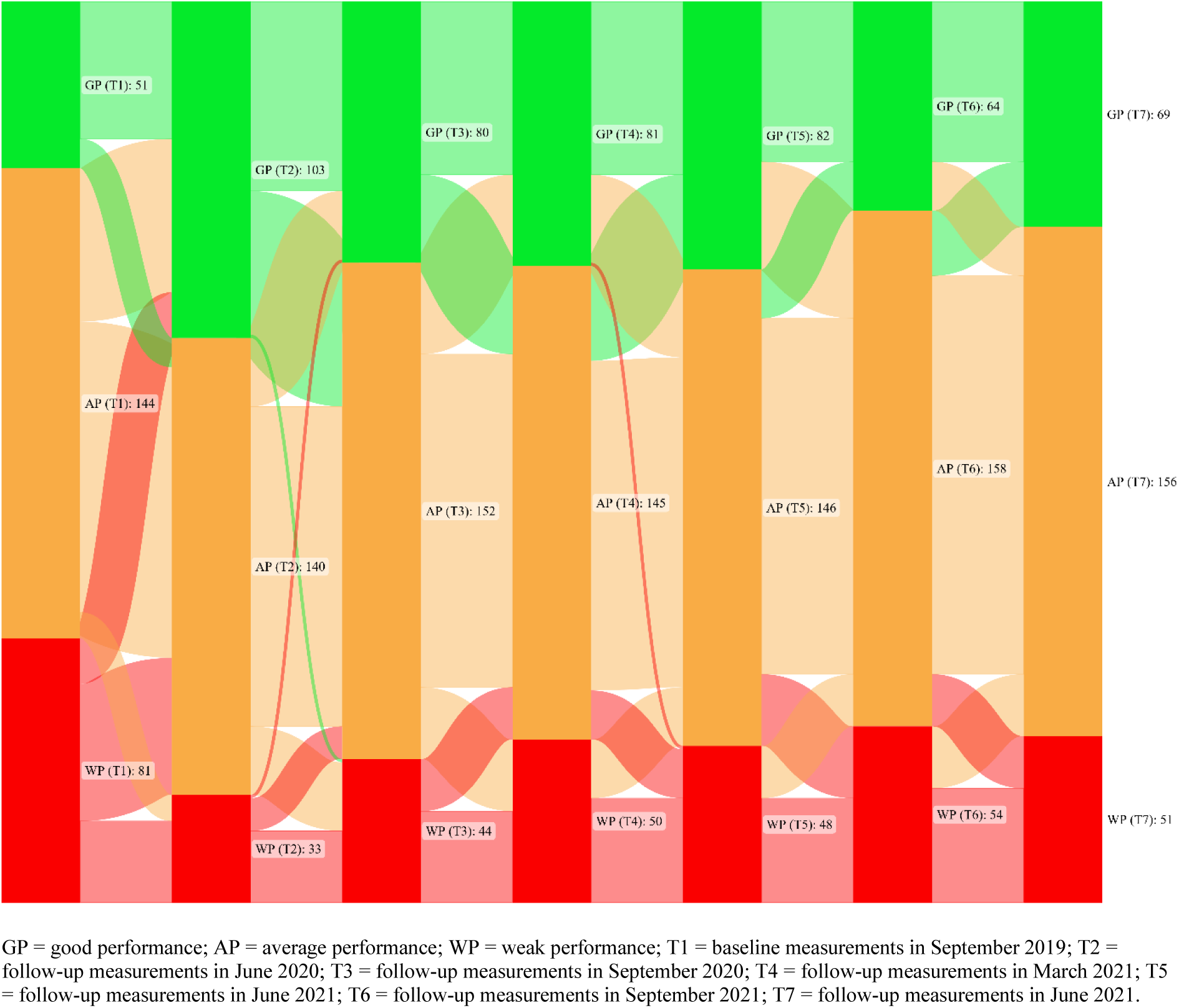
Changes in standing long jump performance groups between September 2019 and June 2022.

Between the baseline measurement in September 2019 and the first measurement point (June 2020) after the COVID-19 pandemic outbreak, an increase was observed in the number of children in the good performance groups (September 2019 (*n* = 51), June 2020 (*n* = 103), p < 0.001); in contrast, the number of children in the poor performance groups decreased (September 2019 (*n* = 81), June 2020 (*n* = 33), *p* < 0.001) (Table R21 and Figure R25).

From June 2020 to September 2020, the number of children in the good performance groups decreased significantly (*z* = –5.64, *p* < 0.001). At the subsequent test time points in March 2021 (*z* = –0.85, *p* > 0.99) and June 2021 (*z* = –0.49, *p* > 0.99), no significant changes in the performance classification groups were observed compared to the previous testing time point. Between June 2021 and September 2021, a significant improvement was observed (*z* = –3.39, *p* = 0.015). In the last observation period between September 2021 and June 2022, no significant changes in performance classification groups were observed (*z* = –1.42, *p* > 0.99) (Table R21 and Figure R25).

Comparing the number of children in all three classification groups (weak, average, and good performance) from baseline (September 2019) before COVID-19 against the last test time point (June 2022), highly significant changes were seen over the entire observation period (*z* = –4.43, *p* < 0.001) (Table R21 and Figure R25).

#### Medicine Ball Throw (MB1kg)

Highly significant changes (Friedman test: Chi-square (6) = 56.06, *p* < 0.001) were observed in the medicine ball throw (1kg) over the overall observation time period (September 2019 to June 2022) (Table R20 and Figure R26).

**Figure R26.**
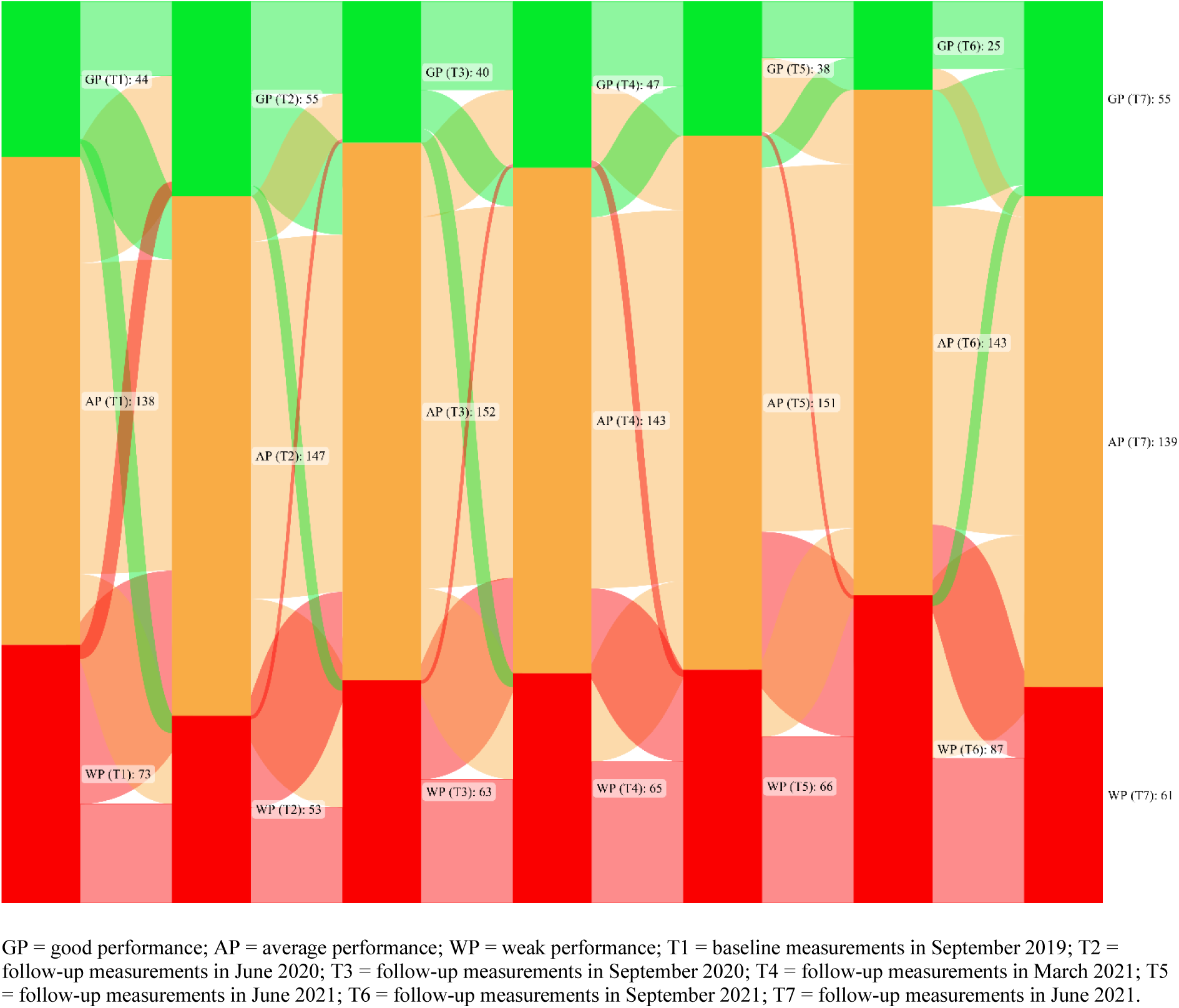
Changes in medicine ball throw (1kg) performance groups between September 2019 and June 2022.

No significant changes were found between the baseline measurement in September 2019 (T1) and the follow-up measurement time points in June 2020 (T2), September 2020 (T3), March 2021 (T4), and June 2021 (T5) (T1–T2: *z* = –2.53, *p* = 0.24; T2–T3: *z* = –2.56, *p* = 0.22; T3–T4: *z* = –0.54, *p* > 0.99; T4–T5: *z* = –0.34, *p* > 0.99) (Table R21 and Figure R26).

A significant decline was observed between June 2021 and September 2021 (T6) (*z* = –4.59, *p* < 0.001). This was followed by a period of significant improvement in performance between September 2021 and June 2022 (*z* = –6.20; *p* < 0.001). This improvement reached, in June 2022, the level of measurement in June 2021 (*z* = –1.39, *p* > 0.99) (Table R21 and Figure R26).

Comparing the number of children in all three classification groups (weak, average, and good performance) from baseline (September 2019) before COVID-19 against the last test time point (June 2022), there were no significant changes over the overall observation time period (*z* = –1.65, *p* > 0.99) (Table R21 and Figure R26).

#### 4 × 10 m Shuttle Run (4×10SHR)

Highly significant changes (Friedman test: Chi-square (6) = 387.99, *p* < 0.001) were observed in action speed (4 × 10 m run) over the overall observation time period (September 2019 to June 2022) (Table R20 and Figure R27).

**Figure R27.**
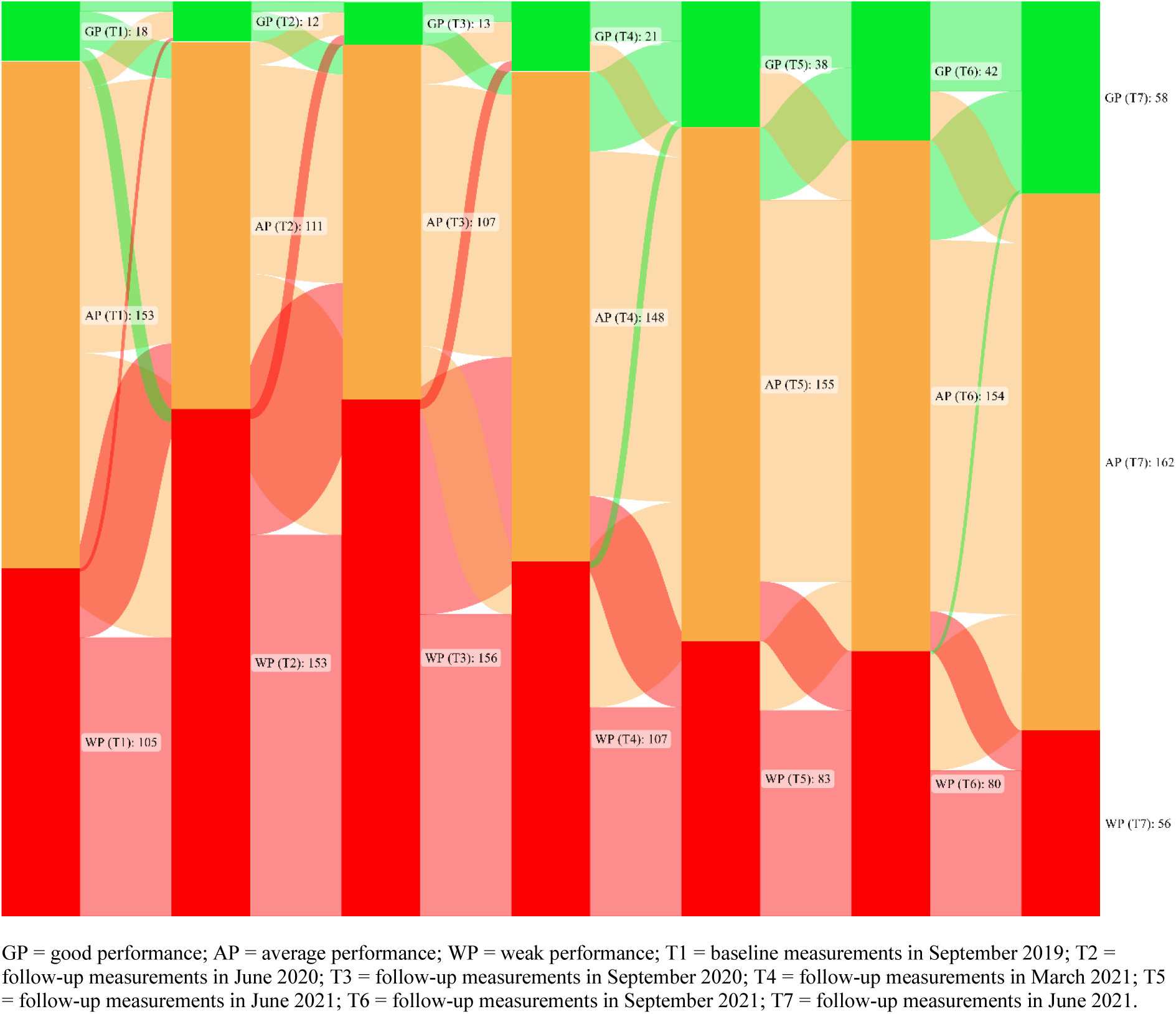
Changes in action speed performance groups between September 2019 and June 2022.

A significant decline in performance (*z* = –5.19, *p* < 0.001) was found between the baseline measurement in September 2019 and the first measurement time point (June 2020) after the outbreak of the COVID-19 pandemic, with a dramatic increase of children in the weak performance groups (September 2019 (*n* = 105), June 2020 (*n* = 153), *p* < 0.001). In contrast, the number of children in the good performance groups decreased sharply (September 2019 (*n* = 18), June 2020 (*n* = 12), *p* < 0.001). As the COVID-19 pandemic continued, continuous improvement was observed, reaching baseline levels at time T4 (March 2021) (T1–T4: *z* = – 0.44, *p* > 0.99). The continuous increase in performance was ongoing and at the last measurement time point in June 2022, a significant improvement in action speed was shown compared to the baseline measurement in September 2019 (*z* = –8.98, *p* < 0.001) (Table R21 and Figure R27).

#### Jumping Sideways (JS)

Significant changes (Friedman test: Chi-square (5) = 19.52, *p* = 0.002) were observed in jumping sideways over the overall observation time period (September 2019 to June 2022) (Table R20 and Figure R28).

**Figure R28.**
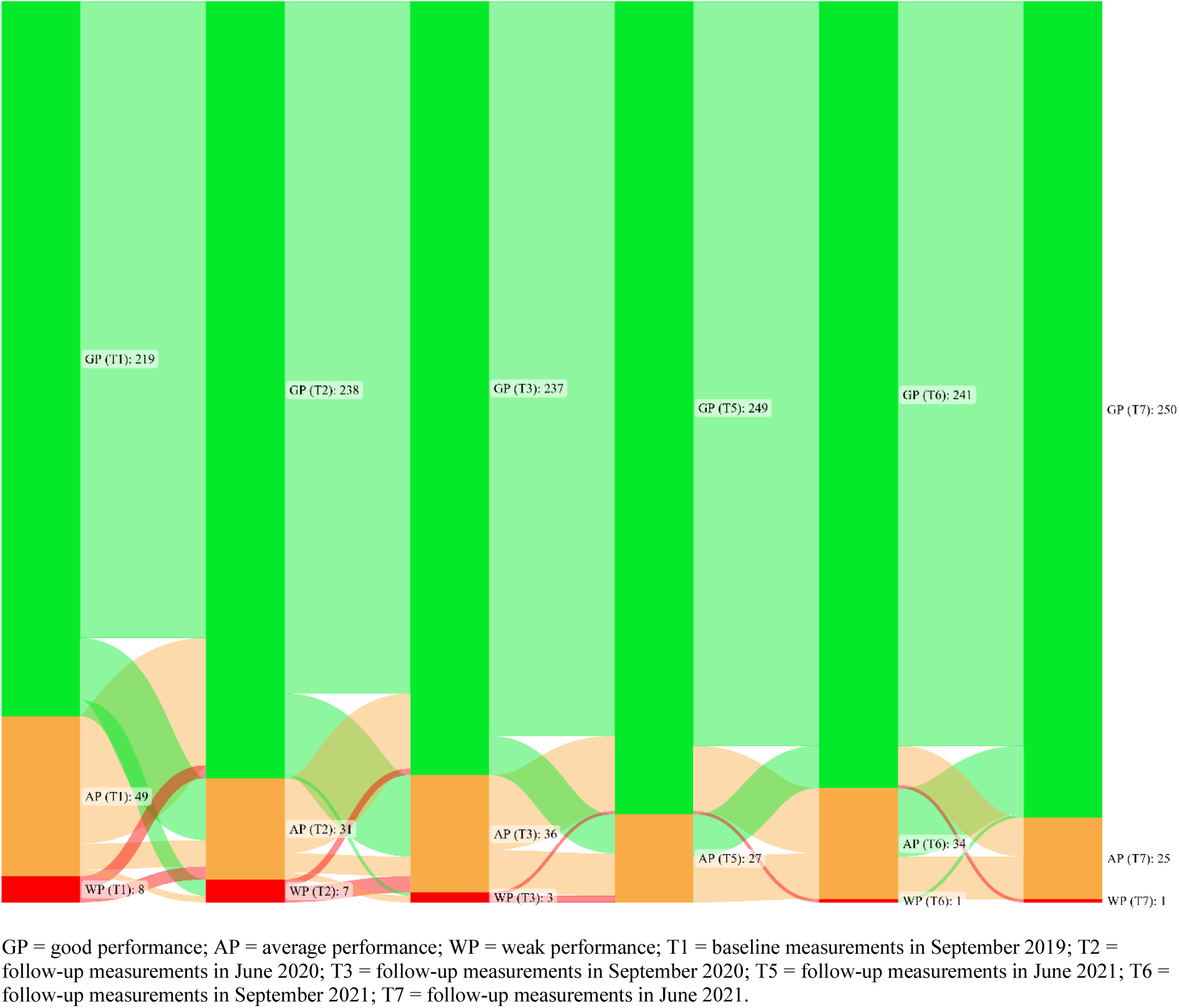
Changes in whole-body coordination performance groups between September 2019 and June 2022.

In the last observation time period between September 2021 and June 2022, no significant changes were found in the performance classification groups (T6–T7: *z* = –2.86, *p* = 0.06). (Table R21 and Figure R28).

Whole-body coordination showed significant changes in the different performance groups during the observation period (T1–T7: *z* = –4.04, *p* < 0.001). The number of children in the good performance groups increased continuously, these results have to be considered with caution as reported by Jarnig et al. [1] and (Table R21 and Figure R28).

#### V Sit-and-Reach Test (VSR)

Highly significant changes in flexibility (Friedman test: Chi-squared (5) = 70.21, *p* < 0.001) were observed over the overall observation time period (September 2019 to June 2022) (Table R20 and Figure R29).

**Figure R29.**
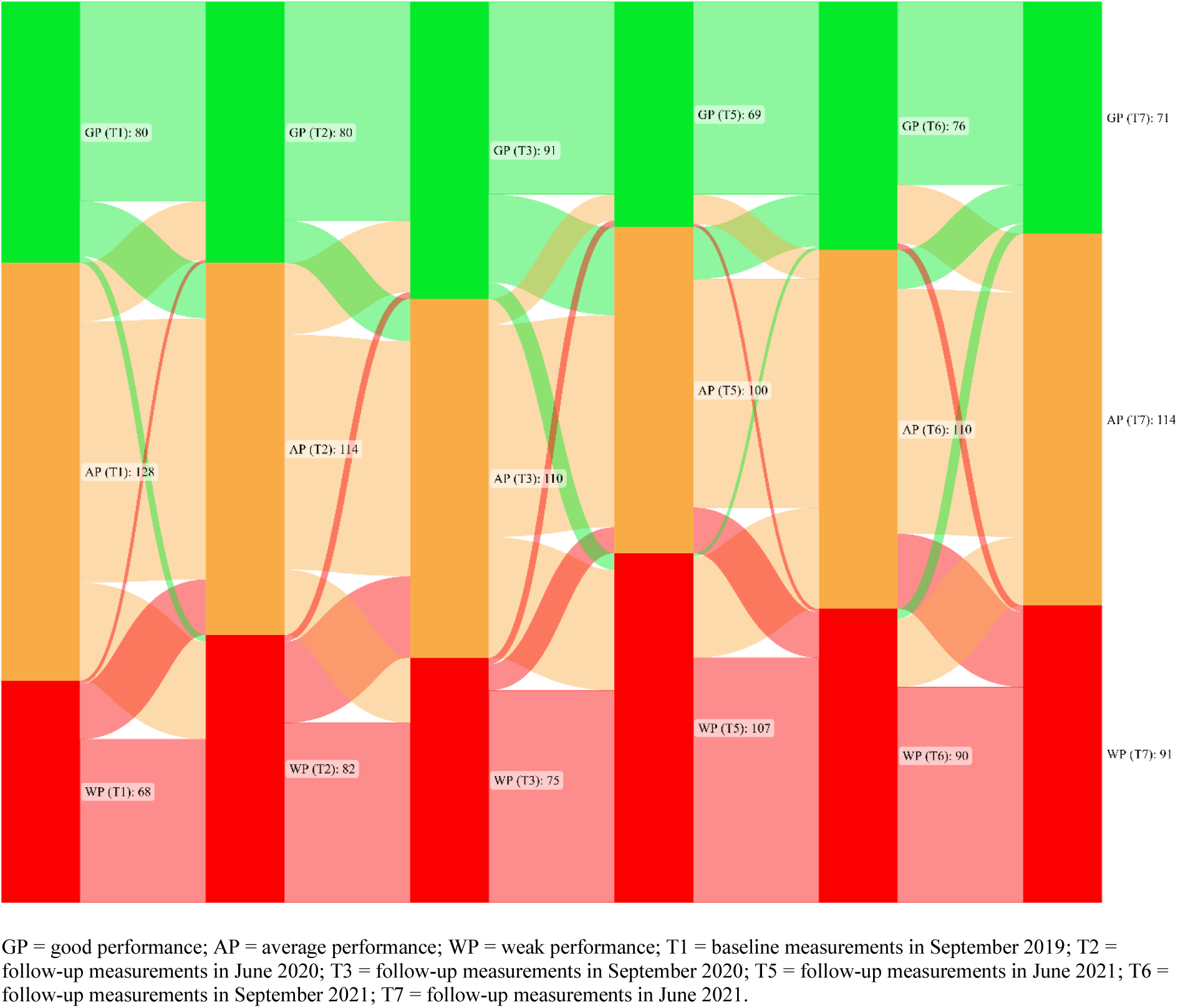
Changes in flexibility performance groups between September 2019 and June 2022.

A slight increase in the number of children in the weak performance groups was found after the first lockdown period in June 2020, but this did not represent a significant change (September 2019 (*n* = 68), June 2020 (*n* = 82), *p* > 0.99) from the baseline measurement. No significant changes were found in the performance classification groups from June 2020 to September 2020 (*z* = –2.29, *p* = 0.33) (Table R21 and Figure R29).

At the measurement time point after the second lockdown period in June 2021, there was a significant decline in flexibility with an increase in the number of children in the low-performing groups (WP: September 2020 (*n* = 75), June 2021 (*n* = 107); *z* = –6.97, *p* < 0.001). This negative trend ended in the period June 2021 to September 2021 and there was a significant improvement in flexibility in September 2021 compared to June 2021(*z* = –4.44, *p* < 0.001) (Table R21 and Figure R29).

In the last observation time period between September 2021 and June 2022, no significant changes were found in the performance classification groups (T6–T7: *z* = –1.32, *p* > 0.99). (Table R21 and Figure R29).

#### Reaction Speed (RD)

Overall, the reaction time showed highly significant increases and decreases in the number of children in each performance group during the observation period between the individual time spans of the measurement time points (Friedman test: Chi-square (5) = 179.79, *p* < 0.001), these results must be viewed with caution as reported in Jarnig et al. [1] (Table R20 and Figure R30).

**Figure R30.**
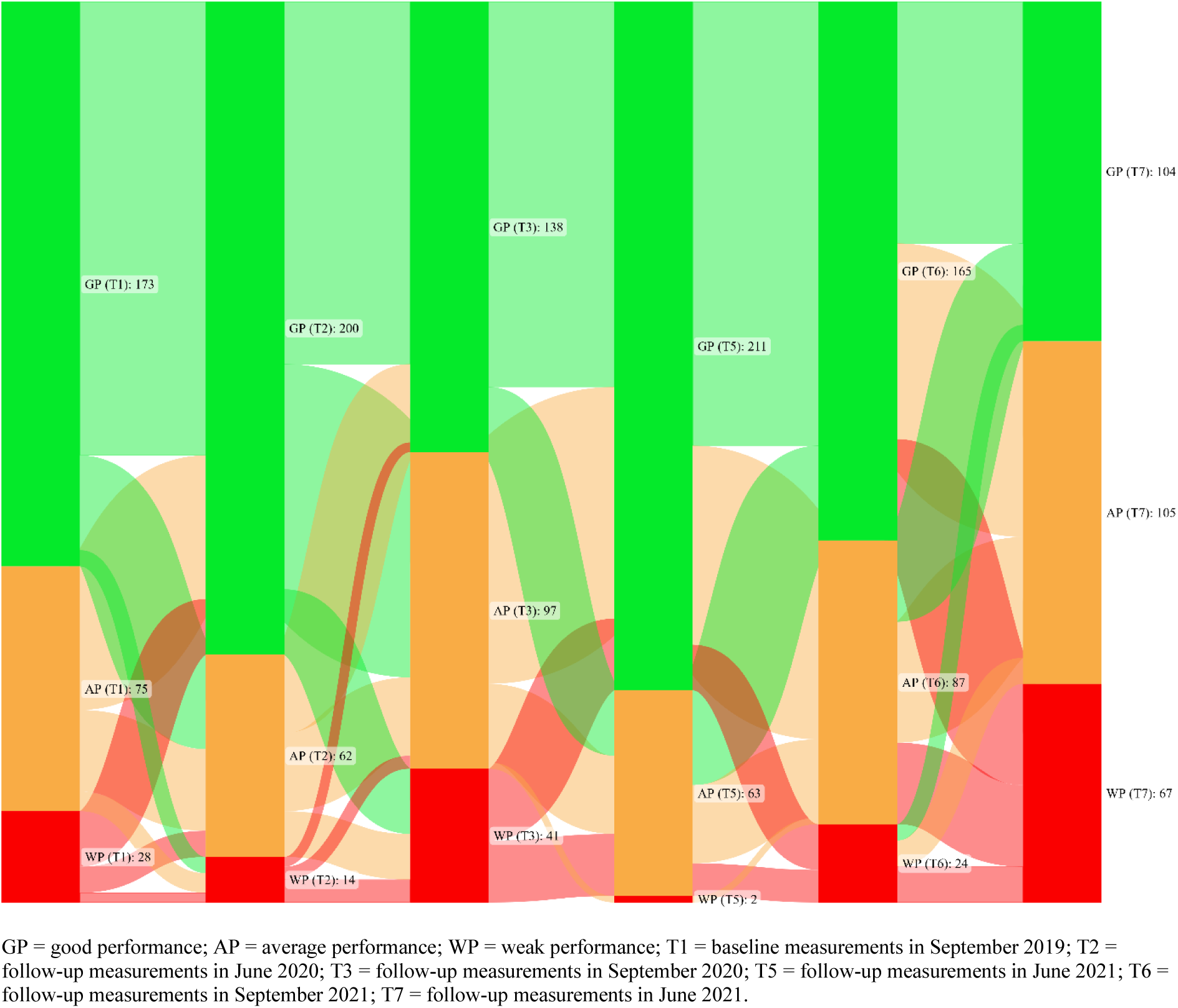
Changes in reaction time performance groups between September 2019 and June 2022.

In the last observation period between September 2021 and June 2022, a highly significant decline (T6–T7: *z* = –6.44, *p* < 0.001) in reaction speed was observed (Table R21 and Figure R30).

#### Balance (SLS)

Highly significant changes in balance (Friedman test: Chi-squared (5) = 279.42, p < 0.001) were observed over the overall observation time period (September 2019 to June 2022) (Table R20 and Figure R31).

**Figure R31.**
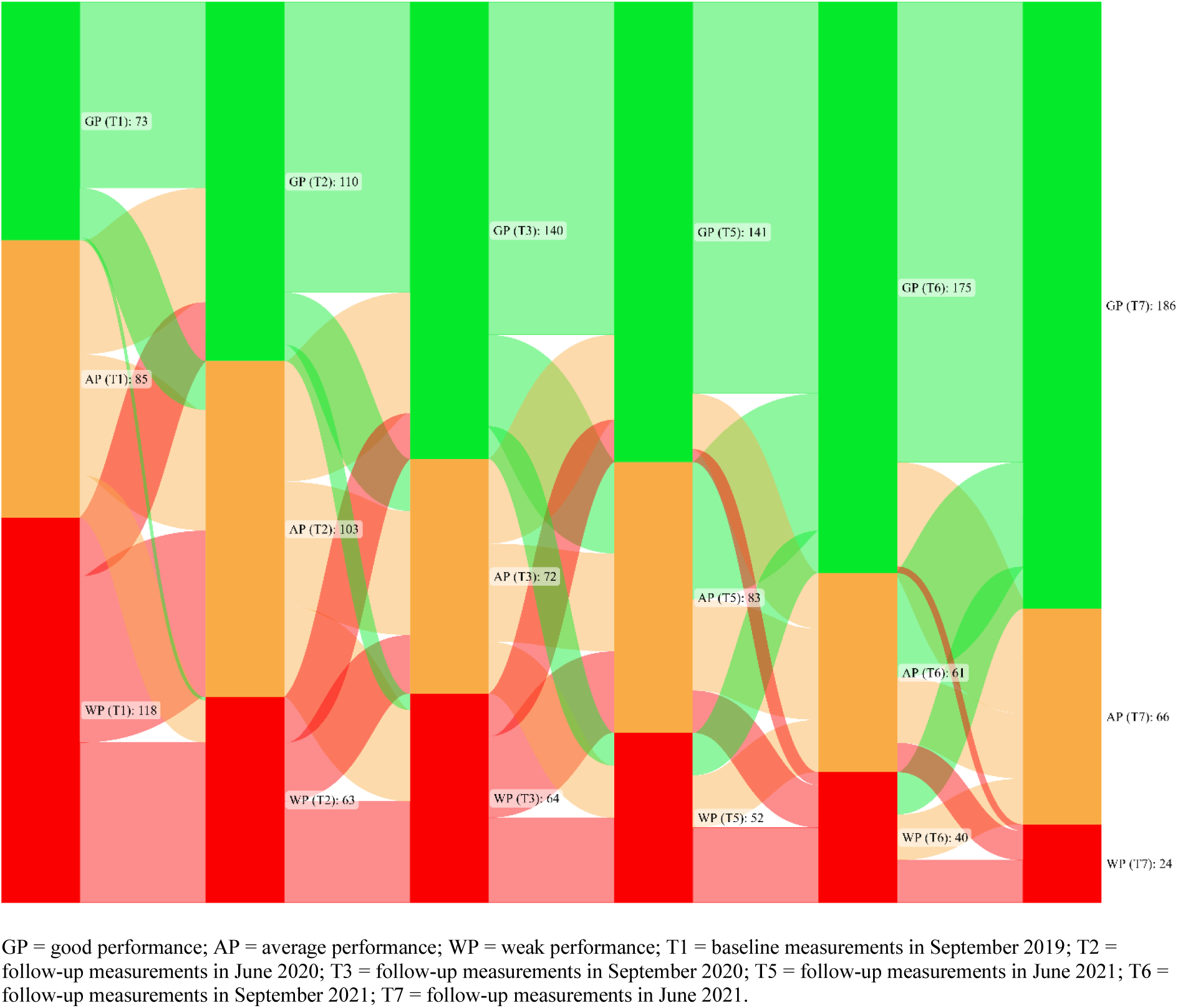
Changes in the balance performance groups between September 2019 and June 2022.

Small significant changes in the performance classification groups (T6–T7: *z* = –3.00, *p* = 0.041) were observed in the last observation time period between September 2021 and June 2022 (Table R21 and Figure R31).

The number of children in the good performance groups increased continuously throughout the observation period from September 2019 to June 2022 (September 2019 (*n* = 73), June 2022 (*n* = 186), *p* < 0.001) (Table R21 and Figure R31).

## DISCUSSION

In line with our findings at the beginning of the pandemic, there are already a number of studies reporting alarming increases in BMI associated with the COVID-19 pandemic [18,20,21,23]. To the best of our knowledge, our study is the first worldwide to provide data on children’s anthropometric and athletic development after relaxation of the stringent COVID-19 mitigation measures (starting June 2021).

We found an increase in BMI during the COVID-19 pandemic until March 2021, and as a result, the number of children with overweight or obesity increased steadily from 14.2% to 21.1% during the period from September 2019 to March 2021. The significant BMI increase at the beginning of the pandemic was followed by a marked improvement in weight classification toward (June 2022. In June 2022, a small increase in the number of children with overweight or obesity was noted compared to prior to COVID-19—September 2019, but these changes were not statistically significant (using a Bonferroni correction for all pairwise comparison). The results suggest that the negative health effects of the pandemic are reparable, at least in “very” young populations.

Changes in waist circumference are reported to be more useful than BMI changes in detecting behavioral changes associated with changes in energy balance. However, there are weaknesses in measuring waist circumference, because waist circumference tends to vary with the time of day and with eating and drinking patterns, and is subject to seasonal variation. Therefore, the waist-to-height ratio has been referred to in scientific reports as a better screening tool for cardiometabolic risk factors than BMI or waist circumference alone. Different trends in the WtHR were observed during the pandemic. The WtHR increased during the lockdown periods (March 2020 to May 2020 and November 2020 to February 2021) and decreased after relaxation of the COVID-19 mitigation measures. It is important to note that the number of children suffering from increased and high health risk also increased during the period from June 2021 to September 2021, suggesting that the increase during the lockdown periods was not exclusively due to the COVID-19 mitigation measures. It appeared, in the context of the waist-to-height ratio (WtHR), that the negative effects, which may have also been predominantly due to by the stringent COVID-19 mitigation measures, seem reparable in primary school children.

Similar results can be observed for the health-related fitness parameter cardiorespiratory endurance (six-minute run) and for the performance-related parameter action speed. A dramatic loss in cardiorespiratory fitness between September 2019 and September 2020 was followed by a steady improvement toward exit testing (June 2022). However, for cardiorespiratory fitness performance, even in June 2022, a significantly lower level than before COVID-19 was found. For action speed, at the beginning of the pandemic, a worrying decrease in performance was observed; at the end of the observation period (June 2022) the children had compensated for this drastic loss in performance and fortunately showed a significant improvement in performance compared to the baseline level before COVID-19 (September 2019). However, the action speed results must be interpreted cautiously, as it is possible that repeated performance and associated technique improvement in the 4 × 10 m run had positive effects on the overall results.

For the standing long jump at the beginning of the observation period, taking into account the results of weight classification, health risk estimation, cardiorespiratory fitness, and action speed, there was a surprisingly strong improvement in performance, which was probably due to an improvement in the technique of jumping execution. Minor changes in standing long jump performance were observed later in the pandemic, most of which were not significant.

A significant improvement in performance was observed in the medicine ball throw at the beginning of the pandemic. In scientific studies, it has been proven that children who are overweight or obese perform better in completing the medicine ball throw than those who are underweight or of normal weight [42]. This may be an explanation for the tremendous improvement in performance. Additionally, improved technique in the execution of the throw may be a factor in the positive performance in the first phase of the COVID-19 pandemic. Between June 2020 and September 2020, a decline in performance in medicine ball throwing was observed, and no significant changes were found at four of the five subsequent testing time points compared to the entry testing prior to COVID-19.

The results in the standing long jump and medicine ball throw might be related to the fact that strength gains in this age group are usually not trained by specific strength training, but are rather achieved by natural growth. Therefore, the COVID-19-related mitigation measures do not seem to have had long-term negative effects on the children’s muscle strength.

When investigating flexibility, slightly significant variations were observed over the course of the pandemic, with a slight decline in performance, related to baseline and exit testing. This is due to a natural evolution, as previous studies have proven that flexibility at a young age decreases with growth without specific training of this [37]. The remarkable decline in flexibility at time T5 (June 2021) compared to time T3 (September 2020) can be explained by a change in the testing procedure at time T5 occurred. In June 2021, due to organizational time restrictions, the testing of flexibility and reaction speed had to be conducted in parallel to regular school lessons. The flexibility results at time T5 (June 2021) can therefore be neglected when summarizing the overall fitness results.

The results of jumping sideways showed no significant changes compared to the performance before the COVID-19 pandemic.

Exciting results were observed in the test of reaction speed, which was performed using the ruler drop test and provided an indication of eye-hand coordination speed. After the lockdown phases (June 2022 and March 2021), highly significant performance gains in eye–hand reaction speed were shown. Studies have reported that during lockdown phases, increased screen time was observed in children and adolescents worldwide [5,6,8–10]. This could explain the strong gains in the ruler drop test, as eye–hand reaction time is permanently trained during computer activities [43]. In contrast, during the phases of relaxation of the COVID-19 mitigation measures, significant performance declines were observed, and at the end of the observation period, a significant performance decline occurred compared to the baseline measurement before COVID-19 in September 2019.

For balance, as described by our group previously, the ceiling effect was reinforced further in the last observation period (September 2021 to June 2022) and a continuous increase in balance ability was monitored, which is not a unique characteristic at this age group but can be explained by developmental steps in growth [39]. There were no abnormalities suggesting a worrisome influence of the COVID-19 conditional mitigation measures on the children’s balance ability.

A major strength of our study is that all data were collected by a team of six trained sports scientists and physical educators and that the individual measurements in the different test phases were conducted by the same test leader. Another strength is that all of the collected data were objectively measured directly on the participants and that there was a randomized and large sample size that allowed a general statement about the development of fitness and health status of primary school children in Austria over the course of the COVID-19 pandemic. The presence of current national reference values (EQUI BMI) can also be mentioned as an additional strength.

On the other side, one limitation is the lack of national reference values for fitness. Another weakness related to the fitness reference values used is the fact that for some fitness tests, only reference values in annual steps are available, which might have an impact on the SDS values and standardized *z*-scores.

Regarding the results of the standing long jump, it would make sense to discuss and pre-test the technique of the standing long jump in future observation regions before testing it for the first time and to let the children practice it thereafter in order to be able to exclude performance increases through technique improvements. For such purposes, it would make sense to have basic athletic elements worked out in a fixed manner in connection with the curriculum as early as kindergarten age.

## CONCLUSION

Our study showed that the COVID-19-related mitigation measures had worrisome negative effects on the health of primary school-aged children in Austria. Fortunately, these seem to be reparable in primary school children. Our findings, in addition to other studies of the effects of specific COVID-19 mitigation measures [44–47], provide a basis for policymakers to target and mandate future virus-related mitigation measures in an evidence-based manner.

Their advantage in terms of virus transmission must be carefully weighed against the long-term disadvantage in terms of BMI and fitness and thus physical health in developing individuals, i.e. children and adolescents. Continuous physical activities should be considered as important as infection prevention, also in schools.

It is satisfying to observe that the worrisome trends in terms of BMI, WtHR and health-related fitness at the beginning of the pandemic, halted and a reversal was initiated in the further course of the pandemic, reducing the health damage caused.

In the future, national and international activities, considerations, and interventions in all age groups will be required in order to promote positive effects on the health development of the whole population and to be able to achieve and guarantee the best possible conditions especially for growing and developing individuals.

## Data Availability

All data produced in the present work are contained in the manuscript

## Supplementary Materials

The tables attached below present the impact of the sports intervention implemented during the last observation period, from September 2021 to June 2022, on the evolution of the children’s fitness and health status.

## Author Contributions

Conceptualization, G.J. and M.N.M.v.P.; methodology, G.J.; formal analysis, G.J.; investigation, G.J.; resources, G.J.; data curation, G.J.; writing—original draft preparation, G.J.; writing—review and editing, G.J., R.K., and M.N.M.v.P.; visualization, G.J.; supervision, M.N.M.v.P.; project administration, G.J.; funding acquisition, G.J. All authors read and agreed to the published version of the manuscript.

## Institutional Review Board Statement

This study was conducted according to the guidelines of the Declaration of Helsinki and approved by the Research Ethics Committee of the University of Graz, Styria, Austria (GZ. 39/23/63 ex 2018/19).

## Informed Consent Statement

Informed consent was obtained from all subjects involved in the study.

## Data Availability Statement

The data presented in this study are available upon request from the corresponding author. The data are not publicly available due to privacy/ethical restriction.

## Acknowledgments

This study was organized by the non-profit association NAMOA— Nachwuchsmodell Austria. The authors would like to thank all participants and their guardians; the trainers and staff of this study; Wolfgang Modritz for the initiation of this study; Peter Hofmann (University of Graz) for the support in the conception phase; Robert Klinglmair (Education Directorate of Carinthia, Austria) for his authorized assessments in schools; Christian Günter (Austrian Federal Ministry of Sport) and Hannes Wolf (Education Directorate of Carinthia, Austria) for helping to continue the assessments after the COVID-19 lockdown. None of the individuals listed were financially compensated.

## Supplementary Materials

Tables X1 and X2 provide an overview of the analysis of the effects already described in the Results section of the sports intervention that took place between T6 and T7.

**Figure X1.**
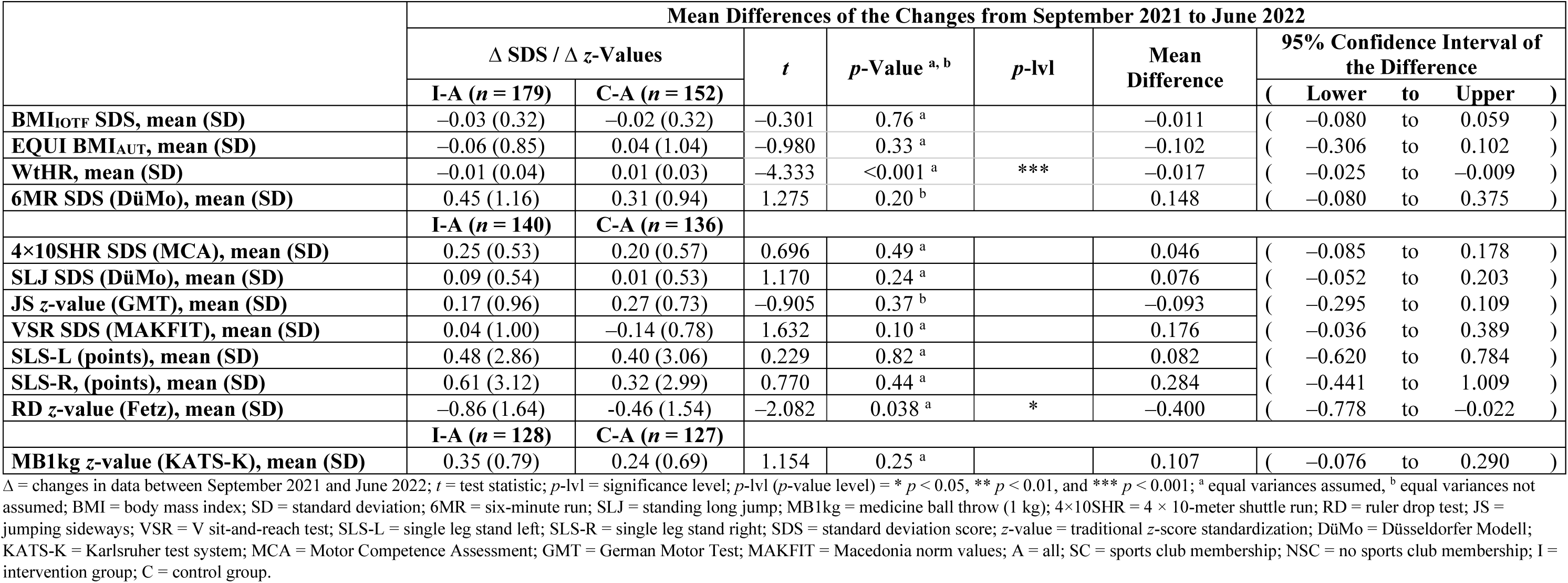
Impact of the athletic intervention in SY 2021–2022 on BMI, WtHR, and health- and performance-related fitness parameters on the study population

**Figure X2.**
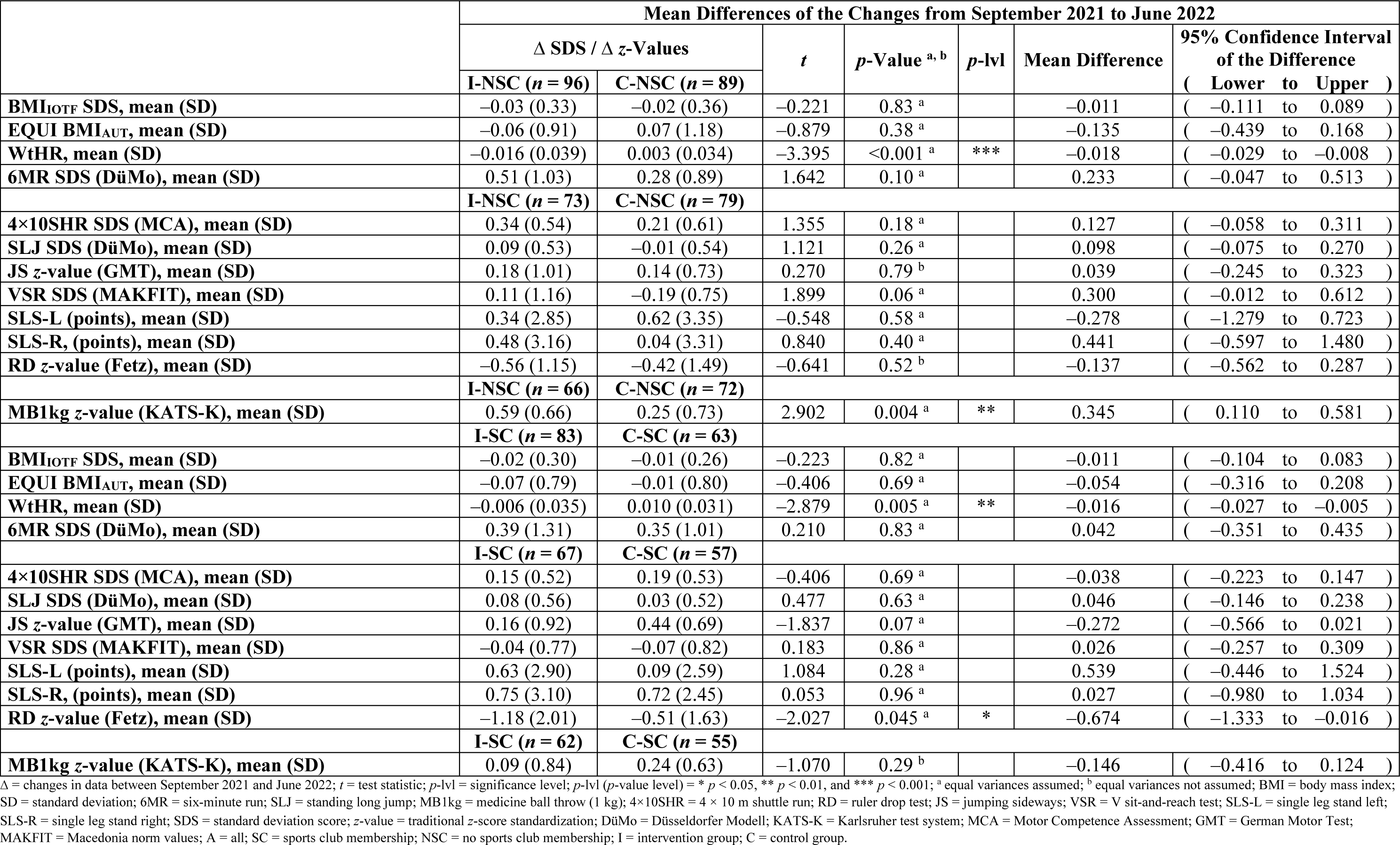
Effects of the sports intervention in SY 2021–2022 on the BMI, WtHR, and health- and performance-related fitness parameters in the subgroups of sports club membership (Yes or No).

